# CIRCULATING IMMUNE DYNAMICS DURING NEOADJUVANT CHEMOIMMUNOTHERAPY IN EARLY TRIPLE-NEGATIVE BREAST CANCER

**DOI:** 10.64898/2026.11.19.26368490

**Authors:** Javier Pascual, Maria Rosario Chica-Parrado, Clotilde Costa, Roberto Pineiro, Maria Elena Quirós-Ortega, Álvaro González-Ortiz, Ana Godoy, Maria Emilia Domínguez, Nuria Ribelles, Bella Pajares, Antonia Márquez-Aragonés, Begoña Jiménez-Rodríguez, Pilar Cantizani, Patricia Palacios-Ozores, Esperanza López-López, Patricia Chaves-Guerrero, Jesús Velasco-Suelto, Maria Victoria Ortega-Jiménez, María Isabel Hierro-Martín, Martina Álvarez, Enrique de Alava, Rafael López-López, Emilio Alba, Iñaki Comino-Méndez

## Abstract

**Background:** Pathological complete response (pCR) after pembrolizumab-based neoadjuvant chemoimmunotherapy predicts improved outcomes in early triple-negative breast cancer (TNBC), but circulating immune determinants of response remain poorly characterized.

**Methods:** The study included 64 patients with early TNBC treated with pembrolizumab-based neoadjuvant chemoimmunotherapy. Flow cytometry analyzed 125 peripheral-blood specimens collected at baseline, during treatment and before surgery. Baseline PBMCs from 22 patients underwent scRNA-seq, generating 135,271 quality-controlled cells. Analyses used nonparametric tests, effect sizes, FDR correction and sensitivity analyses.

**Results:** Baseline scRNA-seq identified response-associated transcriptional differences. Patient-level pseudo-bulk analysis revealed enrichment of TNF-α/NF-κB signaling in cytotoxic T/NK and B cells from non-pCR patients (global q≤3.94×10⁻□), together with MYC targets and mTORC1 signaling in cytotoxic T/NK cells (both global q=7.00×10⁻□). T-cell analyses suggested preserved naïve/memory features in pCR, while a higher proportion of *CX3CR1*-expressing cells within the CD4 memory/activation-like compartment was associated with pCR (q=0.0157; AUC=0.949). In B cells, pCR was associated with a nominally higher naïve-like program score (P=0.029; q=0.059), supported by higher *IGHM* and *IGHD* signals, whereas *XBP1* was higher in non-pCR. Flow cytometry identified treatment-associated B-cell remodeling. At pre-surgery, switched-memory B-cell CD38 balance was higher in pCR than non-pCR patients (62.6% versus 50.0%; P=0.0086; q=0.0667; δ=0.651; AUC=0.825) and remained nominally significant in leave-one-patient-out analyses. Early increases in unswitched-memory B-cell CD38 balance were numerically greater in non-pCR patients, whereas pre-surgery switched-memory B-cell CD38 balance was higher in pCR patients.

**Conclusions:** Baseline inflammatory and metabolic pathway enrichment was associated with non-pCR, whereas pCR-associated T-cell findings included a CD4 memory/activation-associated *CX3CR1* signal and trends towards preserved naïve/memory features. Together with time- and compartment-dependent B-cell remodeling, these findings nominate circulating transcriptional states and CD38-defined B-cell dynamics for prospective validation as candidate biomarkers of pathological response.

## INTRODUCTION

Pembrolizumab plus neoadjuvant chemotherapy is a standard treatment for high-risk early triple-negative breast cancer (TNBC), improving pathological complete response (pCR), event-free survival and overall survival in KEYNOTE-522 ^1,2^. Although pCR predicts favorable long-term outcomes, patients with residual disease remain at risk of recurrence ^3^. However, the immune features associated with pathological response to pembrolizumab-based neoadjuvant chemoimmunotherapy remain incompletely characterized ^2^. Biomarkers reflecting immune competence are therefore needed.

TNBC is among the most immunogenic breast cancer subtypes, and higher tumor-infiltrating lymphocyte levels are associated with neoadjuvant treatment sensitivity and improved outcomes ^4^. Beyond the overall abundance of immune cells, response to immune-checkpoint blockade has been associated with specific functional states and coordinated interactions among T cells, B cells, dendritic cells and myeloid populations ^5–7^. As these features can evolve during treatment, longitudinal immune profiling may reveal response-associated changes that are not captured by a single pretreatment assessment ^8^. However, much of this evidence comes from tumor tissue studies, often conducted in advanced disease or heterogeneous clinical settings. Repeated tumor biopsies are invasive and capture only a limited region of the tumor, limiting their suitability for longitudinal immune monitoring.

Peripheral blood offers a minimally invasive means of assessing systemic immunity and tracking its evolution during treatment, with studies in advanced cancer and pan-cancer cohorts linking circulating T- and B-cell profiles to response to immune-checkpoint inhibitors ^9–11^. However, given the recent adoption of pembrolizumab-based neoadjuvant chemoimmunotherapy in early TNBC, evidence relating pretreatment circulating immune states and their evolution during therapy to pathological response remains limited.

Here, we integrated single-cell RNA sequencing of baseline peripheral blood mononuclear cells (PBMCs) with multiparametric flow cytometry performed in an expanded cohort at baseline, at an intermediate timepoint after the first therapeutic block and after completion of neoadjuvant treatment before surgery. We sought to define baseline circulating immune states associated with pCR, characterize systemic immune remodeling during treatment and identify longitudinally measurable surface phenotypes associated with pathological response.

## METHODS

Detailed experimental and analytical procedures are provided in the **Supplementary Methods.**

### Study design and patient population

This study included patients with early-stage TNBC treated with pembrolizumab-based neoadjuvant chemoimmunotherapy at the Virgen de la Victoria University Hospital in Málaga and the University Hospital Complex of Santiago de Compostela. TNBC was defined according to institutional pathological criteria as the absence of estrogen receptor and progesterone receptor expression and lack of HER2 overexpression or amplification. Patients were eligible if they were aged ≥18 years, had a diagnosis of early-stage TNBC, were scheduled to receive pembrolizumab-based neoadjuvant chemoimmunotherapy as per international guideline recommendations, and had provided written informed consent for the donation and research use of biological samples. Patients with synchronous malignancies, or with a diagnosis of or treatment for another type of malignancy at the time of donation or during the preceding 5 years, were excluded. Patients received pembrolizumab-based neoadjuvant chemoimmunotherapy according to a KEYNOTE-522 clinical trial–based regimen. The first treatment block consisted of pembrolizumab administered every 3 weeks for four cycles in combination with weekly paclitaxel for 12 weeks and carboplatin administered either weekly or every 3 weeks. This was followed by four cycles of pembrolizumab combined with an anthracycline and cyclophosphamide, administered every 3 weeks, before surgery. The actual number of treatment cycles administered varied among patients (**Table 1**).

**TABLE 1.**
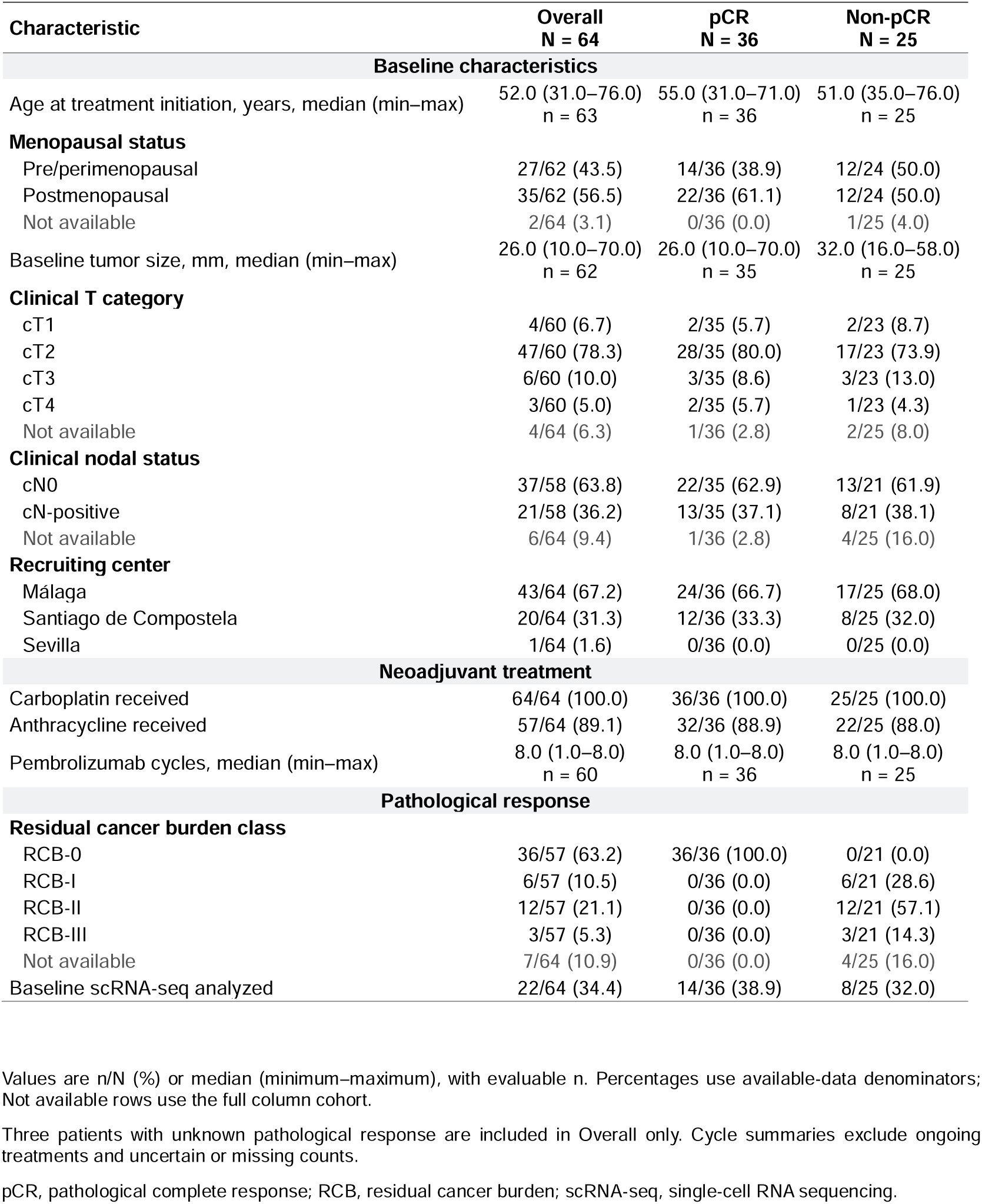
xxxxx

| Characteristic | **Overall**<br>**N = 64** | **pCR**<br>**N = 36** | **Non-pCR**<br>**N = 25** |
|---|---|---|---|
| **Baseline characteristics** |  |  |  |
| Age at treatment initiation, years, median (min–max) | 52.0 (31.0–76.0)<br>n = 63 | 55.0 (31.0–71.0)<br>n = 36 | 51.0 (35.0–76.0)<br>n = 25 |
| **Menopausal status** |  |  |  |
| Pre/perimenopausal | 27/62 (43.5) | 14/36 (38.9) | 12/24 (50.0) |
| Postmenopausal | 35/62 (56.5) | 22/36 (61.1) | 12/24 (50.0) |
| Not available | 2/64 (3.1) | 0/36 (0.0) | 1/25 (4.0) |
| Baseline tumor size, mm, median (min–max) | 26.0 (10.0–70.0)<br>n = 62 | 26.0 (10.0–70.0)<br>n = 35 | 32.0 (16.0–58.0)<br>n = 25 |
| **Clinical T category** |  |  |  |
| cT1 | 4/60 (6.7) | 2/35 (5.7) | 2/23 (8.7) |
| cT2 | 47/60 (78.3) | 28/35 (80.0) | 17/23 (73.9) |
| cT3 | 6/60 (10.0) | 3/35 (8.6) | 3/23 (13.0) |
| cT4 | 3/60 (5.0) | 2/35 (5.7) | 1/23 (4.3) |
| Not available | 4/64 (6.3) | 1/36 (2.8) | 2/25 (8.0) |
| **Clinical nodal status** |  |  |  |
| cN0 | 37/58 (63.8) | 22/35 (62.9) | 13/21 (61.9) |
| cN-positive | 21/58 (36.2) | 13/35 (37.1) | 8/21 (38.1) |
| Not available | 6/64 (9.4) | 1/36 (2.8) | 4/25 (16.0) |
| **Recruiting center** |  |  |  |
| Málaga | 43/64 (67.2) | 24/36 (66.7) | 17/25 (68.0) |
| Santiago de Compostela | 20/64 (31.3) | 12/36 (33.3) | 8/25 (32.0) |
| Sevilla | 1/64 (1.6) | 0/36 (0.0) | 0/25 (0.0) |
| **Neoadjuvant treatment** |  |  |  |
| Carboplatin received | 64/64 (100.0) | 36/36 (100.0) | 25/25 (100.0) |
| Anthracycline received | 57/64 (89.1) | 32/36 (88.9) | 22/25 (88.0) |
| Pembrolizumab cycles, median (min–max) | 8.0 (1.0–8.0)<br>n = 60 | 8.0 (1.0–8.0)<br>n = 36 | 8.0 (1.0–8.0)<br>n = 25 |
| **Pathological response** |  |  |  |
| **Residual cancer burden class** |  |  |  |
| RCB-0 | 36/57 (63.2) | 36/36 (100.0) | 0/21 (0.0) |
| RCB-I | 6/57 (10.5) | 0/36 (0.0) | 6/21 (28.6) |
| RCB-II | 12/57 (21.1) | 0/36 (0.0) | 12/21 (57.1) |
| RCB-III | 3/57 (5.3) | 0/36 (0.0) | 3/21 (14.3) |
| Not available | 7/64 (10.9) | 0/36 (0.0) | 4/25 (16.0) |
| Baseline scRNA-seq analyzed | 22/64 (34.4) | 14/36 (38.9) | 8/25 (32.0) |

Values are n/N (%) or median (minimum–maximum), with evaluable n. Percentages use available-data denominators; Not available rows use the full column cohort.
Three patients with unknown pathological response are included in Overall only. Cycle summaries exclude ongoing treatments and uncertain or missing counts.
pCR, pathological complete response; RCB, residual cancer burden; scRNA-seq, single-cell RNA sequencing.

Baseline PBMC samples from 22 patients were included in the single-cell RNA-sequencing analysis. The flow-cytometry dataset comprised 125 primary PBMC specimens from 64 patients also with early TNBC treated with chemoimmunotherapy, including 55 samples collected before treatment (T1), 40 collected after completion of the first therapeutic block, 3 months after treatment initiation (T2), and 30 collected after completion of neoadjuvant treatment but before surgery (T3). Of the 64 patients, 36 achieved pCR, 25 had residual disease and three had unknown pathological response. Two of these patients were still receiving neoadjuvant treatment at the time of analysis, and the third had transferred to another hospital before pathological-response data became available. The four specimens from patients with unknown response were retained in the dataset for response-independent analyses but excluded from pCR versus non-pCR comparisons. Twenty-two patients underwent both baseline scRNA-seq and flow cytometry, whereas 42 underwent flow cytometry alone (**Supplementary Fig. 1**).

Pathological response was assessed in the surgical specimen after completion of neoadjuvant treatment. pCR was defined as the absence of residual invasive carcinoma in the breast and sampled regional lymph nodes, corresponding to residual cancer burden (RCB) class 0. Patients with residual invasive disease, corresponding to RCB classes I–III, were classified as non-pCR.

### PBMCs isolation and cryopreservation

Peripheral blood was collected in a minimum of three and up to five 10-mL cell-preserving CellSave tubes (Menarini) and maintained at room temperature until processing, which was performed up to 72 hours after collection. Whole-blood samples were centrifuged at 596 × *g* for 10 min at 20°C. After plasma removal, the buffy-coat fraction was collected, diluted 1:1 with Dulbecco’s phosphate-buffered saline (DPBS; Sigma-Aldrich), and carefully layered over Lymphoprep (STEMCELL Technologies) without disturbing the density gradient. Samples were centrifuged at 840 × *g* for 30 min at 4°C with the centrifuge brake disabled. The PBMCs-containing interface was recovered, transferred to a fresh tube, brought to a final volume of 15 mL with DPBS, and centrifuged at 1,341 × *g* for 8 min at 20°C with the brake enabled. The cell pellet was resuspended in 1–5 mL of DPBS, and cell concentration and viability were assessed by Trypan Blue exclusion using a CellDrop automated cell counter (DeNovix). PBMCs were then subjected to a second wash with 15 mL of DPBS under the same centrifugation conditions.

PBMCs were cryopreserved in heat-inactivated fetal bovine serum containing 10% dimethyl sulfoxide. Cells were kept in a freezing container at −80°C for up to 4 days before storage in liquid nitrogen. Post-thaw viability was assessed before scRNA-seq or flow cytometry.

### Single cell RNA sequencing and analysis

Baseline PBMC samples from 27 patients were initially considered for single-cell RNA sequencing. After thawing, dead cells were depleted with the MACS Dead Cell Removal Kit (Miltenyi Biotec), and suspensions with post-depletion viability greater than 80% were processed. Single-cell 5′ gene-expression libraries were prepared with the Chromium Next GEM Single Cell 5′ Kit v2, Dual Index (10x Genomics), targeting recovery of up to 25,000 cells per sample, and sequenced on an Illumina NovaSeq X Plus using the recommended paired-end dual-index configuration. FASTQ files were aligned to GRCh38 and filtered gene-by-cell matrices were generated with Cell Ranger 9.0.1.

Five samples failed sample-level quality control, leaving 22 patients (14 pCR, 8 non-pCR). Analyses used Scanpy 1.12.1. Highly variable genes were selected from raw counts using Seurat v3. The full matrix was normalized to 10,000 counts per cell and log1p-transformed. The scaled highly variable-gene matrix underwent principal-component analysis, with the first 30 components used to construct a 15-nearest-neighbor graph for UMAP and Leiden clustering.

Atlas identities were assigned using canonical lineage markers. T/NK, conventional T-cell and B-cell subsets were independently extracted from the atlas and reclustered at Leiden resolutions 0.4, 0.3 and 0.3, respectively. Patient-level analyses quantified subcluster abundance relative to the parent compartment and summarized functional-program scores and gene-level metrics within eligible T- and B-cell subclusters.

### Flow cytometry and analytical quality control

Multiparameter flow cytometry was performed on thawed unfixed T1, T2 and T3 PBMCs using two surface-staining panels. Cells were stained for 20–30 min at 4°C and analyzed on a CytoFLEX S V4-B4-R3-I2 flow cytometer (Beckman Coulter), with data processed using FlowJo v10. Panel 1 characterized CD4+ and CD8+ T-cell differentiation and checkpoint phenotypes, whereas Panel 2 characterized B-cell, NK-cell, and dendritic-cell-enriched populations.

For focused CD38 analyses, CD38-positive and CD38-negative branches were evaluated within naive, unswitched-memory, switched-memory, and double-negative B-cell populations. CD38 compositional balance was calculated as:

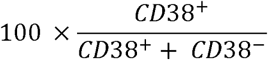

### Statistical analysis

Analyses were performed at the patient level. Continuous clinicopathological variables were summarized as medians (minimum–maximum), whereas flow-cytometry measurements were summarized as medians and interquartile ranges. Cross-sectional comparisons between pCR and non-pCR used two-sided Mann-Whitney U tests, calculated exactly when no ties were present and with the asymptotic tie correction otherwise. For flow-cytometry endpoints, Cliff’s delta was reported with positive values indicating higher values in pCR. Receiver operating characteristic curves treated pCR as the positive outcome, and AUC was derived from the Mann-Whitney statistic. Analyses required at least five evaluable patients in each response group.

Paired changes between T1, T2, and T3 were evaluated with two-sided Wilcoxon signed-rank tests and matched rank-biserial correlations. Response-group differences in within-patient change were evaluated from later-minus-earlier change scores using Mann-Whitney U tests and Cliff’s delta. Benjamini-Hochberg correction was applied within the prespecified scRNA-seq and flow-cytometry analysis families; flow-cytometry families were defined by panel and timepoint or longitudinal interval.

The primary switched-memory CD38 balance at T3, 95% confidence intervals for Cliff’s delta and AUC were estimated by 10,000 stratified patient-level bootstrap resamples. Robustness was assessed by leave-one-patient-out analysis, recalculating the Mann-Whitney test, Cliff’s delta, and AUC after removing each patient in turn. Tests were two-sided; P<0.05 denoted nominal significance, and q<0.05 denoted significance after FDR correction. Pseudo-bulk differential-expression and gene-set analyses were performed in R 4.5.3 using edgeR 4.8.2 and limma 3.66.0. Python-based analyses used Python 3.13.9, SciPy 1.16.3, pandas 2.3.3, statsmodels 0.14.5, and scikit-learn 1.7.

## RESULTS

### Global PBMCs and T/NK profiling reveals broadly shared immune architecture across response groups

Baseline scRNA-seq of PBMCs from 22 patients receiving pembrolizumab-based neoadjuvant chemoimmunotherapy (14 pCR and 8 non-pCR; **Fig. 1a**) yielded 135,271 quality-controlled cells (**Supplementary Fig. 2**). Response-blind clustering at Leiden resolution 0.5 identified 18 transcriptional clusters, consolidated into 11 major populations within a predominantly lymphoid atlas (**Fig. 1b**). Cells from both response groups were broadly intermingled across the major immune compartments, without global response-specific segregation (**Fig. 1c**). Canonical lineage-marker expression supported the annotated population identities (**Fig. 1d**).

**Figure 1.**
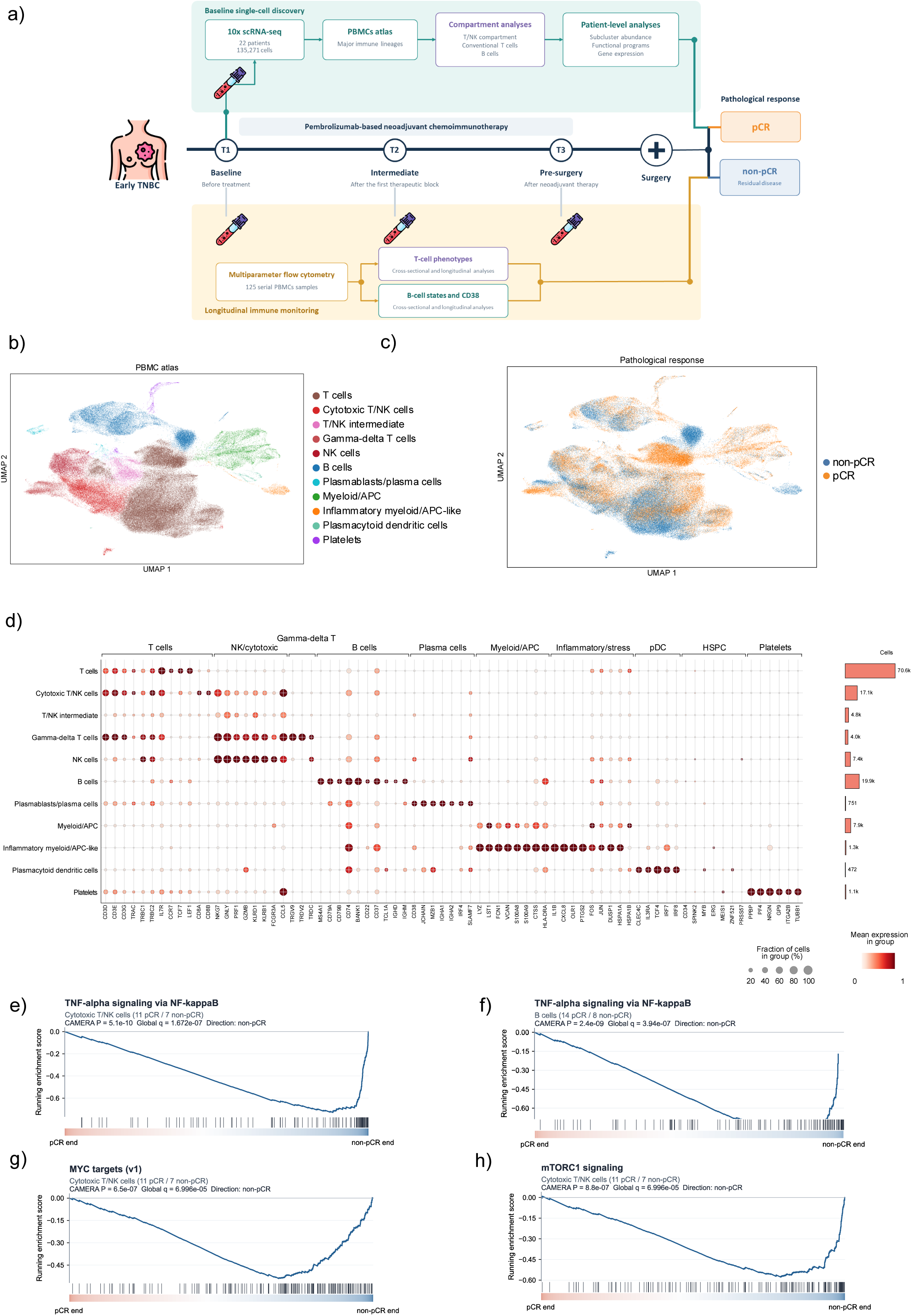
Study design and baseline single-cell characterization of the circulating immune landscape. **(a)** Schematic representation of the study design, treatment course, blood-sampling timepoints and immune-profiling strategy. Baseline PBMC samples from 22 patients treated with pembrolizumab-based neoadjuvant chemoimmunotherapy were analyzed by single-cell RNA sequencing, generating 135,271 quality-controlled cells. Multiparametric flow cytometry was used for longitudinal immune profiling of 125 PBMC specimens from 64 patients, including 55 collected at baseline, 40 after the first therapeutic block and 30 after completion of neoadjuvant therapy but before surgery. **(b)** UMAP representation of the baseline PBMCs atlas showing the 11 major annotated immune populations. **(c)** Global PBMCs UMAP colored according to pathological response. **(d)** Dot plot showing canonical lineage-marker expression across the annotated PBMC populations. **(e–h)** Descriptive enrichment curves for selected Hallmark gene sets: **(e)** TNF-α signaling via NF-κB in cytotoxic T/NK cells; **(f)** the same pathway in B cells; and **(g)** MYC targets (v1) and **(h)** mTORC1 signaling in cytotoxic T/NK cells. Genes were ranked using patient-level pseudo-bulk differential-expression statistics. Displayed P values were obtained from CAMERA; global q values were calculated using Benjamini–Hochberg correction across all tested pathway–population combinations.

Patient-level pseudo-bulk pathway analysis identified compartment-specific transcriptional differences associated with pathological response at baseline. TNF-α signaling via NF-κB was enriched in non-pCR patients within both cytotoxic T/NK cells (global q=1.67×10⁻□; **Fig. 1e**) and B cells (global q=3.94×10⁻□; **Fig. 1f**). Cytotoxic T/NK cells from non-pCR patients also showed enrichment of MYC targets (v1) and mTORC1 signaling (both global q=7.00×10⁻□; **Fig. 1 g,h**).

In addition, the pathway map showed broader non-pCR-oriented patterns involving MYC targets, mTORC1 signaling and oxidative phosphorylation across several lymphoid populations. Conversely, inflammatory programs in myeloid/APC cells were preferentially associated with pCR, including IL-6/JAK–STAT3 signaling (p=4.1×10⁻□) and inflammatory response (p=6.3×10⁻□) (**Supplementary Fig. 3**).

To examine heterogeneity within the dominant lymphoid compartment, we reconstructed a combined T/NK object comprising 103,935 atlas-defined T, cytotoxic T/NK, NK, T/NK-intermediate and γδ T cells. Reclustering at Leiden resolution 0.4 resolved 12 subclusters, C0–C11, spanning conventional T-cell, cytotoxic, NK, γδ T-cell and intermediate transcriptional states. Cells from both response groups remained broadly intermingled on the T/NK UMAP (**Supplementary Fig. 4)**. Subclusters C3 and C11 were excluded from abundance inference because they were predominantly contributed by individual patients. None of the remaining ten T/NK subclusters differed significantly in patient-level abundance between response groups (**Supplementary Fig. 5, Supplementary Table 1**). We therefore restricted subsequent analyses to atlas-defined conventional T cells to examine T-cell-intrinsic heterogeneity without conflating transcriptionally related NK and cytotoxic populations.

### Baseline T-cell profiling identifies preserved naïve/memory features and a pCR-associated CX3CR1 signal

The single-cell RNA sequencing lineage-restricted T-cell object comprised 70,567 cells and resolved seven subclusters at Leiden resolution 0.3, C0–C6 (**Fig. 2a**). Marker expression supported their annotation as C0, CD4 IL7R naïve/memory-like; C1, CD4 memory/activation-like; C2, preserved/quiescent-like; C3, heat-shock/stress-activated; C4, CD8 naïve/memory-like; C5, preserved naïve/memory-like; and C6, cytotoxic/stress-like (**Fig. 2b,c**). Cells from pCR and non-pCR patients remained broadly distributed across the UMAP without global response-specific segregation (**Fig. 2d**).

**Figure 2.**
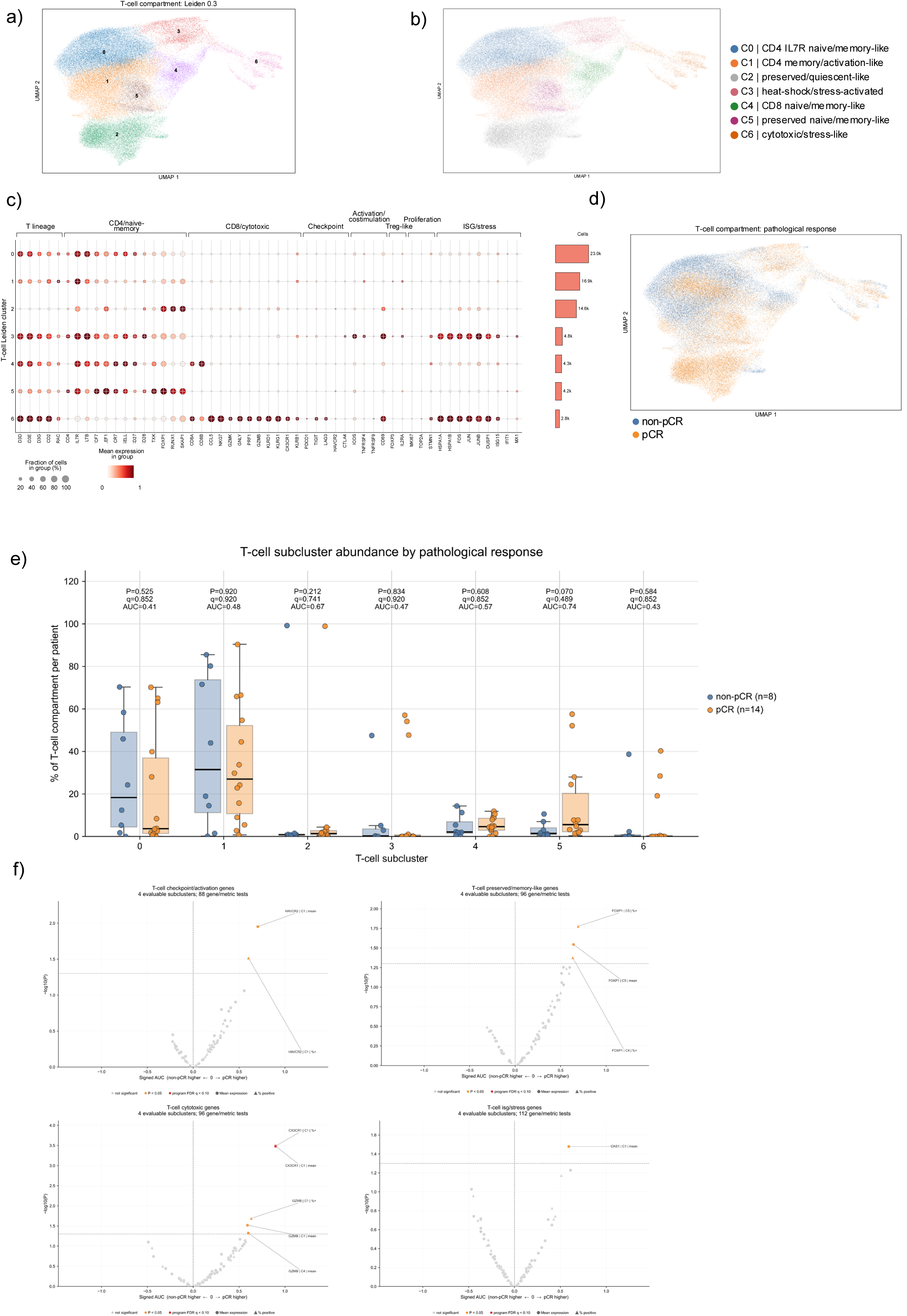
Lineage-restricted T-cell analysis identifies a pCR-associated *CX3CR1* signal within CD4 memory/activation-like cells. **(a)** UMAP representation of the independently reclustered T-cell compartment at Leiden resolution 0.3, showing seven subclusters, C0–C6. **(b)** T-cell UMAP showing the provisional functional annotation of the subclusters. **(c)** Dot plot showing genes associated with T-cell lineage, CD4/naïve-memory, CD8/cytotoxic, checkpoint/activation, proliferation and ISG/stress programs. **(d)** T-cell UMAP colored according to pathological response. **(e)** Patient-level relative abundance of each T-cell subcluster in pCR and non-pCR patients. For each patient, abundance was calculated as the percentage of the total T-cell compartment assigned to the corresponding subcluster. P values were obtained using two-sided Mann– Whitney U tests and q values by Benjamini–Hochberg correction across the seven T-cell subclusters. **(f)** Program-wide gene-level analyses for the checkpoint/activation, preserved/memory-like, cytotoxic and ISG/stress programs across the four evaluable subclusters, C0, C1, C4 and C5. Each point represents a gene–subcluster comparison using patient-level values. Circles indicate mean log-normalized expression and triangles indicate the percentage of expressing cells. Signed AUC was calculated as 2 × AUC − 1, with positive values indicating higher values in pCR and negative values indicating higher values in non-pCR. The y-axis shows −log10(P), and horizontal dashed lines denote nominal P=0.05. Red symbols highlight associations with program-level q<0.10, orange symbols indicate P<0.05 with q≥0.10, and grey symbols indicate the remaining comparisons. Benjamini–Hochberg correction was applied across all genes, evaluable subclusters and both expression metrics within each functional program.

Patient-level subcluster abundances were compared between response groups using the complete T-cell compartment from each patient as the denominator. No T-cell subcluster remained significantly associated with pathological response (**Fig. 2e, Supplementary Table 2**). The largest difference was observed for C5 preserved naïve/memory-like cells, which were more abundant in pCR patients (median, 5.60% versus 1.44%; P=0.0698; q=0.489; AUC=0.741; Cliff’s δ=0.482).

Four biologically defined transcriptional programs, preserved/memory-like, cytotoxic, checkpoint/activation and ISG/stress, were subsequently evaluated using patient-level mean scores. C2 was excluded from these analyses because a single patient contributed 90.4% of its cells, whereas C3 and C6 lacked sufficient evaluable patients in at least one response group. Consequently, C0, C1, C4 and C5 were eligible for functional-program comparisons. None of the four programs differed significantly between response groups. The strongest program-level trend was a higher preserved/memory-like score in pCR within C4 CD8 naïve/memory-like cells (median, 1.260 versus 1.081; P=0.0727; q=0.291; AUC=0.783; Cliff’s δ=0.567; **Supplementary Fig. 6, Supplementary Table 3**).

Gene-level analysis identified a response-associated *CX3CR1* signal within the CD4 memory/activation-like subcluster (**Fig. 2f, Supplementary Table 4**). Although *CX3CR1* transcripts were detected in a small fraction of cells, the median percentage of positive cells was higher in pCR than in non-pCR patients (0.96% versus 0.24%; P=0.00033; gene-family FDR q=0.0157; AUC=0.949; Cliff’s δ=0.898). Other nominal signals within this subcluster also involved relatively small cellular fractions, including *HAVCR2* (*TIM3*) (1.31% versus 0.60%; P=0.030) and *GZMB* (0.40% versus 0%; P=0.020). In contrast, *FOXP1* represented a more broadly expressed preserved/memory-associated signal, where the percentage of positive cells was higher in pCR within both the CD8 naïve/memory-like subcluster (79.92% versus 63.28%; P=0.042) and the preserved naïve/memory-like subcluster (94.87% versus 90.71%; P=0.017).

### Baseline B-cell profiling reveals a pCR-associated naïve-like transcriptional program

The single-cell RNA sequencing B-cell compartment comprised 20,608 cells annotated in the global atlas as conventional B cells or plasmablasts/plasma cells. B-cell-specific reclustering at Leiden resolution 0.3 identified 11 subclusters, C0–C10 (**Fig. 3a**). Source-identity projection and marker expression revealed conventional B-cell, naïve-like, memory-like, activated/APC-like, proliferative and plasma-cell-associated regions (**Fig. 3b,c**). Cells from pCR and non-pCR patients remained broadly intermingled on the UMAP (**Fig. 3d**).

**Figure 3.**
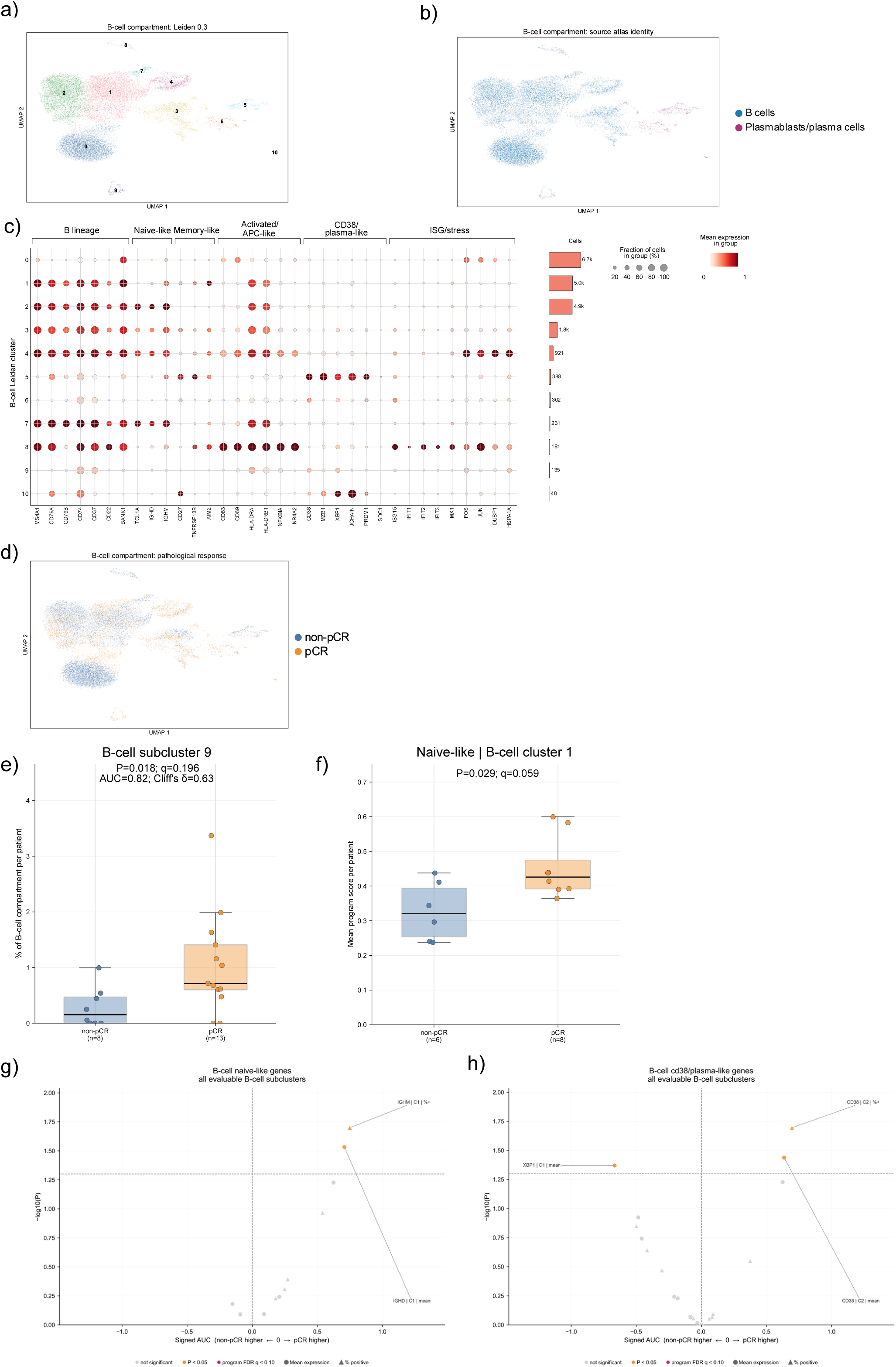
Baseline B-cell states show exploratory associations with pathological response. **(a)** UMAP representation of the independently reclustered B-cell compartment at Leiden resolution 0.3, showing 11 subclusters, C0–C10. **(b)** B-cell UMAP colored according to source atlas identity as conventional B cells or plasmablasts/plasma cells. **(c)** Dot plot showing B-lineage genes and genes associated with naïve-like, memory-like, activated/APC-like, CD38/plasma-like and ISG/stress programs across B-cell subclusters. **(d)** B-cell UMAP colored according to pathological response. **(e)** Patient-level abundance of C9 as a percentage of the total B-cell compartment. **(f)** Patient-level naïve-like program score within C1. The score was defined as the mean log-normalized expression of *TCL1A*, *IGHD* and *IGHM* across all C1 cells contributed by each patient. **(g,h)** Program-wide gene-level analyses for **(g)** the naïve-like and **(h)** CD38/plasma-like programs across the two evaluable subclusters, C1 and C2. Each point represents a patient-level gene–subcluster comparison based on either mean log-normalized expression or the percentage of positive cells. Signed AUC values greater than zero indicate higher values in pCR and negative values indicate higher values in non-pCR. Horizontal lines denote nominal P=0.05. Groups were compared using two-sided Mann–Whitney U tests. Program-score q values were calculated across evaluable subclusters within each program, and gene-level q values across all genes, evaluable subclusters and both expression metrics within the corresponding functional program. Individual points in box plots represent patients; boxes show the median and interquartile range. AUC, area under the receiver operating characteristic curve; pCR, pathological complete response

Quality-control review identified C0, C8 and C10 as sample-dominated and C3 as a mixed B/T-lineage cluster. These four clusters were retained in descriptive visualizations but excluded from response inference. One patient contributed fewer than 50 B-compartment cells and was excluded from patient-level response comparisons, leaving 13 pCR and 8 non-pCR patients for the abundance analysis. Among the seven evaluable B-cell subclusters, the small C9 population was more abundant in pCR patients than in non-pCR patients (median, 0.716% versus 0.152%; P=0.0178; AUC=0.817; Cliff’s δ=0.635; **Fig. 3e, Supplementary Table 5**).

Functional-program analyses were restricted to C1 and C2, the only QC-valid subclusters that met the minimum cell-count and group-size requirements for these comparisons. Within C1, the naïve-like program, defined by *TCL1A*, *IGHD* and *IGHM*, was higher in pCR patients (median score, 0.426 versus 0.320; P=0.0293; within-program q=0.0586; AUC=0.854; Cliff’s δ=0.708; n=8 pCR and 6 non-pCR; **Fig. 3f, Supplementary Table 6**). No other functional-program comparison was associated with response.

Gene-level analyses across the two evaluable subclusters identified several nominal associations (**Fig. 3g,h**; **Supplementary Table 7**), but not in the memory-like, activated/APC-like or ISG/stress programs (**Supplementary Fig. 7)**. Within B-cell C1, *IGHM* was detected in a larger proportion of cells from pCR patients than from non-pCR patients (median, 46.26% versus 32.53%; P=0.020), while *IGHD* showed nominally higher mean expression in pCR (P=0.029), although the difference in the percentage of positive cells was not significant (11.46% versus 7.89%; P=0.108). Within B-cell C2, *CD38* was also detected in a larger cellular fraction in pCR patients (15.56% versus 8.94%; P=0.020). Conversely, *XBP1* showed nominally higher mean expression in non-pCR within C1 (P=0.043) but was detected in only a small proportion of cells (4.35% in non-pCR versus 3.01% in pCR). Nevertheless, their direction was consistent with the higher naïve-like programme score observed in pCR within C1 (P=0.029; q=0.059).

### Neoadjuvant chemoimmunotherapy induces marked longitudinal remodeling of the circulating B-cell compartment

To characterize treatment-associated immune dynamics independently of pathological response, we analyzed paired flow-cytometry samples from the complete cohort across the T1–T2, T2–T3 and T1–T3 intervals (**Supplementary Fig. 8, Supplementary Table 8**).

During the early T1–T2 interval, switched-memory B cells decreased (matched rank-biserial r=−0.76; P=0.00024; q=0.0020), as did unswitched-memory B cells (r=−0.74; P=0.00043; q=0.0024). Total CD3^⁺^ T cells also decreased (r=−0.66; P=0.0019; q=0.0079). Immature dendritic cells declined (r=−0.53; P=0.015; q=0.042), whereas CD83^⁺^HLA-DR^high^ APC-like B cells increased (r=0.52; P=0.017; q=0.042). Several T-cell phenotypes showed nominal changes, but none remained significant after FDR correction.

The most extensive remodeling occurred between T2 and T3. Naive B cells showed the largest decline (r=−0.92; P<0.0001; q=0.00021). CD20^⁺^CD38^⁺^ mature B cells also decreased (r=−0.79; P=0.00041; q=0.0022), whereas plasmablasts increased with a comparable effect magnitude (r=0.79; P=0.00041; q=0.0022). Double-negative B cells increased (r=0.77; P=0.00064; q=0.0026), while unswitched-memory B cells (r=−0.72; P=0.0025; q=0.0062), total B cells (r=−0.69; P=0.0027; q=0.0062) and CD20^⁺^CD38⁻ memory B cells (r=−0.60; P=0.010; q=0.020) decreased.

Consistent changes were observed across the complete T1–T3 interval. CD20^⁺^CD38^⁺^ mature B cells (r=−0.85; P=0.00042; q=0.0053), total B cells (r=−0.82; P=0.00079; q=0.0053), unswitched-memory B cells (r=−0.91; P=0.0010; q=0.0053), naive B cells (r=−0.82; P=0.0021; q=0.0065) and CD20^⁺^CD38⁻ memory B cells (r=−0.76; P=0.0024; q=0.0065) decreased. Conversely, plasmablasts and double-negative B cells increased (both r=0.75; P=0.0028; q=0.0065). Overall, these findings demonstrate broad treatment-associated remodeling of the circulating B-cell compartment, characterized by contraction of conventional B-cell populations and relative enrichment of plasmablast and double-negative states.

### Cross-sectional screening identifies response-associated immune phenotypes at specific treatment timepoints

To identify circulating immune phenotypes associated with pathological response, we compared pCR and non-pCR patients independently at baseline, the intermediate treatment timepoint and pre-surgery samples (**Supplementary Fig. 9, Supplementary Table 9**).

At baseline, CD4^⁺^TIGIT^⁺^ T cells were more abundant in pCR patients than in non-pCR patients (median, 18.95% versus 11.10%; Cliff’s δ=0.390; P=0.0459, **Supplementary Fig. 9**). At the intermediate treatment timepoint, no population reached nominal statistical significance. The lowest P value was observed for CD4^⁺^CD161^⁺^ T cells, which showed numerically higher frequencies in pCR patients (median, 10.40% versus 7.25%; n=21 versus 17; Cliff’s δ=0.311; P=0.1063; q=0.9682, **Supplementary Fig. 9**).

The strongest response-associated differences emerged at pre-surgery and predominantly involved the B-cell compartment. CD20^⁺^CD38^⁻^ memory B cells were less abundant in pCR patients (median, 8.46% versus 30.20%; Cliff’s δ=−0.596; P=0.0070; q=0.0667), whereas plasmablasts were more abundant (median, 69.05% versus 41.70%; Cliff’s δ=0.556; P=0.0125; q=0.0667). Unswitched-memory B cells (median, 0.15% versus 1.02%; Cliff’s δ=−0.465; P=0.0388; q=0.124) and CD20^⁺^CD38^⁺^ mature B cells (median, 6.48% versus 20.50%; Cliff’s δ=−0.444; P=0.0494; q=0.132) were also nominally lower in pCR patients (**Supplementary Fig. 9)**.

### Longitudinal analysis identifies response-associated patterns in B-cell remodeling

To determine whether treatment-associated immune remodeling differed according to pathological response, within-patient changes were calculated for each interval and compared directly between pCR and non-pCR patients (**Fig. 4a–c**; **Supplementary Fig. 10, Supplementary Table 10**).

**Figure 4.**
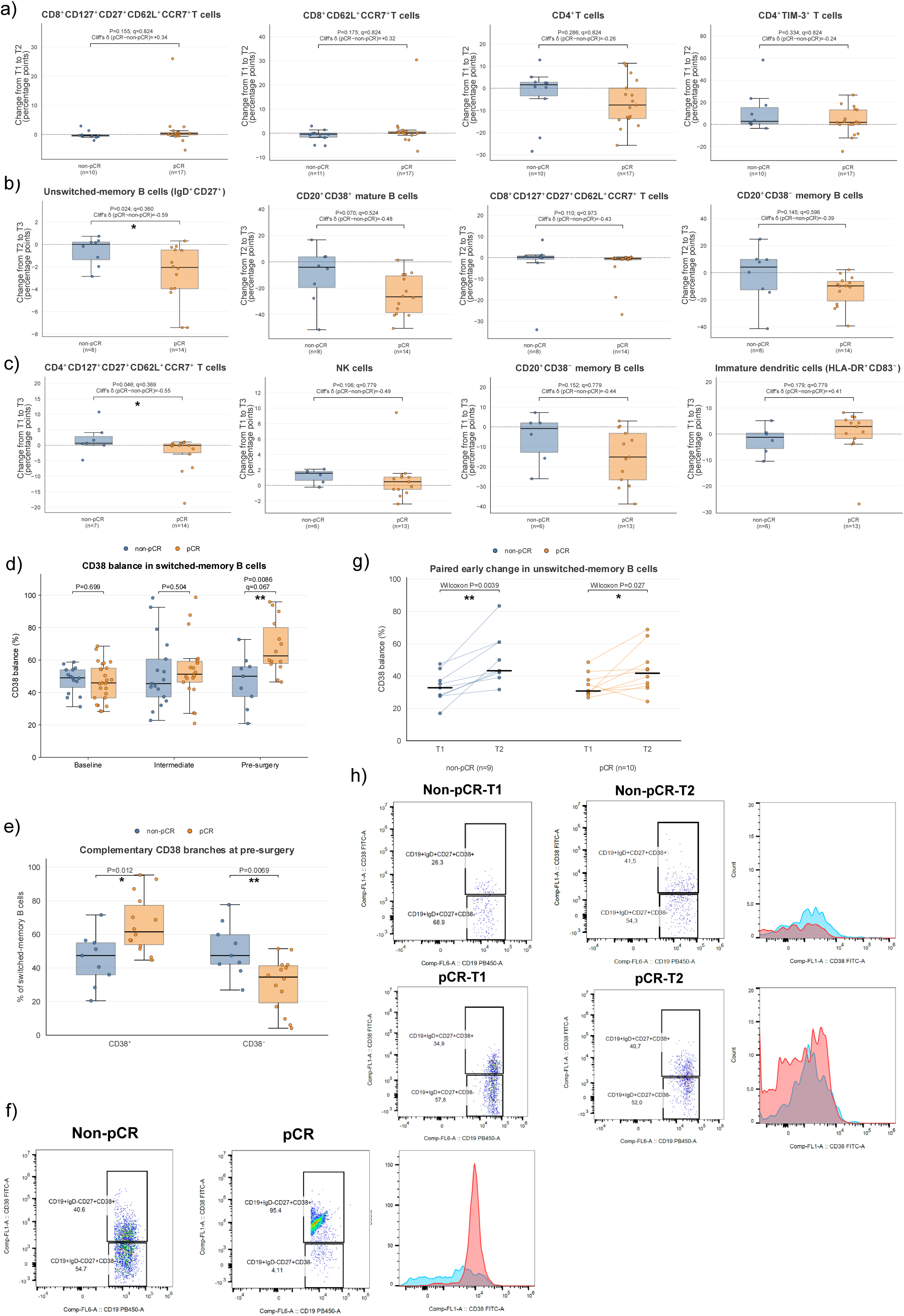
Longitudinal immune remodeling and focused analysis of CD38-defined B-cell states. (a–c) The four populations with the lowest nominal P values for between-group differences in within-patient change during **(a)** T1–T2, **(b)** T2–T3 and **(c)** T1–T3. Changes were calculated as the later minus the earlier value and expressed in percentage points. **(d)** CD38 balance within switched-memory B cells at baseline, the intermediate treatment timepoint and pre-surgery. CD38 balance was calculated as 100 × CD38^⁺^/(CD38^⁺^ + CD38^⁻^). **(e)** Complementary CD38^⁺^ and CD38^⁻^ switched-memory B-cell fractions at pre-surgery. **(f)** Representative switched-memory B-cell CD38 plots at pre-surgery from one non-pCR and one pCR patient, with an overlaid histogram of CD38 fluorescence intensity (blue, non-pCR; red, pCR). **(g)** Paired T1–T2 change in unswitched-memory B-cell CD38 balance, shown separately by response group. Lines connect samples from the same patient. **(h)** Representative paired unswitched-memory B-cell CD38 plots at T1 and T2 from one non-pCR patient (upper row) and one pCR patient (lower row). Overlaid histograms show CD38 fluorescence intensity at T1 (red) and T2 (blue) for each patient. Numbers in the flow-cytometry plots indicate gated percentages within the corresponding parent population. Between-group comparisons of longitudinal change in panels a–c and cross-sectional comparisons in panels d–e used two-sided Mann–Whitney U tests. Within-group paired changes in panel g used two-sided Wilcoxon signed-rank tests. For panels a–c, negative Cliff’s delta values indicate a lower change score in pCR than in non-pCR. Individual points represent patients; boxes show the median and interquartile range. *P<0.05; exact FDR-adjusted q values are shown where applicable.

During the early T1–T2 interval, none of the evaluated immune populations showed a nominally significant difference in change between response groups (**Fig. 4a**). This indicates that the early treatment-associated remodeling observed in the complete cohort was broadly comparable between pCR and non-pCR patients.

The strongest response-associated difference emerged between T2 and T3. Unswitched-memory B cells decreased more markedly in pCR patients than in patients with residual disease (median change, −2.06 versus 0.00 percentage points; Cliff’s δ=−0.589; P=0.0240; q=0.360; **Fig. 4b**). CD20^⁺^CD38^⁺^ mature B cells showed a similar tendency towards a greater decline in pCR patients (median change, −26.60 versus −4.13 percentage points; Cliff’s δ=−0.482; P=0.0698; q=0.524).

Across the complete T1–T3 interval, CD4^⁺^CD127^⁺^CD27^⁺^CD62L^⁺^CCR7^⁺^ T cells showed a nominally different change between response groups. This population remained relatively stable in pCR patients but increased in non-pCR patients (median change, −0.08 versus 0.69 percentage points; Cliff’s δ=−0.551; P=0.0461; q=0.369; **Fig. 4c**).

Overall, the most prominent response-associated longitudinal pattern involved late remodeling of the B-cell compartment, accompanied by an exploratory T-cell difference across the complete treatment interval.

### Focused CD38 analysis identifies a pre-surgery response-associated pattern in switched-memory B cells

The exploratory baseline scRNA-seq signal of higher *CD38* expression in B-cell subcluster C2 in pCR patients, together with the pre-surgery B-cell differences detected by flow cytometry, prompted a focused analysis of CD38-defined B-cell states. No differences in switched-memory B-cell CD38 balance were detected between response groups at baseline or at the intermediate treatment timepoint. At pre-surgery, CD38 balance was higher in pCR than in non-pCR patients (median, 62.59% versus 50.00%; n=14 versus 9; Cliff’s δ=0.651; P=0.0086; q=0.0667; **Fig. 4d**).

Analysis of the complementary branches showed a higher frequency of CD38^⁺^ switched-memory B cells in pCR patients (median, 61.55% versus 47.40%; Cliff’s δ=0.643; P=0.0117) and a lower frequency of the corresponding CD38⁻ branch (median, 34.60% versus 47.40%; Cliff’s δ=−0.667; P=0.0069; **Fig. 4e**). Representative flow-cytometry plots illustrate the contrasting CD38 distributions observed in non-pCR and pCR patients at pre-surgery (**Fig. 4f**).

CD38 balance within unswitched-memory B cells increased from T1 to T2 in both response groups. The median increased from 32.78% to 43.32% in non-pCR patients (n=9; paired Wilcoxon P=0.0039) and from 30.77% to 41.77% in pCR patients (n=10; P=0.027; **Fig. 4g**). The median change was larger in non-pCR than in pCR patients (13.94 versus 6.82 percentage points). Representative paired plots illustrate the early increase in unswitched-memory CD38 balance (**Fig. 4h**).

The pre-surgery switched-memory CD38 association remained stable in sensitivity analyses (**Supplementary Fig. 11**). Stratified patient-level bootstrap analysis yielded a 95% confidence interval of 0.254–0.937 for Cliff’s δ and 0.627–0.968 for the AUC. In leave-one-patient-out analyses, Cliff’s δ ranged from 0.607 to 0.768 and the AUC from 0.804 to 0.884. All 23 leave-one-patient-out comparisons remained nominally significant, with a maximum P value of 0.0197. Thus, the pre-surgery association remained detectable after exclusion of any individual patient.

## DISCUSSION

In this study, we integrated baseline single-cell transcriptomics with longitudinal flow cytometry to identify circulating immune states associated with pathological response to pembrolizumab-based neoadjuvant chemoimmunotherapy. Baseline associations involved specific transcriptional programs and cellular states, while longitudinal profiling revealed extensive B-cell remodeling, with response-associated differences most evident in CD38-defined switched-memory states before surgery.

Non-pCR-associated TNF-α/NF-κB and MYC/mTORC1 enrichment in lymphoid cells may reflect baseline inflammatory and metabolic stimulation. TNF-mediated immunosuppression and mTORC1 regulation of T-cell persistence provide biological parallels ^12,13^. Conversely, inflammatory programs in myeloid/APC cells were oriented towards pCR, suggesting that the relationship between baseline immune activation and pathological response depends on the cellular compartment.

At baseline, pCR was associated with a *CX3CR1* signal within CD4 memory/activation-like cells, alongside exploratory trends towards preserved naïve/memory-like T-cell states. Previous studies linked circulating CX3CR1^⁺^ CD8^⁺^ T cells to checkpoint-blockade response and persistence during chemoimmunotherapy ^14,15^ , although the functional significance of our CD4-enriched signal remains unresolved. In other T-cell subclusters, nominally higher *FOXP1*-positive fractions in pCR were consistent with the preserved-state interpretation, given the role of *FOXP1* in maintaining naïve T-cell quiescence ^16^.

Exploratory findings within B-cell subcluster C1 suggested a more naïve/unswitched-like configuration in pCR and higher *XBP1* expression in non-pCR. At the broader B-cell level, enrichment of the unfolded protein response, MYC targets and mTORC1 signaling in non-pCR patients was consistent with increased secretory adaptation and biosynthetic activity. Given the role of *XBP1* in linking the unfolded protein response to plasma-cell development ^17,18^, these findings raise the possibility of a greater baseline tendency towards antibody-secreting differentiation in non-pCR.

Flow cytometry showed broad treatment-associated B-cell remodeling, with early decreases in memory B-cell frequencies and later redistribution towards plasmablast and double-negative compartments. Response-associated differences were more restricted. These dynamics are consistent with Barth et al., who linked on-treatment peripheral B-cell phenotypes to checkpoint-inhibitor response ^10^.

The clearest response-associated flow-cytometric finding was a higher pre-surgery CD38 balance within switched-memory B cells in pCR, which remained stable in leave-one-patient-out analyses. During the early interval, unswitched-memory CD38 balance increased in both groups, with a larger median change in non-pCR. This early pattern, together with the late switched-memory association in pCR, suggests that CD38 remodeling may have different response associations depending on B-cell subset and treatment phase. Related evidence links CD38-high plasmablast-like B cells in melanoma lymph nodes to favorable outcomes following adjuvant PD-1 blockade. These findings support further investigation of CD38-defined B-cell states ^19^.

Several limitations should be considered. The cohort was modest and imbalanced for pathological response and longitudinal sample availability varied. The scRNA-seq and flow-cytometry measurements also differed in cellular resolution, marker definition and sampling time. Although flow cytometry incorporated IgD/CD27-based B-cell classification, IgM protein expression, B-cell-receptor repertoire and antibody specificity were not evaluated. Finally, the study design cannot separate immune changes attributable to chemotherapy from those specifically caused by pembrolizumab, and peripheral-blood measurements may not directly reproduce immune changes occurring within the tumor microenvironment.

Overall, these exploratory findings link pathological response to baseline transcriptional states and CD38-defined B-cell patterns that vary across subsets and treatment phases. Prospective validation is required to establish the reproducibility and clinical relevance of these associations.

## Data Availability

The raw data supporting the findings of this study have been deposited in Zenodo under DOI: 10.5281/zenodo.23947326

https://doi.org/10.5281/zenodo.23947326

## DECLARATIONS

### Ethics approval and consent to participate

The study was approved by the Provincial Research Ethics Committee of Málaga and the Santiago-Lugo Research Ethics Committee (CEI-SL; approval number 2024/036) and was conducted in accordance with the Declaration of Helsinki. All participants provided written informed consent for the collection and molecular analysis of their biological samples.

## Consent for publication

Not applicable

## Authors’ contributions

**Resources:** JP, CC, RP, RLL, EA, AGO, AG, MED, NR, BP, AMA, BJ, PC and PPO contributed to patient recruitment and sample acquisition. **Investigation:** MRCP and MEQO performed the experimental work, with support from JVS. MVOJ and MA contributed to the pathological assessment. **Methodology:** ICM developed bioinformatic and statistical analysis strategies, and PCG contributed expertise in flow cytometry. **Data curation:** AGO extracted clinical data, and ICM prepared and curated the analytical datasets. **Formal analysis:** ICM led and performed the majority of the bioinformatic and statistical analyses, with bioinformatic support from ELL. **Visualization:** ICM. **Funding acquisition:** EdA, EA and RLL. **Writing – original draft:** ICM. **Writing – review & editing:** MRCP, JP and EA.

## ACKNOWLEDGEMENTS

The authors sincerely thank all the patients who participated in this study and donated biological samples. ICM acknowledges support from the Ramón y Cajal program through grant RYC2023-044581-I, funded by MICIU/AEI/10.13039/501100011033 and the European Social Fund Plus (ESF+). CC was supported by ISCIII (CP23/00002).

## LIST OF ABBREVIATIONS

7-AAD: 7-aminoactinomycin D.
APC: antigen-presenting cell.
AUC: area under the receiver operating characteristic curve.
DMSO: dimethyl sulfoxide.
DPBS: Dulbecco’s phosphate-buffered saline.
FACS: flow cytometry.
FBS: fetal bovine serum.
FDR: false discovery rate.
FITC: fluorescein isothiocyanate.
GEM: gel bead-in-emulsion.
GRCh38: Genome Reference Consortium human genome assembly 38.
ISG: interferon-stimulated gene.
MACS: magnetic-activated cell sorting.
NK: natural killer.
PBMCs: peripheral blood mononuclear cells.
PE: phycoerythrin.
scRNA-seq: single-cell RNA sequencing.
UMAP: uniform manifold approximation and projection.
UMI: unique molecular identifier.

