## Supplementary Methods for "CIRCULATING IMMUNE DYNAMICS DURING NEOADJUVANT CHEMOIMMUNOTHERAPY IN EARLY TRIPLE-NEGATIVE BREAST CANCER"

The sections below provide additional technical information for the single-cell RNA-sequencing, flow-cytometry, and statistical procedures summarized in the main Methods.

**Additional single-cell RNA sequencing details**

Following the dead-cell depletion and viability-selection procedure, the recovered cells were washed, resuspended in phosphate-buffered saline containing 0.04% bovine serum albumin, and counted. Cell suspensions were loaded onto Chromium Next GEM Chip K cartridges. Gel-bead-in-emulsion generation, cellular barcoding, reverse transcription, cDNA amplification, and 5′ gene-expression library construction followed the manufacturer’s workflow. Sequencing used 26 cycles for Read 1, 10 cycles for each index read, and 90 cycles for Read 2, with a target depth of at least 20,000 read pairs per cell. Raw data were demultiplexed and converted to FASTQ files before the alignment and count-matrix generation summarized in the main text.

**Single-cell quality control and preprocessing**

Cell-level quality control considered the number of detected genes, total transcript counts and mitochondrial-transcript percentage. Cells with fewer than 200 detected genes, fewer than 500 total counts or more than 20% mitochondrial counts were excluded. Ribosomal- and haemoglobin-transcript fractions were calculated for quality-control assessment but were not used as exclusion thresholds. Filtered Cell Ranger count matrices were used and sample-level assessment considered cell recovery, transcriptomic complexity, mitochondrial RNA content and the proportion of cells retained after filtering. After cell- and sample-level quality control, the final baseline scRNA-seq cohort comprised 22 TNBC patients treated with pembrolizumab-based neoadjuvant chemoimmunotherapy, including 14 patients achieving pCR and eight with residual disease, and contained 135,271 quality-controlled cells. No additional cells were excluded during the atlas reconstruction described below.

Single-cell analyses used Scanpy 1.12.1. The gene-by-cell matrix was verified to contain non-negative, integer-like raw counts and was retained in a dedicated AnnData counts layer. Before normalization, 3,000 highly variable genes were selected from raw counts using the Seurat v3 method; the batch-aware LOESS fit used a span of 0.5. Full-gene counts were then normalized to 10,000 counts per cell and log1p-transformed, and the resulting full-gene log-normalized matrix was retained for marker visualization, functional scoring and gene-level analyses. Expression of the selected genes was zero-centered and scaled to unit variance, with scaled values clipped at 10. Principal-component analysis used the ARPACK solver. A 15-nearest-neighbor graph was constructed from the first 30 principal components and used for UMAP embedding. Leiden clustering resolution at 0.5 was used for the primary atlas.

**PBMCs atlas annotation**

Cell identities in the reconstructed PBMCs atlas were assigned manually at cluster level using canonical lineage markers, marker dot plots and genes ranked by one-versus-rest Wilcoxon tests on log-normalized expression. T-cell annotation used *CD3D, CD3E, CD3G, TRAC, TRBC1, TRBC2, IL7R, CCR7, LTB, TCF7* and *LEF1*. Cytotoxic T/NK- and NK-cell annotation used *NKG7, GNLY, PRF1, GZMB, KLRD1, KLRB1* and *FCGR3A*. B-cell annotation used *MS4A1, CD79A, CD79B, BANK1, CD22, CD37, CD74, TCL1A*, *IGHD* and *IGHM*. Plasmablast/plasma-cell annotation used *CD38, MZB1, XBP1, JCHAIN, PRDM1* and *SDC1*, and plasmacytoid dendritic-cell annotation used *GZMB*, *GZMA, IL3RA, CLEC4C, TCF4* and *IRF7*. Myeloid/APC annotation used *LYZ, LST1, S100A8, S100A9, MS4A7, CTSS, HLA-DRA* and *HLA-DRB1*, whereas platelet annotation used *PPBP, PF4, NRGN, GP9, ITGA2B* and *TUBB1*. An inflammatory/stress-associated myeloid/APC cluster co-expressed myeloid/APC markers and *FOS*, *JUN, JUNB, DUSP1, HSPA1A, HSPA1B, ISG15, IFIT1* and *MX1*. The final atlas distinguished 11 major populations: conventional T cells, cytotoxic T/NK cells, T/NK-intermediate cells, gamma-delta T cells, NK cells, B cells, myeloid/APC cells, inflammatory/stress-associated myeloid/APC cells, platelets, plasmablasts/plasma cells and plasmacytoid dendritic cells.

**Pseudobulk differential expression and pathway analysis**

Raw UMI counts were summed separately for each patient and annotated atlas population. Patient–population profiles required at least 30 cells, and populations were eligible for comparison when at least five patients per response group met this threshold. Eligible populations comprised conventional T cells, NK cells, B cells, myeloid/APC cells, cytotoxic T/NK cells and T/NK-intermediate cells.

Lowly expressed genes were removed using edgeR’s filterByExpr function, and library normalization factors were calculated using the trimmed mean of M-values method. Differential expression was assessed using robust negative-binomial quasi-likelihood models, with pathological-response group as the explanatory variable and the contrast oriented as pCR versus non-pCR.

The gene-set panel comprised all 50 Hallmark sets and nine selected immune-related Gene Ontology Biological Process sets from MSigDB release 2025.1.Hs. These covered T-cell receptor signalling, T-cell activation, T-cell differentiation, B-cell activation, natural killer cell-mediated immunity, peptide antigen processing and presentation, phagocytosis, lymphocyte migration and leukocyte chemotaxis. For each population, the gene universe comprised the genes retained by edgeR’s filterByExpr function. Gene sets were intersected with this universe and retained when they contained 15–500 genes. HALLMARK_ANGIOGENESIS and HALLMARK_PANCREAS_BETA_CELLS did not meet the minimum gene-overlap criterion in any population. Consequently, 57 distinct gene sets (48 Hallmark and nine GO) were eligible in at least one of the six populations, yielding 331 evaluable gene-set–population combinations. The inter-gene correlation parameter for CAMERA was fixed at 0.01.

For enrichment-curve visualization, all genes retained after expression filtering were ranked in descending order by sign(log2 fold change) × √F, using the edgeR quasi-likelihood F statistic. Positive values indicated higher expression in pCR. Weighted running-sum curves used the absolute ranking statistic to weight gene-set members, with uniform decrements for non-members. These curves were descriptive; displayed P values and adjusted q values were obtained from CAMERA.

**T/NK subclustering and abundance analysis**

The T/NK compartment was extracted directly from the PBMCs atlas and comprised 103,935 cells annotated as conventional T cells, cytotoxic T/NK cells, T/NK-intermediate cells, gamma-delta T cells or NK cells. A total of 3,000 compartment-specific highly variable genes were selected from raw counts using the Seurat v3 method. Log-normalized expression of the selected genes was scaled with values clipped at 10, followed by principal-component analysis, construction of a 15-nearest-neighbor graph from the first 30 principal components and UMAP embedding. Leiden clustering resolution at 0.4 was used for the primary T/NK analysis.

T/NK subclusters were interpreted using T-lineage genes (*CD3D, CD3E, CD3G, TRAC, TRBC1* and *TRBC2*), naive/memory-associated genes (*IL7R, CCR7, LTB, TCF7, LEF1* and *SELL*), cytotoxic T-cell genes (*CD8A, CD8B, GZMK* and *CCL5*), NK/cytotoxic genes (*NKG7, GNLY, PRF1, GZMB, KLRD1, KLRB1, FCGR3A* and *TYROBP*), checkpoint/activation genes (*PDCD1, TIGIT, HAVCR2, LAG3, CTLA4, ICOS* and *TOX*), proliferation-associated genes (*MKI67, TOP2A, STMN1* and *TYMS*), and interferon/stress-associated genes (*ISG15, IFIT1, IFIT2, IFIT3, MX1, FOS, JUN* and *DUSP1*). For each patient, subcluster abundance was calculated as the percentage of all T/NK cells assigned to that subcluster. All patient-by-subcluster combinations were retained, and an absent subcluster was assigned an abundance of zero. Clusters in which one patient contributed at least 70% of all cells were classified as sample-dominated. C3 and C11 met this criterion and were retained in descriptive visualizations but excluded from response-associated inference. The remaining ten subclusters were compared between pCR and non-pCR using two-sided Mann-Whitney U tests. AUC treated pCR as the positive outcome, Cliff's delta was calculated as 2 x AUC - 1, and Benjamini-Hochberg correction was applied across the ten evaluable T/NK subclusters.

**Conventional T-cell reclustering and abundance analysis**

The 70,567 cells assigned as 'T cells' were extracted directly from the PBMCs atlas. Cells classified as cytotoxic T/NK, T/NK intermediate, gamma-delta T or NK cells were not included. The subset was reprocessed using 3,000 T-cell-specific highly variable genes selected from raw counts. Scaling, principal-component analysis, nearest-neighbor graph construction and UMAP embedding used the same parameters as the T/NK analysis. Leiden clustering was calculated at resolution 0.3; the seven-subcluster solution was used for primary analyses. Marker genes for each subcluster were ranked using response-blind one-versus-rest Wilcoxon tests.

Subclusters were provisionally annotated from their marker-expression profiles as C0, CD4 IL7R naive/memory-like; C1, CD4 memory/activation-like; C2, preserved/quiescent-like; C3, heat-shock/stress-activated; C4, CD8 naive/memory-like; C5, preserved naive/memory-like; and C6, cytotoxic/stress-like. For each patient, conventional T-cell-subcluster abundance was calculated as the percentage of all reviewed conventional T cells. Missing patient-by-subcluster combinations were assigned an abundance of zero. Abundance was compared between response groups using two-sided Mann-Whitney U tests, with AUC and Cliff's delta as effect-size measures. Benjamini-Hochberg correction was applied across the seven T-cell subclusters. No minimum of 50 T cells per patient was imposed for these abundance comparisons.

**T-cell functional programs and targeted gene-level analyses**

Four biologically predefined T-cell programs were evaluated to represent complementary dimensions of T-cell state. The preserved/memory-like program comprised *IL7R, CCR7, SELL, TCF7, LEF1, LTB, CD27, CD28, TXK, FOXP1, RUNX1* and *SKAP1*. The cytotoxic program comprised *CCL5, NKG7, GZMK, GNLY, PRF1, GZMB, GZMH, KLRD1, KLRG1, CX3CR1, CTSW* and *FGFBP2*. The checkpoint/activation program comprised *PDCD1, TIGIT, LAG3, HAVCR2, CTLA4, ICOS, TNFRSF4, TNFRSF9, CD69, TOX* and *BTLA*. The ISG/stress program comprised *HSPA1A, HSPA1B, FOS, JUN, JUNB, DUSP1, ISG15, IFIT1, IFIT2, IFIT3, MX1, OAS1, IFITM1* and *IFITM2*. Gene sets were fixed before response-associated testing. Only genes present in the log-normalized expression matrix were retained, and at least 80% of the genes in a program were required to be available.

For each cell, the program score was calculated as the arithmetic mean of log-normalized expression across the available genes in the corresponding fixed gene set. No gene-wise standardization, background-gene subtraction or response-dependent weighting was applied. The patient-level value was the mean per-cell program score among that patient's cells within the corresponding T-cell subcluster. A patient was eligible for a subcluster-specific comparison when at least 30 cells from that patient were assigned to the subcluster. A subcluster was excluded from inferential program analyses if one patient contributed at least 70% of all cells in that subcluster or if fewer than five patients contributed any cells. Each pCR versus non-pCR comparison additionally required at least five eligible patients in each response group. C0, C1, C4 and C5 met the criteria for functional-program comparisons. C2 was excluded because one patient contributed 90.4% of its cells, whereas C3 and C6 did not meet the minimum number of eligible patients in at least one response group.

Evaluable program-level comparisons used two-sided Mann-Whitney U tests. AUC treated pCR as the positive outcome, and Cliff's delta was oriented as pCR minus non-pCR. Benjamini-Hochberg correction was performed across the four evaluable subclusters separately within each program; a secondary global correction was applied across all evaluable program-by-subcluster tests. Nominal P<0.05 and within-program q<0.10 were used as exploratory prioritization thresholds for individual box plots.

Targeted gene-level analyses were performed for every program-by-subcluster combination classified as evaluable, irrespective of the P or q value of the aggregate program score. For every available gene in the corresponding program, two patient-level metrics were calculated: mean log-normalized expression across all eligible cells, including zero values, and the percentage of cells with a raw count greater than zero. The same requirements of at least 30 cells per patient and subcluster and at least five eligible patients per response group were applied. Each gene-by-subcluster-by-metric combination was compared using a two-sided Mann-Whitney U test. Effect direction was represented by signed AUC, calculated as 2 x AUC - 1, such that negative values indicated higher values in non-pCR and positive values indicated higher values in pCR. Benjamini-Hochberg correction was applied within each program across all available genes, the four evaluable subclusters and both metrics; a secondary global correction included all tests from the four programs. Volcano plots displayed signed AUC against -log10(P). Across the four evaluable T-cell subclusters, this analysis comprised 392 gene-by-subcluster-by-metric tests involving 49 unique program genes.

**B-cell subclustering and functional analyses**

The manually reviewed B-cell compartment was extracted from the PBMCs atlas using the final compartment annotation. This selection retained conventional B cells and the reviewed plasmablast/plasma-cell population, yielding 20,608 cells. Raw-count values were retained in a dedicated AnnData layer and were verified to be non-negative and integer-like.

A total of 2,000 B-cell-specific highly variable genes were selected from raw counts using the Seurat v3 method. Log-normalized expression of the selected genes was zero-centered and scaled to unit variance, with scaled values clipped at 10. Principal-component analysis used the ARPACK solver. A 15-nearest-neighbor graph was constructed from the first 30 principal components and used for UMAP embedding. Leiden clustering was calculated at resolutions 0.3 and 0.6; resolution 0.3 was used for the primary B-cell analyses and resolution 0.6 for higher-resolution review. The primary solution comprised 11 subclusters, C0-C10. No expression-level integration or batch correction was applied; sample identity was used only during highly variable-gene selection.

B-cell subclusters were interpreted using genes representing B-cell lineage identity (*MS4A1, CD79A, CD79B, CD74, CD37, CD22* and *BANK1*), naive-like differentiation (*TCL1A, IGHD* and *IGHM*), memory-associated differentiation (*CD27, TNFRSF13B* and *AIM2*), activation and antigen presentation (*CD83, CD69, HLA-DRA, HLA-DRB1,* *NFKBIA* and *NR4A2*), CD38/plasma-cell differentiation (*CD38, MZB1, XBP1, JCHAIN, PRDM1* and *SDC1*), and interferon/stress responses (*ISG15, IFIT1, IFIT2, IFIT3, MX1, FOS, JUN, DUSP1* and *HSPA1A*). Marker genes were ranked by response-blind one-versus-rest Wilcoxon tests. In marker dot plots, dot size represented the percentage of cells with detectable expression and color represented mean expression scaled from 0 to 1 across subclusters separately for each gene.

Potential lineage contamination and sample dominance were evaluated before response-associated inference. Subclusters were excluded from inferential analyses when one patient contributed at least 70% of their cells or fewer than five patients contributed cells. C0, C8 and C10 met these sample-dominance or sample-specificity criteria. C3 was additionally excluded after manual review identified a mixed B/T-lineage signal, including *CD3D, CD3E, IL7R* and *GIMAP7* expression. These four subclusters were retained in descriptive UMAP and marker visualizations but were excluded from response-associated statistical testing and FDR correction. The seven QC-valid subclusters were C1, C2, C4, C5, C6, C7 and C9.

For B-cell-subcluster abundance analyses, patients with a known pathological-response classification and at least 50 reviewed B-compartment cells were included. One patient did not meet this threshold, leaving 13 pCR and eight non-pCR patients for inference. For each patient, subcluster abundance was calculated as the percentage of all reviewed B-compartment cells assigned to that subcluster. All patient-by-subcluster combinations were retained, and an absent subcluster was assigned an abundance of zero. Abundance was compared between pCR and non-pCR using two-sided Mann-Whitney U tests. AUC treated pCR as the positive outcome, and Cliff's delta was calculated as 2 x AUC - 1. Benjamini-Hochberg correction was applied across the seven QC-valid B-cell subclusters.

Five biologically predefined B-cell programs were subsequently examined. The naive-like program comprised *TCL1A*, *IGHD* and *IGHM*. The memory-like program comprised *CD27, TNFRSF13B* and *AIM2*. The activated/APC-like program comprised *CD83, CD69, HLA-DRA, HLA-DRB1, NFKBIA* and *NR4A2*. The CD38/plasma-like program comprised *CD38, MZB1, XBP1, JCHAIN, PRDM1* and *SDC1*. The ISG/stress program comprised I*SG15, IFIT1, IFIT2, IFIT3, MX1, FOS, JUN, DUSP1* and *HSPA1A*. These gene sets were fixed before response-associated testing. Only genes present in both the log-normalized expression matrix and raw-count layer were used, and at least 80% of the genes in a program were required to be available.

For each cell, the program score was calculated as the arithmetic mean of log-normalized expression across the available genes in the corresponding fixed gene set. No gene-wise standardization, background-gene subtraction or response-dependent weighting was applied. The patient-level value was the mean per-cell program score among that patient's cells within the corresponding B-cell subcluster. A patient was eligible for a subcluster-specific analysis when at least 30 cells from that patient were assigned to the subcluster. Each pCR versus non-pCR comparison required at least five eligible patients in each response group. C1 and C2 were the only QC-valid subclusters that met the patient-level cell-count and group-size requirements for functional-program inference.

Evaluable program-score comparisons used two-sided Mann-Whitney U tests. AUC treated pCR as the positive outcome, and Cliff's delta was oriented as pCR minus non-pCR. Benjamini-Hochberg correction was applied across C1 and C2 separately within each program; a secondary global correction was applied across all ten evaluable program-by-subcluster comparisons. Nominal P<0.05 and within-program q<0.10 were used as exploratory prioritization thresholds for individual box plots.

Targeted gene-level analyses were performed for every evaluable program-by-subcluster combination, irrespective of the P or q value of the aggregate program score. For every available gene in the corresponding program, two patient-level metrics were calculated: mean log-normalized expression across all eligible cells, including zero values, and the percentage of cells with a raw count greater than zero. The same requirements of at least 30 cells per patient and subcluster and at least five eligible patients per response group were applied. Each gene-by-subcluster-by-metric combination was compared using a two-sided Mann-Whitney U test. Effect direction was represented by signed AUC, calculated as 2 x AUC - 1, such that negative values indicated higher values in non-pCR and positive values indicated higher values in pCR. Benjamini-Hochberg correction was applied within each program across all available program genes, C1 and C2, and both patient-level metrics; a secondary global correction included all tests from the five programs. Volcano plots displayed signed AUC against -log10(P). Up to 20 nominally significant points per program were labelled, prioritizing comparisons with within-program q<0.10 and then the smallest P values. Across the two evaluable B-cell subclusters, this analysis comprised 108 gene-by-subcluster-by-metric tests involving 27 unique program genes.

**Flow-cytometry quality control and sample adjudication**

Panel 1 included the following antibodies from BioLegend, with fluorochrome, clone and catalogue number indicated in parentheses: CD3 (Alexa Fluor 700; UCHT1; 300424), CD4 (FITC; OKT4; 317408), CD8α (Brilliant Violet 421; RPA-T8; 301036), CD62L (Brilliant Violet 510; DREG-56; 304844), CD127 (Brilliant Violet 650; A019D5; 351326), CD27 (APC; M-T271; 356410), CD161 (Brilliant Violet 605; HP-3G10; 339916), CD279/PD-1 (PE; EH12.2H7; 329906), TIGIT (PE-Cy7; A15153G; 372714) and CD366/TIM-3 (APC-Fire 750; F38-2E2; 345044). CD197/CCR7 was detected using a PE-Vio615-conjugated antibody from Miltenyi Biotec (clone REA546; catalogue no. 130-120-465).

Panel 2 included antibodies against CD3 (Alexa Fluor 700; UCHT1; 300424), CD27 (APC; M-T271; 356410), CD19 (Pacific Blue; HIB19; 302232), IgD (PE; IA6-2; 348204), CD83 (Brilliant Violet 510; HB15e; 305348), HLA-DR (Brilliant Violet 650; L243; 307650), CD38 (FITC; HIT2; 303504), CD94 (PE-Cy7; DX22; 305516) and CD20 (Brilliant Violet 605; 2H7; 302334), all obtained from BioLegend.

Cell viability was assessed in both panels using 7-aminoactinomycin D (7-AAD; Thermo Fisher Scientific; catalogue no. 00-6993-50), with viable cells identified as 7-AAD-negative.

Only the adjudicated primary acquisition was retained for each patient and time point. When an original acquisition was invalid, a designated repeat-primary acquisition was used in its place. Samples were excluded when viability was ≤10%, data were missing or non-interpretable. Response comparisons required at least five evaluable patients in each group.

Endpoint-specific eligibility followed the gating hierarchy. Major-population estimates required at least 500 viable events. CD4+ and CD8+ frequencies required at least 500 total T cells, and first-level subset analyses required at least 200 CD4+ or 100 CD8+ cells. Nested T-cell phenotypes required at least 50 events in the relevant parent population. B-cell subsets and nested CD38 branches required at least 50 CD19+ cells or at least 50 cells in the corresponding B-cell parent, as applicable.

PD-1-containing phenotypes were analyzed only at baseline (T1), because pembrolizumab could interfere with post-treatment detection by clone EH12.2H7. These phenotypes were excluded from cross-sectional analyses at T2 and T3 and from all longitudinal analyses.

**Additional statistical specifications**

Benjamini–Hochberg correction was applied separately across the ten evaluable T/NK subclusters, all seven conventional T-cell subclusters and the seven QC-valid B-cell subclusters for abundance comparisons. For functional-program analyses, correction was applied across evaluable subclusters separately within each program and cell compartment. For targeted gene-level analyses, each correction family included all available program genes, all evaluable subclusters and both patient-level metrics, separately for T and B cells.

For general flow-cytometry endpoints, 95% confidence intervals for Cliff’s delta were estimated using 2,000 bootstrap resamples. The primary T3 switched-memory CD38 balance used the stratified bootstrap procedure specified in the main Methods. Bootstrap confidence intervals were calculated using the percentile method.
