## Supplementary Tables for "CIRCULATING IMMUNE DYNAMICS DURING NEOADJUVANT CHEMOIMMUNOTHERAPY IN EARLY TRIPLE-NEGATIVE BREAST CANCER"

**Supplementary Table 1. Baseline T/NK subcluster abundance according to pathological response.**

| TNK subcluster | Inference status | Inference note | n pCR | n non-pCR | Mean abundance, pCR (%) | Mean abundance, non-pCR (%) | Median abundance, pCR (%) | Median abundance, non-pCR (%) | Median difference (pCR − non-pCR), pp | Mann–Whitney U | Two-sided P | AUC (pCR higher) | Cliff's δ (pCR − non-pCR) | Benjamini–Hochberg q | Total cells | Contributing patients |
|---|---|---|---|---|---|---|---|---|---|---|---|---|---|---|---|---|
| 0 | Evaluable | No prespecified sample-dominance exclusion. | 14 | 8 | 12.21 | 20.97 | 3.85 | 9.63 | -5.77 | 41 | 0.3301 | 0.366 | -0.268 | 0.7352 | 25,072 | 21 |
| 1 | Evaluable | No prespecified sample-dominance exclusion. | 14 | 8 | 29.71 | 25.55 | 16.62 | 18.10 | -1.48 | 59 | 0.8676 | 0.527 | 0.054 | 0.8676 | 17,916 | 22 |
| 2 | Evaluable | No prespecified sample-dominance exclusion. | 14 | 8 | 10.72 | 11.26 | 5.05 | 10.15 | -5.10 | 49 | 0.6642 | 0.438 | -0.125 | 0.7810 | 15,496 | 21 |
| 3 | *Non-evaluable* | *Non-evaluable: cluster driven by individual samples* | 14 | 8 | 7.98 | 12.72 | 0.60 | 0.42 | 0.18 | | | | | | 14,907 | 22 |
| 4 | Evaluable | No prespecified sample-dominance exclusion. | 14 | 8 | 15.35 | 9.72 | 0.02 | 0.25 | -0.23 | 43 | 0.3820 | 0.384 | -0.232 | 0.7352 | 7,981 | 14 |
| 5 | Evaluable | No prespecified sample-dominance exclusion. | 14 | 8 | 8.81 | 5.95 | 7.71 | 5.06 | 2.65 | 70 | 0.3650 | 0.625 | 0.250 | 0.7352 | 7,507 | 22 |
| 6 | Evaluable | No prespecified sample-dominance exclusion. | 14 | 8 | 8.47 | 5.12 | 6.60 | 5.33 | 1.27 | 70 | 0.3650 | 0.625 | 0.250 | 0.7352 | 4,755 | 22 |
| 7 | Evaluable with caution | Evaluable with caution: coherent gamma-delta T-cell phenotype, but P67 contributes 61.7% of all cluster cells. | 14 | 8 | 1.59 | 1.50 | 0.08 | 0.21 | -0.13 | 50 | 0.7029 | 0.446 | -0.107 | 0.7810 | 4,079 | 15 |
| 8 | Evaluable | No prespecified sample-dominance exclusion. | 14 | 8 | 3.02 | 2.87 | 2.39 | 0.94 | 1.45 | 63 | 0.6642 | 0.563 | 0.125 | 0.7810 | 3,655 | 21 |
| 9 | Evaluable | No prespecified sample-dominance exclusion. | 14 | 8 | 1.23 | 2.06 | 0.94 | 1.68 | -0.74 | 38 | 0.2382 | 0.339 | -0.321 | 0.7352 | 1,583 | 22 |
| 10 | Evaluable | No prespecified sample-dominance exclusion. | 14 | 8 | 0.91 | 0.61 | 0.52 | 0.29 | 0.24 | 68 | 0.4411 | 0.607 | 0.214 | 0.7352 | 553 | 22 |
| 11 | *Non-evaluable* | *Non-evaluable: cluster driven by individual samples* | 14 | 8 | 0.00 | 1.69 | 0.00 | 0.00 | 0.00 | | | | | | 431 | 1 |

AUC, area under the receiver operating characteristic curve; n, number of patients; pCR, pathological complete response; non-pCR, absence of pCR; pp, percentage points; q, Benjamini–Hochberg-adjusted P value; TNK, combined T-cell/natural killer-cell compartment.

**Supplementary Table 2. Baseline conventional T-cell subcluster abundance according to pathological response.**

| T-cell subcluster | Functional annotation | Inference status | n pCR | n non-pCR | Median abundance, pCR (%) | Median abundance, non-pCR (%) | Median difference (pCR − non-pCR), pp | Mann–Whitney U | Two-sided P | Benjamini–Hochberg q | AUC (pCR higher) | Cliff's δ (pCR − non-pCR) | QC excluded from cluster-specific inference |
|---|---|---|---|---|---|---|---|---|---|---|---|---|---|
| 0 | CD4 IL7R naive/memory-like | Evaluable | 14 | 8 | 3.67 | 18.32 | -14.65 | 46.0 | 0.5252 | 0.8516 | 0.411 | -0.179 | No |
| 1 | CD4 memory/activation-like | Evaluable | 14 | 8 | 26.99 | 31.48 | -4.49 | 54.0 | 0.9203 | 0.9203 | 0.482 | -0.036 | No |
| *2* | *preserved/quiescent-like* | *QC-excluded* | *14* | *8* | *1.38* | *0.86* | *0.53* | *75.0* | *0.2119* | *0.7415* | *0.670* | *0.339* | *Yes* |
| 3 | heat-shock/stress-activated | Evaluable | 14 | 8 | 0.09 | 0.22 | -0.14 | 52.5 | 0.8338 | 0.9203 | 0.469 | -0.063 | No |
| 4 | CD8 naive/memory-like | Evaluable | 14 | 8 | 4.64 | 2.12 | 2.52 | 64.0 | 0.6083 | 0.8516 | 0.571 | 0.143 | No |
| 5 | preserved naive/memory-like | Evaluable | 14 | 8 | 5.60 | 1.44 | 4.16 | 83.0 | 0.0698 | 0.4888 | 0.741 | 0.482 | No |
| 6 | cytotoxic/stress-like | Evaluable | 14 | 8 | 0.00 | 0.08 | -0.08 | 48.0 | 0.5844 | 0.8516 | 0.429 | -0.143 | No |

AUC, area under the receiver operating characteristic curve; n, number of patients; pCR, pathological complete response; non-pCR, absence of pCR; pp, percentage points; QC, quality control; q, Benjamini–Hochberg-adjusted P value.

**Supplementary Table 3. Baseline T-cell functional program scores according to pathological response.**

| T-cell program | T-cell subcluster | Functional annotation | Evaluation status | n pCR | n non-pCR | Median score, pCR | Median score, non-pCR | Mann–Whitney U | Two-sided P | Benjamini–Hochberg q within program | AUC (pCR higher) | Cliff's δ (pCR − non-pCR) |
|---|---|---|---|---|---|---|---|---|---|---|---|---|
| **Checkpoint/activation** | 0 | CD4 IL7R naive/memory-like | Evaluable | 9 | 6 | 0.110 | 0.101 | 36.0 | 0.3277 | 1.0000 | 0.667 | 0.333 |
| **Checkpoint/activation** | 1 | CD4 memory/activation-like | Evaluable | 14 | 7 | 0.121 | 0.091 | 57.0 | 0.5846 | 1.0000 | 0.582 | 0.163 |
| **Checkpoint/activation** | 2 | *preserved/quiescent-like* | *QC-excluded sample-dominated cluster* | *6* | *2* | *0.206* | *0.215* | | | | | |
| **Checkpoint/activation** | 3 | *heat-shock/stress-activated* | *Fewer than five eligible patients in at least one group* | *3* | *3* | *0.318* | *0.243* | | | | | |
| **Checkpoint/activation** | 4 | CD8 naive/memory-like | Evaluable | 10 | 6 | 0.085 | 0.093 | 30.0 | 1.0000 | 1.0000 | 0.500 | 0.000 |
| **Checkpoint/activation** | 5 | preserved naive/memory-like | Evaluable | 13 | 6 | 0.079 | 0.106 | 36.0 | 0.8314 | 1.0000 | 0.462 | -0.077 |
| **Checkpoint/activation** | 6 | *cytotoxic/stress-like* | *Fewer than five eligible patients in at least one group* | *3* | *2* | *0.301* | *0.223* | | | | | |
| **Cytotoxic** | 0 | CD4 IL7R naive/memory-like | Evaluable | 9 | 6 | 0.058 | 0.045 | 39.0 | 0.1810 | 0.3620 | 0.722 | 0.444 |
| **Cytotoxic** | 1 | CD4 memory/activation-like | Evaluable | 14 | 7 | 0.108 | 0.083 | 59.0 | 0.4880 | 0.4880 | 0.602 | 0.204 |
| **Cytotoxic** | 2 | *preserved/quiescent-like* | *QC-excluded sample-dominated cluster* | *6* | *2* | *0.213* | *0.176* | | | | | |
| **Cytotoxic** | 3 | *heat-shock/stress-activated* | *Fewer than five eligible patients in at least one group* | *3* | *3* | *0.095* | *0.052* | | | | | |
| **Cytotoxic** | 4 | CD8 naive/memory-like | Evaluable | 10 | 6 | 0.101 | 0.124 | 20.0 | 0.3132 | 0.4176 | 0.333 | -0.333 |
| **Cytotoxic** | 5 | preserved naive/memory-like | Evaluable | 13 | 6 | 0.037 | 0.025 | 55.0 | 0.1791 | 0.3620 | 0.705 | 0.410 |
| **Cytotoxic** | 6 | *cytotoxic/stress-like* | *Fewer than five eligible patients in at least one group* | *3* | *2* | *1.548* | *1.771* | | | | | |
| **ISG/stress** | 0 | CD4 IL7R naive/memory-like | Evaluable | 9 | 6 | 0.510 | 0.559 | 26.0 | 0.9546 | 0.9546 | 0.481 | -0.037 |
| **ISG/stress** | 1 | CD4 memory/activation-like | Evaluable | 14 | 7 | 0.264 | 0.361 | 27.0 | 0.1101 | 0.4403 | 0.276 | -0.449 |
| **ISG/stress** | 2 | *preserved/quiescent-like* | *QC-excluded sample-dominated cluster* | *6* | *2* | *0.086* | *0.207* | | | | | |
| **ISG/stress** | 3 | *heat-shock/stress-activated* | *Fewer than five eligible patients in at least one group* | *3* | *3* | *1.276* | *0.994* | | | | | |
| **ISG/stress** | 4 | CD8 naive/memory-like | Evaluable | 10 | 6 | 0.386 | 0.500 | 24.0 | 0.5622 | 0.9546 | 0.400 | -0.200 |
| **ISG/stress** | 5 | preserved naive/memory-like | Evaluable | 13 | 6 | 0.197 | 0.158 | 43.0 | 0.7654 | 0.9546 | 0.551 | 0.103 |
| **ISG/stress** | 6 | *cytotoxic/stress-like* | *Fewer than five eligible patients in at least one group* | *3* | *2* | *1.526* | *1.294* | | | | | |
| **Preserved/memory-like** | 0 | CD4 IL7R naive/memory-like | Evaluable | 9 | 6 | 1.178 | 1.053 | 34.0 | 0.4559 | 0.4670 | 0.630 | 0.259 |
| **Preserved/memory-like** | 1 | CD4 memory/activation-like | Evaluable | 14 | 7 | 0.988 | 0.897 | 64.0 | 0.2872 | 0.4670 | 0.653 | 0.306 |
| **Preserved/memory-like** | 2 | *preserved/quiescent-like* | *QC-excluded sample-dominated cluster* | *6* | *2* | *1.097* | *1.008* | | | | | |
| **Preserved/memory-like** | 3 | *heat-shock/stress-activated* | *Fewer than five eligible patients in at least one group* | *3* | *3* | *1.121* | *1.260* | | | | | |
| **Preserved/memory-like** | 4 | CD8 naive/memory-like | Evaluable | 10 | 6 | 1.260 | 1.081 | 47.0 | 0.0727 | 0.2907 | 0.783 | 0.567 |
| **Preserved/memory-like** | 5 | preserved naive/memory-like | Evaluable | 13 | 6 | 1.485 | 1.423 | 48.0 | 0.4670 | 0.4670 | 0.615 | 0.231 |
| **Preserved/memory-like** | 6 | *cytotoxic/stress-like* | *Fewer than five eligible patients in at least one group* | *3* | *2* | *0.574* | *0.601* | | | | | |

AUC, area under the receiver operating characteristic curve; ISG, interferon-stimulated gene; n, number of patients; pCR, pathological complete response; non-pCR, absence of pCR; QC, quality control; q, Benjamini–Hochberg-adjusted P value.

**Supplementary Table 4. Gene-level comparisons within T-cell functional programs according to pathological response.**

| T-cell program | T-cell subcluster | Functional annotation | Gene | Metric | Evaluation status | n pCR | n non-pCR | Mean, pCR | Mean, non-pCR | Median, pCR | Median, non-pCR | Mann–Whitney U | Two-sided P | AUC (pCR higher) | Signed AUC | Cliff's δ (pCR − non-pCR) | Benjamini–Hochberg q within program | Global Benjamini–Hochberg q | Nominal P <0.05 | Within-program FDR q <0.10 | Global FDR q <0.10 |
|---|---|---|---|---|---|---|---|---|---|---|---|---|---|---|---|---|---|---|---|---|---|
| **Cytotoxic** | 1 | CD4 memory/activation-like | *CX3CR1* | Mean expression | Evaluable | 14 | 7 | 0.024 | 0.005 | 0.021 | 0.005 | 93. | 0.0003 | 0.9490 | 0.8980 | 0.8980 | 0.0157 | 0.0641 | Yes | Yes | Yes |
| **Cytotoxic** | 1 | CD4 memory/activation-like | *CX3CR1* | % positive | Evaluable | 14 | 7 | 1.171 | 0.203 | 0.964 | 0.239 | 93. | 0.0003 | 0.9490 | 0.8980 | 0.8980 | 0.0157 | 0.0641 | Yes | Yes | Yes |
| **Checkpoint/activation** | 1 | CD4 memory/activation-like | *HAVCR2* | Mean expression | Evaluable | 14 | 7 | 0.028 | 0.013 | 0.026 | 0.011 | 83.5 | 0.0112 | 0.8520 | 0.7041 | 0.7041 | 0.9627 | 0.9462 | Yes | No | No |
| **Preserved/memory-like** | 5 | preserved naive/memory-like | *FOXP1* | % positive | Evaluable | 13 | 6 | 93.500 | 88.645 | 94.872 | 90.709 | 66. | 0.0167 | 0.8462 | 0.6923 | 0.6923 | 0.7098 | 0.9462 | Yes | No | No |
| **Cytotoxic** | 1 | CD4 memory/activation-like | *GZMB* | % positive | Evaluable | 14 | 7 | 0.578 | 0.062 | 0.398 | 0.000 | 80. | 0.0205 | 0.8163 | 0.6327 | 0.6327 | 0.6066 | 0.9462 | Yes | No | No |
| **Preserved/memory-like** | 5 | preserved naive/memory-like | *FOXP1* | Mean expression | Evaluable | 13 | 6 | 3.010 | 2.695 | 3.006 | 2.698 | 64. | 0.0285 | 0.8205 | 0.6410 | 0.6410 | 0.7098 | 0.9462 | Yes | No | No |
| **Cytotoxic** | 1 | CD4 memory/activation-like | *GZMB* | Mean expression | Evaluable | 14 | 7 | 0.012 | 0.002 | 0.007 | 0.000 | 78. | 0.0303 | 0.7959 | 0.5918 | 0.5918 | 0.6066 | 0.9462 | Yes | No | No |
| **Checkpoint/activation** | 1 | CD4 memory/activation-like | *HAVCR2* | % positive | Evaluable | 14 | 7 | 1.534 | 0.648 | 1.314 | 0.597 | 78.5 | 0.0304 | 0.8010 | 0.6020 | 0.6020 | 0.9627 | 0.9462 | Yes | No | No |
| **ISG/stress** | 1 | CD4 memory/activation-like | *OAS1* | Mean expression | Evaluable | 14 | 7 | 0.060 | 0.031 | 0.059 | 0.029 | 78. | 0.0333 | 0.7959 | 0.5918 | 0.5918 | 0.9798 | 0.9462 | Yes | No | No |
| **Preserved/memory-like** | 4 | CD8 naive/memory-like | *FOXP1* | % positive | Evaluable | 10 | 6 | 83.308 | 66.094 | 79.921 | 63.277 | 49. | 0.0420 | 0.8167 | 0.6333 | 0.6333 | 0.7098 | 0.9462 | Yes | No | No |
| **Cytotoxic** | 4 | CD8 naive/memory-like | *GZMB* | Mean expression | Evaluable | 10 | 6 | 0.014 | 0.001 | 0.008 | 0.000 | 48. | 0.0475 | 0.8000 | 0.6000 | 0.6000 | 0.6066 | 0.9462 | Yes | No | No |
| **Preserved/memory-like** | 1 | CD4 memory/activation-like | *LEF1* | Mean expression | Evaluable | 14 | 7 | 1.072 | 0.847 | 1.033 | 0.754 | 75. | 0.0556 | 0.7653 | 0.5306 | 0.5306 | 0.7098 | 0.9462 | No | No | No |
| **Preserved/memory-like** | 4 | CD8 naive/memory-like | *LEF1* | Mean expression | Evaluable | 10 | 6 | 1.867 | 1.439 | 1.730 | 1.389 | 48. | 0.0559 | 0.8000 | 0.6000 | 0.6000 | 0.7098 | 0.9462 | No | No | No |
| **Preserved/memory-like** | 5 | preserved naive/memory-like | *RUNX1* | % positive | Evaluable | 13 | 6 | 73.586 | 63.104 | 71.875 | 67.375 | 61. | 0.0577 | 0.7821 | 0.5641 | 0.5641 | 0.7098 | 0.9462 | No | No | No |
| **ISG/stress** | 0 | CD4 IL7R naive/memory-like | *OAS1* | Mean expression | Evaluable | 9 | 6 | 0.049 | 0.021 | 0.034 | 0.026 | 43.5 | 0.0591 | 0.8056 | 0.6111 | 0.6111 | 0.9798 | 0.9462 | No | No | No |
| **Preserved/memory-like** | 0 | CD4 IL7R naive/memory-like | *LTB* | % positive | Evaluable | 9 | 6 | 85.022 | 75.955 | 88.837 | 79.254 | 43. | 0.0663 | 0.7963 | 0.5926 | 0.5926 | 0.7098 | 0.9462 | No | No | No |
| **Cytotoxic** | 1 | CD4 memory/activation-like | *KLRD1* | Mean expression | Evaluable | 14 | 7 | 0.059 | 0.030 | 0.044 | 0.024 | 74. | 0.0667 | 0.7551 | 0.5102 | 0.5102 | 0.6066 | 0.9462 | No | No | No |
| **Cytotoxic** | 1 | CD4 memory/activation-like | *KLRD1* | % positive | Evaluable | 14 | 7 | 2.468 | 1.443 | 1.825 | 1.127 | 74. | 0.0667 | 0.7551 | 0.5102 | 0.5102 | 0.6066 | 0.9462 | No | No | No |
| **Preserved/memory-like** | 1 | CD4 memory/activation-like | *FOXP1* | Mean expression | Evaluable | 14 | 7 | 1.704 | 1.318 | 1.650 | 1.414 | 74. | 0.0667 | 0.7551 | 0.5102 | 0.5102 | 0.7098 | 0.9462 | No | No | No |
| **ISG/stress** | 1 | CD4 memory/activation-like | *OAS1* | % positive | Evaluable | 14 | 7 | 3.132 | 1.560 | 2.741 | 1.356 | 74. | 0.0672 | 0.7551 | 0.5102 | 0.5102 | 0.9798 | 0.9462 | No | No | No |
| **Cytotoxic** | 4 | CD8 naive/memory-like | *PRF1* | Mean expression | Evaluable | 10 | 6 | 0.071 | 0.045 | 0.066 | 0.047 | 47. | 0.0727 | 0.7833 | 0.5667 | 0.5667 | 0.6066 | 0.9462 | No | No | No |
| **Cytotoxic** | 0 | CD4 IL7R naive/memory-like | *CX3CR1* | % positive | Evaluable | 9 | 6 | 0.823 | 0.235 | 0.451 | 0.100 | 42.5 | 0.0768 | 0.7870 | 0.5741 | 0.5741 | 0.6066 | 0.9462 | No | No | No |
| **Cytotoxic** | 1 | CD4 memory/activation-like | *CTSW* | Mean expression | Evaluable | 14 | 7 | 0.100 | 0.159 | 0.086 | 0.183 | 25. | 0.0793 | 0.2551 | -0.4898 | -0.4898 | 0.6066 | 0.9462 | No | No | No |
| **Checkpoint/activation** | 0 | CD4 IL7R naive/memory-like | *HAVCR2* | Mean expression | Evaluable | 9 | 6 | 0.024 | 0.010 | 0.020 | 0.007 | 42. | 0.0863 | 0.7778 | 0.5556 | 0.5556 | 0.9627 | 0.9462 | No | No | No |
| **Checkpoint/activation** | 0 | CD4 IL7R naive/memory-like | *HAVCR2* | % positive | Evaluable | 9 | 6 | 1.693 | 0.647 | 1.343 | 0.365 | 42. | 0.0863 | 0.7778 | 0.5556 | 0.5556 | 0.9627 | 0.9462 | No | No | No |
| **Preserved/memory-like** | 5 | preserved naive/memory-like | *RUNX1* | Mean expression | Evaluable | 13 | 6 | 1.923 | 1.605 | 1.836 | 1.654 | 59. | 0.0874 | 0.7564 | 0.5128 | 0.5128 | 0.7098 | 0.9462 | No | No | No |
| **Cytotoxic** | 0 | CD4 IL7R naive/memory-like | *GNLY* | Mean expression | Evaluable | 9 | 6 | 0.043 | 0.021 | 0.045 | 0.018 | 42. | 0.0879 | 0.7778 | 0.5556 | 0.5556 | 0.6066 | 0.9462 | No | No | No |
| **Preserved/memory-like** | 4 | CD8 naive/memory-like | *FOXP1* | Mean expression | Evaluable | 10 | 6 | 1.848 | 1.347 | 1.721 | 1.280 | 46. | 0.0934 | 0.7667 | 0.5333 | 0.5333 | 0.7098 | 0.9462 | No | No | No |
| **Preserved/memory-like** | 4 | CD8 naive/memory-like | *TXK* | Mean expression | Evaluable | 10 | 6 | 1.289 | 0.976 | 1.235 | 0.964 | 46. | 0.0934 | 0.7667 | 0.5333 | 0.5333 | 0.7098 | 0.9462 | No | No | No |
| **ISG/stress** | 1 | CD4 memory/activation-like | *IFITM1* | Mean expression | Evaluable | 14 | 7 | 1.413 | 1.792 | 1.540 | 2.001 | 26. | 0.0938 | 0.2653 | -0.4694 | -0.4694 | 0.9798 | 0.9462 | No | No | No |
| **ISG/stress** | 1 | CD4 memory/activation-like | *IFITM1* | % positive | Evaluable | 14 | 7 | 51.437 | 63.316 | 56.387 | 68.854 | 26. | 0.0938 | 0.2653 | -0.4694 | -0.4694 | 0.9798 | 0.9462 | No | No | No |
| **ISG/stress** | 1 | CD4 memory/activation-like | *IFITM2* | Mean expression | Evaluable | 14 | 7 | 0.723 | 1.035 | 0.852 | 0.986 | 26. | 0.0938 | 0.2653 | -0.4694 | -0.4694 | 0.9798 | 0.9462 | No | No | No |
| **Cytotoxic** | 0 | CD4 IL7R naive/memory-like | *CX3CR1* | Mean expression | Evaluable | 9 | 6 | 0.012 | 0.004 | 0.008 | 0.001 | 41.5 | 0.0987 | 0.7685 | 0.5370 | 0.5370 | 0.6066 | 0.9462 | No | No | No |
| **Cytotoxic** | 0 | CD4 IL7R naive/memory-like | *PRF1* | Mean expression | Evaluable | 9 | 6 | 0.031 | 0.018 | 0.036 | 0.021 | 41.5 | 0.0987 | 0.7685 | 0.5370 | 0.5370 | 0.6066 | 0.9462 | No | No | No |
| **Cytotoxic** | 1 | CD4 memory/activation-like | *CTSW* | % positive | Evaluable | 14 | 7 | 5.262 | 8.360 | 4.626 | 7.948 | 27. | 0.1101 | 0.2755 | -0.4490 | -0.4490 | 0.6066 | 0.9462 | No | No | No |
| **Cytotoxic** | 1 | CD4 memory/activation-like | *GNLY* | Mean expression | Evaluable | 14 | 7 | 0.126 | 0.094 | 0.109 | 0.065 | 71. | 0.1101 | 0.7245 | 0.4490 | 0.4490 | 0.6066 | 0.9462 | No | No | No |
| **Cytotoxic** | 1 | CD4 memory/activation-like | *GNLY* | % positive | Evaluable | 14 | 7 | 5.143 | 3.972 | 4.342 | 2.303 | 71. | 0.1101 | 0.7245 | 0.4490 | 0.4490 | 0.6066 | 0.9462 | No | No | No |
| **ISG/stress** | 1 | CD4 memory/activation-like | *IFITM2* | % positive | Evaluable | 14 | 7 | 32.239 | 43.137 | 28.873 | 43.911 | 27. | 0.1101 | 0.2755 | -0.4490 | -0.4490 | 0.9798 | 0.9462 | No | No | No |
| **Cytotoxic** | 0 | CD4 IL7R naive/memory-like | *GNLY* | % positive | Evaluable | 9 | 6 | 2.584 | 1.193 | 2.388 | 1.189 | 41. | 0.1135 | 0.7593 | 0.5185 | 0.5185 | 0.6066 | 0.9462 | No | No | No |
| **Cytotoxic** | 5 | preserved naive/memory-like | *GNLY* | Mean expression | Evaluable | 13 | 6 | 0.046 | 0.012 | 0.040 | 0.005 | 57. | 0.1155 | 0.7308 | 0.4615 | 0.4615 | 0.6066 | 0.9462 | No | No | No |
| **Preserved/memory-like** | 4 | CD8 naive/memory-like | *LEF1* | % positive | Evaluable | 10 | 6 | 85.070 | 69.719 | 85.931 | 63.480 | 45. | 0.1179 | 0.7500 | 0.5000 | 0.5000 | 0.7098 | 0.9462 | No | No | No |
| **Preserved/memory-like** | 4 | CD8 naive/memory-like | *TXK* | % positive | Evaluable | 10 | 6 | 69.053 | 53.067 | 68.842 | 47.872 | 45. | 0.1179 | 0.7500 | 0.5000 | 0.5000 | 0.7098 | 0.9462 | No | No | No |
| **Checkpoint/activation** | 5 | preserved naive/memory-like | *HAVCR2* | Mean expression | Evaluable | 13 | 6 | 0.062 | 0.033 | 0.055 | 0.037 | 57. | 0.1247 | 0.7308 | 0.4615 | 0.4615 | 0.9627 | 0.9462 | No | No | No |
| **Cytotoxic** | 0 | CD4 IL7R naive/memory-like | *PRF1* | % positive | Evaluable | 9 | 6 | 2.168 | 1.207 | 2.099 | 1.219 | 40.5 | 0.1252 | 0.7500 | 0.5000 | 0.5000 | 0.6066 | 0.9462 | No | No | No |
| **Cytotoxic** | 4 | CD8 naive/memory-like | *GZMB* | % positive | Evaluable | 10 | 6 | 0.925 | 0.137 | 0.491 | 0.000 | 44. | 0.1264 | 0.7333 | 0.4667 | 0.4667 | 0.6066 | 0.9462 | No | No | No |
| **Preserved/memory-like** | 5 | preserved naive/memory-like | *TXK* | Mean expression | Evaluable | 13 | 6 | 1.512 | 1.318 | 1.465 | 1.237 | 57. | 0.1274 | 0.7308 | 0.4615 | 0.4615 | 0.7098 | 0.9462 | No | No | No |
| **Cytotoxic** | 5 | preserved naive/memory-like | *NKG7* | Mean expression | Evaluable | 13 | 6 | 0.043 | 0.012 | 0.031 | 0.000 | 56. | 0.1327 | 0.7179 | 0.4359 | 0.4359 | 0.6066 | 0.9462 | No | No | No |
| **Preserved/memory-like** | 4 | CD8 naive/memory-like | *TCF7* | % positive | Evaluable | 10 | 6 | 69.046 | 60.597 | 68.772 | 57.000 | 44. | 0.1428 | 0.7333 | 0.4667 | 0.4667 | 0.7098 | 0.9462 | No | No | No |
| **Preserved/memory-like** | 0 | CD4 IL7R naive/memory-like | *TXK* | Mean expression | Evaluable | 9 | 6 | 0.828 | 0.671 | 0.788 | 0.643 | 40. | 0.1447 | 0.7407 | 0.4815 | 0.4815 | 0.7098 | 0.9462 | No | No | No |
| **Preserved/memory-like** | 0 | CD4 IL7R naive/memory-like | *TXK* | % positive | Evaluable | 9 | 6 | 46.892 | 37.027 | 43.057 | 35.606 | 40. | 0.1447 | 0.7407 | 0.4815 | 0.4815 | 0.7098 | 0.9462 | No | No | No |
| **Preserved/memory-like** | 4 | CD8 naive/memory-like | *TCF7* | Mean expression | Evaluable | 10 | 6 | 1.257 | 1.117 | 1.188 | 1.110 | 44. | 0.1471 | 0.7333 | 0.4667 | 0.4667 | 0.7098 | 0.9462 | No | No | No |
| **Cytotoxic** | 5 | preserved naive/memory-like | *CCL5* | % positive | Evaluable | 13 | 6 | 3.017 | 1.776 | 3.125 | 1.812 | 56. | 0.1471 | 0.7179 | 0.4359 | 0.4359 | 0.6085 | 0.9462 | No | No | No |
| **Checkpoint/activation** | 1 | CD4 memory/activation-like | *CTLA4* | Mean expression | Evaluable | 14 | 7 | 0.087 | 0.042 | 0.052 | 0.037 | 69. | 0.1490 | 0.7041 | 0.4082 | 0.4082 | 0.9627 | 0.9462 | No | No | No |
| **Checkpoint/activation** | 1 | CD4 memory/activation-like | *CTLA4* | % positive | Evaluable | 14 | 7 | 4.718 | 1.989 | 2.950 | 1.568 | 69. | 0.1490 | 0.7041 | 0.4082 | 0.4082 | 0.9627 | 0.9462 | No | No | No |
| **ISG/stress** | 1 | CD4 memory/activation-like | *MX1* | % positive | Evaluable | 14 | 7 | 5.884 | 3.766 | 4.822 | 3.368 | 69. | 0.1490 | 0.7041 | 0.4082 | 0.4082 | 0.9798 | 0.9462 | No | No | No |
| **Preserved/memory-like** | 1 | CD4 memory/activation-like | *LEF1* | % positive | Evaluable | 14 | 7 | 43.895 | 36.107 | 42.641 | 32.258 | 69. | 0.1490 | 0.7041 | 0.4082 | 0.4082 | 0.7098 | 0.9462 | No | No | No |
| **Checkpoint/activation** | 5 | preserved naive/memory-like | *PDCD1* | Mean expression | Evaluable | 13 | 6 | 0.003 | 0.000 | 0.000 | 0.000 | 51. | 0.1573 | 0.6538 | 0.3077 | 0.3077 | 0.9627 | 0.9462 | No | No | No |
| **Checkpoint/activation** | 5 | preserved naive/memory-like | *PDCD1* | % positive | Evaluable | 13 | 6 | 0.210 | 0.000 | 0.000 | 0.000 | 51. | 0.1573 | 0.6538 | 0.3077 | 0.3077 | 0.9627 | 0.9462 | No | No | No |
| **Cytotoxic** | 5 | preserved naive/memory-like | *GZMB* | Mean expression | Evaluable | 13 | 6 | 0.011 | 0.000 | 0.000 | 0.000 | 51. | 0.1573 | 0.6538 | 0.3077 | 0.3077 | 0.6085 | 0.9462 | No | No | No |
| **Cytotoxic** | 5 | preserved naive/memory-like | *GZMB* | % positive | Evaluable | 13 | 6 | 0.493 | 0.000 | 0.000 | 0.000 | 51. | 0.1573 | 0.6538 | 0.3077 | 0.3077 | 0.6085 | 0.9462 | No | No | No |
| **Cytotoxic** | 5 | preserved naive/memory-like | *GNLY* | % positive | Evaluable | 13 | 6 | 2.301 | 0.691 | 1.754 | 0.355 | 55. | 0.1633 | 0.7051 | 0.4103 | 0.4103 | 0.6085 | 0.9462 | No | No | No |
| **Checkpoint/activation** | 1 | CD4 memory/activation-like | *PDCD1* | Mean expression | Evaluable | 14 | 7 | 0.026 | 0.016 | 0.026 | 0.014 | 68. | 0.1674 | 0.6939 | 0.3878 | 0.3878 | 0.9627 | 0.9462 | No | No | No |
| **Preserved/memory-like** | 1 | CD4 memory/activation-like | *FOXP1* | % positive | Evaluable | 14 | 7 | 63.925 | 52.267 | 65.608 | 56.205 | 68. | 0.1718 | 0.6939 | 0.3878 | 0.3878 | 0.7098 | 0.9462 | No | No | No |
| **Cytotoxic** | 5 | preserved naive/memory-like | *KLRG1* | Mean expression | Evaluable | 13 | 6 | 0.193 | 0.134 | 0.162 | 0.144 | 55. | 0.1791 | 0.7051 | 0.4103 | 0.4103 | 0.6085 | 0.9462 | No | No | No |
| **Cytotoxic** | 4 | CD8 naive/memory-like | *CCL5* | Mean expression | Evaluable | 10 | 6 | 0.336 | 0.602 | 0.310 | 0.602 | 17. | 0.1806 | 0.2833 | -0.4333 | -0.4333 | 0.6085 | 0.9462 | No | No | No |
| **Preserved/memory-like** | 4 | CD8 naive/memory-like | *LTB* | Mean expression | Evaluable | 10 | 6 | 1.551 | 1.460 | 1.609 | 1.459 | 43. | 0.1806 | 0.7167 | 0.4333 | 0.4333 | 0.7098 | 0.9462 | No | No | No |
| **Cytotoxic** | 0 | CD4 IL7R naive/memory-like | *KLRG1* | % positive | Evaluable | 9 | 6 | 12.989 | 9.648 | 12.500 | 7.444 | 39. | 0.1810 | 0.7222 | 0.4444 | 0.4444 | 0.6085 | 0.9462 | No | No | No |
| **ISG/stress** | 0 | CD4 IL7R naive/memory-like | *IFITM2* | Mean expression | Evaluable | 9 | 6 | 1.333 | 1.499 | 1.283 | 1.513 | 15. | 0.1810 | 0.2778 | -0.4444 | -0.4444 | 0.9798 | 0.9462 | No | No | No |
| **ISG/stress** | 0 | CD4 IL7R naive/memory-like | *MX1* | % positive | Evaluable | 9 | 6 | 8.555 | 6.414 | 7.916 | 5.589 | 39. | 0.1810 | 0.7222 | 0.4444 | 0.4444 | 0.9798 | 0.9462 | No | No | No |
| **Cytotoxic** | 5 | preserved naive/memory-like | *NKG7* | % positive | Evaluable | 13 | 6 | 1.869 | 0.615 | 1.356 | 0.000 | 54. | 0.1864 | 0.6923 | 0.3846 | 0.3846 | 0.6085 | 0.9462 | No | No | No |
| **ISG/stress** | 1 | CD4 memory/activation-like | *HSPA1B* | Mean expression | Evaluable | 14 | 7 | 0.284 | 0.566 | 0.112 | 0.471 | 31. | 0.1916 | 0.3163 | -0.3673 | -0.3673 | 0.9798 | 0.9462 | No | No | No |
| **ISG/stress** | 1 | CD4 memory/activation-like | *HSPA1B* | % positive | Evaluable | 14 | 7 | 11.449 | 20.746 | 4.749 | 17.246 | 31. | 0.1916 | 0.3163 | -0.3673 | -0.3673 | 0.9798 | 0.9462 | No | No | No |

| | | | | | | | | | | | | | | | | | | | | | |
|---|---|---|---|---|---|---|---|---|---|---|---|---|---|---|---|---|---|---|---|---|---|
| **ISG/stress** | 0 | CD4 IL7R naive/memory-like | *OAS1* | % positive | Evaluable | 9 | 6 | 3.410 | 1.519 | 2.188 | 1.649 | 38.5 | 0.1945 | 0.7130 | 0.4259 | 0.4259 | 0.9798 | 0.9462 | No | No | No |
| **ISG/stress** | 1 | CD4 memory/activation-like | *HSPA1A* | Mean expression | Evaluable | 14 | 7 | 0.278 | 0.531 | 0.132 | 0.436 | 31. | 0.1969 | 0.3163 | -0.3673 | -0.3673 | 0.9798 | 0.9462 | No | No | No |
| **ISG/stress** | 1 | CD4 memory/activation-like | *MX1* | Mean expression | Evaluable | 14 | 7 | 0.112 | 0.076 | 0.096 | 0.068 | 67. | 0.1969 | 0.6837 | 0.3673 | 0.3673 | 0.9798 | 0.9462 | No | No | No |
| **Preserved/memory-like** | 1 | CD4 memory/activation-like | *TXK* | Mean expression | Evaluable | 14 | 7 | 0.553 | 0.455 | 0.491 | 0.399 | 67. | 0.1969 | 0.6837 | 0.3673 | 0.3673 | 0.7098 | 0.9462 | No | No | No |
| **ISG/stress** | 4 | CD8 naive/memory-like | *IFIT1* | % positive | Evaluable | 10 | 6 | 0.710 | 2.302 | 0.680 | 1.569 | 18. | 0.2003 | 0.3000 | -0.4000 | -0.4000 | 0.9798 | 0.9462 | No | No | No |
| **Cytotoxic** | 5 | preserved naive/memory-like | *CCL5* | Mean expression | Evaluable | 13 | 6 | 0.068 | 0.035 | 0.062 | 0.036 | 54. | 0.2027 | 0.6923 | 0.3846 | 0.3846 | 0.6085 | 0.9462 | No | No | No |
| **Cytotoxic** | 5 | preserved naive/memory-like | *KLRG1* | % positive | Evaluable | 13 | 6 | 9.315 | 6.293 | 8.824 | 6.854 | 54. | 0.2100 | 0.6923 | 0.3846 | 0.3846 | 0.6085 | 0.9462 | No | No | No |
| **Cytotoxic** | 4 | CD8 naive/memory-like | *GZMK* | % positive | Evaluable | 10 | 6 | 2.170 | 1.069 | 1.711 | 1.103 | 42. | 0.2109 | 0.7000 | 0.4000 | 0.4000 | 0.6085 | 0.9462 | No | No | No |
| **Cytotoxic** | 4 | CD8 naive/memory-like | *PRF1* | % positive | Evaluable | 10 | 6 | 5.409 | 3.215 | 5.035 | 3.331 | 42. | 0.2109 | 0.7000 | 0.4000 | 0.4000 | 0.6085 | 0.9462 | No | No | No |
| **Cytotoxic** | 0 | CD4 IL7R naive/memory-like | *NKG7* | % positive | Evaluable | 9 | 6 | 2.482 | 1.536 | 2.095 | 1.248 | 38. | 0.2155 | 0.7037 | 0.4074 | 0.4074 | 0.6085 | 0.9462 | No | No | No |
| **Preserved/memory-like** | 4 | CD8 naive/memory-like | *CD28* | Mean expression | Evaluable | 10 | 6 | 0.311 | 0.258 | 0.315 | 0.227 | 42. | 0.2198 | 0.7000 | 0.4000 | 0.4000 | 0.7098 | 0.9462 | No | No | No |
| **Preserved/memory-like** | 4 | CD8 naive/memory-like | *LTB* | % positive | Evaluable | 10 | 6 | 74.582 | 70.688 | 78.920 | 67.054 | 42. | 0.2198 | 0.7000 | 0.4000 | 0.4000 | 0.7098 | 0.9462 | No | No | No |
| **Preserved/memory-like** | 4 | CD8 naive/memory-like | *RUNX1* | % positive | Evaluable | 10 | 6 | 43.006 | 33.714 | 40.782 | 29.041 | 42. | 0.2198 | 0.7000 | 0.4000 | 0.4000 | 0.7098 | 0.9462 | No | No | No |
| **Preserved/memory-like** | 4 | CD8 naive/memory-like | *SKAP1* | % positive | Evaluable | 10 | 6 | 70.152 | 61.066 | 68.218 | 58.500 | 42. | 0.2198 | 0.7000 | 0.4000 | 0.4000 | 0.7098 | 0.9462 | No | No | No |
| **Checkpoint/activation** | 0 | CD4 IL7R naive/memory-like | *TOX* | % positive | Evaluable | 9 | 6 | 7.919 | 5.713 | 8.507 | 4.897 | 38. | 0.2238 | 0.7037 | 0.4074 | 0.4074 | 0.9627 | 0.9462 | No | No | No |
| **Cytotoxic** | 0 | CD4 IL7R naive/memory-like | *GZMK* | % positive | Evaluable | 9 | 6 | 2.659 | 1.844 | 3.070 | 1.420 | 38. | 0.2238 | 0.7037 | 0.4074 | 0.4074 | 0.6138 | 0.9462 | No | No | No |
| **ISG/stress** | 0 | CD4 IL7R naive/memory-like | *FOS* | Mean expression | Evaluable | 9 | 6 | 0.654 | 0.338 | 0.664 | 0.392 | 38. | 0.2238 | 0.7037 | 0.4074 | 0.4074 | 0.9798 | 0.9462 | No | No | No |
| **ISG/stress** | 0 | CD4 IL7R naive/memory-like | *FOS* | % positive | Evaluable | 9 | 6 | 31.554 | 17.635 | 34.483 | 20.450 | 38. | 0.2238 | 0.7037 | 0.4074 | 0.4074 | 0.9798 | 0.9462 | No | No | No |
| **Preserved/memory-like** | 0 | CD4 IL7R naive/memory-like | *IL7R* | % positive | Evaluable | 9 | 6 | 90.017 | 85.992 | 89.655 | 85.563 | 38. | 0.2238 | 0.7037 | 0.4074 | 0.4074 | 0.7098 | 0.9462 | No | No | No |
| **Preserved/memory-like** | 0 | CD4 IL7R naive/memory-like | *TCF7* | % positive | Evaluable | 9 | 6 | 63.806 | 53.311 | 63.912 | 55.401 | 38. | 0.2238 | 0.7037 | 0.4074 | 0.4074 | 0.7098 | 0.9462 | No | No | No |
| **Checkpoint/activation** | 1 | CD4 memory/activation-like | *TIGIT* | Mean expression | Evaluable | 14 | 7 | 0.122 | 0.088 | 0.118 | 0.087 | 66. | 0.2245 | 0.6735 | 0.3469 | 0.3469 | 0.9627 | 0.9462 | No | No | No |
| **ISG/stress** | 4 | CD8 naive/memory-like | *IFIT1* | Mean expression | Evaluable | 10 | 6 | 0.011 | 0.029 | 0.009 | 0.023 | 19. | 0.2423 | 0.3167 | -0.3667 | -0.3667 | 0.9798 | 0.9549 | No | No | No |
| **Checkpoint/activation** | 1 | CD4 memory/activation-like | *PDCD1* | % positive | Evaluable | 14 | 7 | 1.415 | 0.938 | 1.222 | 0.661 | 65. | 0.2474 | 0.6633 | 0.3265 | 0.3265 | 0.9627 | 0.9549 | No | No | No |
| **ISG/stress** | 4 | CD8 naive/memory-like | *HSPA1B* | Mean expression | Evaluable | 10 | 6 | 0.606 | 0.582 | 0.092 | 0.525 | 19. | 0.2635 | 0.3167 | -0.3667 | -0.3667 | 0.9798 | 0.9549 | No | No | No |
| **ISG/stress** | 4 | CD8 naive/memory-like | *IFITM1* | Mean expression | Evaluable | 10 | 6 | 2.294 | 2.460 | 2.299 | 2.543 | 19. | 0.2635 | 0.3167 | -0.3667 | -0.3667 | 0.9798 | 0.9549 | No | No | No |
| **ISG/stress** | 4 | CD8 naive/memory-like | *IFITM2* | Mean expression | Evaluable | 10 | 6 | 1.229 | 1.350 | 1.261 | 1.465 | 19. | 0.2635 | 0.3167 | -0.3667 | -0.3667 | 0.9798 | 0.9549 | No | No | No |
| **Preserved/memory-like** | 4 | CD8 naive/memory-like | *SKAP1* | Mean expression | Evaluable | 10 | 6 | 1.360 | 1.161 | 1.248 | 1.102 | 41. | 0.2635 | 0.6833 | 0.3667 | 0.3667 | 0.7098 | 0.9549 | No | No | No |
| **Checkpoint/activation** | 0 | CD4 IL7R naive/memory-like | *BTLA* | Mean expression | Evaluable | 9 | 6 | 0.095 | 0.081 | 0.110 | 0.084 | 37. | 0.2721 | 0.6852 | 0.3704 | 0.3704 | 0.9627 | 0.9549 | No | No | No |
| **Preserved/memory-like** | 0 | CD4 IL7R naive/memory-like | *FOXP1* | % positive | Evaluable | 9 | 6 | 73.766 | 60.830 | 72.992 | 59.210 | 37. | 0.2721 | 0.6852 | 0.3704 | 0.3704 | 0.7098 | 0.9549 | No | No | No |
| **Preserved/memory-like** | 0 | CD4 IL7R naive/memory-like | *LEF1* | % positive | Evaluable | 9 | 6 | 67.725 | 58.585 | 66.667 | 57.789 | 37. | 0.2721 | 0.6852 | 0.3704 | 0.3704 | 0.7098 | 0.9549 | No | No | No |
| **Preserved/memory-like** | 0 | CD4 IL7R naive/memory-like | *LTB* | Mean expression | Evaluable | 9 | 6 | 1.991 | 1.771 | 1.892 | 1.809 | 37. | 0.2721 | 0.6852 | 0.3704 | 0.3704 | 0.7098 | 0.9549 | No | No | No |
| **Preserved/memory-like** | 0 | CD4 IL7R naive/memory-like | *RUNX1* | % positive | Evaluable | 9 | 6 | 46.259 | 37.971 | 44.776 | 33.372 | 37. | 0.2721 | 0.6852 | 0.3704 | 0.3704 | 0.7098 | 0.9549 | No | No | No |
| **Cytotoxic** | 1 | CD4 memory/activation-like | *GZMH* | Mean expression | Evaluable | 14 | 7 | 0.011 | 0.005 | 0.009 | 0.004 | 64. | 0.2738 | 0.6531 | 0.3061 | 0.3061 | 0.7105 | 0.9549 | No | No | No |
| **Cytotoxic** | 1 | CD4 memory/activation-like | *GZMH* | % positive | Evaluable | 14 | 7 | 0.518 | 0.233 | 0.412 | 0.185 | 64. | 0.2738 | 0.6531 | 0.3061 | 0.3061 | 0.7105 | 0.9549 | No | No | No |
| **ISG/stress** | 5 | preserved naive/memory-like | *IFITM1* | Mean expression | Evaluable | 13 | 6 | 0.766 | 0.976 | 0.482 | 0.886 | 26. | 0.2818 | 0.3333 | -0.3333 | -0.3333 | 0.9798 | 0.9549 | No | No | No |
| **Preserved/memory-like** | 1 | CD4 memory/activation-like | *RUNX1* | Mean expression | Evaluable | 14 | 7 | 1.253 | 0.995 | 1.062 | 1.120 | 64. | 0.2872 | 0.6531 | 0.3061 | 0.3061 | 0.7098 | 0.9549 | No | No | No |
| **Checkpoint/activation** | 5 | preserved naive/memory-like | *HAVCR2* | % positive | Evaluable | 13 | 6 | 3.095 | 1.838 | 2.632 | 1.846 | 51. | 0.3129 | 0.6538 | 0.3077 | 0.3077 | 0.9627 | 0.9549 | No | No | No |
| **Cytotoxic** | 4 | CD8 naive/memory-like | *CCL5* | % positive | Evaluable | 10 | 6 | 12.618 | 18.389 | 11.321 | 18.230 | 20. | 0.3132 | 0.3333 | -0.3333 | -0.3333 | 0.7779 | 0.9549 | No | No | No |
| **ISG/stress** | 4 | CD8 naive/memory-like | *HSPA1A* | Mean expression | Evaluable | 10 | 6 | 0.545 | 0.522 | 0.084 | 0.369 | 20. | 0.3132 | 0.3333 | -0.3333 | -0.3333 | 0.9798 | 0.9549 | No | No | No |
| **ISG/stress** | 4 | CD8 naive/memory-like | *IFITM1* | % positive | Evaluable | 10 | 6 | 91.650 | 93.084 | 92.344 | 95.244 | 20. | 0.3132 | 0.3333 | -0.3333 | -0.3333 | 0.9798 | 0.9549 | No | No | No |
| **Preserved/memory-like** | 4 | CD8 naive/memory-like | *CD28* | % positive | Evaluable | 10 | 6 | 23.038 | 19.441 | 20.994 | 15.263 | 40. | 0.3132 | 0.6667 | 0.3333 | 0.3333 | 0.7098 | 0.9549 | No | No | No |
| **Preserved/memory-like** | 4 | CD8 naive/memory-like | *IL7R* | % positive | Evaluable | 10 | 6 | 91.720 | 87.729 | 92.839 | 88.115 | 40. | 0.3132 | 0.6667 | 0.3333 | 0.3333 | 0.7098 | 0.9549 | No | No | No |
| **Preserved/memory-like** | 4 | CD8 naive/memory-like | *RUNX1* | Mean expression | Evaluable | 10 | 6 | 0.716 | 0.551 | 0.665 | 0.491 | 40. | 0.3132 | 0.6667 | 0.3333 | 0.3333 | 0.7098 | 0.9549 | No | No | No |
| **Preserved/memory-like** | 4 | CD8 naive/memory-like | *SELL* | % positive | Evaluable | 10 | 6 | 67.284 | 61.334 | 66.092 | 58.000 | 40. | 0.3132 | 0.6667 | 0.3333 | 0.3333 | 0.7098 | 0.9549 | No | No | No |
| **Cytotoxic** | 0 | CD4 IL7R naive/memory-like | *NKG7* | Mean expression | Evaluable | 9 | 6 | 0.039 | 0.025 | 0.040 | 0.022 | 36. | 0.3160 | 0.6667 | 0.3333 | 0.3333 | 0.7779 | 0.9549 | No | No | No |
| **ISG/stress** | 1 | CD4 memory/activation-like | *HSPA1A* | % positive | Evaluable | 14 | 7 | 11.479 | 20.329 | 6.102 | 18.324 | 35. | 0.3223 | 0.3571 | -0.2857 | -0.2857 | 0.9798 | 0.9549 | No | No | No |
| **Preserved/memory-like** | 5 | preserved naive/memory-like | *CCR7* | % positive | Evaluable | 13 | 6 | 39.736 | 34.904 | 35.135 | 31.774 | 51. | 0.3229 | 0.6538 | 0.3077 | 0.3077 | 0.7098 | 0.9549 | No | No | No |
| **Preserved/memory-like** | 5 | preserved naive/memory-like | *CD27* | % positive | Evaluable | 13 | 6 | 10.715 | 14.001 | 8.108 | 15.166 | 27. | 0.3229 | 0.3462 | -0.3077 | -0.3077 | 0.7098 | 0.9549 | No | No | No |
| **Preserved/memory-like** | 5 | preserved naive/memory-like | *TXK* | % positive | Evaluable | 13 | 6 | 61.139 | 54.033 | 60.084 | 50.760 | 51. | 0.3229 | 0.6538 | 0.3077 | 0.3077 | 0.7098 | 0.9549 | No | No | No |
| **Checkpoint/activation** | 0 | CD4 IL7R naive/memory-like | *BTLA* | % positive | Evaluable | 9 | 6 | 6.975 | 5.265 | 7.389 | 5.230 | 36. | 0.3277 | 0.6667 | 0.3333 | 0.3333 | 0.9627 | 0.9549 | No | No | No |
| **ISG/stress** | 0 | CD4 IL7R naive/memory-like | *DUSP1* | Mean expression | Evaluable | 9 | 6 | 0.346 | 0.189 | 0.247 | 0.142 | 36. | 0.3277 | 0.6667 | 0.3333 | 0.3333 | 0.9798 | 0.9549 | No | No | No |
| **ISG/stress** | 0 | CD4 IL7R naive/memory-like | *DUSP1* | % positive | Evaluable | 9 | 6 | 20.382 | 10.645 | 15.373 | 8.405 | 36. | 0.3277 | 0.6667 | 0.3333 | 0.3333 | 0.9798 | 0.9549 | No | No | No |
| **Preserved/memory-like** | 0 | CD4 IL7R naive/memory-like | *CCR7* | Mean expression | Evaluable | 9 | 6 | 0.813 | 0.717 | 0.753 | 0.697 | 36. | 0.3277 | 0.6667 | 0.3333 | 0.3333 | 0.7098 | 0.9549 | No | No | No |
| **Preserved/memory-like** | 0 | CD4 IL7R naive/memory-like | *FOXP1* | Mean expression | Evaluable | 9 | 6 | 1.500 | 1.297 | 1.517 | 1.251 | 36. | 0.3277 | 0.6667 | 0.3333 | 0.3333 | 0.7098 | 0.9549 | No | No | No |
| **Preserved/memory-like** | 0 | CD4 IL7R naive/memory-like | *SKAP1* | % positive | Evaluable | 9 | 6 | 69.358 | 59.977 | 69.302 | 56.289 | 36. | 0.3277 | 0.6667 | 0.3333 | 0.3333 | 0.7098 | 0.9549 | No | No | No |
| **Checkpoint/activation** | 0 | CD4 IL7R naive/memory-like | *TNFRSF9* | Mean expression | Evaluable | 9 | 6 | 0.000 | 0.002 | 0.000 | 0.000 | 21. | 0.3541 | 0.3889 | -0.2222 | -0.2222 | 0.9627 | 0.9549 | No | No | No |
| **Checkpoint/activation** | 0 | CD4 IL7R naive/memory-like | *TNFRSF9* | % positive | Evaluable | 9 | 6 | 0.006 | 0.073 | 0.000 | 0.000 | 21. | 0.3541 | 0.3889 | -0.2222 | -0.2222 | 0.9627 | 0.9549 | No | No | No |
| **Cytotoxic** | 4 | CD8 naive/memory-like | *KLRD1* | Mean expression | Evaluable | 10 | 6 | 0.051 | 0.059 | 0.023 | 0.045 | 21. | 0.3562 | 0.3500 | -0.3000 | -0.3000 | 0.8286 | 0.9549 | No | No | No |
| **Cytotoxic** | 4 | CD8 naive/memory-like | *KLRD1* | % positive | Evaluable | 10 | 6 | 3.060 | 3.168 | 1.254 | 2.601 | 21. | 0.3562 | 0.3500 | -0.3000 | -0.3000 | 0.8286 | 0.9549 | No | No | No |
| **Preserved/memory-like** | 1 | CD4 memory/activation-like | *LTB* | Mean expression | Evaluable | 14 | 7 | 0.955 | 1.181 | 0.894 | 1.192 | 36. | 0.3601 | 0.3673 | -0.2653 | -0.2653 | 0.7098 | 0.9549 | No | No | No |
| **Preserved/memory-like** | 1 | CD4 memory/activation-like | *SKAP1* | % positive | Evaluable | 14 | 7 | 67.098 | 60.913 | 71.568 | 64.200 | 62. | 0.3601 | 0.6327 | 0.2653 | 0.2653 | 0.7098 | 0.9549 | No | No | No |
| **Checkpoint/activation** | 5 | preserved naive/memory-like | *LAG3* | Mean expression | Evaluable | 13 | 6 | 0.008 | 0.002 | 0.000 | 0.000 | 48. | 0.3662 | 0.6154 | 0.2308 | 0.2308 | 0.9627 | 0.9549 | No | No | No |
| **ISG/stress** | 5 | preserved naive/memory-like | *IFITM2* | Mean expression | Evaluable | 13 | 6 | 0.359 | 0.475 | 0.191 | 0.508 | 28. | 0.3676 | 0.3590 | -0.2821 | -0.2821 | 0.9798 | 0.9549 | No | No | No |
| **Preserved/memory-like** | 5 | preserved naive/memory-like | *CD27* | Mean expression | Evaluable | 13 | 6 | 0.195 | 0.259 | 0.190 | 0.308 | 28. | 0.3676 | 0.3590 | -0.2821 | -0.2821 | 0.7098 | 0.9549 | No | No | No |
| **ISG/stress** | 4 | CD8 naive/memory-like | *HSPA1B* | % positive | Evaluable | 10 | 6 | 25.837 | 26.394 | 5.608 | 25.283 | 21. | 0.3676 | 0.3500 | -0.3000 | -0.3000 | 0.9798 | 0.9549 | No | No | No |
| **Checkpoint/activation** | 0 | CD4 IL7R naive/memory-like | *PDCD1* | Mean expression | Evaluable | 9 | 6 | 0.017 | 0.005 | 0.013 | 0.005 | 35. | 0.3681 | 0.6481 | 0.2963 | 0.2963 | 0.9627 | 0.9549 | No | No | No |
| **Cytotoxic** | 0 | CD4 IL7R naive/memory-like | *KLRD1* | Mean expression | Evaluable | 9 | 6 | 0.008 | 0.006 | 0.007 | 0.006 | 35. | 0.3725 | 0.6481 | 0.2963 | 0.2963 | 0.8286 | 0.9549 | No | No | No |
| **Cytotoxic** | 0 | CD4 IL7R naive/memory-like | *KLRD1* | % positive | Evaluable | 9 | 6 | 0.575 | 0.352 | 0.466 | 0.342 | 35. | 0.3725 | 0.6481 | 0.2963 | 0.2963 | 0.8286 | 0.9549 | No | No | No |
| **Checkpoint/activation** | 0 | CD4 IL7R naive/memory-like | *CD69* | % positive | Evaluable | 9 | 6 | 29.736 | 20.252 | 22.637 | 19.860 | 35. | 0.3884 | 0.6481 | 0.2963 | 0.2963 | 0.9627 | 0.9549 | No | No | No |
| **Checkpoint/activation** | 0 | CD4 IL7R naive/memory-like | *TIGIT* | % positive | Evaluable | 9 | 6 | 5.531 | 3.919 | 4.598 | 3.624 | 35. | 0.3884 | 0.6481 | 0.2963 | 0.2963 | 0.9627 | 0.9549 | No | No | No |
| **Checkpoint/activation** | 0 | CD4 IL7R naive/memory-like | *TNFRSF4* | % positive | Evaluable | 9 | 6 | 8.549 | 5.635 | 6.242 | 4.603 | 35. | 0.3884 | 0.6481 | 0.2963 | 0.2963 | 0.9627 | 0.9549 | No | No | No |
| **Checkpoint/activation** | 0 | CD4 IL7R naive/memory-like | *TOX* | Mean expression | Evaluable | 9 | 6 | 0.132 | 0.098 | 0.140 | 0.084 | 35. | 0.3884 | 0.6481 | 0.2963 | 0.2963 | 0.9627 | 0.9549 | No | No | No |
| **Cytotoxic** | 0 | CD4 IL7R naive/memory-like | *CTSW* | % positive | Evaluable | 9 | 6 | 11.798 | 9.335 | 9.375 | 8.018 | 35. | 0.3884 | 0.6481 | 0.2963 | 0.2963 | 0.8286 | 0.9549 | No | No | No |
| **Cytotoxic** | 0 | CD4 IL7R naive/memory-like | *GZMK* | Mean expression | Evaluable | 9 | 6 | 0.048 | 0.036 | 0.051 | 0.030 | 35. | 0.3884 | 0.6481 | 0.2963 | 0.2963 | 0.8286 | 0.9549 | No | No | No |

| | | | | | | | | | | | | | | | | | | | | | |
|---|---|---|---|---|---|---|---|---|---|---|---|---|---|---|---|---|---|---|---|---|---|
| **ISG/stress** | 0 | CD4 IL7R naive/memory-like | *MX1* | Mean expression | Evaluable | 9 | 6 | 0.124 | 0.103 | 0.124 | 0.101 | 35. | 0.3884 | 0.6481 | 0.2963 | 0.2963 | 0.9798 | 0.9549 | No | No | No |
| **Cytotoxic** | 1 | CD4 memory/activation-like | *KLRG1* | Mean expression | Evaluable | 14 | 7 | 0.252 | 0.259 | 0.247 | 0.223 | 61. | 0.4003 | 0.6224 | 0.2449 | 0.2449 | 0.8353 | 0.9549 | No | No | No |
| **Preserved/memory-like** | 1 | CD4 memory/activation-like | *CD28* | Mean expression | Evaluable | 14 | 7 | 0.405 | 0.358 | 0.433 | 0.354 | 61. | 0.4003 | 0.6224 | 0.2449 | 0.2449 | 0.7098 | 0.9549 | No | No | No |
| **Preserved/memory-like** | 1 | CD4 memory/activation-like | *IL7R* | % positive | Evaluable | 14 | 7 | 71.259 | 77.432 | 72.351 | 81.384 | 37. | 0.4003 | 0.3776 | -0.2449 | -0.2449 | 0.7098 | 0.9549 | No | No | No |
| **Preserved/memory-like** | 1 | CD4 memory/activation-like | *RUNX1* | % positive | Evaluable | 14 | 7 | 49.663 | 41.993 | 47.460 | 43.079 | 61. | 0.4003 | 0.6224 | 0.2449 | 0.2449 | 0.7098 | 0.9549 | No | No | No |
| **ISG/stress** | 5 | preserved naive/memory-like | *IFITM1* | % positive | Evaluable | 13 | 6 | 32.028 | 39.874 | 24.242 | 37.162 | 29. | 0.4155 | 0.3718 | -0.2564 | -0.2564 | 0.9798 | 0.9549 | No | No | No |
| **Preserved/memory-like** | 5 | preserved naive/memory-like | *CD28* | Mean expression | Evaluable | 13 | 6 | 0.492 | 0.424 | 0.478 | 0.421 | 49. | 0.4155 | 0.6282 | 0.2564 | 0.2564 | 0.7098 | 0.9549 | No | No | No |
| **Preserved/memory-like** | 5 | preserved naive/memory-like | *LEF1* | Mean expression | Evaluable | 13 | 6 | 2.515 | 2.359 | 2.610 | 2.220 | 49. | 0.4155 | 0.6282 | 0.2564 | 0.2564 | 0.7098 | 0.9549 | No | No | No |
| **Preserved/memory-like** | 5 | preserved naive/memory-like | *LEF1* | % positive | Evaluable | 13 | 6 | 85.451 | 81.168 | 87.879 | 85.624 | 49. | 0.4155 | 0.6282 | 0.2564 | 0.2564 | 0.7098 | 0.9549 | No | No | No |
| **Cytotoxic** | 5 | preserved naive/memory-like | *CX3CR1* | Mean expression | Evaluable | 13 | 6 | 0.013 | 0.003 | 0.000 | 0.000 | 47. | 0.4253 | 0.6026 | 0.2051 | 0.2051 | 0.8577 | 0.9549 | No | No | No |
| **Checkpoint/activation** | 4 | CD8 naive/memory-like | *TOX* | Mean expression | Evaluable | 10 | 6 | 0.066 | 0.058 | 0.043 | 0.034 | 38. | 0.4278 | 0.6333 | 0.2667 | 0.2667 | 0.9627 | 0.9549 | No | No | No |
| **Preserved/memory-like** | 4 | CD8 naive/memory-like | *CCR7* | % positive | Evaluable | 10 | 6 | 61.820 | 55.380 | 62.555 | 50.027 | 38. | 0.4278 | 0.6333 | 0.2667 | 0.2667 | 0.7098 | 0.9549 | No | No | No |
| **Cytotoxic** | 0 | CD4 IL7R naive/memory-like | *GZMB* | Mean expression | Evaluable | 9 | 6 | 0.004 | 0.001 | 0.003 | 0.001 | 34. | 0.4289 | 0.6296 | 0.2593 | 0.2593 | 0.8577 | 0.9549 | No | No | No |
| **Checkpoint/activation** | 0 | CD4 IL7R naive/memory-like | *PDCD1* | % positive | Evaluable | 9 | 6 | 1.205 | 0.314 | 0.746 | 0.302 | 34. | 0.4353 | 0.6296 | 0.2593 | 0.2593 | 0.9627 | 0.9549 | No | No | No |
| **ISG/stress** | 0 | CD4 IL7R naive/memory-like | *IFIT3* | Mean expression | Evaluable | 9 | 6 | 0.015 | 0.012 | 0.011 | 0.013 | 34. | 0.4420 | 0.6296 | 0.2593 | 0.2593 | 0.9798 | 0.9549 | No | No | No |
| **ISG/stress** | 0 | CD4 IL7R naive/memory-like | *IFIT3* | % positive | Evaluable | 9 | 6 | 1.057 | 0.750 | 0.624 | 0.614 | 34. | 0.4420 | 0.6296 | 0.2593 | 0.2593 | 0.9798 | 0.9549 | No | No | No |
| **Checkpoint/activation** | 1 | CD4 memory/activation-like | *BTLA* | Mean expression | Evaluable | 14 | 7 | 0.081 | 0.089 | 0.072 | 0.096 | 38. | 0.4430 | 0.3878 | -0.2245 | -0.2245 | 0.9627 | 0.9549 | No | No | No |
| **Checkpoint/activation** | 1 | CD4 memory/activation-like | *TOX* | Mean expression | Evaluable | 14 | 7 | 0.402 | 0.317 | 0.342 | 0.326 | 60. | 0.4430 | 0.6122 | 0.2245 | 0.2245 | 0.9627 | 0.9549 | No | No | No |
| **Preserved/memory-like** | 1 | CD4 memory/activation-like | *IL7R* | Mean expression | Evaluable | 14 | 7 | 2.170 | 2.389 | 2.276 | 2.396 | 38. | 0.4430 | 0.3878 | -0.2245 | -0.2245 | 0.7098 | 0.9549 | No | No | No |
| **Preserved/memory-like** | 1 | CD4 memory/activation-like | *TXK* | % positive | Evaluable | 14 | 7 | 24.712 | 22.018 | 23.056 | 16.348 | 60. | 0.4430 | 0.6122 | 0.2245 | 0.2245 | 0.7098 | 0.9549 | No | No | No |
| **Checkpoint/activation** | 0 | CD4 IL7R naive/memory-like | *CD69* | Mean expression | Evaluable | 9 | 6 | 0.521 | 0.358 | 0.442 | 0.347 | 34. | 0.4559 | 0.6296 | 0.2593 | 0.2593 | 0.9627 | 0.9549 | No | No | No |
| **Checkpoint/activation** | 0 | CD4 IL7R naive/memory-like | *ICOS* | % positive | Evaluable | 9 | 6 | 22.058 | 18.584 | 17.070 | 16.239 | 34. | 0.4559 | 0.6296 | 0.2593 | 0.2593 | 0.9627 | 0.9549 | No | No | No |
| **Checkpoint/activation** | 0 | CD4 IL7R naive/memory-like | *TIGIT* | Mean expression | Evaluable | 9 | 6 | 0.085 | 0.064 | 0.083 | 0.065 | 34. | 0.4559 | 0.6296 | 0.2593 | 0.2593 | 0.9627 | 0.9549 | No | No | No |
| **Checkpoint/activation** | 0 | CD4 IL7R naive/memory-like | *TNFRSF4* | Mean expression | Evaluable | 9 | 6 | 0.128 | 0.093 | 0.105 | 0.075 | 34. | 0.4559 | 0.6296 | 0.2593 | 0.2593 | 0.9627 | 0.9549 | No | No | No |
| **Cytotoxic** | 0 | CD4 IL7R naive/memory-like | *KLRG1* | Mean expression | Evaluable | 9 | 6 | 0.192 | 0.154 | 0.169 | 0.139 | 34. | 0.4559 | 0.6296 | 0.2593 | 0.2593 | 0.8930 | 0.9549 | No | No | No |
| **ISG/stress** | 0 | CD4 IL7R naive/memory-like | *IFITM1* | Mean expression | Evaluable | 9 | 6 | 2.503 | 2.576 | 2.576 | 2.675 | 20. | 0.4559 | 0.3704 | -0.2593 | -0.2593 | 0.9798 | 0.9549 | No | No | No |
| **Preserved/memory-like** | 0 | CD4 IL7R naive/memory-like | *CD28* | % positive | Evaluable | 9 | 6 | 26.107 | 22.300 | 24.204 | 21.752 | 34. | 0.4559 | 0.6296 | 0.2593 | 0.2593 | 0.7098 | 0.9549 | No | No | No |
| **Preserved/memory-like** | 0 | CD4 IL7R naive/memory-like | *LEF1* | Mean expression | Evaluable | 9 | 6 | 1.374 | 1.211 | 1.297 | 1.187 | 34. | 0.4559 | 0.6296 | 0.2593 | 0.2593 | 0.7098 | 0.9549 | No | No | No |
| **Preserved/memory-like** | 0 | CD4 IL7R naive/memory-like | *RUNX1* | Mean expression | Evaluable | 9 | 6 | 0.793 | 0.685 | 0.735 | 0.658 | 34. | 0.4559 | 0.6296 | 0.2593 | 0.2593 | 0.7098 | 0.9549 | No | No | No |
| **Preserved/memory-like** | 0 | CD4 IL7R naive/memory-like | *SKAP1* | Mean expression | Evaluable | 9 | 6 | 1.338 | 1.214 | 1.279 | 1.230 | 34. | 0.4559 | 0.6296 | 0.2593 | 0.2593 | 0.7098 | 0.9549 | No | No | No |
| **Preserved/memory-like** | 0 | CD4 IL7R naive/memory-like | *TCF7* | Mean expression | Evaluable | 9 | 6 | 1.150 | 1.020 | 1.121 | 1.067 | 34. | 0.4559 | 0.6296 | 0.2593 | 0.2593 | 0.7098 | 0.9549 | No | No | No |
| **Checkpoint/activation** | 5 | preserved naive/memory-like | *ICOS* | % positive | Evaluable | 13 | 6 | 21.086 | 15.471 | 15.709 | 14.926 | 48. | 0.4670 | 0.6154 | 0.2308 | 0.2308 | 0.9627 | 0.9549 | No | No | No |
| **Preserved/memory-like** | 5 | preserved naive/memory-like | *CCR7* | Mean expression | Evaluable | 13 | 6 | 0.850 | 0.745 | 0.792 | 0.733 | 48. | 0.4670 | 0.6154 | 0.2308 | 0.2308 | 0.7098 | 0.9549 | No | No | No |
| **Checkpoint/activation** | 1 | CD4 memory/activation-like | *TNFRSF9* | % positive | Evaluable | 14 | 7 | 0.519 | 0.601 | 0.111 | 0.185 | 39. | 0.4703 | 0.3980 | -0.2041 | -0.2041 | 0.9627 | 0.9549 | No | No | No |
| **Checkpoint/activation** | 0 | CD4 IL7R naive/memory-like | *LAG3* | % positive | Evaluable | 9 | 6 | 0.728 | 0.445 | 0.607 | 0.502 | 33.5 | 0.4755 | 0.6204 | 0.2407 | 0.2407 | 0.9627 | 0.9549 | No | No | No |
| **Cytotoxic** | 4 | CD8 naive/memory-like | *GZMK* | Mean expression | Evaluable | 10 | 6 | 0.034 | 0.019 | 0.025 | 0.021 | 37. | 0.4795 | 0.6167 | 0.2333 | 0.2333 | 0.8930 | 0.9549 | No | No | No |
| **ISG/stress** | 5 | preserved naive/memory-like | *HSPA1B* | Mean expression | Evaluable | 13 | 6 | 0.359 | 0.234 | 0.055 | 0.225 | 30.5 | 0.4810 | 0.3910 | -0.2179 | -0.2179 | 0.9798 | 0.9549 | No | No | No |
| **Preserved/memory-like** | 5 | preserved naive/memory-like | *SELL* | % positive | Evaluable | 13 | 6 | 38.470 | 40.862 | 40.845 | 43.226 | 30.5 | 0.4827 | 0.3910 | -0.2179 | -0.2179 | 0.7098 | 0.9549 | No | No | No |
| **Checkpoint/activation** | 1 | CD4 memory/activation-like | *TIGIT* | % positive | Evaluable | 14 | 7 | 6.225 | 4.593 | 5.652 | 4.165 | 59. | 0.4880 | 0.6020 | 0.2041 | 0.2041 | 0.9627 | 0.9549 | No | No | No |
| **Preserved/memory-like** | 1 | CD4 memory/activation-like | *CD27* | Mean expression | Evaluable | 14 | 7 | 0.214 | 0.239 | 0.161 | 0.211 | 39. | 0.4880 | 0.3980 | -0.2041 | -0.2041 | 0.7098 | 0.9549 | No | No | No |
| **Preserved/memory-like** | 1 | CD4 memory/activation-like | *CD28* | % positive | Evaluable | 14 | 7 | 19.884 | 17.716 | 22.979 | 17.564 | 59. | 0.4880 | 0.6020 | 0.2041 | 0.2041 | 0.7098 | 0.9549 | No | No | No |
| **Preserved/memory-like** | 1 | CD4 memory/activation-like | *SKAP1* | Mean expression | Evaluable | 14 | 7 | 1.832 | 1.575 | 1.693 | 1.675 | 59. | 0.4880 | 0.6020 | 0.2041 | 0.2041 | 0.7098 | 0.9549 | No | No | No |
| **Checkpoint/activation** | 5 | preserved naive/memory-like | *LAG3* | % positive | Evaluable | 13 | 6 | 0.472 | 0.118 | 0.000 | 0.000 | 46. | 0.4896 | 0.5897 | 0.1795 | 0.1795 | 0.9627 | 0.9549 | No | No | No |
| **Cytotoxic** | 5 | preserved naive/memory-like | *CX3CR1* | % positive | Evaluable | 13 | 6 | 0.565 | 0.225 | 0.000 | 0.000 | 46. | 0.4896 | 0.5897 | 0.1795 | 0.1795 | 0.8930 | 0.9549 | No | No | No |
| **Cytotoxic** | 4 | CD8 naive/memory-like | *KLRG1* | Mean expression | Evaluable | 10 | 6 | 0.110 | 0.135 | 0.097 | 0.111 | 23. | 0.4923 | 0.3833 | -0.2333 | -0.2333 | 0.8930 | 0.9549 | No | No | No |
| **ISG/stress** | 4 | CD8 naive/memory-like | *HSPA1A* | % positive | Evaluable | 10 | 6 | 23.023 | 23.682 | 5.277 | 17.863 | 23. | 0.4923 | 0.3833 | -0.2333 | -0.2333 | 0.9798 | 0.9549 | No | No | No |
| **ISG/stress** | 4 | CD8 naive/memory-like | *IFIT2* | Mean expression | Evaluable | 10 | 6 | 0.043 | 0.046 | 0.032 | 0.041 | 23. | 0.4923 | 0.3833 | -0.2333 | -0.2333 | 0.9798 | 0.9549 | No | No | No |
| **ISG/stress** | 4 | CD8 naive/memory-like | *MX1* | Mean expression | Evaluable | 10 | 6 | 0.084 | 0.126 | 0.097 | 0.107 | 23. | 0.4923 | 0.3833 | -0.2333 | -0.2333 | 0.9798 | 0.9549 | No | No | No |
| **Cytotoxic** | 4 | CD8 naive/memory-like | *GZMH* | Mean expression | Evaluable | 10 | 6 | 0.007 | 0.001 | 0.000 | 0.000 | 36. | 0.4930 | 0.6000 | 0.2000 | 0.2000 | 0.8930 | 0.9549 | No | No | No |
| **Checkpoint/activation** | 5 | preserved naive/memory-like | *TNFRSF4* | Mean expression | Evaluable | 13 | 6 | 0.016 | 0.041 | 0.000 | 0.017 | 31.5 | 0.5068 | 0.4038 | -0.1923 | -0.1923 | 0.9627 | 0.9549 | No | No | No |
| **ISG/stress** | 4 | CD8 naive/memory-like | *OAS1* | Mean expression | Evaluable | 10 | 6 | 0.018 | 0.024 | 0.017 | 0.027 | 23.5 | 0.5149 | 0.3917 | -0.2167 | -0.2167 | 0.9798 | 0.9549 | No | No | No |
| **Checkpoint/activation** | 1 | CD4 memory/activation-like | *TNFRSF9* | Mean expression | Evaluable | 14 | 7 | 0.009 | 0.008 | 0.002 | 0.004 | 40. | 0.5183 | 0.4082 | -0.1837 | -0.1837 | 0.9627 | 0.9549 | No | No | No |
| **Checkpoint/activation** | 5 | preserved naive/memory-like | *BTLA* | Mean expression | Evaluable | 13 | 6 | 0.135 | 0.124 | 0.131 | 0.107 | 47. | 0.5214 | 0.6026 | 0.2051 | 0.2051 | 0.9627 | 0.9549 | No | No | No |
| **ISG/stress** | 5 | preserved naive/memory-like | *DUSP1* | % positive | Evaluable | 13 | 6 | 8.539 | 3.098 | 2.632 | 1.351 | 46.5 | 0.5235 | 0.5962 | 0.1923 | 0.1923 | 0.9798 | 0.9549 | No | No | No |
| **Checkpoint/activation** | 0 | CD4 IL7R naive/memory-like | *CTLA4* | Mean expression | Evaluable | 9 | 6 | 0.048 | 0.037 | 0.044 | 0.036 | 33. | 0.5287 | 0.6111 | 0.2222 | 0.2222 | 0.9627 | 0.9549 | No | No | No |
| **ISG/stress** | 0 | CD4 IL7R naive/memory-like | *ISG15* | % positive | Evaluable | 9 | 6 | 13.394 | 11.185 | 12.806 | 11.960 | 33. | 0.5287 | 0.6111 | 0.2222 | 0.2222 | 0.9798 | 0.9549 | No | No | No |
| **ISG/stress** | 0 | CD4 IL7R naive/memory-like | *JUNB* | Mean expression | Evaluable | 9 | 6 | 0.687 | 0.644 | 0.723 | 0.748 | 33. | 0.5287 | 0.6111 | 0.2222 | 0.2222 | 0.9798 | 0.9549 | No | No | No |
| **ISG/stress** | 0 | CD4 IL7R naive/memory-like | *JUNB* | % positive | Evaluable | 9 | 6 | 33.948 | 29.463 | 36.269 | 33.876 | 33. | 0.5287 | 0.6111 | 0.2222 | 0.2222 | 0.9798 | 0.9549 | No | No | No |
| **Preserved/memory-like** | 0 | CD4 IL7R naive/memory-like | *CCR7* | % positive | Evaluable | 9 | 6 | 45.060 | 38.775 | 41.194 | 38.281 | 33. | 0.5287 | 0.6111 | 0.2222 | 0.2222 | 0.7464 | 0.9549 | No | No | No |
| **Preserved/memory-like** | 0 | CD4 IL7R naive/memory-like | *SELL* | % positive | Evaluable | 9 | 6 | 57.877 | 52.296 | 55.646 | 50.927 | 33. | 0.5287 | 0.6111 | 0.2222 | 0.2222 | 0.7464 | 0.9549 | No | No | No |
| **Checkpoint/activation** | 1 | CD4 memory/activation-like | *ICOS* | % positive | Evaluable | 14 | 7 | 12.492 | 10.862 | 11.797 | 8.721 | 58. | 0.5353 | 0.5918 | 0.1837 | 0.1837 | 0.9627 | 0.9549 | No | No | No |
| **Cytotoxic** | 1 | CD4 memory/activation-like | *CCL5* | Mean expression | Evaluable | 14 | 7 | 0.396 | 0.396 | 0.435 | 0.300 | 58. | 0.5353 | 0.5918 | 0.1837 | 0.1837 | 0.9124 | 0.9549 | No | No | No |
| **Cytotoxic** | 1 | CD4 memory/activation-like | *KLRG1* | % positive | Evaluable | 14 | 7 | 12.380 | 12.856 | 13.002 | 10.859 | 58. | 0.5353 | 0.5918 | 0.1837 | 0.1837 | 0.9124 | 0.9549 | No | No | No |
| **Cytotoxic** | 1 | CD4 memory/activation-like | *NKG7* | % positive | Evaluable | 14 | 7 | 5.132 | 5.055 | 5.296 | 4.092 | 58. | 0.5353 | 0.5918 | 0.1837 | 0.1837 | 0.9124 | 0.9549 | No | No | No |
| **ISG/stress** | 1 | CD4 memory/activation-like | *IFIT2* | Mean expression | Evaluable | 14 | 7 | 0.028 | 0.023 | 0.026 | 0.021 | 58. | 0.5353 | 0.5918 | 0.1837 | 0.1837 | 0.9798 | 0.9549 | No | No | No |
| **Checkpoint/activation** | 5 | preserved naive/memory-like | *TIGIT* | Mean expression | Evaluable | 13 | 6 | 0.045 | 0.060 | 0.051 | 0.056 | 31.5 | 0.5391 | 0.4038 | -0.1923 | -0.1923 | 0.9627 | 0.9549 | No | No | No |
| **Checkpoint/activation** | 4 | CD8 naive/memory-like | *TOX* | % positive | Evaluable | 10 | 6 | 3.904 | 3.307 | 2.853 | 2.403 | 36. | 0.5622 | 0.6000 | 0.2000 | 0.2000 | 0.9627 | 0.9549 | No | No | No |
| **ISG/stress** | 4 | CD8 naive/memory-like | *JUN* | Mean expression | Evaluable | 10 | 6 | 0.602 | 0.534 | 0.142 | 0.390 | 24. | 0.5622 | 0.4000 | -0.2000 | -0.2000 | 0.9798 | 0.9549 | No | No | No |
| **ISG/stress** | 4 | CD8 naive/memory-like | *JUN* | % positive | Evaluable | 10 | 6 | 28.297 | 25.921 | 8.405 | 20.180 | 24. | 0.5622 | 0.4000 | -0.2000 | -0.2000 | 0.9798 | 0.9549 | No | No | No |
| **ISG/stress** | 4 | CD8 naive/memory-like | *JUNB* | Mean expression | Evaluable | 10 | 6 | 0.936 | 0.889 | 0.472 | 0.821 | 24. | 0.5622 | 0.4000 | -0.2000 | -0.2000 | 0.9798 | 0.9549 | No | No | No |
| **Checkpoint/activation** | 5 | preserved naive/memory-like | *TIGIT* | % positive | Evaluable | 13 | 6 | 2.381 | 3.185 | 2.326 | 2.678 | 32. | 0.5681 | 0.4103 | -0.1795 | -0.1795 | 0.9627 | 0.9549 | No | No | No |
| **Checkpoint/activation** | 5 | preserved naive/memory-like | *TNFRSF4* | % positive | Evaluable | 13 | 6 | 0.872 | 1.708 | 0.000 | 0.676 | 32.5 | 0.5693 | 0.4167 | -0.1667 | -0.1667 | 0.9627 | 0.9549 | No | No | No |
| **Checkpoint/activation** | 1 | CD4 memory/activation-like | *LAG3* | Mean expression | Evaluable | 14 | 7 | 0.019 | 0.013 | 0.017 | 0.013 | 57. | 0.5746 | 0.5816 | 0.1633 | 0.1633 | 0.9627 | 0.9549 | No | No | No |
| **Checkpoint/activation** | 1 | CD4 memory/activation-like | *LAG3* | % positive | Evaluable | 14 | 7 | 0.820 | 0.660 | 0.733 | 0.676 | 57. | 0.5746 | 0.5816 | 0.1633 | 0.1633 | 0.9627 | 0.9549 | No | No | No |

| | | | | | | | | | | | | | | | | | | | | | |
|---|---|---|---|---|---|---|---|---|---|---|---|---|---|---|---|---|---|---|---|---|---|
| **Cytotoxic** | 4 | CD8 naive/memory-like | *CX3CR1* | Mean expression | Evaluable | 10 | 6 | 0.008 | 0.001 | 0.000 | 0.000 | 35. | 0.5749 | 0.5833 | 0.1667 | 0.1667 | 0.9124 | 0.9549 | No | No | No |
| **Cytotoxic** | 4 | CD8 naive/memory-like | *CX3CR1* | % positive | Evaluable | 10 | 6 | 0.611 | 0.112 | 0.000 | 0.000 | 35. | 0.5749 | 0.5833 | 0.1667 | 0.1667 | 0.9124 | 0.9549 | No | No | No |
| **Cytotoxic** | 4 | CD8 naive/memory-like | *GZMH* | % positive | Evaluable | 10 | 6 | 0.504 | 0.120 | 0.000 | 0.000 | 35. | 0.5749 | 0.5833 | 0.1667 | 0.1667 | 0.9124 | 0.9549 | No | No | No |
| **Checkpoint/activation** | 5 | preserved naive/memory-like | *CD69* | Mean expression | Evaluable | 13 | 6 | 0.419 | 0.311 | 0.147 | 0.269 | 32. | 0.5789 | 0.4103 | -0.1795 | -0.1795 | 0.9627 | 0.9549 | No | No | No |
| **Checkpoint/activation** | 5 | preserved naive/memory-like | *ICOS* | Mean expression | Evaluable | 13 | 6 | 0.422 | 0.331 | 0.323 | 0.325 | 46. | 0.5789 | 0.5897 | 0.1795 | 0.1795 | 0.9627 | 0.9549 | No | No | No |
| **Preserved/memory-like** | 5 | preserved naive/memory-like | *CD28* | % positive | Evaluable | 13 | 6 | 24.925 | 21.170 | 21.875 | 23.138 | 46. | 0.5789 | 0.5897 | 0.1795 | 0.1795 | 0.7827 | 0.9549 | No | No | No |
| **Preserved/memory-like** | 5 | preserved naive/memory-like | *LTB* | Mean expression | Evaluable | 13 | 6 | 0.560 | 0.669 | 0.463 | 0.532 | 32. | 0.5789 | 0.4103 | -0.1795 | -0.1795 | 0.7827 | 0.9549 | No | No | No |
| **Preserved/memory-like** | 5 | preserved naive/memory-like | *SKAP1* | Mean expression | Evaluable | 13 | 6 | 2.268 | 2.214 | 2.288 | 2.188 | 46. | 0.5789 | 0.5897 | 0.1795 | 0.1795 | 0.7827 | 0.9549 | No | No | No |
| **Cytotoxic** | 0 | CD4 IL7R naive/memory-like | *FGFBP2* | Mean expression | Evaluable | 9 | 6 | 0.003 | 0.002 | 0.002 | 0.001 | 32. | 0.5839 | 0.5926 | 0.1852 | 0.1852 | 0.9124 | 0.9549 | No | No | No |
| **Cytotoxic** | 0 | CD4 IL7R naive/memory-like | *GZMB* | % positive | Evaluable | 9 | 6 | 0.281 | 0.089 | 0.149 | 0.065 | 32. | 0.5839 | 0.5926 | 0.1852 | 0.1852 | 0.9124 | 0.9549 | No | No | No |
| **ISG/stress** | 5 | preserved naive/memory-like | *DUSP1* | Mean expression | Evaluable | 13 | 6 | 0.158 | 0.060 | 0.043 | 0.033 | 45.5 | 0.5845 | 0.5833 | 0.1667 | 0.1667 | 0.9798 | 0.9549 | No | No | No |
| **Checkpoint/activation** | 1 | CD4 memory/activation-like | *ICOS* | Mean expression | Evaluable | 14 | 7 | 0.250 | 0.225 | 0.234 | 0.208 | 57. | 0.5846 | 0.5816 | 0.1633 | 0.1633 | 0.9627 | 0.9549 | No | No | No |
| **Checkpoint/activation** | 1 | CD4 memory/activation-like | *TOX* | % positive | Evaluable | 14 | 7 | 16.381 | 13.523 | 14.894 | 14.320 | 57. | 0.5846 | 0.5816 | 0.1633 | 0.1633 | 0.9627 | 0.9549 | No | No | No |
| **Checkpoint/activation** | 4 | CD8 naive/memory-like | *CTLA4* | Mean expression | Evaluable | 10 | 6 | 0.040 | 0.023 | 0.027 | 0.015 | 35.5 | 0.5873 | 0.5917 | 0.1833 | 0.1833 | 0.9627 | 0.9549 | No | No | No |
| **Checkpoint/activation** | 5 | preserved naive/memory-like | *BTLA* | % positive | Evaluable | 13 | 6 | 6.911 | 6.434 | 7.042 | 6.076 | 45.5 | 0.5986 | 0.5833 | 0.1667 | 0.1667 | 0.9627 | 0.9549 | No | No | No |
| **ISG/stress** | 5 | preserved naive/memory-like | *IFITM2* | % positive | Evaluable | 13 | 6 | 18.215 | 22.257 | 9.859 | 22.552 | 32.5 | 0.5986 | 0.4167 | -0.1667 | -0.1667 | 0.9798 | 0.9549 | No | No | No |
| **Checkpoint/activation** | 0 | CD4 IL7R naive/memory-like | *CTLA4* | % positive | Evaluable | 9 | 6 | 3.297 | 2.368 | 2.394 | 2.219 | 32. | 0.6070 | 0.5926 | 0.1852 | 0.1852 | 0.9627 | 0.9549 | No | No | No |
| **Cytotoxic** | 0 | CD4 IL7R naive/memory-like | *CCL5* | % positive | Evaluable | 9 | 6 | 8.021 | 5.072 | 6.886 | 5.044 | 32. | 0.6070 | 0.5926 | 0.1852 | 0.1852 | 0.9124 | 0.9549 | No | No | No |
| **ISG/stress** | 0 | CD4 IL7R naive/memory-like | *HSPA1A* | Mean expression | Evaluable | 9 | 6 | 0.460 | 0.589 | 0.178 | 0.704 | 22. | 0.6070 | 0.4074 | -0.1852 | -0.1852 | 0.9798 | 0.9549 | No | No | No |
| **ISG/stress** | 0 | CD4 IL7R naive/memory-like | *IFIT2* | % positive | Evaluable | 9 | 6 | 3.596 | 2.787 | 2.675 | 2.623 | 32. | 0.6070 | 0.5926 | 0.1852 | 0.1852 | 0.9798 | 0.9549 | No | No | No |
| **ISG/stress** | 0 | CD4 IL7R naive/memory-like | *IFITM2* | % positive | Evaluable | 9 | 6 | 67.481 | 68.872 | 65.658 | 68.535 | 22. | 0.6070 | 0.4074 | -0.1852 | -0.1852 | 0.9798 | 0.9549 | No | No | No |
| **Preserved/memory-like** | 0 | CD4 IL7R naive/memory-like | *CD27* | % positive | Evaluable | 9 | 6 | 33.673 | 30.206 | 33.178 | 32.362 | 32. | 0.6070 | 0.5926 | 0.1852 | 0.1852 | 0.8093 | 0.9549 | No | No | No |
| **Cytotoxic** | 5 | preserved naive/memory-like | *KLRD1* | Mean expression | Evaluable | 13 | 6 | 0.017 | 0.017 | 0.014 | 0.000 | 45. | 0.6101 | 0.5769 | 0.1538 | 0.1538 | 0.9124 | 0.9549 | No | No | No |
| **Checkpoint/activation** | 4 | CD8 naive/memory-like | *PDCD1* | Mean expression | Evaluable | 10 | 6 | 0.003 | 0.003 | 0.000 | 0.000 | 26. | 0.6180 | 0.4333 | -0.1333 | -0.1333 | 0.9627 | 0.9549 | No | No | No |
| **ISG/stress** | 4 | CD8 naive/memory-like | *IFIT3* | Mean expression | Evaluable | 10 | 6 | 0.002 | 0.006 | 0.000 | 0.001 | 25.5 | 0.6181 | 0.4250 | -0.1500 | -0.1500 | 0.9798 | 0.9549 | No | No | No |
| **ISG/stress** | 4 | CD8 naive/memory-like | *IFIT3* | % positive | Evaluable | 10 | 6 | 0.143 | 0.535 | 0.000 | 0.099 | 25.5 | 0.6181 | 0.4250 | -0.1500 | -0.1500 | 0.9798 | 0.9549 | No | No | No |
| **Cytotoxic** | 5 | preserved naive/memory-like | *PRF1* | Mean expression | Evaluable | 13 | 6 | 0.018 | 0.029 | 0.011 | 0.012 | 33. | 0.6208 | 0.4231 | -0.1538 | -0.1538 | 0.9124 | 0.9549 | No | No | No |
| **ISG/stress** | 5 | preserved naive/memory-like | *FOS* | % positive | Evaluable | 13 | 6 | 16.550 | 5.356 | 5.402 | 5.037 | 45. | 0.6242 | 0.5769 | 0.1538 | 0.1538 | 0.9798 | 0.9549 | No | No | No |
| **ISG/stress** | 5 | preserved naive/memory-like | *IFIT2* | Mean expression | Evaluable | 13 | 6 | 0.035 | 0.029 | 0.033 | 0.022 | 45. | 0.6265 | 0.5769 | 0.1538 | 0.1538 | 0.9798 | 0.9549 | No | No | No |
| **ISG/stress** | 1 | CD4 memory/activation-like | *JUN* | % positive | Evaluable | 14 | 7 | 8.057 | 9.348 | 3.188 | 5.631 | 42. | 0.6276 | 0.4286 | -0.1429 | -0.1429 | 0.9798 | 0.9549 | No | No | No |
| **ISG/stress** | 5 | preserved naive/memory-like | *ISG15* | % positive | Evaluable | 13 | 6 | 3.595 | 2.741 | 2.632 | 3.059 | 45. | 0.6280 | 0.5769 | 0.1538 | 0.1538 | 0.9798 | 0.9549 | No | No | No |
| **Cytotoxic** | 5 | preserved naive/memory-like | *CTSW* | % positive | Evaluable | 13 | 6 | 3.483 | 4.383 | 2.373 | 3.567 | 33. | 0.6292 | 0.4231 | -0.1538 | -0.1538 | 0.9124 | 0.9549 | No | No | No |
| **ISG/stress** | 4 | CD8 naive/memory-like | *FOS* | Mean expression | Evaluable | 10 | 6 | 0.835 | 0.534 | 0.426 | 0.353 | 35. | 0.6354 | 0.5833 | 0.1667 | 0.1667 | 0.9798 | 0.9549 | No | No | No |
| **ISG/stress** | 4 | CD8 naive/memory-like | *JUNB* | % positive | Evaluable | 10 | 6 | 40.573 | 39.047 | 23.868 | 37.308 | 25. | 0.6354 | 0.4167 | -0.1667 | -0.1667 | 0.9798 | 0.9549 | No | No | No |
| **Checkpoint/activation** | 1 | CD4 memory/activation-like | *TNFRSF4* | Mean expression | Evaluable | 14 | 7 | 0.156 | 0.179 | 0.146 | 0.192 | 42. | 0.6359 | 0.4286 | -0.1429 | -0.1429 | 0.9627 | 0.9549 | No | No | No |
| **Preserved/memory-like** | 1 | CD4 memory/activation-like | *CD27* | % positive | Evaluable | 14 | 7 | 11.563 | 12.485 | 8.324 | 11.017 | 42. | 0.6359 | 0.4286 | -0.1429 | -0.1429 | 0.8287 | 0.9549 | No | No | No |
| **Checkpoint/activation** | 5 | preserved naive/memory-like | *CTLA4* | Mean expression | Evaluable | 13 | 6 | 0.056 | 0.044 | 0.037 | 0.043 | 33. | 0.6388 | 0.4231 | -0.1538 | -0.1538 | 0.9627 | 0.9549 | No | No | No |
| **ISG/stress** | 5 | preserved naive/memory-like | *JUNB* | Mean expression | Evaluable | 13 | 6 | 0.306 | 0.222 | 0.160 | 0.279 | 33. | 0.6388 | 0.4231 | -0.1538 | -0.1538 | 0.9798 | 0.9549 | No | No | No |
| **Preserved/memory-like** | 5 | preserved naive/memory-like | *SELL* | Mean expression | Evaluable | 13 | 6 | 0.828 | 0.906 | 0.855 | 0.961 | 33. | 0.6388 | 0.4231 | -0.1538 | -0.1538 | 0.8287 | 0.9549 | No | No | No |
| **ISG/stress** | 5 | preserved naive/memory-like | *IFIT3* | Mean expression | Evaluable | 13 | 6 | 0.013 | 0.007 | 0.000 | 0.000 | 44. | 0.6484 | 0.5641 | 0.1282 | 0.1282 | 0.9798 | 0.9549 | No | No | No |
| **ISG/stress** | 5 | preserved naive/memory-like | *HSPA1B* | % positive | Evaluable | 13 | 6 | 14.780 | 10.453 | 3.361 | 8.436 | 33.5 | 0.6596 | 0.4295 | -0.1410 | -0.1410 | 0.9798 | 0.9549 | No | No | No |
| **Cytotoxic** | 5 | preserved naive/memory-like | *CTSW* | Mean expression | Evaluable | 13 | 6 | 0.066 | 0.097 | 0.052 | 0.076 | 33.5 | 0.6609 | 0.4295 | -0.1410 | -0.1410 | 0.9124 | 0.9549 | No | No | No |
| **Checkpoint/activation** | 4 | CD8 naive/memory-like | *CTLA4* | % positive | Evaluable | 10 | 6 | 3.464 | 1.767 | 1.777 | 1.020 | 34.5 | 0.6642 | 0.5750 | 0.1500 | 0.1500 | 0.9627 | 0.9549 | No | No | No |
| **Cytotoxic** | 0 | CD4 IL7R naive/memory-like | *FGFBP2* | % positive | Evaluable | 9 | 6 | 0.225 | 0.091 | 0.127 | 0.074 | 31. | 0.6701 | 0.5741 | 0.1481 | 0.1481 | 0.9124 | 0.9549 | No | No | No |
| **Cytotoxic** | 0 | CD4 IL7R naive/memory-like | *GZMH* | % positive | Evaluable | 9 | 6 | 0.333 | 0.172 | 0.382 | 0.100 | 31. | 0.6745 | 0.5741 | 0.1481 | 0.1481 | 0.9124 | 0.9549 | No | No | No |
| **Cytotoxic** | 5 | preserved naive/memory-like | *KLRD1* | % positive | Evaluable | 13 | 6 | 0.723 | 0.741 | 0.678 | 0.000 | 44. | 0.6765 | 0.5641 | 0.1282 | 0.1282 | 0.9124 | 0.9549 | No | No | No |
| **ISG/stress** | 1 | CD4 memory/activation-like | *IFIT1* | Mean expression | Evaluable | 14 | 7 | 0.016 | 0.019 | 0.015 | 0.014 | 43. | 0.6806 | 0.4388 | -0.1224 | -0.1224 | 0.9798 | 0.9549 | No | No | No |
| **ISG/stress** | 1 | CD4 memory/activation-like | *IFIT1* | % positive | Evaluable | 14 | 7 | 0.762 | 1.072 | 0.785 | 0.686 | 43. | 0.6806 | 0.4388 | -0.1224 | -0.1224 | 0.9798 | 0.9549 | No | No | No |
| **ISG/stress** | 5 | preserved naive/memory-like | *FOS* | Mean expression | Evaluable | 13 | 6 | 0.408 | 0.108 | 0.085 | 0.102 | 44. | 0.6885 | 0.5641 | 0.1282 | 0.1282 | 0.9798 | 0.9549 | No | No | No |
| **Cytotoxic** | 1 | CD4 memory/activation-like | *GZMK* | % positive | Evaluable | 14 | 7 | 3.701 | 3.725 | 3.566 | 2.588 | 55. | 0.6888 | 0.5612 | 0.1224 | 0.1224 | 0.9124 | 0.9549 | No | No | No |
| **Cytotoxic** | 1 | CD4 memory/activation-like | *PRF1* | Mean expression | Evaluable | 14 | 7 | 0.056 | 0.058 | 0.051 | 0.033 | 55. | 0.6888 | 0.5612 | 0.1224 | 0.1224 | 0.9124 | 0.9549 | No | No | No |
| **ISG/stress** | 1 | CD4 memory/activation-like | *DUSP1* | Mean expression | Evaluable | 14 | 7 | 0.127 | 0.160 | 0.089 | 0.083 | 43. | 0.6888 | 0.4388 | -0.1224 | -0.1224 | 0.9798 | 0.9549 | No | No | No |
| **Preserved/memory-like** | 1 | CD4 memory/activation-like | *LTB* | % positive | Evaluable | 14 | 7 | 38.954 | 47.144 | 38.539 | 44.978 | 43. | 0.6888 | 0.4388 | -0.1224 | -0.1224 | 0.8521 | 0.9549 | No | No | No |
| **Cytotoxic** | 0 | CD4 IL7R naive/memory-like | *CTSW* | Mean expression | Evaluable | 9 | 6 | 0.173 | 0.155 | 0.149 | 0.134 | 31. | 0.6889 | 0.5741 | 0.1481 | 0.1481 | 0.9124 | 0.9549 | No | No | No |
| **ISG/stress** | 0 | CD4 IL7R naive/memory-like | *ISG15* | Mean expression | Evaluable | 9 | 6 | 0.196 | 0.184 | 0.210 | 0.182 | 31. | 0.6889 | 0.5741 | 0.1481 | 0.1481 | 0.9798 | 0.9549 | No | No | No |
| **Preserved/memory-like** | 0 | CD4 IL7R naive/memory-like | *IL7R* | Mean expression | Evaluable | 9 | 6 | 2.416 | 2.383 | 2.290 | 2.381 | 23. | 0.6889 | 0.4259 | -0.1481 | -0.1481 | 0.8521 | 0.9549 | No | No | No |
| **Preserved/memory-like** | 0 | CD4 IL7R naive/memory-like | *SELL* | Mean expression | Evaluable | 9 | 6 | 1.068 | 1.032 | 1.070 | 0.978 | 31. | 0.6889 | 0.5741 | 0.1481 | 0.1481 | 0.8521 | 0.9549 | No | No | No |
| **ISG/stress** | 5 | preserved naive/memory-like | *IFIT2* | % positive | Evaluable | 13 | 6 | 1.797 | 1.666 | 2.041 | 1.257 | 44. | 0.6905 | 0.5641 | 0.1282 | 0.1282 | 0.9798 | 0.9549 | No | No | No |
| **ISG/stress** | 5 | preserved naive/memory-like | *JUNB* | % positive | Evaluable | 13 | 6 | 14.391 | 10.870 | 9.941 | 12.382 | 34. | 0.7012 | 0.4359 | -0.1282 | -0.1282 | 0.9798 | 0.9549 | No | No | No |
| **Preserved/memory-like** | 5 | preserved naive/memory-like | *LTB* | % positive | Evaluable | 13 | 6 | 27.075 | 30.992 | 24.242 | 26.792 | 34. | 0.7012 | 0.4359 | -0.1282 | -0.1282 | 0.8521 | 0.9549 | No | No | No |
| **Preserved/memory-like** | 5 | preserved naive/memory-like | *SKAP1* | % positive | Evaluable | 13 | 6 | 80.207 | 79.489 | 81.356 | 80.187 | 44. | 0.7012 | 0.5641 | 0.1282 | 0.1282 | 0.8521 | 0.9549 | No | No | No |
| **Checkpoint/activation** | 4 | CD8 naive/memory-like | *TNFRSF9* | Mean expression | Evaluable | 10 | 6 | 0.000 | 0.001 | 0.000 | 0.000 | 27.5 | 0.7061 | 0.4583 | -0.0833 | -0.0833 | 0.9627 | 0.9549 | No | No | No |
| **Checkpoint/activation** | 4 | CD8 naive/memory-like | *TNFRSF9* | % positive | Evaluable | 10 | 6 | 0.041 | 0.142 | 0.000 | 0.000 | 27.5 | 0.7061 | 0.4583 | -0.0833 | -0.0833 | 0.9627 | 0.9549 | No | No | No |
| **Cytotoxic** | 5 | preserved naive/memory-like | *GZMK* | Mean expression | Evaluable | 13 | 6 | 0.004 | 0.017 | 0.000 | 0.000 | 35. | 0.7098 | 0.4487 | -0.1026 | -0.1026 | 0.9124 | 0.9549 | No | No | No |
| **Cytotoxic** | 5 | preserved naive/memory-like | *GZMK* | % positive | Evaluable | 13 | 6 | 0.191 | 0.665 | 0.000 | 0.000 | 35. | 0.7098 | 0.4487 | -0.1026 | -0.1026 | 0.9124 | 0.9549 | No | No | No |
| **Checkpoint/activation** | 4 | CD8 naive/memory-like | *ICOS* | % positive | Evaluable | 10 | 6 | 22.273 | 20.799 | 12.932 | 18.911 | 34. | 0.7128 | 0.5667 | 0.1333 | 0.1333 | 0.9627 | 0.9549 | No | No | No |
| **Cytotoxic** | 4 | CD8 naive/memory-like | *NKG7* | Mean expression | Evaluable | 10 | 6 | 0.123 | 0.168 | 0.109 | 0.133 | 26. | 0.7128 | 0.4333 | -0.1333 | -0.1333 | 0.9124 | 0.9549 | No | No | No |
| **ISG/stress** | 4 | CD8 naive/memory-like | *DUSP1* | Mean expression | Evaluable | 10 | 6 | 0.453 | 0.282 | 0.132 | 0.196 | 34. | 0.7128 | 0.5667 | 0.1333 | 0.1333 | 0.9798 | 0.9549 | No | No | No |
| **ISG/stress** | 4 | CD8 naive/memory-like | *MX1* | % positive | Evaluable | 10 | 6 | 6.346 | 8.318 | 6.304 | 7.158 | 26. | 0.7128 | 0.4333 | -0.1333 | -0.1333 | 0.9798 | 0.9549 | No | No | No |
| **Checkpoint/activation** | 4 | CD8 naive/memory-like | *PDCD1* | % positive | Evaluable | 10 | 6 | 0.242 | 0.183 | 0.000 | 0.000 | 27. | 0.7217 | 0.4500 | -0.1000 | -0.1000 | 0.9627 | 0.9549 | No | No | No |
| **ISG/stress** | 5 | preserved naive/memory-like | *OAS1* | Mean expression | Evaluable | 13 | 6 | 0.037 | 0.035 | 0.026 | 0.032 | 34.5 | 0.7246 | 0.4423 | -0.1154 | -0.1154 | 0.9798 | 0.9549 | No | No | No |
| **ISG/stress** | 5 | preserved naive/memory-like | *HSPA1A* | Mean expression | Evaluable | 13 | 6 | 0.310 | 0.207 | 0.075 | 0.199 | 34.5 | 0.7256 | 0.4423 | -0.1154 | -0.1154 | 0.9798 | 0.9549 | No | No | No |
| **Cytotoxic** | 1 | CD4 memory/activation-like | *FGFBP2* | % positive | Evaluable | 14 | 7 | 0.349 | 0.102 | 0.106 | 0.063 | 54. | 0.7266 | 0.5510 | 0.1020 | 0.1020 | 0.9166 | 0.9549 | No | No | No |
| **ISG/stress** | 1 | CD4 memory/activation-like | *JUN* | Mean expression | Evaluable | 14 | 7 | 0.197 | 0.226 | 0.067 | 0.104 | 44. | 0.7370 | 0.4490 | -0.1020 | -0.1020 | 0.9798 | 0.9549 | No | No | No |

| | | | | | | | | | | | | | | | | | | | | | |
|---|---|---|---|---|---|---|---|---|---|---|---|---|---|---|---|---|---|---|---|---|---|
| **Checkpoint/activation** | 1 | CD4 memory/activation-like | *BTLA* | % positive | Evaluable | 14 | 7 | 4.313 | 4.392 | 3.764 | 4.654 | 44. | 0.7433 | 0.4490 | -0.1020 | -0.1020 | 0.9627 | 0.9549 | No | No | No |
| **Checkpoint/activation** | 1 | CD4 memory/activation-like | *CD69* | Mean expression | Evaluable | 14 | 7 | 0.249 | 0.262 | 0.183 | 0.192 | 44. | 0.7433 | 0.4490 | -0.1020 | -0.1020 | 0.9627 | 0.9549 | No | No | No |
| **Checkpoint/activation** | 1 | CD4 memory/activation-like | *TNFRSF4* | % positive | Evaluable | 14 | 7 | 8.146 | 8.794 | 8.504 | 9.677 | 44. | 0.7433 | 0.4490 | -0.1020 | -0.1020 | 0.9627 | 0.9549 | No | No | No |
| **Cytotoxic** | 1 | CD4 memory/activation-like | *PRF1* | % positive | Evaluable | 14 | 7 | 2.945 | 3.114 | 2.206 | 1.802 | 54. | 0.7433 | 0.5510 | 0.1020 | 0.1020 | 0.9166 | 0.9549 | No | No | No |
| **ISG/stress** | 1 | CD4 memory/activation-like | *JUNB* | % positive | Evaluable | 14 | 7 | 13.283 | 14.495 | 10.011 | 15.878 | 44. | 0.7433 | 0.4490 | -0.1020 | -0.1020 | 0.9798 | 0.9549 | No | No | No |
| **Checkpoint/activation** | 4 | CD8 naive/memory-like | *TIGIT* | Mean expression | Evaluable | 10 | 6 | 0.030 | 0.048 | 0.024 | 0.035 | 26.5 | 0.7447 | 0.4417 | -0.1167 | -0.1167 | 0.9627 | 0.9549 | No | No | No |
| **ISG/stress** | 4 | CD8 naive/memory-like | *OAS1* | % positive | Evaluable | 10 | 6 | 1.625 | 1.725 | 1.600 | 1.844 | 26.5 | 0.7447 | 0.4417 | -0.1167 | -0.1167 | 0.9798 | 0.9549 | No | No | No |
| **Cytotoxic** | 5 | preserved naive/memory-like | *PRF1* | % positive | Evaluable | 13 | 6 | 0.955 | 1.239 | 0.817 | 0.835 | 35. | 0.7529 | 0.4487 | -0.1026 | -0.1026 | 0.9166 | 0.9549 | No | No | No |
| **ISG/stress** | 5 | preserved naive/memory-like | *ISG15* | Mean expression | Evaluable | 13 | 6 | 0.069 | 0.047 | 0.053 | 0.057 | 43. | 0.7578 | 0.5513 | 0.1026 | 0.1026 | 0.9798 | 0.9549 | No | No | No |
| **ISG/stress** | 5 | preserved naive/memory-like | *MX1* | Mean expression | Evaluable | 13 | 6 | 0.100 | 0.081 | 0.097 | 0.084 | 43. | 0.7585 | 0.5513 | 0.1026 | 0.1026 | 0.9798 | 0.9549 | No | No | No |
| **ISG/stress** | 1 | CD4 memory/activation-like | *IFIT3* | % positive | Evaluable | 14 | 7 | 0.675 | 0.427 | 0.532 | 0.450 | 53.5 | 0.7628 | 0.5459 | 0.0918 | 0.0918 | 0.9798 | 0.9549 | No | No | No |
| **Cytotoxic** | 4 | CD8 naive/memory-like | *FGFBP2* | Mean expression | Evaluable | 10 | 6 | 0.007 | 0.007 | 0.002 | 0.000 | 33. | 0.7651 | 0.5500 | 0.1000 | 0.1000 | 0.9166 | 0.9549 | No | No | No |
| **Cytotoxic** | 4 | CD8 naive/memory-like | *FGFBP2* | % positive | Evaluable | 10 | 6 | 0.558 | 0.601 | 0.207 | 0.000 | 33. | 0.7651 | 0.5500 | 0.1000 | 0.1000 | 0.9166 | 0.9549 | No | No | No |
| **Checkpoint/activation** | 0 | CD4 IL7R naive/memory-like | *ICOS* | Mean expression | Evaluable | 9 | 6 | 0.333 | 0.308 | 0.293 | 0.271 | 30. | 0.7756 | 0.5556 | 0.1111 | 0.1111 | 0.9627 | 0.9549 | No | No | No |
| **Cytotoxic** | 0 | CD4 IL7R naive/memory-like | *CCL5* | Mean expression | Evaluable | 9 | 6 | 0.160 | 0.119 | 0.152 | 0.116 | 30. | 0.7756 | 0.5556 | 0.1111 | 0.1111 | 0.9166 | 0.9549 | No | No | No |
| **ISG/stress** | 0 | CD4 IL7R naive/memory-like | *HSPA1B* | Mean expression | Evaluable | 9 | 6 | 0.607 | 0.700 | 0.267 | 0.756 | 24. | 0.7756 | 0.4444 | -0.1111 | -0.1111 | 0.9798 | 0.9549 | No | No | No |
| **ISG/stress** | 0 | CD4 IL7R naive/memory-like | *IFIT1* | % positive | Evaluable | 9 | 6 | 1.587 | 1.386 | 1.146 | 1.531 | 30. | 0.7756 | 0.5556 | 0.1111 | 0.1111 | 0.9798 | 0.9549 | No | No | No |
| **ISG/stress** | 0 | CD4 IL7R naive/memory-like | *IFITM1* | % positive | Evaluable | 9 | 6 | 93.073 | 92.042 | 93.134 | 93.134 | 30. | 0.7756 | 0.5556 | 0.1111 | 0.1111 | 0.9798 | 0.9549 | No | No | No |
| **Checkpoint/activation** | 5 | preserved naive/memory-like | *CTLA4* | % positive | Evaluable | 13 | 6 | 2.841 | 2.248 | 1.563 | 2.299 | 35.5 | 0.7924 | 0.4551 | -0.0897 | -0.0897 | 0.9627 | 0.9549 | No | No | No |
| **ISG/stress** | 5 | preserved naive/memory-like | *HSPA1A* | % positive | Evaluable | 13 | 6 | 12.594 | 9.124 | 3.782 | 7.463 | 35.5 | 0.7924 | 0.4551 | -0.0897 | -0.0897 | 0.9798 | 0.9549 | No | No | No |
| **Checkpoint/activation** | 4 | CD8 naive/memory-like | *CD69* | Mean expression | Evaluable | 10 | 6 | 0.691 | 0.487 | 0.495 | 0.418 | 33. | 0.7925 | 0.5500 | 0.1000 | 0.1000 | 0.9627 | 0.9549 | No | No | No |
| **Checkpoint/activation** | 4 | CD8 naive/memory-like | *CD69* | % positive | Evaluable | 10 | 6 | 36.918 | 29.754 | 28.077 | 23.514 | 33. | 0.7925 | 0.5500 | 0.1000 | 0.1000 | 0.9627 | 0.9549 | No | No | No |
| **Checkpoint/activation** | 4 | CD8 naive/memory-like | *ICOS* | Mean expression | Evaluable | 10 | 6 | 0.312 | 0.301 | 0.215 | 0.293 | 33. | 0.7925 | 0.5500 | 0.1000 | 0.1000 | 0.9627 | 0.9549 | No | No | No |
| **Cytotoxic** | 4 | CD8 naive/memory-like | *CTSW* | Mean expression | Evaluable | 10 | 6 | 0.590 | 0.602 | 0.648 | 0.545 | 33. | 0.7925 | 0.5500 | 0.1000 | 0.1000 | 0.9166 | 0.9549 | No | No | No |
| **Cytotoxic** | 4 | CD8 naive/memory-like | *CTSW* | % positive | Evaluable | 10 | 6 | 39.346 | 38.177 | 45.529 | 33.444 | 33. | 0.7925 | 0.5500 | 0.1000 | 0.1000 | 0.9166 | 0.9549 | No | No | No |
| **ISG/stress** | 4 | CD8 naive/memory-like | *IFIT2* | % positive | Evaluable | 10 | 6 | 3.003 | 3.519 | 2.346 | 3.387 | 27. | 0.7925 | 0.4500 | -0.1000 | -0.1000 | 0.9798 | 0.9549 | No | No | No |
| **ISG/stress** | 4 | CD8 naive/memory-like | *IFITM2* | % positive | Evaluable | 10 | 6 | 67.157 | 68.252 | 67.029 | 70.408 | 27. | 0.7925 | 0.4500 | -0.1000 | -0.1000 | 0.9798 | 0.9549 | No | No | No |
| **ISG/stress** | 4 | CD8 naive/memory-like | *ISG15* | % positive | Evaluable | 10 | 6 | 17.153 | 14.963 | 13.787 | 13.036 | 33. | 0.7925 | 0.5500 | 0.1000 | 0.1000 | 0.9798 | 0.9549 | No | No | No |
| **Preserved/memory-like** | 4 | CD8 naive/memory-like | *CD27* | % positive | Evaluable | 10 | 6 | 40.705 | 39.561 | 40.651 | 37.250 | 33. | 0.7925 | 0.5500 | 0.1000 | 0.1000 | 0.9241 | 0.9549 | No | No | No |
| **Preserved/memory-like** | 4 | CD8 naive/memory-like | *SELL* | Mean expression | Evaluable | 10 | 6 | 1.240 | 1.208 | 1.230 | 1.213 | 33. | 0.7925 | 0.5500 | 0.1000 | 0.1000 | 0.9241 | 0.9549 | No | No | No |
| **ISG/stress** | 1 | CD4 memory/activation-like | *ISG15* | Mean expression | Evaluable | 14 | 7 | 0.126 | 0.124 | 0.139 | 0.152 | 53. | 0.7990 | 0.5408 | 0.0816 | 0.0816 | 0.9798 | 0.9549 | No | No | No |
| **ISG/stress** | 1 | CD4 memory/activation-like | *JUNB* | Mean expression | Evaluable | 14 | 7 | 0.295 | 0.310 | 0.206 | 0.311 | 45. | 0.7990 | 0.4592 | -0.0816 | -0.0816 | 0.9798 | 0.9549 | No | No | No |
| **Preserved/memory-like** | 1 | CD4 memory/activation-like | *CCR7* | Mean expression | Evaluable | 14 | 7 | 0.268 | 0.250 | 0.253 | 0.240 | 53. | 0.7990 | 0.5408 | 0.0816 | 0.0816 | 0.9241 | 0.9549 | No | No | No |
| **Preserved/memory-like** | 1 | CD4 memory/activation-like | *SELL* | Mean expression | Evaluable | 14 | 7 | 0.481 | 0.439 | 0.495 | 0.429 | 53. | 0.7990 | 0.5408 | 0.0816 | 0.0816 | 0.9241 | 0.9549 | No | No | No |
| **ISG/stress** | 1 | CD4 memory/activation-like | *IFIT3* | Mean expression | Evaluable | 14 | 7 | 0.014 | 0.008 | 0.007 | 0.008 | 52.5 | 0.8209 | 0.5357 | 0.0714 | 0.0714 | 0.9798 | 0.9661 | No | No | No |
| **Checkpoint/activation** | 4 | CD8 naive/memory-like | *TIGIT* | % positive | Evaluable | 10 | 6 | 2.310 | 3.222 | 1.467 | 2.855 | 27.5 | 0.8281 | 0.4583 | -0.0833 | -0.0833 | 0.9627 | 0.9661 | No | No | No |
| **Checkpoint/activation** | 5 | preserved naive/memory-like | *CD69* | % positive | Evaluable | 13 | 6 | 17.844 | 13.331 | 9.532 | 12.444 | 36. | 0.8314 | 0.4615 | -0.0769 | -0.0769 | 0.9627 | 0.9661 | No | No | No |
| **Checkpoint/activation** | 5 | preserved naive/memory-like | *TOX* | Mean expression | Evaluable | 13 | 6 | 0.167 | 0.167 | 0.122 | 0.143 | 36. | 0.8314 | 0.4615 | -0.0769 | -0.0769 | 0.9627 | 0.9661 | No | No | No |
| **Checkpoint/activation** | 5 | preserved naive/memory-like | *TOX* | % positive | Evaluable | 13 | 6 | 7.396 | 7.338 | 6.250 | 6.202 | 36. | 0.8314 | 0.4615 | -0.0769 | -0.0769 | 0.9627 | 0.9661 | No | No | No |
| **Preserved/memory-like** | 5 | preserved naive/memory-like | *TCF7* | Mean expression | Evaluable | 13 | 6 | 1.276 | 1.197 | 1.321 | 1.297 | 42. | 0.8314 | 0.5385 | 0.0769 | 0.0769 | 0.9437 | 0.9661 | No | No | No |
| **Cytotoxic** | 1 | CD4 memory/activation-like | *FGFBP2* | Mean expression | Evaluable | 14 | 7 | 0.008 | 0.002 | 0.002 | 0.001 | 52. | 0.8460 | 0.5306 | 0.0612 | 0.0612 | 0.9437 | 0.9661 | No | No | No |
| **Cytotoxic** | 5 | preserved naive/memory-like | *GZMH* | Mean expression | Evaluable | 13 | 6 | 0.004 | 0.004 | 0.000 | 0.000 | 41. | 0.8537 | 0.5256 | 0.0513 | 0.0513 | 0.9437 | 0.9661 | No | No | No |
| **Checkpoint/activation** | 1 | CD4 memory/activation-like | *CD69* | % positive | Evaluable | 14 | 7 | 11.956 | 13.022 | 9.390 | 7.937 | 46. | 0.8557 | 0.4694 | -0.0612 | -0.0612 | 0.9780 | 0.9661 | No | No | No |
| **Cytotoxic** | 1 | CD4 memory/activation-like | *GZMK* | Mean expression | Evaluable | 14 | 7 | 0.076 | 0.086 | 0.062 | 0.070 | 46. | 0.8557 | 0.4694 | -0.0612 | -0.0612 | 0.9437 | 0.9661 | No | No | No |
| **ISG/stress** | 1 | CD4 memory/activation-like | *DUSP1* | % positive | Evaluable | 14 | 7 | 6.734 | 8.053 | 4.530 | 4.505 | 46. | 0.8557 | 0.4694 | -0.0612 | -0.0612 | 0.9798 | 0.9661 | No | No | No |
| **Preserved/memory-like** | 1 | CD4 memory/activation-like | *TCF7* | Mean expression | Evaluable | 14 | 7 | 0.708 | 0.730 | 0.701 | 0.769 | 46. | 0.8557 | 0.4694 | -0.0612 | -0.0612 | 0.9437 | 0.9661 | No | No | No |
| **Cytotoxic** | 0 | CD4 IL7R naive/memory-like | *GZMH* | Mean expression | Evaluable | 9 | 6 | 0.005 | 0.003 | 0.007 | 0.002 | 29. | 0.8571 | 0.5370 | 0.0741 | 0.0741 | 0.9437 | 0.9661 | No | No | No |
| **ISG/stress** | 5 | preserved naive/memory-like | *MX1* | % positive | Evaluable | 13 | 6 | 4.880 | 4.505 | 4.651 | 4.382 | 41.5 | 0.8605 | 0.5321 | 0.0641 | 0.0641 | 0.9798 | 0.9661 | No | No | No |
| **ISG/stress** | 0 | CD4 IL7R naive/memory-like | *HSPA1A* | % positive | Evaluable | 9 | 6 | 21.237 | 25.284 | 13.023 | 30.024 | 25. | 0.8639 | 0.4630 | -0.0741 | -0.0741 | 0.9798 | 0.9661 | No | No | No |
| **ISG/stress** | 0 | CD4 IL7R naive/memory-like | *HSPA1B* | % positive | Evaluable | 9 | 6 | 29.660 | 31.508 | 19.063 | 31.869 | 25. | 0.8639 | 0.4630 | -0.0741 | -0.0741 | 0.9798 | 0.9661 | No | No | No |
| **ISG/stress** | 0 | CD4 IL7R naive/memory-like | *JUN* | Mean expression | Evaluable | 9 | 6 | 0.402 | 0.341 | 0.238 | 0.307 | 29. | 0.8639 | 0.5370 | 0.0741 | 0.0741 | 0.9798 | 0.9661 | No | No | No |
| **ISG/stress** | 0 | CD4 IL7R naive/memory-like | *JUN* | % positive | Evaluable | 9 | 6 | 21.360 | 17.802 | 13.433 | 16.048 | 29. | 0.8639 | 0.5370 | 0.0741 | 0.0741 | 0.9798 | 0.9661 | No | No | No |
| **Preserved/memory-like** | 0 | CD4 IL7R naive/memory-like | *CD27* | Mean expression | Evaluable | 9 | 6 | 0.534 | 0.517 | 0.567 | 0.563 | 29. | 0.8639 | 0.5370 | 0.0741 | 0.0741 | 0.9437 | 0.9661 | No | No | No |
| **Preserved/memory-like** | 0 | CD4 IL7R naive/memory-like | *CD28* | Mean expression | Evaluable | 9 | 6 | 0.392 | 0.371 | 0.394 | 0.363 | 29. | 0.8639 | 0.5370 | 0.0741 | 0.0741 | 0.9437 | 0.9661 | No | No | No |
| **Checkpoint/activation** | 4 | CD8 naive/memory-like | *BTLA* | % positive | Evaluable | 10 | 6 | 8.019 | 7.453 | 8.270 | 6.453 | 32. | 0.8749 | 0.5333 | 0.0667 | 0.0667 | 0.9870 | 0.9661 | No | No | No |
| **Cytotoxic** | 4 | CD8 naive/memory-like | *KLRG1* | % positive | Evaluable | 10 | 6 | 8.403 | 9.581 | 6.980 | 5.699 | 32. | 0.8749 | 0.5333 | 0.0667 | 0.0667 | 0.9437 | 0.9661 | No | No | No |
| **Cytotoxic** | 4 | CD8 naive/memory-like | *NKG7* | % positive | Evaluable | 10 | 6 | 8.413 | 9.616 | 6.881 | 7.373 | 28. | 0.8749 | 0.4667 | -0.0667 | -0.0667 | 0.9437 | 0.9661 | No | No | No |
| **ISG/stress** | 4 | CD8 naive/memory-like | *DUSP1* | % positive | Evaluable | 10 | 6 | 25.575 | 17.693 | 8.364 | 12.106 | 32. | 0.8749 | 0.5333 | 0.0667 | 0.0667 | 0.9798 | 0.9661 | No | No | No |
| **ISG/stress** | 4 | CD8 naive/memory-like | *FOS* | % positive | Evaluable | 10 | 6 | 36.798 | 27.117 | 21.098 | 17.753 | 32. | 0.8749 | 0.5333 | 0.0667 | 0.0667 | 0.9798 | 0.9661 | No | No | No |
| **Preserved/memory-like** | 4 | CD8 naive/memory-like | *CD27* | Mean expression | Evaluable | 10 | 6 | 0.601 | 0.620 | 0.643 | 0.655 | 28. | 0.8749 | 0.4667 | -0.0667 | -0.0667 | 0.9437 | 0.9661 | No | No | No |
| **Preserved/memory-like** | 4 | CD8 naive/memory-like | *IL7R* | Mean expression | Evaluable | 10 | 6 | 2.418 | 2.330 | 2.296 | 2.365 | 32. | 0.8749 | 0.5333 | 0.0667 | 0.0667 | 0.9437 | 0.9661 | No | No | No |
| **Preserved/memory-like** | 5 | preserved naive/memory-like | *IL7R* | Mean expression | Evaluable | 13 | 6 | 2.246 | 2.328 | 2.333 | 2.409 | 37. | 0.8983 | 0.4744 | -0.0513 | -0.0513 | 0.9476 | 0.9838 | No | No | No |
| **Preserved/memory-like** | 5 | preserved naive/memory-like | *TCF7* | % positive | Evaluable | 13 | 6 | 55.765 | 52.205 | 53.125 | 57.299 | 41. | 0.8983 | 0.5256 | 0.0513 | 0.0513 | 0.9476 | 0.9838 | No | No | No |
| **Checkpoint/activation** | 0 | CD4 IL7R naive/memory-like | *LAG3* | Mean expression | Evaluable | 9 | 6 | 0.011 | 0.008 | 0.009 | 0.009 | 28.5 | 0.9053 | 0.5278 | 0.0556 | 0.0556 | 1.0000 | 0.9838 | No | No | No |
| **ISG/stress** | 1 | CD4 memory/activation-like | *FOS* | Mean expression | Evaluable | 14 | 7 | 0.297 | 0.314 | 0.197 | 0.182 | 47. | 0.9106 | 0.4796 | -0.0408 | -0.0408 | 0.9798 | 0.9838 | No | No | No |
| **ISG/stress** | 1 | CD4 memory/activation-like | *FOS* | % positive | Evaluable | 14 | 7 | 12.887 | 13.586 | 9.552 | 9.797 | 47. | 0.9106 | 0.4796 | -0.0408 | -0.0408 | 0.9798 | 0.9838 | No | No | No |
| **ISG/stress** | 1 | CD4 memory/activation-like | *ISG15* | % positive | Evaluable | 14 | 7 | 6.690 | 6.216 | 7.831 | 7.496 | 51. | 0.9131 | 0.5204 | 0.0408 | 0.0408 | 0.9798 | 0.9838 | No | No | No |
| **Preserved/memory-like** | 1 | CD4 memory/activation-like | *TCF7* | % positive | Evaluable | 14 | 7 | 32.212 | 34.157 | 34.246 | 35.484 | 47. | 0.9131 | 0.4796 | -0.0408 | -0.0408 | 0.9528 | 0.9838 | No | No | No |
| **Cytotoxic** | 4 | CD8 naive/memory-like | *GNLY* | Mean expression | Evaluable | 10 | 6 | 0.043 | 0.043 | 0.033 | 0.034 | 28.5 | 0.9136 | 0.4750 | -0.0500 | -0.0500 | 0.9638 | 0.9838 | No | No | No |
| **Cytotoxic** | 4 | CD8 naive/memory-like | *GNLY* | % positive | Evaluable | 10 | 6 | 2.595 | 2.250 | 1.822 | 2.198 | 31.5 | 0.9136 | 0.5250 | 0.0500 | 0.0500 | 0.9638 | 0.9838 | No | No | No |
| **ISG/stress** | 5 | preserved naive/memory-like | *IFIT3* | % positive | Evaluable | 13 | 6 | 0.550 | 0.506 | 0.000 | 0.000 | 40.5 | 0.9192 | 0.5192 | 0.0385 | 0.0385 | 0.9798 | 0.9861 | No | No | No |
| **ISG/stress** | 5 | preserved naive/memory-like | *OAS1* | % positive | Evaluable | 13 | 6 | 1.909 | 1.615 | 1.408 | 1.812 | 37.5 | 0.9298 | 0.4808 | -0.0385 | -0.0385 | 0.9798 | 0.9861 | No | No | No |
| **Cytotoxic** | 5 | preserved naive/memory-like | *FGFBP2* | Mean expression | Evaluable | 13 | 6 | 0.002 | 0.003 | 0.000 | 0.000 | 40. | 0.9510 | 0.5128 | 0.0256 | 0.0256 | 0.9710 | 0.9861 | No | No | No |
| **Cytotoxic** | 5 | preserved naive/memory-like | *FGFBP2* | % positive | Evaluable | 13 | 6 | 0.083 | 0.194 | 0.000 | 0.000 | 40. | 0.9510 | 0.5128 | 0.0256 | 0.0256 | 0.9710 | 0.9861 | No | No | No |

| | | | | | | | | | | | | | | | | | | | | | |
|---|---|---|---|---|---|---|---|---|---|---|---|---|---|---|---|---|---|---|---|---|---|
| **Cytotoxic** | 5 | preserved naive/memory-like | *GZMH* | % positive | Evaluable | 13 | 6 | 0.170 | 0.236 | 0.000 | 0.000 | 40. | 0.9510 | 0.5128 | 0.0256 | 0.0256 | 0.9710 | 0.9861 | No | No | No |
| **ISG/stress** | 0 | CD4 IL7R naive/memory-like | *IFIT1* | Mean expression | Evaluable | 9 | 6 | 0.023 | 0.021 | 0.020 | 0.021 | 26. | 0.9546 | 0.4815 | -0.0370 | -0.0370 | 0.9798 | 0.9861 | No | No | No |
| **ISG/stress** | 0 | CD4 IL7R naive/memory-like | *IFIT2* | Mean expression | Evaluable | 9 | 6 | 0.049 | 0.042 | 0.041 | 0.043 | 28. | 0.9546 | 0.5185 | 0.0370 | 0.0370 | 0.9798 | 0.9861 | No | No | No |
| **Checkpoint/activation** | 4 | CD8 naive/memory-like | *TNFRSF4* | % positive | Evaluable | 10 | 6 | 0.898 | 0.873 | 0.716 | 0.155 | 31. | 0.9549 | 0.5167 | 0.0333 | 0.0333 | 1.0000 | 0.9861 | No | No | No |
| **Checkpoint/activation** | 4 | CD8 naive/memory-like | *HAVCR2* | Mean expression | Evaluable | 10 | 6 | 0.028 | 0.035 | 0.030 | 0.028 | 29. | 0.9566 | 0.4833 | -0.0333 | -0.0333 | 1.0000 | 0.9861 | No | No | No |
| **Checkpoint/activation** | 4 | CD8 naive/memory-like | *LAG3* | Mean expression | Evaluable | 10 | 6 | 0.036 | 0.040 | 0.025 | 0.024 | 31. | 0.9566 | 0.5167 | 0.0333 | 0.0333 | 1.0000 | 0.9861 | No | No | No |
| **Checkpoint/activation** | 4 | CD8 naive/memory-like | *LAG3* | % positive | Evaluable | 10 | 6 | 2.447 | 2.375 | 1.658 | 1.625 | 31. | 0.9566 | 0.5167 | 0.0333 | 0.0333 | 1.0000 | 0.9861 | No | No | No |
| **ISG/stress** | 4 | CD8 naive/memory-like | *ISG15* | Mean expression | Evaluable | 10 | 6 | 0.243 | 0.214 | 0.201 | 0.226 | 29. | 0.9578 | 0.4833 | -0.0333 | -0.0333 | 0.9798 | 0.9861 | No | No | No |
| **Preserved/memory-like** | 4 | CD8 naive/memory-like | *CCR7* | Mean expression | Evaluable | 10 | 6 | 1.083 | 1.014 | 1.082 | 1.058 | 31. | 0.9578 | 0.5167 | 0.0333 | 0.0333 | 0.9710 | 0.9861 | No | No | No |
| **ISG/stress** | 5 | preserved naive/memory-like | *IFIT1* | % positive | Evaluable | 13 | 6 | 1.305 | 1.171 | 0.877 | 0.835 | 38. | 0.9645 | 0.4872 | -0.0256 | -0.0256 | 0.9798 | 0.9861 | No | No | No |
| **ISG/stress** | 5 | preserved naive/memory-like | *JUN* | Mean expression | Evaluable | 13 | 6 | 0.272 | 0.127 | 0.059 | 0.130 | 40. | 0.9645 | 0.5128 | 0.0256 | 0.0256 | 0.9798 | 0.9861 | No | No | No |
| **ISG/stress** | 5 | preserved naive/memory-like | *JUN* | % positive | Evaluable | 13 | 6 | 11.584 | 5.384 | 3.858 | 6.191 | 40. | 0.9645 | 0.5128 | 0.0256 | 0.0256 | 0.9798 | 0.9861 | No | No | No |
| **Preserved/memory-like** | 5 | preserved naive/memory-like | *IL7R* | % positive | Evaluable | 13 | 6 | 79.390 | 80.469 | 76.056 | 82.430 | 40. | 0.9650 | 0.5128 | 0.0256 | 0.0256 | 0.9710 | 0.9861 | No | No | No |
| **Cytotoxic** | 1 | CD4 memory/activation-like | *CCL5* | % positive | Evaluable | 14 | 7 | 14.873 | 14.577 | 15.670 | 11.712 | 50. | 0.9710 | 0.5102 | 0.0204 | 0.0204 | 0.9710 | 0.9861 | No | No | No |
| **Cytotoxic** | 1 | CD4 memory/activation-like | *NKG7* | Mean expression | Evaluable | 14 | 7 | 0.106 | 0.106 | 0.111 | 0.089 | 50. | 0.9710 | 0.5102 | 0.0204 | 0.0204 | 0.9710 | 0.9861 | No | No | No |
| **ISG/stress** | 1 | CD4 memory/activation-like | *IFIT2* | % positive | Evaluable | 14 | 7 | 1.461 | 1.385 | 1.062 | 1.014 | 50. | 0.9710 | 0.5102 | 0.0204 | 0.0204 | 0.9798 | 0.9861 | No | No | No |
| **Preserved/memory-like** | 1 | CD4 memory/activation-like | *CCR7* | % positive | Evaluable | 14 | 7 | 13.256 | 12.792 | 10.648 | 12.213 | 50. | 0.9710 | 0.5102 | 0.0204 | 0.0204 | 0.9710 | 0.9861 | No | No | No |
| **Preserved/memory-like** | 1 | CD4 memory/activation-like | *SELL* | % positive | Evaluable | 14 | 7 | 22.045 | 21.118 | 21.056 | 19.355 | 50. | 0.9710 | 0.5102 | 0.0204 | 0.0204 | 0.9710 | 0.9861 | No | No | No |
| **Checkpoint/activation** | 4 | CD8 naive/memory-like | *BTLA* | Mean expression | Evaluable | 10 | 6 | 0.103 | 0.099 | 0.102 | 0.104 | 30. | 1.0000 | 0.5000 | 0.0000 | 0.0000 | 1.0000 | 1.0000 | No | No | No |
| **Checkpoint/activation** | 4 | CD8 naive/memory-like | *HAVCR2* | % positive | Evaluable | 10 | 6 | 2.363 | 2.675 | 2.668 | 2.087 | 30. | 1.0000 | 0.5000 | 0.0000 | 0.0000 | 1.0000 | 1.0000 | No | No | No |
| **Checkpoint/activation** | 4 | CD8 naive/memory-like | *TNFRSF4* | Mean expression | Evaluable | 10 | 6 | 0.012 | 0.013 | 0.006 | 0.002 | 30. | 1.0000 | 0.5000 | 0.0000 | 0.0000 | 1.0000 | 1.0000 | No | No | No |
| **Checkpoint/activation** | 5 | preserved naive/memory-like | *TNFRSF9* | Mean expression | Evaluable | 13 | 6 | 0.000 | 0.000 | 0.000 | 0.000 | 39. | 1.0000 | 0.5000 | 0.0000 | 0.0000 | 1.0000 | 1.0000 | No | No | No |
| **Checkpoint/activation** | 5 | preserved naive/memory-like | *TNFRSF9* | % positive | Evaluable | 13 | 6 | 0.000 | 0.000 | 0.000 | 0.000 | 39. | 1.0000 | 0.5000 | 0.0000 | 0.0000 | 1.0000 | 1.0000 | No | No | No |
| **ISG/stress** | 5 | preserved naive/memory-like | *IFIT1* | Mean expression | Evaluable | 13 | 6 | 0.026 | 0.023 | 0.017 | 0.019 | 39. | 1.0000 | 0.5000 | 0.0000 | 0.0000 | 1.0000 | 1.0000 | No | No | No |

Mean expression denotes the patient-level mean of log-normalized expression, including zero values. “% positive” denotes the percentage of cells with at least one raw UMI for the corresponding gene. Signed AUC was calculated as 2 × AUC- 1, with positive values indicating higher values in pCR and negative values indicating higher values in non-pCR. AUC, area under the receiver operating characteristic curve; FDR, false discovery rate; ISG, interferon-stimulated gene; n, number of patients; pCR, pathological complete response; non-pCR, absence of pCR; q, Benjamini–Hochberg-adjusted P value.

**Supplementary Table 5. Baseline B-cell subcluster abundance according to pathological response.**

| B-cell subcluster | n pCR | n non-pCR | Mean abundance, pCR (%) | Mean abundance, non-pCR (%) | Median abundance, pCR (%) | Median abundance, non-pCR (%) | Median difference (pCR − non-pCR), pp | Mann–Whitney U | Two-sided P | Benjamini–Hochberg q | AUC (pCR higher) | Cliff's δ (pCR − non-pCR) | Nominal P <0.05 | FDR q <0.10 | Direction | Analysis status |
|---|---|---|---|---|---|---|---|---|---|---|---|---|---|---|---|---|
| 0 | 13 | 8 | 7.53 | 12.79 | 0.72 | 0.25 | 0.46 | — | — | — | — | — | — | — | — | Descriptive only |
| 1 | 13 | 8 | 29.60 | 31.65 | 28.81 | 32.29 | -3.49 | 48.0 | 0.8044 | 0.8044 | 0.462 | -0.077 | No | No | Higher in non-pCR | Evaluable |
| 2 | 13 | 8 | 27.00 | 30.65 | 20.20 | 34.28 | -14.08 | 44.0 | 0.5951 | 0.6942 | 0.423 | -0.154 | No | No | Higher in non-pCR | Evaluable |
| 3 | 13 | 8 | 12.05 | 7.01 | 10.06 | 8.83 | 1.23 | — | — | — | — | — | — | — | — | Descriptive only |
| 4 | 13 | 8 | 15.60 | 10.54 | 0.00 | 0.12 | -0.12 | 36.0 | 0.1990 | 0.3482 | 0.346 | -0.308 | No | No | Higher in non-pCR | Evaluable |
| 5 | 13 | 8 | 2.15 | 1.86 | 1.42 | 1.11 | 0.31 | 62.0 | 0.5002 | 0.6942 | 0.596 | 0.192 | No | No | Higher in pCR | Evaluable |
| 6 | 13 | 8 | 3.07 | 1.23 | 2.10 | 1.11 | 0.98 | 75.0 | 0.1042 | 0.3482 | 0.721 | 0.442 | No | No | Higher in pCR | Evaluable |
| 7 | 13 | 8 | 0.76 | 1.77 | 0.36 | 1.42 | -1.06 | 33.0 | 0.1682 | 0.3482 | 0.317 | -0.365 | No | No | Higher in non-pCR | Evaluable |
| 8 | 13 | 8 | 0.00 | 2.22 | 0.00 | 0.00 | 0.00 | — | — | — | — | — | — | — | — | Descriptive only |
| 9 | 13 | 8 | 1.05 | 0.29 | 0.72 | 0.15 | 0.56 | 85.0 | **0.0178** | **0.1248** | **0.817** | **0.635** | **Yes** | **No** | Higher in pCR | Evaluable |
| 10 | 13 | 8 | 1.19 | 0.00 | 0.00 | 0.00 | 0.00 | — | — | — | — | — | — | — | — | Descriptive only |

AUC, area under the receiver operating characteristic curve; FDR, false discovery rate; n, number of patients; pCR, pathological complete response; non-pCR, absence of pCR; pp, percentage points; q, Benjamini–Hochberg-adjusted P value. Dashes indicate statistics not reported for descriptive-only subclusters.

**Supplementary Table 6. Baseline B-cell functional program scores according to pathological response.**

| B-cell program | Biological dimension | B-cell subcluster | Evaluation status | Exclusion reason | Program genes requested | Program genes used | Gene coverage | Genes used | n pCR | n non-pCR | Mean score, pCR | Mean score, non-pCR | Median score, pCR | Median score, non-pCR | Median difference (pCR − non-pCR) | Mann–Whitney U | Two-sided P | AUC (pCR higher) | Signed AUC | Cliff's δ (pCR − non-pCR) | BH q within program | Global BH q | Nominal P <0.05 | Within-program FDR q <0.10 | Global FDR q <0.10 | Direction |
|---|---|---|---|---|---|---|---|---|---|---|---|---|---|---|---|---|---|---|---|---|---|---|---|---|---|---|
| **Naive-like** | Naive and unswitched B-cell differentiation state. | 0 | *Not evaluable* | *one patient contributes 87.7% of cells; pCR n=1; requires >= 5; non-pCR n=1; requires >= 5* | 3 | 3 | 100% | *TCL1A, IGHD, IGHM* | 1 | 1 | 0.187 | 0.188 | 0.187 | 0.188 | 0.000 | | | | | | | | No | No | No | |
| **Naive-like** | Naive and unswitched B-cell differentiation state. | 1 | Evaluable | | 3 | 3 | 100% | *TCL1A, IGHD, IGHM* | 8 | 6 | 0.453 | 0.328 | 0.426 | 0.320 | 0.106 | 41.0 | **0.0293** | 0.8542 | 0.7083 | 0.7083 | 0.0586 | 0.2930 | Yes | Yes | No | Higher in pCR |
| **Naive-like** | Naive and unswitched B-cell differentiation state. | 2 | Evaluable | | 3 | 3 | 100% | *TCL1A, IGHD, IGHM* | 11 | 6 | 1.364 | 1.392 | 1.442 | 1.382 | 0.060 | 31.0 | 0.8836 | 0.470 | -0.061 | -0.061 | 0.8836 | 1.0000 | No | No | No | Higher in non-pCR |
| **Naive-like** | Naive and unswitched B-cell differentiation state. | 3 | *Not evaluable* | *mixed B/T-lineage cluster: CD3D, CD3E, IL7R and GIMAP7 signal* | 3 | 3 | 100% | *TCL1A, IGHD, IGHM* | 9 | 5 | 0.602 | 0.649 | 0.685 | 0.603 | 0.082 | | | | | | | | No | No | No | |
| **Naive-like** | Naive and unswitched B-cell differentiation state. | 4 | *Not evaluable* | *pCR n=3; requires >= 5; non-pCR n=1; requires >= 5* | 3 | 3 | 100% | *TCL1A, IGHD, IGHM* | 3 | 1 | 0.945 | 0.822 | 1.009 | 0.822 | 0.187 | | | | | | | | No | No | No | |
| **Naive-like** | Naive and unswitched B-cell differentiation state. | 5 | *Not evaluable* | *pCR n=2; requires >= 5; non-pCR n=2; requires >= 5* | 3 | 3 | 100% | *TCL1A, IGHD, IGHM* | 2 | 2 | 0.156 | 0.660 | 0.156 | 0.660 | -0.503 | | | | | | | | No | No | No | |
| **Naive-like** | Naive and unswitched B-cell differentiation state. | 6 | *Not evaluable* | *pCR n=3; requires >= 5; non-pCR n=1; requires >= 5* | 3 | 3 | 100% | *TCL1A, IGHD, IGHM* | 3 | 1 | 0.039 | 0.055 | 0.031 | 0.055 | -0.023 | | | | | | | | No | No | No | |
| **Naive-like** | Naive and unswitched B-cell differentiation state. | 7 | *Not evaluable* | *pCR n=1; requires >= 5; non-pCR n=3; requires >= 5* | 3 | 3 | 100% | *TCL1A, IGHD, IGHM* | 1 | 3 | 1.310 | 1.343 | 1.310 | 1.277 | 0.032 | | | | | | | | No | No | No | |
| **Naive-like** | Naive and unswitched B-cell differentiation state. | 8 | *Not evaluable* | *one patient contributes 100.0% of cells; pCR n=0; requires >= 5; non-pCR n=1; requires >= 5* | 3 | 3 | 100% | *TCL1A, IGHD, IGHM* | 0 | 1 | | 0.224 | | 0.224 | | | | | | | | | No | No | No | |
| **Naive-like** | Naive and unswitched B-cell differentiation state. | 9 | *Not evaluable* | *pCR n=1; requires >= 5; non-pCR n=0; requires >= 5* | 3 | 3 | 100% | *TCL1A, IGHD, IGHM* | 1 | 0 | 0.025 | | 0.025 | | | | | | | | | | No | No | No | |
| **Naive-like** | Naive and unswitched B-cell differentiation state. | 10 | *Not evaluable* | *one patient contributes 83.3% of cells; only 2 patients contribute cells; pCR n=1; requires >= 5; non-pCR n=0; requires >= 5* | 3 | 3 | 100% | *TCL1A, IGHD, IGHM* | 1 | 0 | 0.000 | | 0.000 | | | | | | | | | | No | No | No | |
| **Memory-like** | Antigen-experienced and memory-related B-cell state. | 0 | *Not evaluable* | *one patient contributes 87.7% of cells; pCR n=1; requires >= 5; non-pCR n=1; requires >= 5* | 3 | 3 | 100% | *CD27, TNFRSF13B, AIM2* | 1 | 1 | 0.136 | 0.032 | 0.136 | 0.032 | 0.103 | | | | | | | | No | No | No | |
| **Memory-like** | Antigen-experienced and memory-related B-cell state. | 1 | Evaluable | | 3 | 3 | 100% | *CD27, TNFRSF13B, AIM2* | 8 | 6 | 0.482 | 0.427 | 0.514 | 0.414 | 0.100 | 29.0 | 0.5728 | 0.604 | 0.208 | 0.208 | 1.0000 | 1.0000 | No | No | No | Higher in pCR |
| **Memory-like** | Antigen-experienced and memory-related B-cell state. | 2 | Evaluable | | 3 | 3 | 100% | *CD27, TNFRSF13B, AIM2* | 11 | 6 | 0.041 | 0.034 | 0.035 | 0.033 | 0.002 | 33.0 | 1.0000 | 0.500 | 0.000 | 0.000 | 1.0000 | 1.0000 | No | No | No | |
| **Memory-like** | Antigen-experienced and memory-related B-cell state. | 3 | *Not evaluable* | *mixed B/T-lineage cluster: CD3D, CD3E, IL7R and GIMAP7 signal* | 3 | 3 | 100% | *CD27, TNFRSF13B, AIM2* | 9 | 5 | 0.266 | 0.258 | 0.274 | 0.227 | 0.047 | | | | | | | | No | No | No | |
| **Memory-like** | Antigen-experienced and memory-related B-cell state. | 4 | *Not evaluable* | *pCR n=3; requires >= 5; non-pCR n=1; requires >= 5* | 3 | 3 | 100% | *CD27, TNFRSF13B, AIM2* | 3 | 1 | 0.236 | 0.441 | 0.239 | 0.441 | -0.202 | | | | | | | | No | No | No | |
| **Memory-like** | Antigen-experienced and memory-related B-cell state. | 5 | *Not evaluable* | *pCR n=2; requires >= 5; non-pCR n=2; requires >= 5* | 3 | 3 | 100% | *CD27, TNFRSF13B, AIM2* | 2 | 2 | 0.617 | 0.454 | 0.617 | 0.454 | 0.163 | | | | | | | | No | No | No | |
| **Memory-like** | Antigen-experienced and memory-related B-cell state. | 6 | *Not evaluable* | *pCR n=3; requires >= 5; non-pCR n=1; requires >= 5* | 3 | 3 | 100% | *CD27, TNFRSF13B, AIM2* | 3 | 1 | 0.151 | 0.241 | 0.131 | 0.241 | -0.110 | | | | | | | | No | No | No | |
| **Memory-like** | Antigen-experienced and memory-related B-cell state. | 7 | *Not evaluable* | *pCR n=1; requires >= 5; non-pCR n=3; requires >= 5* | 3 | 3 | 100% | *CD27, TNFRSF13B, AIM2* | 1 | 3 | 0.128 | 0.082 | 0.128 | 0.097 | 0.031 | | | | | | | | No | No | No | |
| **Memory-like** | Antigen-experienced and memory-related B-cell state. | 8 | *Not evaluable* | *one patient contributes 100.0% of cells; pCR n=0; requires >= 5; non-pCR n=1; requires >= 5* | 3 | 3 | 100% | *CD27, TNFRSF13B, AIM2* | 0 | 1 | | 0.323 | | 0.323 | | | | | | | | | No | No | No | |
| **Memory-like** | Antigen-experienced and memory-related B-cell state. | 9 | *Not evaluable* | *pCR n=1; requires >= 5; non-pCR n=0; requires >= 5* | 3 | 3 | 100% | *CD27, TNFRSF13B, AIM2* | 1 | 0 | 0.003 | | 0.003 | | | | | | | | | | No | No | No | |
| **Memory-like** | Antigen-experienced and memory-related B-cell state. | 10 | *Not evaluable* | *one patient contributes 83.3% of cells; only 2 patients contribute cells; pCR n=1; requires >= 5; non-pCR n=0; requires >= 5* | 3 | 3 | 100% | *CD27, TNFRSF13B, AIM2* | 1 | 0 | 0.307 | | 0.307 | | | | | | | | | | No | No | No | |
| **Activated/APC-like** | B-cell activation and antigen-presentation program. | 0 | *Not evaluable* | *one patient contributes 87.7% of cells; pCR n=1; requires >= 5; non-pCR n=1; requires >= 5* | 6 | 6 | 100% | *CD83, CD69, HLA-DRA, HLA-DRB1, NFKBIA, NR4A2* | 1 | 1 | 0.519 | 0.692 | 0.519 | 0.692 | -0.173 | | | | | | | | No | No | No | |
| **Activated/APC-like** | B-cell activation and antigen-presentation program. | 1 | Evaluable | | 6 | 6 | 100% | *CD83, CD69, HLA-DRA, HLA-DRB1, NFKBIA, NR4A2* | 8 | 6 | 0.924 | 0.916 | 0.977 | 0.957 | 0.020 | 24.0 | 1.0000 | 0.500 | 0.000 | 0.000 | 1.0000 | 1.0000 | No | No | No | |
| **Activated/APC-like** | B-cell activation and antigen-presentation program. | 2 | Evaluable | | 6 | 6 | 100% | *CD83, CD69, HLA-DRA, HLA-DRB1, NFKBIA, NR4A2* | 11 | 6 | 0.985 | 1.024 | 1.097 | 1.059 | 0.038 | 32.0 | 0.9612 | 0.485 | -0.030 | -0.030 | 1.0000 | 1.0000 | No | No | No | Higher in non-pCR |
| **Activated/APC-like** | B-cell activation and antigen-presentation program. | 3 | *Not evaluable* | *mixed B/T-lineage cluster: CD3D, CD3E, IL7R and GIMAP7 signal* | 6 | 6 | 100% | *CD83, CD69, HLA-DRA, HLA-DRB1, NFKBIA, NR4A2* | 9 | 5 | 0.992 | 0.869 | 0.857 | 0.948 | -0.091 | | | | | | | | No | No | No | |
| **Activated/APC-like** | B-cell activation and antigen-presentation program. | 4 | *Not evaluable* | *pCR n=3; requires >= 5; non-pCR n=1; requires >= 5* | 6 | 6 | 100% | *CD83, CD69, HLA-DRA, HLA-DRB1, NFKBIA, NR4A2* | 3 | 1 | 2.006 | 1.788 | 2.055 | 1.788 | 0.267 | | | | | | | | No | No | No | |
| **Activated/APC-like** | B-cell activation and antigen-presentation program. | 5 | *Not evaluable* | *pCR n=2; requires >= 5; non-pCR n=2; requires >= 5* | 6 | 6 | 100% | *CD83, CD69, HLA-DRA, HLA-DRB1, NFKBIA, NR4A2* | 2 | 2 | 0.231 | 0.176 | 0.231 | 0.176 | 0.054 | | | | | | | | No | No | No | |
| **Activated/APC-like** | B-cell activation and antigen-presentation program. | 6 | *Not evaluable* | *pCR n=3; requires >= 5; non-pCR n=1; requires >= 5* | 6 | 6 | 100% | *CD83, CD69, HLA-DRA, HLA-DRB1, NFKBIA, NR4A2* | 3 | 1 | 0.325 | 0.311 | 0.374 | 0.311 | 0.063 | | | | | | | | No | No | No | |
| **Activated/APC-like** | B-cell activation and antigen-presentation program. | 7 | *Not evaluable* | *pCR n=1; requires >= 5; non-pCR n=3; requires >= 5* | 6 | 6 | 100% | *CD83, CD69, HLA-DRA, HLA-DRB1, NFKBIA, NR4A2* | 1 | 3 | 1.288 | 1.287 | 1.288 | 1.365 | -0.077 | | | | | | | | No | No | No | |
| **Activated/APC-like** | B-cell activation and antigen-presentation program. | 8 | *Not evaluable* | *one patient contributes 100.0% of cells; pCR n=0; requires >= 5; non-pCR n=1; requires >= 5* | 6 | 6 | 100% | *CD83, CD69, HLA-DRA, HLA-DRB1, NFKBIA, NR4A2* | 0 | 1 | | 3.371 | | 3.371 | | | | | | | | | No | No | No | |
| **Activated/APC-like** | B-cell activation and antigen-presentation program. | 9 | *Not evaluable* | *pCR n=1; requires >= 5; non-pCR n=0; requires >= 5* | 6 | 6 | 100% | *CD83, CD69, HLA-DRA, HLA-DRB1, NFKBIA, NR4A2* | 1 | 0 | 0.519 | | 0.519 | | | | | | | | | | No | No | No | |
| **Activated/APC-like** | B-cell activation and antigen-presentation program. | 10 | *Not evaluable* | *one patient contributes 83.3% of cells; only 2 patients contribute cells; pCR n=1; requires >= 5; non-pCR n=0; requires >= 5* | 6 | 6 | 100% | *CD83, CD69, HLA-DRA, HLA-DRB1, NFKBIA, NR4A2* | 1 | 0 | 0.177 | | 0.177 | | | | | | | | | | No | No | No | |
| **CD38/plasma-like** | CD38-associated antibody-secreting/plasma-cell differentiation. | 0 | *Not evaluable* | *one patient contributes 87.7% of cells; pCR n=1; requires >= 5; non-pCR n=1; requires >= 5* | 6 | 6 | 100% | *CD38, MZB1, XBP1, JCHAIN, PRDM1, SDC1* | 1 | 1 | 0.084 | 0.068 | 0.084 | 0.068 | 0.016 | | | | | | | | No | No | No | |
| **CD38/plasma-like** | CD38-associated antibody-secreting/plasma-cell differentiation. | 1 | Evaluable | | 6 | 6 | 100% | *CD38, MZB1, XBP1, JCHAIN, PRDM1, SDC1* | 8 | 6 | 0.122 | 0.160 | 0.121 | 0.181 | -0.060 | 10.0 | 0.0813 | 0.208 | -0.583 | -0.583 | 0.1625 | 0.4063 | No | No | No | Higher in non-pCR |
| **CD38/plasma-like** | CD38-associated antibody-secreting/plasma-cell differentiation. | 2 | Evaluable | | 6 | 6 | 100% | *CD38, MZB1, XBP1, JCHAIN, PRDM1, SDC1* | 11 | 6 | 0.127 | 0.123 | 0.124 | 0.124 | -0.001 | 36.0 | 0.8075 | 0.545 | 0.091 | 0.091 | 0.8075 | 1.0000 | No | No | No | Higher in pCR |
| **CD38/plasma-like** | CD38-associated antibody-secreting/plasma-cell differentiation. | 3 | *Not evaluable* | *mixed B/T-lineage cluster: CD3D, CD3E, IL7R and GIMAP7 signal* | 6 | 6 | 100% | *CD38, MZB1, XBP1, JCHAIN, PRDM1, SDC1* | 9 | 5 | 0.176 | 0.157 | 0.154 | 0.174 | -0.020 | | | | | | | | No | No | No | |
| **CD38/plasma-like** | CD38-associated antibody-secreting/plasma-cell differentiation. | 4 | *Not evaluable* | *pCR n=3; requires >= 5; non-pCR n=1; requires >= 5* | 6 | 6 | 100% | *CD38, MZB1, XBP1, JCHAIN, PRDM1, SDC1* | 3 | 1 | 0.199 | 0.197 | 0.188 | 0.197 | -0.009 | | | | | | | | No | No | No | |
| **CD38/plasma-like** | CD38-associated antibody-secreting/plasma-cell differentiation. | 5 | *Not evaluable* | *pCR n=2; requires >= 5; non-pCR n=2; requires >= 5* | 6 | 6 | 100% | *CD38, MZB1, XBP1, JCHAIN, PRDM1, SDC1* | 2 | 2 | 1.633 | 1.428 | 1.633 | 1.428 | 0.206 | | | | | | | | No | No | No | |
| **CD38/plasma-like** | CD38-associated antibody-secreting/plasma-cell differentiation. | 6 | *Not evaluable* | *pCR n=3; requires >= 5; non-pCR n=1; requires >= 5* | 6 | 6 | 100% | *CD38, MZB1, XBP1, JCHAIN, PRDM1, SDC1* | 3 | 1 | 0.185 | 0.222 | 0.186 | 0.222 | -0.036 | | | | | | | | No | No | No | |
| **CD38/plasma-like** | CD38-associated antibody-secreting/plasma-cell differentiation. | 7 | *Not evaluable* | *pCR n=1; requires >= 5; non-pCR n=3; requires >= 5* | 6 | 6 | 100% | *CD38, MZB1, XBP1, JCHAIN, PRDM1, SDC1* | 1 | 3 | 0.062 | 0.135 | 0.062 | 0.114 | -0.051 | | | | | | | | No | No | No | |
| **CD38/plasma-like** | CD38-associated antibody-secreting/plasma-cell differentiation. | 8 | *Not evaluable* | *one patient contributes 100.0% of cells; pCR n=0; requires >= 5; non-pCR n=1; requires >= 5* | 6 | 6 | 100% | *CD38, MZB1, XBP1, JCHAIN, PRDM1, SDC1* | 0 | 1 | | 0.173 | | 0.173 | | | | | | | | | No | No | No | |
| **CD38/plasma-like** | CD38-associated antibody-secreting/plasma-cell differentiation. | 9 | *Not evaluable* | *pCR n=1; requires >= 5; non-pCR n=0; requires >= 5* | 6 | 6 | 100% | *CD38, MZB1, XBP1, JCHAIN, PRDM1, SDC1* | 1 | 0 | 0.192 | | 0.192 | | | | | | | | | | No | No | No | |
| **CD38/plasma-like** | CD38-associated antibody-secreting/plasma-cell differentiation. | 10 | *Not evaluable* | *one patient contributes 83.3% of cells; only 2 patients contribute cells; pCR n=1; requires >= 5; non-pCR n=0; requires >= 5* | 6 | 6 | 100% | *CD38, MZB1, XBP1, JCHAIN, PRDM1, SDC1* | 1 | 0 | 1.238 | | 1.238 | | | | | | | | | | No | No | No | |
| **ISG/stress** | Interferon-response and immediate cellular-stress program. | 0 | *Not evaluable* | *one patient contributes 87.7% of cells; pCR n=1; requires >= 5; non-pCR n=1; requires >= 5* | 9 | 9 | 100% | *ISG15, IFIT1, IFIT2, IFIT3, MX1, FOS, JUN, DUSP1, HSPA1A* | 1 | 1 | 0.171 | 0.411 | 0.171 | 0.411 | -0.240 | | | | | | | | No | No | No | |
| **ISG/stress** | Interferon-response and immediate cellular-stress program. | 1 | Evaluable | | 9 | 9 | 100% | *ISG15, IFIT1, IFIT2, IFIT3, MX1, FOS, JUN, DUSP1, HSPA1A* | 8 | 6 | 0.094 | 0.150 | 0.092 | 0.116 | -0.024 | 23.0 | 0.9497 | 0.479 | -0.042 | -0.042 | 0.9497 | 1.0000 | No | No | No | Higher in non-pCR |
| **ISG/stress** | Interferon-response and immediate cellular-stress program. | 2 | Evaluable | | 9 | 9 | 100% | *ISG15, IFIT1, IFIT2, IFIT3, MX1, FOS, JUN, DUSP1, HSPA1A* | 11 | 6 | 0.116 | 0.102 | 0.072 | 0.078 | -0.006 | 36.0 | 0.8075 | 0.545 | 0.091 | 0.091 | 0.9497 | 1.0000 | No | No | No | Higher in pCR |
| **ISG/stress** | Interferon-response and immediate cellular-stress program. | 3 | *Not evaluable* | *mixed B/T-lineage cluster: CD3D, CD3E, IL7R and GIMAP7 signal* | 9 | 9 | 100% | *ISG15, IFIT1, IFIT2, IFIT3, MX1, FOS, JUN, DUSP1, HSPA1A* | 9 | 5 | 0.343 | 0.195 | 0.144 | 0.235 | -0.090 | | | | | | | | No | No | No | |
| **ISG/stress** | Interferon-response and immediate cellular-stress program. | 4 | *Not evaluable* | *pCR n=3; requires >= 5; non-pCR n=1; requires >= 5* | 9 | 9 | 100% | *ISG15, IFIT1, IFIT2, IFIT3, MX1, FOS, JUN, DUSP1, HSPA1A* | 3 | 1 | 1.014 | 1.009 | 1.080 | 1.009 | 0.071 | | | | | | | | No | No | No | |
| **ISG/stress** | Interferon-response and immediate cellular-stress program. | 5 | *Not evaluable* | *pCR n=2; requires >= 5; non-pCR n=2; requires >= 5* | 9 | 9 | 100% | *ISG15, IFIT1, IFIT2, IFIT3, MX1, FOS, JUN, DUSP1, HSPA1A* | 2 | 2 | 0.100 | 0.122 | 0.100 | 0.122 | -0.022 | | | | | | | | No | No | No | |
| **ISG/stress** | Interferon-response and immediate cellular-stress program. | 6 | *Not evaluable* | *pCR n=3; requires >= 5; non-pCR n=1; requires >= 5* | 9 | 9 | 100% | *ISG15, IFIT1, IFIT2, IFIT3, MX1, FOS, JUN, DUSP1, HSPA1A* | 3 | 1 | 0.139 | 0.182 | 0.134 | 0.182 | -0.049 | | | | | | | | No | No | No | |
| **ISG/stress** | Interferon-response and immediate cellular-stress program. | 7 | *Not evaluable* | *pCR n=1; requires >= 5; non-pCR n=3; requires >= 5* | 9 | 9 | 100% | *ISG15, IFIT1, IFIT2, IFIT3, MX1, FOS, JUN, DUSP1, HSPA1A* | 1 | 3 | 0.120 | 0.127 | 0.120 | 0.114 | 0.006 | | | | | | | | No | No | No | |
| **ISG/stress** | Interferon-response and immediate cellular-stress program. | 8 | *Not evaluable* | *one patient contributes 100.0% of cells; pCR n=0; requires >= 5; non-pCR n=1; requires >= 5* | 9 | 9 | 100% | *ISG15, IFIT1, IFIT2, IFIT3, MX1, FOS, JUN, DUSP1, HSPA1A* | 0 | 1 | | 0.856 | | 0.856 | | | | | | | | | No | No | No | |
| **ISG/stress** | Interferon-response and immediate cellular-stress program. | 9 | *Not evaluable* | *pCR n=1; requires >= 5; non-pCR n=0; requires >= 5* | 9 | 9 | 100% | *ISG15, IFIT1, IFIT2, IFIT3, MX1, FOS, JUN, DUSP1, HSPA1A* | 1 | 0 | 0.122 | | 0.122 | | | | | | | | | | No | No | No | |
| **ISG/stress** | Interferon-response and immediate cellular-stress program. | 10 | *Not evaluable* | *one patient contributes 83.3% of cells; only 2 patients contribute cells; pCR n=1; requires >= 5; non-pCR n=0; requires >= 5* | 9 | 9 | 100% | *ISG15, IFIT1, IFIT2, IFIT3, MX1, FOS, JUN, DUSP1, HSPA1A* | 1 | 0 | 0.093 | | 0.093 | | | | | | | | | | No | No | No | |

APC, antigen-presenting cell; AUC, area under the receiver operating characteristic curve; BH, Benjamini–Hochberg; FDR, false discovery rate; ISG, interferon-stimulated gene; n, number of patients; pCR, pathological complete response; non-pCR, absence of pCR; q, FDR-adjusted P value.

**Supplementary Table 7. Gene-level comparisons within B-cell functional programs according to pathological response.**

| B-cell program | B-cell subcluster | Gene | Metric | n pCR | n non-pCR | Mean, pCR | Mean, non-pCR | Median, pCR | Median, non-pCR | Mann–Whitney U | Two-sided P | AUC (pCR higher) | Signed AUC | Cliff's δ (pCR − non-pCR) | Benjamini–Hochberg q within program | Global Benjamini–Hochberg q | Nominal P <0.05 | Within-program FDR q <0.10 | Global FDR q <0.10 |
|---|---|---|---|---|---|---|---|---|---|---|---|---|---|---|---|---|---|---|---|
| Naive-like | 1 | *IGHM* | % positive | 8 | 6 | 47.082 | 33.198 | 46.257 | 32.530 | 42. | 0.0200 | 0.8750 | 0.7500 | 0.7500 | 0.1758 | 0.9145 | Yes | No | No |
| CD38/plasma-like | 2 | *CD38* | % positive | 11 | 6 | 15.325 | 9.438 | 15.556 | 8.942 | 56. | 0.0202 | 0.8485 | 0.6970 | 0.6970 | 0.3410 | 0.9145 | Yes | No | No |
| Naive-like | 1 | *IGHD* | Mean expression | 8 | 6 | 0.203 | 0.133 | 0.201 | 0.133 | 41. | 0.0293 | 0.8542 | 0.7083 | 0.7083 | 0.1758 | 0.9145 | Yes | No | No |
| CD38/plasma-like | 2 | *CD38* | Mean expression | 11 | 6 | 0.302 | 0.195 | 0.321 | 0.209 | 54. | 0.0365 | 0.8182 | 0.6364 | 0.6364 | 0.3410 | 0.9145 | Yes | No | No |
| CD38/plasma-like | 1 | *XBP1* | Mean expression | 8 | 6 | 0.046 | 0.083 | 0.043 | 0.074 | 8. | 0.0426 | 0.1667 | -0.6667 | -0.6667 | 0.3410 | 0.9145 | Yes | No | No |
| CD38/plasma-like | 1 | *PRDM1* | Mean expression | 8 | 6 | 0.032 | 0.014 | 0.027 | 0.015 | 39. | 0.0593 | 0.8125 | 0.6250 | 0.6250 | 0.3556 | 0.9145 | No | No | No |
| Naive-like | 1 | *IGHM* | Mean expression | 8 | 6 | 1.102 | 0.809 | 0.981 | 0.807 | 39. | 0.0593 | 0.8125 | 0.6250 | 0.6250 | 0.2371 | 0.9145 | No | No | No |
| Naive-like | 1 | *IGHD* | % positive | 8 | 6 | 11.830 | 7.409 | 11.464 | 7.886 | 37. | 0.1079 | 0.7708 | 0.5417 | 0.5417 | 0.3237 | 1.0000 | No | No | No |
| CD38/plasma-like | 2 | *XBP1* | Mean expression | 11 | 6 | 0.046 | 0.069 | 0.022 | 0.065 | 17. | 0.1191 | 0.2576 | -0.4848 | -0.4848 | 0.5674 | 1.0000 | No | No | No |
| CD38/plasma-like | 1 | *JCHAIN* | % positive | 8 | 6 | 22.330 | 30.344 | 20.198 | 35.620 | 12. | 0.1419 | 0.2500 | -0.5000 | -0.5000 | 0.5674 | 1.0000 | No | No | No |
| CD38/plasma-like | 1 | *JCHAIN* | Mean expression | 8 | 6 | 0.431 | 0.641 | 0.343 | 0.682 | 13. | 0.1812 | 0.2708 | -0.4583 | -0.4583 | 0.6211 | 1.0000 | No | No | No |
| ISG/stress | 1 | *MX1* | % positive | 8 | 6 | 16.552 | 9.589 | 12.653 | 7.479 | 35. | 0.1812 | 0.7292 | 0.4583 | 0.4583 | 1.0000 | 1.0000 | No | No | No |
| ISG/stress | 1 | *DUSP1* | Mean expression | 8 | 6 | 0.059 | 0.116 | 0.043 | 0.064 | 14. | 0.2195 | 0.2917 | -0.4167 | -0.4167 | 1.0000 | 1.0000 | No | No | No |
| ISG/stress | 1 | *DUSP1* | % positive | 8 | 6 | 3.948 | 6.647 | 2.776 | 3.375 | 14. | 0.2195 | 0.2917 | -0.4167 | -0.4167 | 1.0000 | 1.0000 | No | No | No |
| CD38/plasma-like | 1 | *XBP1* | % positive | 8 | 6 | 3.270 | 5.085 | 3.008 | 4.345 | 14. | 0.2284 | 0.2917 | -0.4167 | -0.4167 | 0.6853 | 1.0000 | No | No | No |
| ISG/stress | 1 | *MX1* | Mean expression | 8 | 6 | 0.267 | 0.168 | 0.264 | 0.154 | 34. | 0.2284 | 0.7083 | 0.4167 | 0.4167 | 1.0000 | 1.0000 | No | No | No |
| Memory-like | 1 | *TNFRSF13B* | Mean expression | 8 | 6 | 0.346 | 0.277 | 0.378 | 0.264 | 34. | 0.2284 | 0.7083 | 0.4167 | 0.4167 | 1.0000 | 1.0000 | No | No | No |
| ISG/stress | 2 | *HSPA1A* | Mean expression | 11 | 6 | 0.110 | 0.132 | 0.030 | 0.087 | 21. | 0.2449 | 0.3182 | -0.3636 | -0.3636 | 1.0000 | 1.0000 | No | No | No |
| ISG/stress | 2 | *MX1* | Mean expression | 11 | 6 | 0.184 | 0.125 | 0.181 | 0.116 | 45. | 0.2561 | 0.6818 | 0.3636 | 0.3636 | 1.0000 | 1.0000 | No | No | No |
| CD38/plasma-like | 1 | *PRDM1* | % positive | 8 | 6 | 2.065 | 0.921 | 1.279 | 0.776 | 33. | 0.2824 | 0.6875 | 0.3750 | 0.3750 | 0.7530 | 1.0000 | No | No | No |
| ISG/stress | 1 | *HSPA1A* | Mean expression | 8 | 6 | 0.058 | 0.367 | 0.063 | 0.276 | 15. | 0.2824 | 0.3125 | -0.3750 | -0.3750 | 1.0000 | 1.0000 | No | No | No |
| ISG/stress | 2 | *MX1* | % positive | 11 | 6 | 9.747 | 6.261 | 9.259 | 5.360 | 44. | 0.3011 | 0.6667 | 0.3333 | 0.3333 | 1.0000 | 1.0000 | No | No | No |
| ISG/stress | 1 | *JUN* | Mean expression | 8 | 6 | 0.082 | 0.232 | 0.094 | 0.189 | 16. | 0.3324 | 0.3333 | -0.3333 | -0.3333 | 1.0000 | 1.0000 | No | No | No |
| ISG/stress | 1 | *JUN* | % positive | 8 | 6 | 5.408 | 12.270 | 5.852 | 9.538 | 16. | 0.3324 | 0.3333 | -0.3333 | -0.3333 | 1.0000 | 1.0000 | No | No | No |
| CD38/plasma-like | 2 | *XBP1* | % positive | 11 | 6 | 2.820 | 3.592 | 1.639 | 3.137 | 23. | 0.3394 | 0.3485 | -0.3030 | -0.3030 | 0.8145 | 1.0000 | No | No | No |
| Activated/APC-like | 1 | *NR4A2* | Mean expression | 8 | 6 | 0.064 | 0.086 | 0.027 | 0.060 | 16. | 0.3450 | 0.3333 | -0.3333 | -0.3333 | 1.0000 | 1.0000 | No | No | No |
| ISG/stress | 2 | *HSPA1A* | % positive | 11 | 6 | 5.455 | 5.983 | 1.639 | 4.259 | 24. | 0.3900 | 0.3636 | -0.2727 | -0.2727 | 1.0000 | 1.0000 | No | No | No |
| ISG/stress | 1 | *IFIT1* | Mean expression | 8 | 6 | 0.029 | 0.015 | 0.020 | 0.013 | 31. | 0.4009 | 0.6458 | 0.2917 | 0.2917 | 1.0000 | 1.0000 | No | No | No |
| ISG/stress | 1 | *IFIT1* | % positive | 8 | 6 | 1.618 | 0.924 | 1.487 | 0.693 | 31. | 0.4009 | 0.6458 | 0.2917 | 0.2917 | 1.0000 | 1.0000 | No | No | No |
| Naive-like | 2 | *IGHD* | % positive | 11 | 6 | 45.579 | 40.100 | 42.683 | 40.510 | 42. | 0.4043 | 0.6364 | 0.2727 | 0.2727 | 0.8075 | 1.0000 | No | No | No |
| Activated/APC-like | 1 | *NR4A2* | % positive | 8 | 6 | 4.457 | 5.091 | 1.732 | 3.160 | 17. | 0.4136 | 0.3542 | -0.2917 | -0.2917 | 1.0000 | 1.0000 | No | No | No |
| Memory-like | 1 | *AIM2* | Mean expression | 8 | 6 | 0.749 | 0.698 | 0.814 | 0.651 | 31. | 0.4136 | 0.6458 | 0.2917 | 0.2917 | 1.0000 | 1.0000 | No | No | No |
| Memory-like | 1 | *AIM2* | % positive | 8 | 6 | 41.357 | 35.841 | 45.531 | 34.769 | 31. | 0.4136 | 0.6458 | 0.2917 | 0.2917 | 1.0000 | 1.0000 | No | No | No |
| Memory-like | 1 | *TNFRSF13B* | % positive | 8 | 6 | 21.336 | 16.205 | 22.441 | 15.317 | 31. | 0.4136 | 0.6458 | 0.2917 | 0.2917 | 1.0000 | 1.0000 | No | No | No |
| Activated/APC-like | 2 | *CD83* | Mean expression | 11 | 6 | 0.323 | 0.243 | 0.286 | 0.192 | 41. | 0.4623 | 0.6212 | 0.2424 | 0.2424 | 1.0000 | 1.0000 | No | No | No |
| Activated/APC-like | 2 | *CD83* | % positive | 11 | 6 | 17.451 | 11.994 | 16.079 | 9.703 | 41. | 0.4623 | 0.6212 | 0.2424 | 0.2424 | 1.0000 | 1.0000 | No | No | No |
| ISG/stress | 1 | *FOS* | Mean expression | 8 | 6 | 0.118 | 0.200 | 0.066 | 0.131 | 18. | 0.4772 | 0.3750 | -0.2500 | -0.2500 | 1.0000 | 1.0000 | No | No | No |
| Activated/APC-like | 1 | *CD69* | Mean expression | 8 | 6 | 0.155 | 0.191 | 0.146 | 0.171 | 18. | 0.4908 | 0.3750 | -0.2500 | -0.2500 | 1.0000 | 1.0000 | No | No | No |
| ISG/stress | 1 | *HSPA1A* | % positive | 8 | 6 | 4.058 | 16.393 | 4.173 | 12.740 | 18. | 0.4908 | 0.3750 | -0.2500 | -0.2500 | 1.0000 | 1.0000 | No | No | No |
| Memory-like | 1 | *CD27* | % positive | 8 | 6 | 21.972 | 17.126 | 20.149 | 18.406 | 30. | 0.4908 | 0.6250 | 0.2500 | 0.2500 | 1.0000 | 1.0000 | No | No | No |
| Naive-like | 1 | *TCL1A* | % positive | 8 | 6 | 2.821 | 2.072 | 2.780 | 2.218 | 30. | 0.4908 | 0.6250 | 0.2500 | 0.2500 | 0.8075 | 1.0000 | No | No | No |
| ISG/stress | 2 | *IFIT3* | Mean expression | 11 | 6 | 0.027 | 0.011 | 0.014 | 0.009 | 40. | 0.5043 | 0.6061 | 0.2121 | 0.2121 | 1.0000 | 1.0000 | No | No | No |
| ISG/stress | 2 | *IFIT3* | % positive | 11 | 6 | 1.494 | 0.606 | 0.797 | 0.477 | 40. | 0.5043 | 0.6061 | 0.2121 | 0.2121 | 1.0000 | 1.0000 | No | No | No |
| Memory-like | 2 | *CD27* | % positive | 11 | 6 | 0.827 | 0.594 | 1.163 | 0.529 | 40. | 0.5083 | 0.6061 | 0.2121 | 0.2121 | 1.0000 | 1.0000 | No | No | No |
| ISG/stress | 1 | *FOS* | % positive | 8 | 6 | 7.063 | 10.319 | 4.096 | 6.261 | 19. | 0.5608 | 0.3958 | -0.2083 | -0.2083 | 1.0000 | 1.0000 | No | No | No |
| CD38/plasma-like | 1 | *CD38* | Mean expression | 8 | 6 | 0.062 | 0.057 | 0.049 | 0.068 | 19. | 0.5728 | 0.3958 | -0.2083 | -0.2083 | 1.0000 | 1.0000 | No | No | No |
| ISG/stress | 1 | *ISG15* | Mean expression | 8 | 6 | 0.115 | 0.141 | 0.116 | 0.153 | 19. | 0.5728 | 0.3958 | -0.2083 | -0.2083 | 1.0000 | 1.0000 | No | No | No |
| Naive-like | 1 | *TCL1A* | Mean expression | 8 | 6 | 0.053 | 0.042 | 0.056 | 0.045 | 29. | 0.5728 | 0.6042 | 0.2083 | 0.2083 | 0.8075 | 1.0000 | No | No | No |
| ISG/stress | 2 | *IFIT1* | % positive | 11 | 6 | 1.143 | 0.679 | 0.881 | 0.512 | 39. | 0.5781 | 0.5909 | 0.1818 | 0.1818 | 1.0000 | 1.0000 | No | No | No |
| CD38/plasma-like | 2 | *JCHAIN* | Mean expression | 11 | 6 | 0.183 | 0.231 | 0.133 | 0.239 | 27. | 0.5908 | 0.4091 | -0.1818 | -0.1818 | 1.0000 | 1.0000 | No | No | No |
| Naive-like | 2 | *IGHM* | % positive | 11 | 6 | 75.341 | 72.752 | 78.689 | 73.459 | 39. | 0.5908 | 0.5909 | 0.1818 | 0.1818 | 0.8075 | 1.0000 | No | No | No |
| Activated/APC-like | 2 | *NR4A2* | Mean expression | 11 | 6 | 0.089 | 0.082 | 0.021 | 0.069 | 28. | 0.6470 | 0.4242 | -0.1515 | -0.1515 | 1.0000 | 1.0000 | No | No | No |
| Activated/APC-like | 2 | *HLA-DRA* | % positive | 11 | 6 | 79.211 | 84.728 | 88.421 | 93.752 | 28. | 0.6605 | 0.4242 | -0.1515 | -0.1515 | 1.0000 | 1.0000 | No | No | No |
| Naive-like | 2 | *IGHM* | Mean expression | 11 | 6 | 1.895 | 1.940 | 1.943 | 1.837 | 28. | 0.6605 | 0.4242 | -0.1515 | -0.1515 | 0.8075 | 1.0000 | No | No | No |
| Activated/APC-like | 1 | *CD83* | Mean expression | 8 | 6 | 0.144 | 0.145 | 0.148 | 0.119 | 28. | 0.6620 | 0.5833 | 0.1667 | 0.1667 | 1.0000 | 1.0000 | No | No | No |
| Activated/APC-like | 1 | *HLA-DRB1* | Mean expression | 8 | 6 | 2.083 | 1.937 | 2.330 | 2.111 | 28. | 0.6620 | 0.5833 | 0.1667 | 0.1667 | 1.0000 | 1.0000 | No | No | No |
| Activated/APC-like | 1 | *HLA-DRB1* | % positive | 8 | 6 | 77.563 | 73.089 | 87.110 | 84.468 | 28. | 0.6620 | 0.5833 | 0.1667 | 0.1667 | 1.0000 | 1.0000 | No | No | No |
| Activated/APC-like | 1 | *NFKBIA* | Mean expression | 8 | 6 | 0.314 | 0.338 | 0.341 | 0.351 | 20. | 0.6620 | 0.4167 | -0.1667 | -0.1667 | 1.0000 | 1.0000 | No | No | No |
| ISG/stress | 1 | *ISG15* | % positive | 8 | 6 | 7.341 | 8.369 | 6.559 | 9.500 | 20. | 0.6620 | 0.4167 | -0.1667 | -0.1667 | 1.0000 | 1.0000 | No | No | No |
| Memory-like | 1 | *CD27* | Mean expression | 8 | 6 | 0.351 | 0.304 | 0.354 | 0.328 | 28. | 0.6620 | 0.5833 | 0.1667 | 0.1667 | 1.0000 | 1.0000 | No | No | No |
| ISG/stress | 2 | *IFIT2* | % positive | 11 | 6 | 5.267 | 4.998 | 4.918 | 3.973 | 37. | 0.7249 | 0.5606 | 0.1212 | 0.1212 | 1.0000 | 1.0000 | No | No | No |
| ISG/stress | 2 | *JUN* | Mean expression | 11 | 6 | 0.165 | 0.162 | 0.067 | 0.113 | 29. | 0.7249 | 0.4394 | -0.1212 | -0.1212 | 1.0000 | 1.0000 | No | No | No |
| Activated/APC-like | 2 | *HLA-DRA* | Mean expression | 11 | 6 | 2.655 | 2.877 | 2.885 | 3.132 | 29. | 0.7325 | 0.4394 | -0.1212 | -0.1212 | 1.0000 | 1.0000 | No | No | No |
| Activated/APC-like | 2 | *NFKBIA* | Mean expression | 11 | 6 | 0.352 | 0.369 | 0.292 | 0.396 | 29. | 0.7325 | 0.4394 | -0.1212 | -0.1212 | 1.0000 | 1.0000 | No | No | No |
| Activated/APC-like | 1 | *CD69* | % positive | 8 | 6 | 9.352 | 10.647 | 8.864 | 10.475 | 21. | 0.7546 | 0.4375 | -0.1250 | -0.1250 | 1.0000 | 1.0000 | No | No | No |
| ISG/stress | 1 | *IFIT2* | % positive | 8 | 6 | 5.405 | 4.988 | 5.254 | 4.466 | 27. | 0.7546 | 0.5625 | 0.1250 | 0.1250 | 1.0000 | 1.0000 | No | No | No |
| Activated/APC-like | 2 | *NR4A2* | % positive | 11 | 6 | 4.728 | 4.471 | 1.116 | 3.782 | 30. | 0.7992 | 0.4545 | -0.0909 | -0.0909 | 1.0000 | 1.0000 | No | No | No |
| ISG/stress | 2 | *IFIT1* | Mean expression | 11 | 6 | 0.022 | 0.013 | 0.014 | 0.011 | 36. | 0.8004 | 0.5455 | 0.0909 | 0.0909 | 1.0000 | 1.0000 | No | No | No |
| ISG/stress | 2 | *ISG15* | Mean expression | 11 | 6 | 0.086 | 0.098 | 0.095 | 0.091 | 30. | 0.8011 | 0.4545 | -0.0909 | -0.0909 | 1.0000 | 1.0000 | No | No | No |
| Activated/APC-like | 2 | *CD69* | Mean expression | 11 | 6 | 0.298 | 0.327 | 0.247 | 0.267 | 30. | 0.8075 | 0.4545 | -0.0909 | -0.0909 | 1.0000 | 1.0000 | No | No | No |
| CD38/plasma-like | 2 | *JCHAIN* | % positive | 11 | 6 | 10.384 | 10.751 | 7.368 | 12.350 | 36. | 0.8075 | 0.5455 | 0.0909 | 0.0909 | 1.0000 | 1.0000 | No | No | No |
| Memory-like | 2 | *AIM2* | Mean expression | 11 | 6 | 0.093 | 0.076 | 0.080 | 0.068 | 36. | 0.8075 | 0.5455 | 0.0909 | 0.0909 | 1.0000 | 1.0000 | No | No | No |

| | | | | | | | | | | | | | | | | | | | |
|---|---|---|---|---|---|---|---|---|---|---|---|---|---|---|---|---|---|---|---|
| **Memory-like** | 2 | *AIM2* | % positive | 11 | 6 | 4.537 | 3.453 | 3.791 | 3.544 | 36. | 0.8075 | 0.5455 | 0.0909 | 0.0909 | 1.0000 | 1.0000 | No | No | No |
| **Naive-like** | 2 | *IGHD* | Mean expression | 11 | 6 | 0.947 | 0.895 | 0.979 | 0.931 | 36. | 0.8075 | 0.5455 | 0.0909 | 0.0909 | 0.8075 | 1.0000 | No | No | No |
| **Naive-like** | 2 | *TCL1A* | Mean expression | 11 | 6 | 1.250 | 1.341 | 1.219 | 1.511 | 30. | 0.8075 | 0.4545 | -0.0909 | -0.0909 | 0.8075 | 1.0000 | No | No | No |
| **Naive-like** | 2 | *TCL1A* | % positive | 11 | 6 | 53.164 | 54.950 | 56.778 | 63.996 | 30. | 0.8075 | 0.4545 | -0.0909 | -0.0909 | 0.8075 | 1.0000 | No | No | No |
| **Activated/APC-like** | 1 | *CD83* | % positive | 8 | 6 | 8.974 | 8.426 | 8.250 | 7.244 | 26. | 0.8518 | 0.5417 | 0.0833 | 0.0833 | 1.0000 | 1.0000 | No | No | No |
| **CD38/plasma-like** | 1 | *CD38* | % positive | 8 | 6 | 3.389 | 3.143 | 2.484 | 3.091 | 22. | 0.8518 | 0.4583 | -0.0833 | -0.0833 | 1.0000 | 1.0000 | No | No | No |
| **CD38/plasma-like** | 1 | *MZB1* | Mean expression | 8 | 6 | 0.163 | 0.164 | 0.145 | 0.182 | 22. | 0.8518 | 0.4583 | -0.0833 | -0.0833 | 1.0000 | 1.0000 | No | No | No |
| **CD38/plasma-like** | 1 | *MZB1* | % positive | 8 | 6 | 10.698 | 9.431 | 9.401 | 10.555 | 26. | 0.8518 | 0.5417 | 0.0833 | 0.0833 | 1.0000 | 1.0000 | No | No | No |
| **Memory-like** | 2 | *TNFRSF13B* | % positive | 11 | 6 | 0.915 | 0.841 | 1.116 | 0.794 | 35. | 0.8787 | 0.5303 | 0.0606 | 0.0606 | 1.0000 | 1.0000 | No | No | No |
| **CD38/plasma-like** | 2 | *PRDM1* | % positive | 11 | 6 | 0.606 | 0.599 | 0.474 | 0.425 | 31. | 0.8794 | 0.4697 | -0.0606 | -0.0606 | 1.0000 | 1.0000 | No | No | No |
| **ISG/stress** | 2 | *IFIT2* | Mean expression | 11 | 6 | 0.089 | 0.098 | 0.088 | 0.087 | 31. | 0.8801 | 0.4697 | -0.0606 | -0.0606 | 1.0000 | 1.0000 | No | No | No |
| **Activated/APC-like** | 2 | *HLA-DRB1* | Mean expression | 11 | 6 | 2.194 | 2.245 | 2.336 | 2.348 | 31. | 0.8836 | 0.4697 | -0.0606 | -0.0606 | 1.0000 | 1.0000 | No | No | No |
| **Activated/APC-like** | 2 | *NFKBIA* | % positive | 11 | 6 | 19.853 | 17.858 | 17.073 | 18.753 | 31. | 0.8836 | 0.4697 | -0.0606 | -0.0606 | 1.0000 | 1.0000 | No | No | No |
| **CD38/plasma-like** | 2 | *MZB1* | % positive | 11 | 6 | 12.338 | 11.671 | 11.475 | 9.258 | 35. | 0.8836 | 0.5303 | 0.0606 | 0.0606 | 1.0000 | 1.0000 | No | No | No |
| **ISG/stress** | 1 | *IFIT3* | % positive | 8 | 6 | 2.157 | 2.238 | 2.144 | 1.331 | 25.5 | 0.8972 | 0.5313 | 0.0625 | 0.0625 | 1.0000 | 1.0000 | No | No | No |
| **ISG/stress** | 2 | *DUSP1* | Mean expression | 11 | 6 | 0.108 | 0.092 | 0.054 | 0.061 | 31.5 | 0.9195 | 0.4773 | -0.0455 | -0.0455 | 1.0000 | 1.0000 | No | No | No |
| **Activated/APC-like** | 1 | *HLA-DRA* | % positive | 8 | 6 | 86.514 | 87.617 | 92.011 | 94.035 | 25. | 0.9497 | 0.5208 | 0.0417 | 0.0417 | 1.0000 | 1.0000 | No | No | No |
| **ISG/stress** | 1 | *IFIT3* | Mean expression | 8 | 6 | 0.035 | 0.035 | 0.024 | 0.026 | 23. | 0.9497 | 0.4792 | -0.0417 | -0.0417 | 1.0000 | 1.0000 | No | No | No |
| **Memory-like** | 2 | *CD27* | Mean expression | 11 | 6 | 0.015 | 0.012 | 0.018 | 0.010 | 32. | 0.9594 | 0.4848 | -0.0303 | -0.0303 | 1.0000 | 1.0000 | No | No | No |
| **Activated/APC-like** | 2 | *CD69* | % positive | 11 | 6 | 16.272 | 16.168 | 14.634 | 14.280 | 32. | 0.9612 | 0.4848 | -0.0303 | -0.0303 | 1.0000 | 1.0000 | No | No | No |
| **Activated/APC-like** | 2 | *HLA-DRB1* | % positive | 11 | 6 | 76.281 | 74.503 | 80.233 | 83.339 | 32. | 0.9612 | 0.4848 | -0.0303 | -0.0303 | 1.0000 | 1.0000 | No | No | No |
| **CD38/plasma-like** | 2 | *MZB1* | Mean expression | 11 | 6 | 0.218 | 0.231 | 0.208 | 0.202 | 32. | 0.9612 | 0.4848 | -0.0303 | -0.0303 | 1.0000 | 1.0000 | No | No | No |
| **Activated/APC-like** | 1 | *HLA-DRA* | Mean expression | 8 | 6 | 2.782 | 2.798 | 2.970 | 2.984 | 24. | 1.0000 | 0.5000 | 0.0000 | 0.0000 | 1.0000 | 1.0000 | No | No | No |
| **Activated/APC-like** | 1 | *NFKBIA* | % positive | 8 | 6 | 19.741 | 18.705 | 20.599 | 17.683 | 24. | 1.0000 | 0.5000 | 0.0000 | 0.0000 | 1.0000 | 1.0000 | No | No | No |
| **CD38/plasma-like** | 1 | *SDC1* | Mean expression | 8 | 6 | 0.000 | 0.000 | 0.000 | 0.000 | 24. | 1.0000 | 0.5000 | 0.0000 | 0.0000 | 1.0000 | 1.0000 | No | No | No |
| **CD38/plasma-like** | 1 | *SDC1* | % positive | 8 | 6 | 0.000 | 0.000 | 0.000 | 0.000 | 24. | 1.0000 | 0.5000 | 0.0000 | 0.0000 | 1.0000 | 1.0000 | No | No | No |
| **CD38/plasma-like** | 2 | *PRDM1* | Mean expression | 11 | 6 | 0.010 | 0.011 | 0.011 | 0.008 | 33. | 1.0000 | 0.5000 | 0.0000 | 0.0000 | 1.0000 | 1.0000 | No | No | No |
| **CD38/plasma-like** | 2 | *SDC1* | Mean expression | 11 | 6 | 0.000 | 0.000 | 0.000 | 0.000 | 33. | 1.0000 | 0.5000 | 0.0000 | 0.0000 | 1.0000 | 1.0000 | No | No | No |
| **CD38/plasma-like** | 2 | *SDC1* | % positive | 11 | 6 | 0.000 | 0.000 | 0.000 | 0.000 | 33. | 1.0000 | 0.5000 | 0.0000 | 0.0000 | 1.0000 | 1.0000 | No | No | No |
| **ISG/stress** | 1 | *IFIT2* | Mean expression | 8 | 6 | 0.085 | 0.079 | 0.094 | 0.075 | 24. | 1.0000 | 0.5000 | 0.0000 | 0.0000 | 1.0000 | 1.0000 | No | No | No |
| **ISG/stress** | 2 | *DUSP1* | % positive | 11 | 6 | 6.190 | 4.745 | 3.279 | 3.144 | 33.5 | 1.0000 | 0.5076 | 0.0152 | 0.0152 | 1.0000 | 1.0000 | No | No | No |
| **ISG/stress** | 2 | *FOS* | Mean expression | 11 | 6 | 0.251 | 0.184 | 0.115 | 0.128 | 32.5 | 1.0000 | 0.4924 | -0.0152 | -0.0152 | 1.0000 | 1.0000 | No | No | No |
| **ISG/stress** | 2 | *FOS* | % positive | 11 | 6 | 11.721 | 8.369 | 5.742 | 6.459 | 32.5 | 1.0000 | 0.4924 | -0.0152 | -0.0152 | 1.0000 | 1.0000 | No | No | No |
| **ISG/stress** | 2 | *ISG15* | % positive | 11 | 6 | 4.970 | 5.074 | 5.213 | 4.830 | 33. | 1.0000 | 0.5000 | 0.0000 | 0.0000 | 1.0000 | 1.0000 | No | No | No |
| **ISG/stress** | 2 | *JUN* | % positive | 11 | 6 | 9.003 | 7.821 | 3.659 | 5.593 | 33. | 1.0000 | 0.5000 | 0.0000 | 0.0000 | 1.0000 | 1.0000 | No | No | No |
| **Memory-like** | 2 | *TNFRSF13B* | Mean expression | 11 | 6 | 0.015 | 0.015 | 0.016 | 0.015 | 33. | 1.0000 | 0.5000 | 0.0000 | 0.0000 | 1.0000 | 1.0000 | No | No | No |

Mean expression denotes the patient-level mean of log-normalized expression, including zero values. "% positive" denotes the percentage of cells with at least one raw UMI for the corresponding gene. Signed AUC was calculated as 2 × AUC − 1, with positive values indicating higher values in pCR and negative values indicating higher values in non-pCR. APC, antigen-presenting cell; AUC, area under the receiver operating characteristic curve; FDR, false discovery rate; ISG, interferon-stimulated gene; n, number of patients; pCR, pathological complete response; non-pCR, absence of pCR; q, Benjamini–Hochberg-adjusted P value.

**Supplementary Table 8. Longitudinal changes in circulating immune populations in the complete flow cytometry cohort.**

| Panel | Immune population | Metric | Interval | Paired patients, n | Median at earlier time point (%) | Median at later time point (%) | Median paired change (percentage points) | Q1 paired change | Q3 paired change | Matched rank-biserial r | Wilcoxon P value | FDR-adjusted q value | Evaluation status |
|---|---|---|---|---|---|---|---|---|---|---|---|---|---|
| Panel 1 | Total CD3$^{+}$ T cells | Population frequency (%) | T1→T2 | 28 | 75.300 | 72.900 | -1.600 | -8.675 | 4.350 | -0.233 | 0.290431 | 0.457437 | Evaluable |
| Panel 1 | Total CD3$^{+}$ T cells | Population frequency (%) | T2→T3 | 23 | 78.100 | 75.600 | -1.300 | -4.950 | 3.800 | -0.123 | 0.614811 | 0.983698 | Evaluable |
| Panel 1 | Total CD3$^{+}$ T cells | Population frequency (%) | T1→T3 | 19 | 74.200 | 75.300 | 0.600 | -13.900 | 6.550 | -0.158 | 0.546047 | 0.863854 | Evaluable |
| Panel 1 | CD4$^{+}$ T cells | Population frequency (%) | T1→T2 | 27 | 59.900 | 55.800 | -3.100 | -13.400 | 2.650 | -0.437 | 0.047464 | 0.151886 | Evaluable |
| Panel 1 | CD4$^{+}$ T cells | Population frequency (%) | T2→T3 | 23 | 46.500 | 48.500 | 0.100 | -8.200 | 9.800 | -0.022 | 0.940555 | 0.988101 | Evaluable |
| Panel 1 | CD4$^{+}$ T cells | Population frequency (%) | T1→T3 | 18 | 57.200 | 52.750 | -4.750 | -9.725 | 0.725 | -0.368 | 0.181465 | 0.580688 | Evaluable |
| Panel 1 | CD4$^{+}$CCR7$^{+}$ T cells | Population frequency (%) | T1→T2 | 27 | 55.900 | 46.100 | -9.200 | -31.250 | 15.050 | -0.254 | 0.258371 | 0.457437 | Evaluable |
| Panel 1 | CD4$^{+}$CCR7$^{+}$ T cells | Population frequency (%) | T2→T3 | 23 | 63.200 | 52.300 | -9.100 | -37.250 | 13.100 | -0.217 | 0.376522 | 0.890498 | Evaluable |
| Panel 1 | CD4$^{+}$CCR7$^{+}$ T cells | Population frequency (%) | T1→T3 | 18 | 63.100 | 48.700 | -16.750 | -37.950 | 5.400 | -0.544 | 0.043167 | 0.230225 | Evaluable |
| Panel 1 | CD4$^{+}$CD161$^{+}$ T cells | Population frequency (%) | T1→T2 | 27 | 8.650 | 9.300 | -0.300 | -4.490 | 3.100 | -0.190 | 0.399743 | 0.532990 | Evaluable |
| Panel 1 | CD4$^{+}$CD161$^{+}$ T cells | Population frequency (%) | T2→T3 | 23 | 10.800 | 9.670 | 0.140 | -3.405 | 6.830 | 0.188 | 0.445200 | 0.890498 | Evaluable |
| Panel 1 | CD4$^{+}$CD161$^{+}$ T cells | Population frequency (%) | T1→T3 | 18 | 10.405 | 8.755 | 0.415 | -3.515 | 3.160 | 0.029 | 0.932281 | 0.945745 | Evaluable |
| Panel 1 | CD4$^{+}$TIM-3$^{+}$ T cells | Population frequency (%) | T1→T2 | 27 | 1.490 | 7.220 | 2.100 | 0.070 | 13.540 | 0.519 | 0.017386 | 0.092724 | Evaluable |
| Panel 1 | CD4$^{+}$TIM-3$^{+}$ T cells | Population frequency (%) | T2→T3 | 23 | 7.760 | 2.030 | 0.980 | -8.480 | 7.530 | -0.007 | 0.988101 | 0.988101 | Evaluable |
| Panel 1 | CD4$^{+}$TIM-3$^{+}$ T cells | Population frequency (%) | T1→T3 | 18 | 1.795 | 1.985 | 0.550 | -5.793 | 2.255 | -0.111 | 0.701881 | 0.863854 | Evaluable |
| Panel 1 | CD4$^{+}$TIGIT$^{+}$ T cells | Population frequency (%) | T1→T2 | 27 | 16.700 | 14.200 | -1.900 | -9.500 | 7.040 | -0.117 | 0.602605 | 0.654547 | Evaluable |
| Panel 1 | CD4$^{+}$TIGIT$^{+}$ T cells | Population frequency (%) | T2→T3 | 23 | 12.900 | 16.500 | 6.220 | -20.850 | 18.660 | 0.029 | 0.916847 | 0.988101 | Evaluable |
| Panel 1 | CD4$^{+}$TIGIT$^{+}$ T cells | Population frequency (%) | T1→T3 | 18 | 19.400 | 17.850 | -2.550 | -13.050 | 9.013 | -0.111 | 0.701881 | 0.863854 | Evaluable |
| Panel 1 | CD4$^{+}$CD127$^{+}$CD27$^{+}$ T cells | Population frequency (%) | T1→T2 | 27 | 68.600 | 63.700 | -5.300 | -13.450 | 1.450 | -0.577 | 0.007586 | 0.060686 | Evaluable |
| Panel 1 | CD4$^{+}$CD127$^{+}$CD27$^{+}$ T cells | Population frequency (%) | T2→T3 | 23 | 69.500 | 68.500 | 1.000 | -7.750 | 9.400 | 0.087 | 0.715111 | 0.988101 | Evaluable |
| Panel 1 | CD4$^{+}$CD127$^{+}$CD27$^{+}$ T cells | Population frequency (%) | T1→T3 | 18 | 71.000 | 66.250 | -2.600 | -7.000 | 0.675 | -0.203 | 0.463107 | 0.863854 | Evaluable |
| Panel 1 | CD4$^{+}$CD127$^{+}$CD27$^{+}$CD62L$^{+}$CCR7$^{+}$ T cells | Population frequency (%) | T1→T2 | 29 | 0.620 | 0.450 | -0.260 | -0.812 | 0.390 | -0.218 | 0.314488 | 0.457437 | Evaluable |
| Panel 1 | CD4$^{+}$CD127$^{+}$CD27$^{+}$CD62L$^{+}$CCR7$^{+}$ T cells | Population frequency (%) | T2→T3 | 23 | 0.640 | 0.420 | -0.040 | -1.730 | 0.665 | -0.174 | 0.481952 | 0.890498 | Evaluable |
| Panel 1 | CD4$^{+}$CD127$^{+}$CD27$^{+}$CD62L$^{+}$CCR7$^{+}$ T cells | Population frequency (%) | T1→T3 | 21 | 0.620 | 0.650 | -0.020 | -1.150 | 0.690 | -0.022 | 0.945745 | 0.945745 | Evaluable |
| Panel 1 | CD8$^{+}$ T cells | Population frequency (%) | T1→T2 | 27 | 27.900 | 27.700 | 0.800 | -2.900 | 4.500 | 0.233 | 0.290431 | 0.457437 | Evaluable |
| Panel 1 | CD8$^{+}$ T cells | Population frequency (%) | T2→T3 | 23 | 24.800 | 27.900 | 2.500 | -4.050 | 5.950 | 0.254 | 0.300133 | 0.890498 | Evaluable |
| Panel 1 | CD8$^{+}$ T cells | Population frequency (%) | T1→T3 | 18 | 27.800 | 34.400 | 4.750 | -0.800 | 12.150 | 0.567 | 0.034233 | 0.230225 | Evaluable |
| Panel 1 | CD8$^{+}$CCR7$^{+}$ T cells | Population frequency (%) | T1→T2 | 28 | 33.900 | 22.000 | -9.045 | -21.620 | 7.525 | -0.448 | 0.037930 | 0.151720 | Evaluable |
| Panel 1 | CD8$^{+}$CCR7$^{+}$ T cells | Population frequency (%) | T2→T3 | 23 | 35.500 | 34.400 | -1.100 | -41.815 | 14.150 | -0.188 | 0.445200 | 0.890498 | Evaluable |
| Panel 1 | CD8$^{+}$CCR7$^{+}$ T cells | Population frequency (%) | T1→T3 | 19 | 41.700 | 27.500 | -11.330 | -25.450 | -2.900 | -0.600 | 0.020412 | 0.230225 | Evaluable |
| Panel 1 | CD8$^{+}$CD161$^{+}$ T cells | Population frequency (%) | T1→T2 | 28 | 6.130 | 5.910 | -1.150 | -5.143 | 1.088 | -0.281 | 0.201568 | 0.457437 | Evaluable |
| Panel 1 | CD8$^{+}$CD161$^{+}$ T cells | Population frequency (%) | T2→T3 | 23 | 5.430 | 7.010 | 1.090 | -2.345 | 5.985 | 0.268 | 0.272583 | 0.890498 | Evaluable |
| Panel 1 | CD8$^{+}$CD161$^{+}$ T cells | Population frequency (%) | T1→T3 | 19 | 4.990 | 7.010 | 0.810 | -0.985 | 3.290 | 0.158 | 0.567760 | 0.863854 | Evaluable |
| Panel 1 | CD8$^{+}$TIM-3$^{+}$ T cells | Population frequency (%) | T1→T2 | 28 | 2.390 | 2.265 | -0.270 | -2.225 | 3.590 | -0.133 | 0.551874 | 0.654547 | Evaluable |
| Panel 1 | CD8$^{+}$TIM-3$^{+}$ T cells | Population frequency (%) | T2→T3 | 23 | 3.620 | 1.810 | -0.050 | -8.360 | 5.620 | -0.065 | 0.799773 | 0.988101 | Evaluable |
| Panel 1 | CD8$^{+}$TIM-3$^{+}$ T cells | Population frequency (%) | T1→T3 | 19 | 2.130 | 1.430 | 0.180 | -13.535 | 0.720 | -0.137 | 0.622562 | 0.863854 | Evaluable |
| Panel 1 | CD8$^{+}$TIGIT$^{+}$ T cells | Population frequency (%) | T1→T2 | 28 | 30.000 | 25.700 | -3.700 | -18.798 | 4.868 | -0.404 | 0.061857 | 0.164953 | Evaluable |
| Panel 1 | CD8$^{+}$TIGIT$^{+}$ T cells | Population frequency (%) | T2→T3 | 23 | 30.100 | 34.100 | 5.200 | -16.300 | 13.900 | 0.029 | 0.916847 | 0.988101 | Evaluable |
| Panel 1 | CD8$^{+}$TIGIT$^{+}$ T cells | Population frequency (%) | T1→T3 | 19 | 30.200 | 37.900 | -4.300 | -18.650 | 16.800 | -0.063 | 0.828785 | 0.945745 | Evaluable |
| Panel 1 | CD8$^{+}$CD127$^{+}$CD27$^{+}$ T cells | Population frequency (%) | T1→T2 | 28 | 44.900 | 35.950 | -8.200 | -16.850 | -2.000 | -0.576 | 0.006549 | 0.060686 | Evaluable |
| Panel 1 | CD8$^{+}$CD127$^{+}$CD27$^{+}$ T cells | Population frequency (%) | T2→T3 | 23 | 45.600 | 45.100 | -2.800 | -7.850 | 14.900 | 0.167 | 0.500905 | 0.890498 | Evaluable |
| Panel 1 | CD8$^{+}$CD127$^{+}$CD27$^{+}$ T cells | Population frequency (%) | T1→T3 | 19 | 46.200 | 37.900 | -3.500 | -14.900 | 7.500 | -0.263 | 0.332066 | 0.863854 | Evaluable |
| Panel 1 | CD8$^{+}$CD127$^{+}$CD27$^{+}$CD62L$^{+}$CCR7$^{+}$ T cells | Population frequency (%) | T1→T2 | 27 | 0.540 | 0.450 | -0.088 | -0.480 | 0.730 | 0.053 | 0.822338 | 0.822338 | Evaluable |
| Panel 1 | CD8$^{+}$CD127$^{+}$CD27$^{+}$CD62L$^{+}$CCR7$^{+}$ T cells | Population frequency (%) | T2→T3 | 23 | 0.550 | 0.350 | -0.420 | -1.720 | 0.225 | -0.442 | 0.065036 | 0.598953 | Evaluable |
| Panel 1 | CD8$^{+}$CD127$^{+}$CD27$^{+}$CD62L$^{+}$CCR7$^{+}$ T cells | Population frequency (%) | T1→T3 | 18 | 0.605 | 0.555 | -0.014 | -0.678 | 0.320 | -0.135 | 0.639694 | 0.863854 | Evaluable |
| Panel 1 | CD8$^{+}$CD62L$^{+}$CCR7$^{+}$ T cells | Population frequency (%) | T1→T2 | 28 | 0.430 | 0.285 | -0.081 | -1.028 | 0.540 | -0.113 | 0.613638 | 0.654547 | Evaluable |
| Panel 1 | CD8$^{+}$CD62L$^{+}$CCR7$^{+}$ T cells | Population frequency (%) | T2→T3 | 23 | 0.640 | 0.390 | -0.380 | -2.625 | 0.149 | -0.428 | 0.074869 | 0.598953 | Evaluable |
| Panel 1 | CD8$^{+}$CD62L$^{+}$CCR7$^{+}$ T cells | Population frequency (%) | T1→T3 | 19 | 0.950 | 0.390 | -0.320 | -2.800 | 0.200 | -0.450 | 0.093603 | 0.374413 | Evaluable |
| Panel 1 | CD8$^{+}$CD62L$^{+}$CCR7$^{+}$CD127$^{+}$ T cells | Population frequency (%) | T1→T2 | 0 | | | | | | | | | Not evaluable: <5 pairs |
| Panel 1 | CD8$^{+}$CD62L$^{+}$CCR7$^{+}$CD127$^{+}$ T cells | Population frequency (%) | T2→T3 | 3 | 0.510 | 0.580 | 0.070 | -2.430 | 0.255 | | | | Not evaluable: <5 pairs |
| Panel 1 | CD8$^{+}$CD62L$^{+}$CCR7$^{+}$CD127$^{+}$ T cells | Population frequency (%) | T1→T3 | 2 | 1.105 | 0.631 | -0.475 | -0.892 | -0.057 | | | | Not evaluable: <5 pairs |
| Panel 2 | Total CD3$^{+}$ T cells | Population frequency (%) | T1→T2 | 27 | 70.800 | 59.900 | -8.400 | -15.350 | 0.050 | -0.661 | 0.001859 | 0.007899 | Evaluable |
| Panel 2 | Total CD3$^{+}$ T cells | Population frequency (%) | T2→T3 | 23 | 60.800 | 54.500 | -9.100 | -33.350 | 13.150 | -0.225 | 0.360391 | 0.524205 | Evaluable |
| Panel 2 | Total CD3$^{+}$ T cells | Population frequency (%) | T1→T3 | 19 | 69.200 | 60.900 | -6.200 | -37.050 | 4.850 | -0.432 | 0.104156 | 0.151500 | Evaluable |
| Panel 2 | NK cells | Population frequency (%) | T1→T2 | 27 | 3.260 | 2.950 | -0.580 | -2.090 | 1.405 | -0.212 | 0.348328 | 0.422969 | Evaluable |
| Panel 2 | NK cells | Population frequency (%) | T2→T3 | 23 | 1.770 | 3.100 | 0.430 | -1.280 | 3.185 | 0.312 | 0.200155 | 0.355832 | Evaluable |
| Panel 2 | NK cells | Population frequency (%) | T1→T3 | 19 | 2.180 | 3.100 | 0.840 | -0.375 | 1.455 | 0.442 | 0.095512 | 0.151500 | Evaluable |
| Panel 2 | Total B cells | Population frequency (%) | T1→T2 | 27 | 5.530 | 5.430 | -1.060 | -2.885 | 0.735 | -0.381 | 0.085933 | 0.162319 | Evaluable |
| Panel 2 | Total B cells | Population frequency (%) | T2→T3 | 23 | 3.270 | 0.770 | -1.540 | -4.575 | 0.025 | -0.688 | 0.002726 | 0.006232 | Evaluable |
| Panel 2 | Total B cells | Population frequency (%) | T1→T3 | 19 | 5.600 | 0.980 | -4.750 | -5.665 | -1.355 | -0.821 | 0.000790 | 0.005343 | Evaluable |
| Panel 2 | CD20$^{+}$CD38$^{+}$ mature B cells | Population frequency (%) | T1→T2 | 27 | 32.700 | 37.800 | 1.800 | -3.200 | 10.300 | 0.268 | 0.232575 | 0.329481 | Evaluable |
| Panel 2 | CD20$^{+}$CD38$^{+}$ mature B cells | Population frequency (%) | T2→T3 | 23 | 33.300 | 7.450 | -16.750 | -33.610 | -4.125 | -0.790 | 0.000408 | 0.002176 | Evaluable |
| Panel 2 | CD20$^{+}$CD38$^{+}$ mature B cells | Population frequency (%) | T1→T3 | 19 | 28.400 | 13.200 | -10.990 | -20.995 | -1.445 | -0.853 | 0.000420 | 0.005343 | Evaluable |
| Panel 2 | CD20$^{+}$CD38$^{-}$ memory B cells | Population frequency (%) | T1→T2 | 27 | 35.200 | 27.000 | -4.600 | -16.000 | 2.300 | -0.439 | 0.046263 | 0.098308 | Evaluable |
| Panel 2 | CD20$^{+}$CD38$^{-}$ memory B cells | Population frequency (%) | T2→T3 | 23 | 25.800 | 9.550 | -8.650 | -14.119 | -0.070 | -0.601 | 0.010109 | 0.020218 | Evaluable |
| Panel 2 | CD20$^{+}$CD38$^{-}$ memory B cells | Population frequency (%) | T1→T3 | 19 | 28.900 | 10.800 | -8.550 | -24.250 | 0.140 | -0.758 | 0.002399 | 0.006487 | Evaluable |
| Panel 2 | CD20$^{-}$CD38$^{+}$ plasmablasts | Population frequency (%) | T1→T2 | 27 | 15.300 | 19.800 | 1.510 | -9.400 | 12.620 | 0.138 | 0.546018 | 0.580144 | Evaluable |
| Panel 2 | CD20$^{-}$CD38$^{+}$ plasmablasts | Population frequency (%) | T2→T3 | 23 | 23.600 | 56.100 | 28.800 | 15.400 | 45.265 | 0.790 | 0.000408 | 0.002176 | Evaluable |
| Panel 2 | CD20$^{-}$CD38$^{+}$ plasmablasts | Population frequency (%) | T1→T3 | 19 | 22.400 | 56.600 | 30.300 | 11.550 | 45.205 | 0.747 | 0.002838 | 0.006487 | Evaluable |
| Panel 2 | Switched-memory B cells (IgD$^{-}$CD27$^{+}$) | Population frequency (%) | T1→T2 | 27 | 31.700 | 22.500 | -3.900 | -9.050 | -1.650 | -0.762 | 0.000236 | 0.002009 | Evaluable |

| | | | | | | | | | | | | | |
|---|---|---|---|---|---|---|---|---|---|---|---|---|---|
| Panel 2 | Switched-memory B cells (IgD⁻CD27⁺) | Population frequency (%) | T2→T3 | 23 | 21.800 | 23.900 | 0.400 | -10.965 | 17.550 | 0.116 | 0.643390 | 0.686283 | Evaluable |
| Panel 2 | Switched-memory B cells (IgD⁻CD27⁺) | Population frequency (%) | T1→T3 | 19 | 33.600 | 25.400 | -4.140 | -11.500 | 4.800 | -0.211 | 0.441334 | 0.470756 | Evaluable |
| Panel 2 | CD38⁺ switched-memory B cells | Population frequency (%) | T1→T2 | 24 | 44.400 | 46.600 | 1.650 | -3.100 | 8.500 | 0.160 | 0.492871 | | Evaluable |
| Panel 2 | CD38⁺ switched-memory B cells | Population frequency (%) | T2→T3 | 16 | 50.950 | 54.000 | 6.550 | -9.550 | 15.025 | 0.250 | 0.403748 | | Evaluable |
| Panel 2 | CD38⁺ switched-memory B cells | Population frequency (%) | T1→T3 | 12 | 50.900 | 52.100 | 5.300 | -6.225 | 14.525 | 0.385 | 0.266113 | | Evaluable |
| Panel 2 | CD38⁻ switched-memory B cells | Population frequency (%) | T1→T2 | 24 | 51.600 | 47.500 | -1.500 | -7.275 | 3.125 | -0.193 | 0.407323 | | Evaluable |
| Panel 2 | CD38⁻ switched-memory B cells | Population frequency (%) | T2→T3 | 16 | 43.450 | 41.850 | -5.550 | -12.950 | 8.808 | -0.221 | 0.463745 | | Evaluable |
| Panel 2 | CD38⁻ switched-memory B cells | Population frequency (%) | T1→T3 | 12 | 43.900 | 42.100 | -2.550 | -14.250 | 4.075 | -0.282 | 0.423828 | | Evaluable |
| Panel 2 | CD83⁺HLA-DRhi APC-like B cells | Population frequency (%) | T1→T2 | 27 | 23.100 | 27.400 | 6.100 | 2.480 | 14.580 | 0.519 | 0.017386 | 0.042223 | Evaluable |
| Panel 2 | CD83⁺HLA-DRhi APC-like B cells | Population frequency (%) | T2→T3 | 23 | 27.400 | 21.700 | -4.400 | -33.650 | 12.750 | -0.225 | 0.360391 | 0.524205 | Evaluable |
| Panel 2 | CD83⁺HLA-DRhi APC-like B cells | Population frequency (%) | T1→T3 | 19 | 16.600 | 27.200 | 11.760 | -3.050 | 24.340 | 0.474 | 0.070128 | 0.124672 | Evaluable |
| Panel 2 | Naive B cells (IgD⁺CD27⁻) | Population frequency (%) | T1→T2 | 27 | 26.800 | 29.300 | -0.900 | -13.800 | 11.000 | -0.037 | 0.866436 | 0.866436 | Evaluable |
| Panel 2 | Naive B cells (IgD⁺CD27⁻) | Population frequency (%) | T2→T3 | 23 | 28.000 | 0.910 | -24.820 | -35.353 | -6.510 | -0.920 | 0.000013 | 0.000210 | Evaluable |
| Panel 2 | Naive B cells (IgD⁺CD27⁻) | Population frequency (%) | T1→T3 | 19 | 16.800 | 3.340 | -12.820 | -27.655 | 0.008 | -0.825 | 0.002138 | 0.006487 | Evaluable |
| Panel 2 | Double-negative B cells (IgD⁻CD27⁻) | Population frequency (%) | T1→T2 | 27 | 19.000 | 26.800 | 4.600 | -4.870 | 16.500 | 0.323 | 0.148152 | 0.251859 | Evaluable |
| Panel 2 | Double-negative B cells (IgD⁻CD27⁻) | Population frequency (%) | T2→T3 | 23 | 34.800 | 62.300 | 25.200 | 4.000 | 49.700 | 0.768 | 0.000639 | 0.002555 | Evaluable |
| Panel 2 | Double-negative B cells (IgD⁻CD27⁻) | Population frequency (%) | T1→T3 | 19 | 26.900 | 62.300 | 20.000 | 6.550 | 41.150 | 0.747 | 0.002838 | 0.006487 | Evaluable |
| Panel 2 | CD38⁻ double-negative B cells | Population frequency (%) | T1→T2 | 27 | 25.600 | 23.400 | -0.400 | -14.600 | 8.850 | -0.245 | 0.274764 | | Evaluable |
| Panel 2 | CD38⁻ double-negative B cells | Population frequency (%) | T2→T3 | 22 | 26.900 | 9.765 | -14.200 | -21.115 | -3.588 | -0.739 | 0.001455 | | Evaluable |
| Panel 2 | CD38⁻ double-negative B cells | Population frequency (%) | T1→T3 | 18 | 20.400 | 9.285 | -11.720 | -24.438 | -5.898 | -0.731 | 0.004745 | | Evaluable |
| Panel 2 | CD38⁺ double-negative B cells | Population frequency (%) | T1→T2 | 27 | 70.900 | 75.500 | 1.600 | -9.800 | 14.450 | 0.228 | 0.312570 | | Evaluable |
| Panel 2 | CD38⁺ double-negative B cells | Population frequency (%) | T2→T3 | 22 | 69.950 | 88.350 | 15.350 | 3.950 | 22.600 | 0.739 | 0.002399 | | Evaluable |
| Panel 2 | CD38⁺ double-negative B cells | Population frequency (%) | T1→T3 | 18 | 76.100 | 88.900 | 12.900 | 8.650 | 25.075 | 0.708 | 0.006577 | | Evaluable |
| Panel 2 | CD38⁻ naive B cells | Population frequency (%) | T1→T2 | 21 | 18.300 | 15.600 | -4.820 | -8.400 | 0.050 | -0.636 | 0.009016 | | Evaluable |
| Panel 2 | CD38⁻ naive B cells | Population frequency (%) | T2→T3 | 5 | 8.800 | 7.810 | -0.990 | -2.560 | 5.670 | 0.067 | 1.000000 | | Evaluable |
| Panel 2 | CD38⁻ naive B cells | Population frequency (%) | T1→T3 | 5 | 21.500 | 13.700 | -0.940 | -7.800 | 1.100 | -0.333 | 0.625000 | | Evaluable |
| Panel 2 | CD38⁺ naive B cells | Population frequency (%) | T1→T2 | 21 | 76.900 | 81.200 | 6.000 | 0.400 | 11.100 | 0.680 | 0.004879 | | Evaluable |
| Panel 2 | CD38⁺ naive B cells | Population frequency (%) | T2→T3 | 5 | 89.000 | 86.700 | -2.300 | -6.800 | 2.600 | -0.067 | 1.000000 | | Evaluable |
| Panel 2 | CD38⁺ naive B cells | Population frequency (%) | T1→T3 | 5 | 72.300 | 83.100 | 1.900 | 0.400 | 10.800 | 0.733 | 0.187500 | | Evaluable |
| Panel 2 | Unswitched-memory B cells (IgD⁺CD27⁺) | Population frequency (%) | T1→T2 | 27 | 5.550 | 2.870 | -2.560 | -6.165 | 0.235 | -0.735 | 0.000427 | 0.002421 | Evaluable |
| Panel 2 | Unswitched-memory B cells (IgD⁺CD27⁺) | Population frequency (%) | T2→T3 | 23 | 1.610 | 0.600 | -1.150 | -2.775 | 0.000 | -0.721 | 0.002474 | 0.006232 | Evaluable |
| Panel 2 | Unswitched-memory B cells (IgD⁺CD27⁺) | Population frequency (%) | T1→T3 | 19 | 3.400 | 0.300 | -2.110 | -6.680 | -0.551 | -0.908 | 0.001002 | 0.005343 | Evaluable |
| Panel 2 | CD38⁻ unswitched-memory B cells | Population frequency (%) | T1→T2 | 19 | 62.400 | 54.000 | -8.800 | -16.800 | -5.500 | -0.832 | 0.000645 | | Evaluable |
| Panel 2 | CD38⁻ unswitched-memory B cells | Population frequency (%) | T2→T3 | 1 | 54.300 | 60.200 | 5.900 | 5.900 | 5.900 | | | | Not evaluable: <5 pairs |
| Panel 2 | CD38⁻ unswitched-memory B cells | Population frequency (%) | T1→T3 | 2 | 54.400 | 52.600 | -1.800 | -5.250 | 1.650 | | | | Not evaluable: <5 pairs |
| Panel 2 | CD38⁺ unswitched-memory B cells | Population frequency (%) | T1→T2 | 19 | 28.800 | 41.000 | 10.000 | 5.750 | 19.200 | 0.905 | 0.000126 | | Evaluable |
| Panel 2 | CD38⁺ unswitched-memory B cells | Population frequency (%) | T2→T3 | 1 | 41.500 | 35.200 | -6.300 | -6.300 | -6.300 | | | | Not evaluable: <5 pairs |
| Panel 2 | CD38⁺ unswitched-memory B cells | Population frequency (%) | T1→T3 | 2 | 40.500 | 42.100 | 1.600 | -2.050 | 5.250 | | | | Not evaluable: <5 pairs |
| Panel 2 | Mature dendritic cells (HLA-DR⁺CD83⁺) | Population frequency (%) | T1→T2 | 27 | 5.060 | 7.780 | 1.400 | -1.825 | 4.345 | 0.302 | 0.177520 | 0.274349 | Evaluable |
| Panel 2 | Mature dendritic cells (HLA-DR⁺CD83⁺) | Population frequency (%) | T2→T3 | 23 | 8.560 | 9.170 | 1.300 | -2.970 | 6.515 | 0.130 | 0.601046 | 0.686283 | Evaluable |
| Panel 2 | Mature dendritic cells (HLA-DR⁺CD83⁺) | Population frequency (%) | T1→T3 | 19 | 4.180 | 10.000 | 1.890 | -0.820 | 11.285 | 0.379 | 0.156403 | 0.208537 | Evaluable |
| Panel 2 | Immature dendritic cells (HLA-DR⁺CD83⁻) | Population frequency (%) | T1→T2 | 27 | 13.400 | 10.100 | -3.700 | -6.730 | 0.815 | -0.534 | 0.015241 | 0.042223 | Evaluable |
| Panel 2 | Immature dendritic cells (HLA-DR⁺CD83⁻) | Population frequency (%) | T2→T3 | 23 | 9.300 | 11.300 | 2.160 | -3.155 | 6.570 | 0.196 | 0.427418 | 0.533166 | Evaluable |
| Panel 2 | Immature dendritic cells (HLA-DR⁺CD83⁻) | Population frequency (%) | T1→T3 | 19 | 11.200 | 14.000 | 0.500 | -2.910 | 4.680 | 0.147 | 0.594887 | 0.594887 | Evaluable |
| Panel 2 | CD38 balance in switched-memory B cells | CD38 compositional balance (%) | T1→T2 | 24 | 46.253 | 49.385 | 1.983 | -3.154 | 8.340 | 0.193 | 0.422341 | 0.478654 | Evaluable |
| Panel 2 | CD38 balance in switched-memory B cells | CD38 compositional balance (%) | T2→T3 | 16 | 54.007 | 56.736 | 5.925 | -9.092 | 14.576 | 0.235 | 0.433197 | 0.533166 | Evaluable |
| Panel 2 | CD38 balance in switched-memory B cells | CD38 compositional balance (%) | T1→T3 | 12 | 54.173 | 55.304 | 3.914 | -5.427 | 14.806 | 0.282 | 0.423828 | 0.470756 | Evaluable |
| Panel 2 | CD38 balance in double-negative B cells | CD38 compositional balance (%) | T1→T2 | 27 | 73.472 | 76.340 | 0.656 | -9.250 | 15.068 | 0.238 | 0.290081 | 0.379337 | Evaluable |
| Panel 2 | CD38 balance in double-negative B cells | CD38 compositional balance (%) | T2→T3 | 22 | 71.954 | 90.027 | 15.232 | 3.631 | 21.661 | 0.747 | 0.001262 | 0.004037 | Evaluable |
| Panel 2 | CD38 balance in double-negative B cells | CD38 compositional balance (%) | T1→T3 | 18 | 78.965 | 90.543 | 11.971 | 6.422 | 25.826 | 0.719 | 0.005600 | 0.011200 | Evaluable |
| Panel 2 | CD38 balance in naive B cells | CD38 compositional balance (%) | T1→T2 | 21 | 80.938 | 83.817 | 5.008 | -0.010 | 9.508 | 0.654 | 0.007101 | 0.024144 | Evaluable |
| Panel 2 | CD38 balance in naive B cells | CD38 compositional balance (%) | T2→T3 | 5 | 91.002 | 91.736 | 0.734 | -5.953 | 2.560 | 0.067 | 1.000000 | 1.000000 | Evaluable |
| Panel 2 | CD38 balance in naive B cells | CD38 compositional balance (%) | T1→T3 | 5 | 77.079 | 85.847 | 0.725 | -0.418 | 8.768 | 0.467 | 0.437500 | 0.470756 | Evaluable |
| Panel 2 | CD38 balance in unswitched-memory B cells | CD38 compositional balance (%) | T1→T2 | 19 | 30.947 | 43.319 | 11.086 | 5.976 | 18.960 | 0.905 | 0.000126 | 0.002009 | Evaluable |
| Panel 2 | CD38 balance in unswitched-memory B cells | CD38 compositional balance (%) | T2→T3 | 1 | 43.319 | 36.897 | -6.422 | -6.422 | -6.422 | | | | Not evaluable: <5 pairs |
| Panel 2 | CD38 balance in unswitched-memory B cells | CD38 compositional balance (%) | T1→T3 | 2 | 42.724 | 44.512 | 1.788 | -1.953 | 5.530 | | | | Not evaluable: <5 pairs |

Complementary CD38-positive and CD38-negative branches were excluded from FDR correction; their P values are nominal. Blank q values indicate either an unevaluable comparison or an endpoint outside the correction family. APC, antigen-presenting cell; FDR, false discovery rate; n, number of paired patients; NK, natural killer; Q1, first quartile; Q3, third quartile; r, matched rank-biserial correlation; q, Benjamini–Hochberg-adjusted P value; T1, baseline before treatment; T2, after completion of the first treatment block; T3, after completion of neoadjuvant treatment and before surgery.

**Supplementary Table 9. Cross-sectional flow cytometry comparisons of circulating immune populations according to pathological response.**

| Time point | Biological time | Panel | Immune population | Metric | pCR patients, n | non-pCR patients, n | Median pCR (%) | Q1 pCR | Q3 pCR | Median non-pCR (%) | Q1 non-pCR | Q3 non-pCR | Median difference (pCR − non-pCR) | Cliff's delta | Cliff's delta 95% CI, lower | Cliff's delta 95% CI, upper | AUC (pCR higher) | Mann–Whitney P value | Panel-specific FDR q value | Included in FDR family | Evaluation status |
|---|---|---|---|---|---|---|---|---|---|---|---|---|---|---|---|---|---|---|---|---|---|
| T1 | Baseline | Panel 1 | Total CD3$^{+}$ T cells | Population frequency (%) | 30 | 14 | 76.100 | 67.875 | 82.550 | 74.200 | 66.750 | 78.850 | 1.900 | 0.164 | -0.205 | 0.505 | 0.582 | 0.391583 | 0.837146 | Yes | Evaluable |
| T1 | Baseline | Panel 1 | CD4$^{+}$ T cells | Population frequency (%) | 30 | 13 | 58.950 | 52.650 | 62.900 | 62.300 | 48.500 | 66.500 | -3.350 | -0.205 | -0.564 | 0.180 | 0.397 | 0.300376 | 0.837146 | Yes | Evaluable |
| T1 | Baseline | Panel 1 | CD4$^{+}$CCR7$^{+}$ T cells | Population frequency (%) | 30 | 13 | 59.150 | 40.325 | 69.700 | 49.000 | 26.500 | 68.600 | 10.150 | 0.210 | -0.200 | 0.590 | 0.605 | 0.288188 | 0.837146 | Yes | Evaluable |
| T1 | Baseline | Panel 1 | CD4$^{+}$CD161$^{+}$ T cells | Population frequency (%) | 30 | 13 | 8.435 | 3.633 | 13.925 | 7.310 | 5.160 | 12.100 | 1.125 | -0.036 | -0.359 | 0.323 | 0.482 | 0.863515 | 0.907656 | Yes | Evaluable |
| T1 | Baseline | Panel 1 | CD4$^{+}$PD-1$^{+}$ T cells | Population frequency (%) | 30 | 13 | 11.750 | 4.558 | 17.250 | 8.850 | 7.090 | 9.900 | 2.900 | 0.231 | -0.098 | 0.554 | 0.615 | 0.239269 | 0.837146 | Yes | Evaluable |
| T1 | Baseline | Panel 1 | CD4$^{+}$TIM-3$^{+}$ T cells | Population frequency (%) | 30 | 13 | 3.595 | 0.383 | 12.525 | 1.840 | 0.620 | 4.310 | 1.755 | 0.179 | -0.174 | 0.497 | 0.590 | 0.366252 | 0.837146 | Yes | Evaluable |
| T1 | Baseline | Panel 1 | CD4$^{+}$TIGIT$^{+}$ T cells | Population frequency (%) | 30 | 13 | 18.950 | 10.700 | 31.625 | 11.100 | 7.460 | 16.700 | 7.850 | 0.390 | 0.046 | 0.677 | 0.695 | 0.045867 | 0.837146 | Yes | Evaluable |
| T1 | Baseline | Panel 1 | CD4$^{+}$PD-1$^{+}$TIM-3$^{+}$TIGIT$^{+}$ T cells | Population frequency (%) | 26 | 13 | 1.475 | 0.383 | 7.160 | 2.320 | 0.770 | 6.440 | -0.845 | -0.080 | -0.438 | 0.302 | 0.460 | 0.698521 | 0.907656 | Yes | Evaluable |
| T1 | Baseline | Panel 1 | CD4$^{+}$PD-1$^{+}$TIM-3$^{-}$TIGIT$^{+}$ T cells | Population frequency (%) | 26 | 13 | 41.000 | 25.975 | 54.300 | 41.900 | 31.800 | 48.100 | -0.900 | -0.036 | -0.426 | 0.314 | 0.482 | 0.869837 | 0.907656 | Yes | Evaluable |
| T1 | Baseline | Panel 1 | CD4$^{+}$PD-1$^{+}$TIM-3$^{-}$TIGIT$^{-}$ T cells | Population frequency (%) | 26 | 13 | 48.250 | 35.025 | 68.350 | 48.400 | 43.600 | 61.500 | -0.150 | -0.047 | -0.408 | 0.314 | 0.476 | 0.823183 | 0.907656 | Yes | Evaluable |
| T1 | Baseline | Panel 1 | CD4$^{+}$CD127$^{+}$CD27$^{+}$ T cells | Population frequency (%) | 30 | 13 | 69.300 | 56.475 | 74.150 | 66.700 | 54.700 | 81.600 | 2.600 | -0.090 | -0.505 | 0.323 | 0.455 | 0.653008 | 0.907656 | Yes | Evaluable |
| T1 | Baseline | Panel 1 | CD4$^{+}$CD127$^{+}$CD27$^{+}$CD62L$^{+}$CCR7$^{+}$ T cells | Population frequency (%) | 30 | 16 | 0.625 | 0.255 | 1.658 | 0.770 | 0.363 | 1.500 | -0.145 | -0.044 | -0.388 | 0.323 | 0.478 | 0.817545 | 0.907656 | Yes | Evaluable |
| T1 | Baseline | Panel 1 | CD8$^{+}$ T cells | Population frequency (%) | 30 | 13 | 27.850 | 19.175 | 32.850 | 27.800 | 19.900 | 29.800 | 0.050 | -0.003 | -0.395 | 0.377 | 0.499 | 1.000000 | 1.000000 | Yes | Evaluable |
| T1 | Baseline | Panel 1 | CD8$^{+}$CCR7$^{+}$ T cells | Population frequency (%) | 30 | 14 | 29.800 | 19.050 | 56.075 | 27.700 | 11.975 | 48.925 | 2.100 | 0.195 | -0.195 | 0.548 | 0.598 | 0.307455 | 0.837146 | Yes | Evaluable |
| T1 | Baseline | Panel 1 | CD8$^{+}$CD161$^{+}$ T cells | Population frequency (%) | 30 | 14 | 7.085 | 3.403 | 11.450 | 5.315 | 4.540 | 10.685 | 1.770 | -0.040 | -0.369 | 0.319 | 0.480 | 0.840233 | 0.907656 | Yes | Evaluable |
| T1 | Baseline | Panel 1 | CD8$^{+}$PD-1$^{+}$ T cells | Population frequency (%) | 30 | 14 | 21.700 | 11.003 | 29.525 | 18.500 | 12.525 | 26.075 | 3.200 | 0.133 | -0.219 | 0.476 | 0.567 | 0.488320 | 0.837146 | Yes | Evaluable |
| T1 | Baseline | Panel 1 | CD8$^{+}$TIM-3$^{+}$ T cells | Population frequency (%) | 30 | 14 | 3.195 | 0.583 | 10.095 | 2.065 | 1.083 | 3.008 | 1.130 | 0.133 | -0.233 | 0.472 | 0.567 | 0.488335 | 0.837146 | Yes | Evaluable |
| T1 | Baseline | Panel 1 | CD8$^{+}$TIGIT$^{+}$ T cells | Population frequency (%) | 30 | 14 | 30.050 | 24.300 | 53.350 | 28.600 | 14.825 | 31.425 | 1.450 | 0.295 | -0.048 | 0.605 | 0.648 | 0.121211 | 0.837146 | Yes | Evaluable |
| T1 | Baseline | Panel 1 | CD8$^{+}$PD-1$^{+}$TIM-3$^{+}$TIGIT$^{+}$ T cells | Population frequency (%) | 27 | 13 | 1.080 | 0.325 | 5.660 | 1.760 | 1.330 | 3.690 | -0.680 | -0.140 | -0.476 | 0.214 | 0.430 | 0.488266 | 0.837146 | Yes | Evaluable |
| T1 | Baseline | Panel 1 | CD8$^{+}$PD-1$^{+}$TIM-3$^{-}$TIGIT$^{+}$ T cells | Population frequency (%) | 27 | 13 | 55.000 | 46.600 | 68.600 | 43.300 | 36.500 | 62.700 | 11.700 | 0.236 | -0.143 | 0.587 | 0.618 | 0.236372 | 0.837146 | Yes | Evaluable |
| T1 | Baseline | Panel 1 | CD8$^{+}$PD-1$^{+}$TIM-3$^{-}$TIGIT$^{-}$ T cells | Population frequency (%) | 27 | 13 | 31.900 | 15.650 | 42.350 | 38.600 | 26.000 | 52.600 | -6.700 | -0.265 | -0.624 | 0.123 | 0.368 | 0.184052 | 0.837146 | Yes | Evaluable |
| T1 | Baseline | Panel 1 | CD8$^{+}$CD127$^{+}$CD27$^{+}$ T cells | Population frequency (%) | 30 | 14 | 44.750 | 35.025 | 57.025 | 49.100 | 36.150 | 64.025 | -4.350 | -0.105 | -0.471 | 0.276 | 0.448 | 0.587978 | 0.907656 | Yes | Evaluable |
| T1 | Baseline | Panel 1 | CD8$^{+}$CD127$^{+}$CD27$^{+}$CD62L$^{+}$CCR7$^{+}$ T cells | Population frequency (%) | 30 | 13 | 0.510 | 0.165 | 1.300 | 0.680 | 0.440 | 1.550 | -0.170 | -0.144 | -0.479 | 0.205 | 0.428 | 0.466958 | 0.837146 | Yes | Evaluable |
| T1 | Baseline | Panel 1 | CD8$^{+}$CD62L$^{+}$CCR7$^{+}$ T cells | Population frequency (%) | 30 | 14 | 0.355 | 0.163 | 1.443 | 0.795 | 0.260 | 1.375 | -0.440 | -0.093 | -0.443 | 0.255 | 0.454 | 0.632077 | 0.907656 | Yes | Evaluable |
| T1 | Baseline | Panel 1 | CD8$^{+}$CD62L$^{+}$CCR7$^{+}$CD127$^{+}$ T cells | Population frequency (%) | 2 | 2 | | | | | | | | | | | | | | | Not evaluable: <5 samples in one group |
| T1 | Baseline | Panel 2 | Total CD3$^{+}$ T cells | Population frequency (%) | 29 | 16 | 71.300 | 62.400 | 74.900 | 69.700 | 64.550 | 71.800 | 1.600 | 0.164 | -0.172 | 0.487 | 0.582 | 0.373897 | 0.838145 | Yes | Evaluable |
| T1 | Baseline | Panel 2 | NK cells | Population frequency (%) | 29 | 16 | 2.130 | 1.040 | 3.580 | 2.785 | 1.905 | 4.478 | -0.655 | -0.248 | -0.556 | 0.095 | 0.376 | 0.176495 | 0.835446 | Yes | Evaluable |
| T1 | Baseline | Panel 2 | Total B cells | Population frequency (%) | 29 | 16 | 5.190 | 3.740 | 7.080 | 5.565 | 3.180 | 6.623 | -0.375 | -0.009 | -0.358 | 0.358 | 0.496 | 0.971627 | 0.971627 | Yes | Evaluable |
| T1 | Baseline | Panel 2 | CD20$^{+}$CD38$^{+}$ mature B cells | Population frequency (%) | 29 | 16 | 29.300 | 20.100 | 41.900 | 35.850 | 27.825 | 43.800 | -6.550 | -0.194 | -0.517 | 0.142 | 0.403 | 0.294863 | 0.835446 | Yes | Evaluable |
| T1 | Baseline | Panel 2 | CD20$^{+}$CD38$^{-}$ memory B cells | Population frequency (%) | 29 | 16 | 31.600 | 23.100 | 38.600 | 37.400 | 26.925 | 44.575 | -5.800 | -0.203 | -0.548 | 0.159 | 0.399 | 0.273511 | 0.835446 | Yes | Evaluable |
| T1 | Baseline | Panel 2 | CD20$^{-}$CD38$^{+}$ plasmablasts | Population frequency (%) | 29 | 16 | 16.000 | 11.800 | 30.800 | 13.800 | 8.395 | 30.575 | 2.200 | 0.125 | -0.228 | 0.470 | 0.563 | 0.503344 | 0.838145 | Yes | Evaluable |
| T1 | Baseline | Panel 2 | Switched-memory B cells (IgD$^{-}$CD27$^{+}$) | Population frequency (%) | 29 | 16 | 25.600 | 21.400 | 37.700 | 36.150 | 26.650 | 38.700 | -10.550 | -0.226 | -0.556 | 0.134 | 0.387 | 0.217553 | 0.835446 | Yes | Evaluable |
| T1 | Baseline | Panel 2 | CD38$^{+}$ switched-memory B cells | Population frequency (%) | 25 | 15 | 44.400 | 35.100 | 52.900 | 46.700 | 41.150 | 50.600 | -2.300 | -0.088 | -0.440 | 0.256 | 0.456 | 0.654833 | | No | Evaluable |
| T1 | Baseline | Panel 2 | CD38$^{-}$ switched-memory B cells | Population frequency (%) | 25 | 15 | 52.300 | 43.200 | 60.000 | 48.300 | 43.950 | 54.350 | 4.000 | 0.067 | -0.293 | 0.448 | 0.533 | 0.737392 | | No | Evaluable |
| T1 | Baseline | Panel 2 | CD83$^{+}$HLA-DRhi APC-like B cells | Population frequency (%) | 29 | 16 | 24.900 | 12.300 | 49.600 | 20.700 | 7.848 | 40.825 | 4.200 | 0.125 | -0.246 | 0.466 | 0.563 | 0.503344 | 0.838145 | Yes | Evaluable |
| T1 | Baseline | Panel 2 | Naive B cells (IgD$^{+}$CD27$^{-}$) | Population frequency (%) | 29 | 16 | 27.000 | 14.200 | 39.400 | 27.700 | 15.375 | 35.475 | -0.700 | 0.024 | -0.341 | 0.388 | 0.512 | 0.905624 | 0.962226 | Yes | Evaluable |
| T1 | Baseline | Panel 2 | Double-negative B cells (IgD$^{-}$CD27$^{-}$) | Population frequency (%) | 29 | 16 | 20.500 | 16.500 | 29.800 | 19.400 | 15.525 | 34.350 | 1.100 | 0.071 | -0.313 | 0.418 | 0.536 | 0.704398 | 0.838145 | Yes | Evaluable |
| T1 | Baseline | Panel 2 | CD38$^{-}$ double-negative B cells | Population frequency (%) | 29 | 16 | 25.500 | 14.400 | 36.500 | 30.700 | 19.250 | 42.925 | -5.200 | -0.211 | -0.552 | 0.159 | 0.394 | 0.250132 | | No | Evaluable |
| T1 | Baseline | Panel 2 | CD38$^{+}$ double-negative B cells | Population frequency (%) | 29 | 16 | 71.600 | 58.100 | 82.800 | 65.250 | 53.925 | 76.475 | 6.350 | 0.216 | -0.134 | 0.573 | 0.608 | 0.240483 | | No | Evaluable |
| T1 | Baseline | Panel 2 | CD38$^{-}$ naive B cells | Population frequency (%) | 25 | 14 | 18.300 | 13.100 | 22.900 | 20.600 | 13.625 | 25.050 | -2.300 | -0.077 | -0.457 | 0.314 | 0.461 | 0.703485 | | No | Evaluable |
| T1 | Baseline | Panel 2 | CD38$^{+}$ naive B cells | Population frequency (%) | 25 | 14 | 77.700 | 69.300 | 83.800 | 73.800 | 67.950 | 84.275 | 3.900 | 0.046 | -0.354 | 0.420 | 0.523 | 0.826166 | | No | Evaluable |
| T1 | Baseline | Panel 2 | Unswitched-memory B cells (IgD$^{+}$CD27$^{+}$) | Population frequency (%) | 29 | 16 | 4.630 | 2.290 | 7.300 | 5.200 | 3.365 | 7.400 | -0.570 | -0.078 | -0.422 | 0.246 | 0.461 | 0.678142 | 0.838145 | Yes | Evaluable |
| T1 | Baseline | Panel 2 | CD38$^{-}$ unswitched-memory B cells | Population frequency (%) | 22 | 13 | 62.750 | 58.850 | 68.650 | 58.300 | 56.100 | 63.400 | 4.450 | 0.325 | -0.084 | 0.703 | 0.663 | 0.116293 | | No | Evaluable |
| T1 | Baseline | Panel 2 | CD38$^{+}$ unswitched-memory B cells | Population frequency (%) | 22 | 13 | 27.600 | 25.200 | 32.250 | 33.200 | 26.300 | 40.100 | -5.600 | -0.266 | -0.664 | 0.119 | 0.367 | 0.200433 | | No | Evaluable |
| T1 | Baseline | Panel 2 | Mature dendritic cells (HLA-DR$^{+}$CD83$^{+}$) | Population frequency (%) | 29 | 16 | 5.060 | 1.880 | 11.700 | 4.675 | 1.560 | 9.468 | 0.385 | 0.091 | -0.265 | 0.422 | 0.545 | 0.626889 | 0.838145 | Yes | Evaluable |
| T1 | Baseline | Panel 2 | Immature dendritic cells (HLA-DR$^{+}$CD83$^{-}$) | Population frequency (%) | 29 | 16 | 11.200 | 6.800 | 17.800 | 11.750 | 9.490 | 15.650 | -0.550 | -0.114 | -0.446 | 0.233 | 0.443 | 0.537558 | 0.838145 | Yes | Evaluable |
| T1 | Baseline | Panel 2 | CD38 balance in switched-memory B cells | CD38 compositional balance (%) | 25 | 15 | 45.915 | 36.715 | 55.047 | 49.003 | 43.085 | 54.002 | -3.088 | -0.077 | -0.435 | 0.285 | 0.461 | 0.699116 | 0.838145 | Yes | Evaluable |
| T1 | Baseline | Panel 2 | CD38 balance in double-negative B cells | CD38 compositional balance (%) | 29 | 16 | 73.738 | 61.677 | 85.469 | 67.988 | 55.639 | 79.906 | 5.751 | 0.211 | -0.151 | 0.556 | 0.606 | 0.253217 | 0.835446 | Yes | Evaluable |
| T1 | Baseline | Panel 2 | CD38 balance in naive B cells | CD38 compositional balance (%) | 25 | 14 | 80.938 | 75.560 | 86.550 | 78.083 | 72.643 | 86.082 | 2.854 | 0.069 | -0.314 | 0.451 | 0.534 | 0.739540 | 0.838145 | Yes | Evaluable |
| T1 | Baseline | Panel 2 | CD38 balance in unswitched-memory B cells | CD38 compositional balance (%) | 22 | 13 | 30.326 | 27.160 | 34.666 | 35.141 | 28.281 | 41.641 | -4.815 | -0.252 | -0.629 | 0.182 | 0.374 | 0.229010 | 0.835446 | Yes | Evaluable |
| T2 | Intermediate | Panel 1 | Total CD3$^{+}$ T cells | Population frequency (%) | 21 | 17 | 73.900 | 67.400 | 82.700 | 76.700 | 70.000 | 90.300 | -2.800 | -0.140 | -0.490 | 0.238 | 0.430 | 0.471928 | 0.968154 | Yes | Evaluable |
| T2 | Intermediate | Panel 1 | CD4$^{+}$ T cells | Population frequency (%) | 21 | 17 | 48.900 | 39.700 | 65.400 | 50.900 | 45.400 | 62.700 | -2.000 | -0.050 | -0.434 | 0.317 | 0.475 | 0.802921 | 0.968154 | Yes | Evaluable |
| T2 | Intermediate | Panel 1 | CD4$^{+}$CCR7$^{+}$ T cells | Population frequency (%) | 21 | 17 | 50.400 | 37.400 | 67.700 | 68.600 | 29.100 | 86.300 | -18.200 | -0.120 | -0.524 | 0.283 | 0.440 | 0.537533 | 0.968154 | Yes | Evaluable |
| T2 | Intermediate | Panel 1 | CD4$^{+}$CD161$^{+}$ T cells | Population frequency (%) | 21 | 17 | 10.400 | 6.470 | 12.700 | 7.250 | 4.650 | 11.000 | 3.150 | 0.311 | -0.059 | 0.636 | 0.655 | 0.106341 | 0.968154 | Yes | Evaluable |
| T2 | Intermediate | Panel 1 | CD4$^{+}$TIM-3$^{+}$ T cells | Population frequency (%) | 21 | 17 | 7.220 | 1.620 | 15.700 | 8.200 | 1.750 | 18.100 | -0.980 | -0.098 | -0.451 | 0.300 | 0.451 | 0.617682 | 0.968154 | Yes | Evaluable |
| T2 | Intermediate | Panel 1 | CD4$^{+}$TIGIT$^{+}$ T cells | Population frequency (%) | 21 | 17 | 14.900 | 9.040 | 23.100 | 12.600 | 6.800 | 56.100 | 2.300 | 0.025 | -0.373 | 0.412 | 0.513 | 0.907645 | 0.968154 | Yes | Evaluable |
| T2 | Intermediate | Panel 1 | CD4$^{+}$CD127$^{+}$CD27$^{+}$ T cells | Population frequency (%) | 21 | 17 | 68.400 | 54.800 | 71.700 | 65.800 | 57.200 | 72.100 | 2.600 | 0.039 | -0.328 | 0.418 | 0.520 | 0.848637 | 0.968154 | Yes | Evaluable |
| T2 | Intermediate | Panel 1 | CD4$^{+}$CD127$^{+}$CD27$^{+}$CD62L$^{+}$CCR7$^{+}$ T cells | Population frequency (%) | 21 | 17 | 0.550 | 0.140 | 2.690 | 0.450 | 0.150 | 1.430 | 0.100 | 0.092 | -0.272 | 0.474 | 0.546 | 0.638496 | 0.968154 | Yes | Evaluable |
| T2 | Intermediate | Panel 1 | CD8$^{+}$ T cells | Population frequency (%) | 21 | 17 | 25.200 | 21.600 | 34.700 | 25.800 | 15.400 | 30.800 | -0.600 | 0.252 | -0.129 | 0.605 | 0.626 | 0.191360 | 0.968154 | Yes | Evaluable |
| T2 | Intermediate | Panel 1 | CD8$^{+}$CCR7$^{+}$ T cells | Population frequency (%) | 21 | 17 | 30.300 | 19.000 | 46.300 | 35.200 | 14.600 | 88.000 | -4.900 | -0.104 | -0.485 | 0.305 | 0.448 | 0.597173 | 0.968154 | Yes | Evaluable |
| T2 | Intermediate | Panel 1 | CD8$^{+}$CD161$^{+}$ T cells | Population frequency (%) | 21 | 17 | 5.620 | 3.050 | 8.490 | 5.570 | 2.830 | 8.240 | 0.050 | 0.048 | -0.328 | 0.395 | 0.524 | 0.814305 | 0.968154 | Yes | Evaluable |
| T2 | Intermediate | Panel 1 | CD8$^{+}$TIM-3$^{+}$ T cells | Population frequency (%) | 21 | 17 | 2.220 | 0.530 | 5.850 | 3.620 | 1.040 | 11.100 | -1.400 | -0.171 | -0.524 | 0.221 | 0.415 | 0.382824 | 0.968154 | Yes | Evaluable |
| T2 | Intermediate | Panel 1 | CD8$^{+}$TIGIT$^{+}$ T cells | Population frequency (%) | 21 | 17 | 29.400 | 16.600 | 38.200 | 28.900 | 9.210 | 37.400 | 0.500 | 0.053 | -0.311 | 0.429 | 0.527 | 0.794031 | 0.968154 | Yes | Evaluable |
| T2 | Intermediate | Panel 1 | CD8$^{+}$CD127$^{+}$CD27$^{+}$ T cells | Population frequency (%) | 21 | 17 | 37.400 | 22.100 | 51.400 | 39.000 | 31.900 | 55.600 | -1.600 | -0.165 | -0.513 | 0.221 | 0.417 | 0.398980 | 0.968154 | Yes | Evaluable |
| T2 | Intermediate | Panel 1 | CD8$^{+}$CD127$^{+}$CD27$^{+}$CD62L$^{+}$CCR7$^{+}$ T cells | Population frequency (%) | 21 | 17 | 0.500 | 0.180 | 2.480 | 0.270 | 0.140 | 1.870 | 0.230 | 0.070 | -0.289 | 0.434 | 0.535 | 0.724560 | 0.968154 | Yes | Evaluable |
| T2 | Intermediate | Panel 1 | CD8$^{+}$CD62L$^{+}$CCR7$^{+}$ T cells | Population frequency (%) | 21 | 17 | 0.290 | 0.140 | 2.220 | 0.590 | 0.140 | 1.540 | -0.300 | -0.006 | -0.395 | 0.401 | 0.497 | 0.988287 | 0.988287 | Yes | Evaluable |
| T2 | Intermediate | Panel 1 | CD8$^{+}$CD62L$^{+}$CCR7$^{+}$CD127$^{+}$ T cells | Population frequency (%) | 6 | 3 | | | | | | | | | | | | | | | Not evaluable: <5 samples in one group |
| T2 | Intermediate | Panel 2 | Total CD3$^{+}$ T cells | Population frequency (%) | 21 | 18 | 59.700 | 50.200 | 67.400 | 62.850 | 58.550 | 67.450 | -3.150 | -0.183 | -0.545 | 0.172 | 0.409 | 0.338094 | 1.000000 | Yes | Evaluable |
| T2 | Intermediate | Panel 2 | NK cells | Population frequency (%) | 21 | 18 | 2.960 | 1.450 | 3.960 | 1.675 | 0.675 | 3.025 | 1.285 | 0.212 | -0.153 | 0.572 | 0.606 | 0.269175 | 1.000000 | Yes | Evaluable |
| T2 | Intermediate | Panel 2 | Total B cells | Population frequency (%) | 21 | 18 | 5.430 | 2.250 | 6.460 | 3.210 | 1.080 | 5.853 | 2.220 | 0.167 | -0.206 | 0.513 | 0.583 | 0.382461 | 1.000000 | Yes | Evaluable |
| T2 | Intermediate | Panel 2 | CD20$^{+}$CD38$^{+}$ mature B cells | Population frequency (%) | 21 | 18 | 35.000 | 19.300 | 43.400 | 31.000 | 13.925 | 47.800 | 4.000 | 0.005 | -0.370 | 0.376 | 0.503 | 0.988894 | 1.000000 | Yes | Evaluable |
| T2 | Intermediate | Panel 2 | CD20$^{+}$CD38$^{-}$ memory B cells | Population frequency (%) | 21 | 18 | 26.300 | 14.900 | 33.900 | 26.600 | 14.375 | 37.025 | -0.300 | -0.045 | -0.421 | 0.328 | 0.478 | 0.821679 | 1.000000 | Yes | Evaluable |
| T2 | Intermediate | Panel 2 | CD20$^{-}$CD38$^{+}$ plasmablasts | Population frequency (%) | 21 | 18 | 24.900 | 16.200 | 35.200 | 19.950 | 16.500 | 29.175 | 4.950 | 0.116 | -0.259 | 0.487 | 0.558 | 0.548901 | 1.000000 | Yes | Evaluable |
| T2 | Intermediate | Panel 2 | Switched-memory B cells (IgD$^{-}$CD27$^{+}$) | Population frequency (%) | 21 | 18 | 22.500 | 14.000 | 32.900 | 22.600 | 16.750 | 32.825 | -0.100 | -0.016 | -0.386 | 0.355 | 0.492 | 0.943849 | 1.000000 | Yes | Evaluable |
| T2 | Intermediate | Panel 2 | CD38$^{+}$ switched-memory B cells | Population frequency (%) | 21 | 17 | 49.100 | 44.500 | 56.700 | 43.500 | 36.000 | 57.500 | 5.600 | 0.129 | -0.238 | 0.513 | 0.564 | 0.508877 | | No | Evaluable |
| T2 | Intermediate | Panel 2 | CD38$^{-}$ switched-memory B cells | Population frequency (%) | 21 | 17 | 46.700 | 38.900 | 51.500 | 52.100 | 37.300 | 60.500 | -5.400 | -0.137 | -0.524 | 0.238 | 0.431 | 0.485560 | | No | Evaluable |
| T2 | Intermediate | Panel 2 | CD83$^{+}$HLA-DRhi APC-like B cells | Population frequency (%) | 21 | 18 | 26.100 | 18.400 | 47.800 | 32.550 | 19.450 | 55.150 | -6.450 | -0.103 | -0.460 | 0.249 | 0.448 | 0.592448 | 1.000000 | Yes | Evaluable |
| T2 | Intermediate | Panel 2 | Naive B cells (IgD$^{+}$CD27$^{-}$) | Population frequency (%) | 21 | 18 | 29.500 | 13.600 | 38.500 | 23.150 | 12.000 | 38.575 | 6.350 | 0.037 | -0.317 | 0.434 | 0.519 | 0.856379 | 1.000000 | Yes | Evaluable |
| T2 | Intermediate | Panel 2 | Double-negative B cells (IgD$^{-}$CD27$^{-}$) | Population frequency (%) | 21 | 18 | 29.000 | 22.200 | 41.600 | 31.500 | 17.300 | 42.100 | -2.500 | 0.000 | -0.360 | 0.376 | 0.500 | 1.000000 | 1.000000 | Yes | Evaluable |
| T2 | Intermediate | Panel 2 | CD38$^{-}$ double-negative B cells | Population frequency (%) | 21 | 18 | 26.400 | 13.600 | 28.000 | 24.450 | 12.750 | 37.800 | 1.950 | -0.153 | -0.519 | 0.249 | 0.423 | 0.421988 | | No | Evaluable |
| T2 | Intermediate | Panel 2 | CD38$^{+}$ double-negative B cells | Population frequency (%) | 21 | 18 | 70.200 | 67.200 | 84.100 | 71.450 | 58.350 | 84.500 | -1.250 | 0.185 | -0.228 | 0.587 | 0.593 | 0.331062 | | No | Evaluable |
| T2 | Intermediate | Panel 2 | CD38$^{-}$ naive B cells | Population frequency (%) | 19 | 16 | 10.400 | 8.540 | 18.450 | 16.950 | 7.273 | 21.625 | -6.550 | -0.135 | -0.533 | 0.283 | 0.433 | 0.507741 | | No | Evaluable |
| T2 | Intermediate | Panel 2 | CD38$^{+}$ naive B cells | Population frequency (%) | 19 | 16 | 87.100 | 77.300 | 89.750 | 80.000 | 70.250 | 91.200 | 7.100 | 0.128 | -0.309 | 0.539 | 0.564 | 0.529191 | | No | Evaluable |
| T2 | Intermediate | Panel 2 | Unswitched-memory B cells (IgD$^{+}$CD27$^{+}$) | Population frequency (%) | 21 | 18 | 2.690 | 1.450 | 3.930 | 2.535 | 1.173 | 3.815 | 0.155 | 0.079 | -0.302 | 0.439 | 0.540 | 0.686255 | 1.000000 | Yes | Evaluable |
| T2 | Intermediate | Panel 2 | CD38$^{-}$ unswitched-memory B cells | Population frequency (%) | 15 | 11 | 53.700 | 40.200 | 62.150 | 48.000 | 31.700 | 54.450 | 5.700 | 0.309 | -0.164 | 0.709 | 0.655 | 0.198164 | | No | Evaluable |
| T2 | Intermediate | Panel 2 | CD38$^{+}$ unswitched-memory B cells | Population frequency (%) | 15 | 11 | 40.700 | 31.750 | 54.100 | 48.000 | 40.450 | 63.050 | -7.300 | -0.333 | -0.721 | 0.103 | 0.333 | 0.163875 | | No | Evaluable |
| T2 | Intermediate | Panel 2 | Mature dendritic cells (HLA-DR$^{+}$CD83$^{+}$) | Population frequency (%) | 21 | 18 | 9.560 | 5.470 | 12.400 | 8.520 | 6.478 | 13.425 | 1.040 | -0.119 | -0.484 | 0.254 | 0.440 | 0.535382 | 1.000000 | Yes | Evaluable |
| T2 | Intermediate | Panel 2 | Immature dendritic cells (HLA-DR$^{+}$CD83$^{-}$) | Population frequency (%) | 21 | 18 | 10.100 | 7.780 | 12.700 | 9.580 | 3.800 | 14.600 | 0.520 | 0.071 | -0.312 | 0.442 | 0.536 | 0.714149 | 1.000000 | Yes | Evaluable |
| T2 | Intermediate | Panel 2 | CD38 balance in switched-memory B cells | CD38 compositional balance (%) | 21 | 17 | 51.253 | 46.354 | 59.310 | 45.502 | 37.306 | 60.654 | 5.751 | 0.132 | -0.261 | 0.518 | 0.566 | 0.503992 | 1.000000 | Yes | Evaluable |
| T2 | Intermediate | Panel 2 | CD38 balance in double-negative B cells | CD38 compositional balance (%) | 21 | 18 | 72.471 | 70.984 | 86.080 | 74.429 | 60.687 | 86.889 | -1.958 | 0.153 | -0.254 | 0.524 | 0.577 | 0.426373 | 1.000000 | Yes | Evaluable |
| T2 | Intermediate | Panel 2 | CD38 balance in naive B cells | CD38 compositional balance (%) | 19 | 16 | 89.278 | 80.726 | 91.312 | 82.559 | 76.372 | 92.616 | 6.719 | 0.132 | -0.283 | 0.553 | 0.566 | 0.523243 | 1.000000 | Yes | Evaluable |
| T2 | Intermediate | Panel 2 | CD38 balance in unswitched-memory B cells | CD38 compositional balance (%) | 15 | 11 | 43.905 | 33.373 | 57.371 | 50.000 | 42.690 | 66.525 | -6.095 | -0.309 | -0.709 | 0.164 | 0.345 | 0.198164 | 1.000000 | Yes | Evaluable |
| T3 | Pre-surgery | Panel 1 | Total CD3$^{+}$ T cells | Population frequency (%) | 18 | 11 | 75.500 | 70.900 | 82.925 | 76.100 | 71.500 | 95.050 | -0.600 | -0.146 | -0.616 | 0.303 | 0.427 | 0.529133 | 0.751263 | Yes | Evaluable |
| T3 | Pre-surgery | Panel 1 | CD4$^{+}$ T cells | Population frequency (%) | 18 | 11 | 49.550 | 43.000 | 56.775 | 47.500 | 26.850 | 54.600 | 2.050 | 0.278 | -0.182 | 0.692 | 0.639 | 0.224802 | 0.751263 | Yes | Evaluable |
| T3 | Pre-surgery | Panel 1 | CD4$^{+}$CCR7$^{+}$ T cells | Population frequency (%) | 18 | 11 | 51.450 | 44.050 | 72.150 | 55.000 | 42.500 | 72.550 | -3.550 | -0.020 | -0.424 | 0.404 | 0.490 | 0.947159 | 0.964145 | Yes | Evaluable |
| T3 | Pre-surgery | Panel 1 | CD4$^{+}$CD161$^{+}$ T cells | Population frequency (%) | 18 | 11 | 9.935 | 6.963 | 14.550 | 9.430 | 5.005 | 16.900 | 0.505 | -0.020 | -0.485 | 0.424 | 0.490 | 0.947159 | 0.964145 | Yes | Evaluable |
| T3 | Pre-surgery | Panel 1 | CD4$^{+}$TIM-3$^{+}$ T cells | Population frequency (%) | 18 | 11 | 1.985 | 0.883 | 24.700 | 9.700 | 2.160 | 20.650 | -7.715 | -0.152 | -0.616 | 0.323 | 0.424 | 0.520798 | 0.751263 | Yes | Evaluable |
| T3 | Pre-surgery | Panel 1 | CD4$^{+}$TIGIT$^{+}$ T cells | Population frequency (%) | 18 | 11 | 20.500 | 11.675 | 36.625 | 17.000 | 13.250 | 29.750 | 3.500 | 0.015 | -0.424 | 0.470 | 0.508 | 0.964145 | 0.964145 | Yes | Evaluable |
| T3 | Pre-surgery | Panel 1 | CD4$^{+}$CD127$^{+}$CD27$^{+}$ T cells | Population frequency (%) | 18 | 11 | 63.800 | 51.300 | 70.725 | 68.500 | 54.000 | 75.250 | -4.700 | -0.293 | -0.707 | 0.152 | 0.354 | 0.203814 | 0.751263 | Yes | Evaluable |
| T3 | Pre-surgery | Panel 1 | CD4$^{+}$CD127$^{+}$CD27$^{+}$CD62L$^{+}$CCR7$^{+}$ T cells | Population frequency (%) | 18 | 11 | 0.445 | 0.265 | 1.035 | 0.900 | 0.320 | 1.855 | -0.455 | -0.217 | -0.642 | 0.263 | 0.391 | 0.345053 | 0.751263 | Yes | Evaluable |
| T3 | Pre-surgery | Panel 1 | CD8$^{+}$ T cells | Population frequency (%) | 18 | 11 | 28.050 | 20.725 | 37.550 | 27.900 | 14.200 | 35.700 | 0.150 | 0.232 | -0.263 | 0.677 | 0.616 | 0.311813 | 0.751263 | Yes | Evaluable |
| T3 | Pre-surgery | Panel 1 | CD8$^{+}$CCR7$^{+}$ T cells | Population frequency (%) | 18 | 11 | 32.750 | 22.825 | 42.475 | 42.400 | 24.500 | 57.950 | -9.650 | -0.116 | -0.566 | 0.323 | 0.442 | 0.620971 | 0.812039 | Yes | Evaluable |
| T3 | Pre-surgery | Panel 1 | CD8$^{+}$CD161$^{+}$ T cells | Population frequency (%) | 18 | 11 | 6.460 | 4.468 | 11.195 | 7.950 | 6.790 | 11.235 | -1.490 | -0.283 | -0.646 | 0.162 | 0.359 | 0.220433 | 0.751263 | Yes | Evaluable |
| T3 | Pre-surgery | Panel 1 | CD8$^{+}$TIM-3$^{+}$ T cells | Population frequency (%) | 18 | 11 | 1.015 | 0.515 | 4.093 | 7.640 | 2.120 | 22.750 | -6.625 | -0.384 | -0.798 | 0.071 | 0.308 | 0.092197 | 0.751263 | Yes | Evaluable |
| T3 | Pre-surgery | Panel 1 | CD8$^{+}$TIGIT$^{+}$ T cells | Population frequency (%) | 18 | 11 | 36.000 | 23.350 | 45.775 | 36.400 | 22.000 | 43.150 | -0.400 | 0.030 | -0.394 | 0.465 | 0.515 | 0.912045 | 0.964145 | Yes | Evaluable |
| T3 | Pre-surgery | Panel 1 | CD8$^{+}$CD127$^{+}$CD27$^{+}$ T cells | Population frequency (%) | 18 | 11 | 37.700 | 32.025 | 52.175 | 47.200 | 37.800 | 57.650 | -9.500 | -0.152 | -0.586 | 0.354 | 0.424 | 0.520798 | 0.751263 | Yes | Evaluable |
| T3 | Pre-surgery | Panel 1 | CD8$^{+}$CD127$^{+}$CD27$^{+}$CD62L$^{+}$CCR7$^{+}$ T cells | Population frequency (%) | 18 | 10 | 0.440 | 0.185 | 0.755 | 0.820 | 0.198 | 1.245 | -0.380 | -0.189 | -0.656 | 0.289 | 0.406 | 0.428751 | 0.751263 | Yes | Evaluable |
| T3 | Pre-surgery | Panel 1 | CD8$^{+}$CD62L$^{+}$CCR7$^{+}$ T cells | Population frequency (%) | 18 | 11 | 0.425 | 0.185 | 0.730 | 0.500 | 0.290 | 1.940 | -0.075 | -0.197 | -0.637 | 0.263 | 0.402 | 0.392996 | 0.751263 | Yes | Evaluable |

| | | | | | | | | | | | | | | | | | | | | | |
|---|---|---|---|---|---|---|---|---|---|---|---|---|---|---|---|---|---|---|---|---|---|
| T3 | Pre-surgery | Panel 1 | CD8$^{+}$CD62L$^{+}$CCR7$^{+}$CD127$^{+}$ T cells | Population frequency (%) | 7 | 5 | 0.220 | 0.140 | 0.665 | 0.720 | 0.320 | 1.190 | -0.500 | -0.257 | -0.886 | 0.429 | 0.371 | 0.530303 | 0.751263 | Yes | Evaluable |
| T3 | Pre-surgery | Panel 2 | Total CD3$^{+}$ T cells | Population frequency (%) | 18 | 11 | 57.700 | 37.925 | 73.625 | 63.100 | 26.850 | 69.500 | -5.400 | 0.141 | -0.313 | 0.556 | 0.571 | 0.550075 | 0.773487 | Yes | Evaluable |
| T3 | Pre-surgery | Panel 2 | NK cells | Population frequency (%) | 18 | 11 | 2.840 | 1.978 | 3.923 | 3.300 | 1.470 | 4.970 | -0.460 | -0.015 | -0.475 | 0.424 | 0.492 | 0.964145 | 0.964145 | Yes | Evaluable |
| T3 | Pre-surgery | Panel 2 | Total B cells | Population frequency (%) | 18 | 11 | 0.930 | 0.468 | 2.045 | 0.670 | 0.410 | 2.615 | 0.260 | 0.081 | -0.374 | 0.535 | 0.540 | 0.736010 | 0.841155 | Yes | Evaluable |
| T3 | Pre-surgery | Panel 2 | CD20$^{+}$CD38$^{+}$ mature B cells | Population frequency (%) | 18 | 11 | 6.475 | 2.240 | 15.350 | 20.500 | 6.605 | 33.200 | -14.025 | -0.444 | -0.808 | 0.000 | 0.278 | 0.049392 | 0.131712 | Yes | Evaluable |
| T3 | Pre-surgery | Panel 2 | CD20$^{+}$CD38$^{-}$ memory B cells | Population frequency (%) | 18 | 11 | 8.460 | 1.888 | 15.825 | 30.200 | 10.950 | 35.250 | -21.740 | -0.596 | -0.919 | -0.182 | 0.202 | 0.006965 | 0.066656 | Yes | Evaluable |
| T3 | Pre-surgery | Panel 2 | CD20$^{-}$CD38$^{+}$ plasmablasts | Population frequency (%) | 18 | 11 | 69.050 | 55.075 | 82.750 | 41.700 | 28.050 | 56.750 | 27.350 | 0.556 | 0.131 | 0.899 | 0.778 | 0.012498 | 0.066656 | Yes | Evaluable |
| T3 | Pre-surgery | Panel 2 | Switched-memory B cells (IgD$^{-}$CD27$^{+}$) | Population frequency (%) | 18 | 11 | 22.000 | 10.875 | 43.000 | 28.100 | 9.930 | 39.500 | -6.100 | -0.061 | -0.515 | 0.414 | 0.470 | 0.807945 | 0.861808 | Yes | Evaluable |
| T3 | Pre-surgery | Panel 2 | CD38$^{+}$ switched-memory B cells | Population frequency (%) | 14 | 9 | 61.550 | 53.850 | 77.400 | 47.400 | 36.000 | 55.000 | 14.150 | 0.643 | 0.254 | 0.945 | 0.821 | 0.011723 | | No | Evaluable |
| T3 | Pre-surgery | Panel 2 | CD38$^{-}$ switched-memory B cells | Population frequency (%) | 14 | 9 | 34.600 | 19.250 | 41.375 | 47.400 | 42.300 | 59.900 | -12.800 | -0.667 | -0.952 | -0.270 | 0.167 | 0.006887 | | No | Evaluable |
| T3 | Pre-surgery | Panel 2 | CD83$^{+}$HLA-DRhi APC-like B cells | Population frequency (%) | 18 | 11 | 20.650 | 14.775 | 34.400 | 29.000 | 23.550 | 40.100 | -8.350 | -0.192 | -0.596 | 0.253 | 0.404 | 0.411953 | 0.659125 | Yes | Evaluable |
| T3 | Pre-surgery | Panel 2 | Naive B cells (IgD$^{+}$CD27$^{-}$) | Population frequency (%) | 18 | 11 | 1.335 | 0.373 | 4.840 | 4.580 | 1.985 | 10.815 | -3.245 | -0.333 | -0.727 | 0.096 | 0.333 | 0.143981 | 0.287962 | Yes | Evaluable |
| T3 | Pre-surgery | Panel 2 | Double-negative B cells (IgD$^{-}$CD27$^{-}$) | Population frequency (%) | 18 | 11 | 64.350 | 45.925 | 81.625 | 53.200 | 27.300 | 68.000 | 11.150 | 0.343 | -0.111 | 0.737 | 0.672 | 0.132093 | 0.287962 | Yes | Evaluable |
| T3 | Pre-surgery | Panel 2 | CD38$^{-}$ double-negative B cells | Population frequency (%) | 17 | 11 | 9.190 | 1.930 | 12.300 | 16.000 | 9.030 | 20.700 | -6.810 | -0.487 | -0.818 | -0.101 | 0.257 | 0.034251 | | No | Evaluable |
| T3 | Pre-surgery | Panel 2 | CD38$^{+}$ double-negative B cells | Population frequency (%) | 17 | 11 | 89.300 | 86.200 | 97.300 | 82.100 | 74.800 | 89.050 | 7.200 | 0.487 | 0.080 | 0.829 | 0.743 | 0.032596 | | No | Evaluable |
| T3 | Pre-surgery | Panel 2 | CD38$^{-}$ naive B cells | Population frequency (%) | 5 | 5 | 13.700 | 7.810 | 22.700 | 12.100 | 7.690 | 20.700 | 1.600 | 0.200 | -0.600 | 0.840 | 0.600 | 0.690476 | | No | Evaluable |
| T3 | Pre-surgery | Panel 2 | CD38$^{+}$ naive B cells | Population frequency (%) | 5 | 5 | 83.100 | 72.700 | 86.700 | 87.100 | 77.700 | 92.300 | -4.000 | -0.280 | -1.000 | 0.520 | 0.360 | 0.547619 | | No | Evaluable |
| T3 | Pre-surgery | Panel 2 | Unswitched-memory B cells (IgD$^{+}$CD27$^{+}$) | Population frequency (%) | 18 | 11 | 0.145 | 0.000 | 0.698 | 1.020 | 0.540 | 1.445 | -0.875 | -0.465 | -0.833 | -0.045 | 0.268 | 0.038775 | 0.124081 | Yes | Evaluable |
| T3 | Pre-surgery | Panel 2 | CD38$^{-}$ unswitched-memory B cells | Population frequency (%) | 1 | 3 | | | | | | | | | | | | | | No | Not evaluable: <5 samples in one group |
| T3 | Pre-surgery | Panel 2 | CD38$^{+}$ unswitched-memory B cells | Population frequency (%) | 1 | 3 | | | | | | | | | | | | | | No | Not evaluable: <5 samples in one group |
| T3 | Pre-surgery | Panel 2 | Mature dendritic cells (HLA-DR$^{+}$CD83$^{+}$) | Population frequency (%) | 18 | 11 | 8.435 | 4.108 | 13.450 | 11.200 | 5.265 | 14.050 | -2.765 | -0.131 | -0.556 | 0.313 | 0.434 | 0.580115 | 0.773487 | Yes | Evaluable |
| T3 | Pre-surgery | Panel 2 | Immature dendritic cells (HLA-DR$^{+}$CD83$^{-}$) | Population frequency (%) | 18 | 11 | 13.650 | 6.355 | 15.775 | 11.300 | 2.840 | 15.350 | 2.350 | 0.217 | -0.283 | 0.646 | 0.609 | 0.345172 | 0.613639 | Yes | Evaluable |
| T3 | Pre-surgery | Panel 2 | CD38 balance in switched-memory B cells | CD38 compositional balance (%) | 14 | 9 | 62.595 | 57.910 | 80.072 | 50.000 | 37.539 | 55.951 | 12.595 | 0.651 | 0.254 | 0.937 | 0.825 | 0.008568 | 0.066656 | Yes | Evaluable |
| T3 | Pre-surgery | Panel 2 | CD38 balance in double-negative B cells | CD38 compositional balance (%) | 17 | 11 | 90.669 | 87.513 | 98.055 | 83.690 | 78.512 | 90.793 | 6.979 | 0.487 | 0.070 | 0.818 | 0.743 | 0.032596 | 0.124081 | Yes | Evaluable |
| T3 | Pre-surgery | Panel 2 | CD38 balance in naive B cells | CD38 compositional balance (%) | 5 | 5 | 85.847 | 76.205 | 91.736 | 87.802 | 78.963 | 92.309 | -1.955 | -0.200 | -0.840 | 0.600 | 0.400 | 0.690476 | 0.841155 | Yes | Evaluable |
| T3 | Pre-surgery | Panel 2 | CD38 balance in unswitched-memory B cells | CD38 compositional balance (%) | 1 | 3 | | | | | | | | | | | | | | | Not evaluable: <5 samples in one group |

APC, antigen-presenting cell; AUC, area under the receiver operating characteristic curve; CI, confidence interval; FDR, false discovery rate; n, number of patients; NK, natural killer; pCR, pathological complete response; non-pCR, absence of pCR; Q1, first quartile; Q3, third quartile; q, Benjamini–Hochberg-adjusted P value; T1, baseline before treatment; T2, after completion of the first treatment block; T3, after completion of neoadjuvant treatment and before surgery.

**Supplementary Table 10. Comparisons of within-patient changes in circulating immune populations between pCR and non-pCR patients.**

| Interval | Panel | Immune population | Metric | n pCR | n non-pCR | pCR earlier, median [IQR] | pCR later, median [IQR] | pCR paired change, median [IQR] | pCR within-group P | non-pCR earlier, median [IQR] | non-pCR later, median [IQR] | non-pCR paired change, median [IQR] | non-pCR within-group P | Difference in median changes (pCR − non-pCR) | Cliff's δ for paired change | Cliff's δ, 95% CI | AUC (greater change in pCR) | Group with greater change | Between-group P | Panel-specific FDR q | Between-group evaluation status |
|---|---|---|---|---|---|---|---|---|---|---|---|---|---|---|---|---|---|---|---|---|---|
| Early change (T1–T2) | Panel 1 | Total CD3+ T cells | Population frequency | 17 | 11 | 76.70 [68.10–82.10] | 73.70 [66.50–82.60] | -3.40 [-8.40–5.70] | 0.2842 | 73.30 [65.25–78.30] | 72.00 [68.15–75.90] | 0.00 [-7.90–3.65] | 0.8457 | -3.40 | -0.070 | -0.519 to 0.380 | 0.465 | non-pCR | 0.7814 | 0.8245 | Evaluable |
| Early change (T1–T2) | Panel 1 | CD4+ T cells | Population frequency | 17 | 10 | 59.80 [54.40–64.20] | 48.90 [39.70–65.40] | -7.60 [-13.70–0.10] | 0.0267 | 61.40 [50.62–65.60] | 56.90 [51.23–63.83] | 1.55 [-3.42–2.73] | 0.8457 | -9.15 | -0.259 | -0.718 to 0.224 | 0.371 | non-pCR | 0.2860 | 0.8245 | Evaluable |
| Early change (T1–T2) | Panel 1 | CD4+CCR7+ T cells | Population frequency | 17 | 10 | 57.90 [41.90–71.40] | 48.50 [37.40–65.30] | -6.10 [-23.20–4.20] | 0.3060 | 53.85 [29.70–69.58] | 31.85 [20.85–73.47] | -19.00 [-36.27–16.18] | 0.6953 | 12.90 | 0.106 | -0.412 to 0.612 | 0.553 | pCR | 0.6750 | 0.8245 | Evaluable |
| Early change (T1–T2) | Panel 1 | CD4+CD161+ T cells | Population frequency | 17 | 10 | 10.20 [5.07–15.20] | 10.40 [7.30–12.70] | 0.10 [-3.70–2.93] | 0.8900 | 7.25 [5.32–11.24] | 8.05 [4.58–10.57] | -2.21 [-4.62–2.38] | 0.3223 | 2.32 | 0.129 | -0.318 to 0.588 | 0.565 | pCR | 0.6043 | 0.8245 | Evaluable |
| Early change (T1–T2) | Panel 1 | CD4+TIM-3+ T cells | Population frequency | 17 | 10 | 1.49 [0.24–8.18] | 7.76 [2.49–17.90] | 1.90 [-0.14–13.26] | 0.2069 | 1.21 [0.35–3.77] | 4.78 [1.04–15.96] | 2.87 [0.69–15.36] | 0.0195 | -0.97 | -0.235 | -0.647 to 0.224 | 0.382 | non-pCR | 0.3338 | 0.8245 | Evaluable |
| Early change (T1–T2) | Panel 1 | CD4+TIGIT+ T cells | Population frequency | 17 | 10 | 19.10 [11.30–32.30] | 15.30 [10.90–23.10] | -4.10 [-10.80–2.52] | 0.4307 | 11.85 [10.80–16.30] | 8.71 [6.82–18.15] | -0.88 [-6.09–9.62] | 0.8203 | -3.22 | -0.212 | -0.624 to 0.212 | 0.394 | non-pCR | 0.3863 | 0.8245 | Evaluable |
| Early change (T1–T2) | Panel 1 | CD4+CD127+CD27+ T cells | Population frequency | 17 | 10 | 69.30 [62.10–73.70] | 64.60 [53.70–69.70] | -5.30 [-11.90–0.90] | 0.0150 | 65.75 [54.33–79.38] | 60.15 [53.62–67.78] | -5.55 [-16.60–3.10] | 0.2324 | 0.25 | 0.059 | -0.459 to 0.553 | 0.529 | pCR | 0.8245 | 0.8245 | Evaluable |
| Early change (T1–T2) | Panel 1 | CD4+CD127+CD27+CD62L+CCR7+ T cells | Population frequency | 17 | 12 | 0.40 [0.23–1.75] | 0.55 [0.14–3.23] | -0.13 [-1.60–0.52] | 0.9632 | 0.77 [0.42–1.48] | 0.26 [0.12–0.81] | -0.46 [-0.77–0.00] | 0.1763 | 0.33 | 0.206 | -0.225 to 0.618 | 0.603 | pCR | 0.3698 | 0.8245 | Evaluable |
| Early change (T1–T2) | Panel 1 | CD8+ T cells | Population frequency | 17 | 10 | 27.90 [22.80–32.50] | 26.70 [23.00–34.70] | 0.80 [-2.00–6.50] | 0.3060 | 24.95 [18.62–29.70] | 29.60 [22.20–30.80] | 0.95 [-3.20–2.56] | 0.6953 | -0.15 | 0.082 | -0.383 to 0.506 | 0.541 | pCR | 0.7441 | 0.8245 | Evaluable |
| Early change (T1–T2) | Panel 1 | CD8+CCR7+ T cells | Population frequency | 17 | 11 | 31.80 [19.50–56.20] | 29.00 [19.00–43.20] | -8.80 [-21.20–7.90] | 0.1901 | 37.80 [13.30–53.20] | 17.30 [8.07–28.90] | -15.60 [-26.11–3.52] | 0.1230 | 6.80 | 0.123 | -0.326 to 0.561 | 0.561 | pCR | 0.6106 | 0.8245 | Evaluable |
| Early change (T1–T2) | Panel 1 | CD8+CD161+ T cells | Population frequency | 17 | 11 | 6.46 [3.96–10.10] | 5.62 [3.05–8.49] | -1.46 [-5.09–1.41] | 0.3060 | 5.27 [4.22–12.00] | 6.20 [3.95–8.13] | 0.10 [-4.95–0.90] | 0.6377 | -1.56 | -0.070 | -0.540 to 0.380 | 0.465 | non-pCR | 0.7814 | 0.8245 | Evaluable |
| Early change (T1–T2) | Panel 1 | CD8+TIM-3+ T cells | Population frequency | 17 | 11 | 3.32 [0.50–4.68] | 2.38 [1.13–5.85] | -1.09 [-2.30–3.20] | 0.5171 | 2.06 [1.23–3.27] | 1.58 [0.52–10.06] | -0.06 [-1.94–4.22] | 0.8984 | -1.03 | -0.112 | -0.529 to 0.348 | 0.444 | non-pCR | 0.6435 | 0.8245 | Evaluable |
| Early change (T1–T2) | Panel 1 | CD8+TIGIT+ T cells | Population frequency | 17 | 11 | 30.20 [25.70–56.00] | 30.10 [16.60–38.20] | -5.20 [-18.20–7.20] | 0.2868 | 29.80 [24.85–32.35] | 17.70 [8.56–30.25] | -2.20 [-21.20–1.17] | 0.1230 | -3.00 | 0.134 | -0.316 to 0.572 | 0.567 | pCR | 0.5724 | 0.8245 | Evaluable |
| Early change (T1–T2) | Panel 1 | CD8+CD127+CD27+ T cells | Population frequency | 17 | 11 | 42.70 [32.90–52.70] | 36.30 [21.90–49.70] | -6.50 [-16.50–2.80] | 0.0017 | 47.40 [39.00–54.80] | 35.60 [31.80–62.15] | -11.40 [-15.27–7.00] | 0.3652 | 4.90 | -0.112 | -0.561 to 0.358 | 0.444 | non-pCR | 0.6435 | 0.8245 | Evaluable |
| Early change (T1–T2) | Panel 1 | CD8+CD127+CD27+CD62L+CCR7+ T cells | Population frequency | 17 | 10 | 0.37 [0.16–1.03] | 0.50 [0.18–1.12] | 0.28 [-0.10–0.75] | 0.3060 | 0.65 [0.47–0.77] | 0.35 [0.14–0.81] | -0.37 [-0.65–0.18] | 0.3750 | 0.65 | 0.341 | -0.141 to 0.788 | 0.671 | pCR | 0.1550 | 0.8245 | Evaluable |
| Early change (T1–T2) | Panel 1 | CD8+CD62L+CCR7+ T cells | Population frequency | 17 | 11 | 0.22 [0.16–1.53] | 0.29 [0.24–1.70] | 0.10 [-0.14–0.78] | 0.5791 | 0.94 [0.32–1.98] | 0.20 [0.09–1.03] | -0.44 [-1.66–0.02] | 0.1475 | 0.54 | 0.316 | -0.123 to 0.711 | 0.658 | pCR | 0.1755 | 0.8245 | Evaluable |
| Early change (T1–T2) | Panel 1 | CD8+CD62L+CCR7+CD127+ T cells | Population frequency | 0 | 0 | | | | | | | | | | | | | | | | *Not evaluable: <5 paired patients in one group* |
| Early change (T1–T2) | Panel 2 | Total CD3+ T cells | Population frequency | 16 | 11 | 70.80 [61.60–75.40] | 58.20 [46.60–65.30] | -12.45 [-19.43–2.77] | 0.0034 | 69.20 [63.60–71.90] | 63.70 [59.00–67.00] | -7.40 [-9.80–1.70] | 0.2402 | -5.05 | -0.352 | -0.739 to 0.080 | 0.324 | non-pCR | 0.1341 | 0.9807 | Evaluable |
| Early change (T1–T2) | Panel 2 | NK cells | Population frequency | 16 | 11 | 2.59 [1.90–3.82] | 2.65 [1.42–3.73] | -0.58 [-1.67–1.08] | 0.4037 | 3.33 [2.04–4.64] | 2.95 [1.72–4.54] | -1.44 [-2.13–1.68] | 0.7002 | 0.86 | -0.023 | -0.477 to 0.477 | 0.489 | non-pCR | 0.9420 | 0.9807 | Evaluable |
| Early change (T1–T2) | Panel 2 | Total B cells | Population frequency | 16 | 11 | 6.08 [4.18–8.04] | 5.51 [2.27–7.33] | -1.19 [-3.30–0.62] | 0.1928 | 5.53 [2.90–7.04] | 5.09 [3.21–6.65] | -1.06 [-2.55–1.22] | 0.3652 | -0.13 | -0.011 | -0.466 to 0.420 | 0.494 | non-pCR | 0.9807 | 0.9807 | Evaluable |
| Early change (T1–T2) | Panel 2 | CD20+CD38+ mature B cells | Population frequency | 16 | 11 | 31.00 [21.88–35.25] | 36.80 [16.64–44.42] | 0.80 [-8.78–8.57] | 0.6496 | 35.40 [28.35–43.90] | 40.00 [31.00–50.80] | 4.10 [-1.50–11.55] | 0.1816 | -3.30 | -0.148 | -0.591 to 0.307 | 0.426 | non-pCR | 0.5437 | 0.9807 | Evaluable |
| Early change (T1–T2) | Panel 2 | CD20+CD38− memory B cells | Population frequency | 16 | 11 | 33.05 [24.30–41.88] | 26.40 [20.30–34.02] | -0.50 [-12.57–3.46] | 0.4037 | 43.30 [31.35–47.55] | 31.70 [23.15–36.35] | -11.60 [-17.30–1.70] | 0.0537 | 11.10 | 0.205 | -0.261 to 0.636 | 0.602 | pCR | 0.3943 | 0.9807 | Evaluable |
| Early change (T1–T2) | Panel 2 | CD20−CD38+ plasmablasts | Population frequency | 16 | 11 | 18.05 [14.12–30.43] | 22.90 [15.88–35.33] | 0.95 [-7.58–13.53] | 0.5966 | 9.53 [7.83–24.95] | 18.10 [13.70–23.10] | 1.61 [-10.35–12.62] | 0.7646 | -0.66 | 0.034 | -0.420 to 0.489 | 0.517 | pCR | 0.9035 | 0.9807 | Evaluable |
| Early change (T1–T2) | Panel 2 | Switched-memory B cells (IgD−CD27+) | Population frequency | 16 | 11 | 27.55 [22.77–43.55] | 22.15 [15.50–33.23] | -3.45 [-11.53–1.70] | 0.0042 | 36.60 [26.40–39.90] | 25.30 [18.05–35.95] | -3.96 [-7.50–2.35] | 0.0322 | 0.51 | 0.045 | -0.409 to 0.523 | 0.523 | pCR | 0.8653 | 0.9807 | Evaluable |
| Early change (T1–T2) | Panel 2 | CD38+ switched-memory B cells | Population frequency | 14 | 10 | 43.95 [38.00–52.73] | 47.40 [42.33–54.65] | -0.55 [-12.47–10.30] | 0.9515 | 45.20 [35.92–47.30] | 44.60 [41.83–50.77] | 3.30 [-1.07–6.70] | 0.0645 | -3.85 | -0.250 | -0.700 to 0.229 | 0.375 | non-pCR | 0.3194 | | Evaluable |
| Early change (T1–T2) | Panel 2 | CD38− switched-memory B cells | Population frequency | 14 | 10 | 52.55 [43.33–57.05] | 47.30 [40.52–54.10] | 1.05 [-10.57–12.35] | 0.9515 | 50.55 [48.15–59.48] | 50.90 [44.17–54.40] | -2.95 [-5.47–1.05] | 0.0840 | 4.00 | 0.193 | -0.286 to 0.672 | 0.596 | pCR | 0.4464 | | Evaluable |
| Early change (T1–T2) | Panel 2 | CD83+HLA-DRhi APC-like B cells | Population frequency | 16 | 11 | 22.75 [14.03–34.77] | 26.75 [18.85–42.40] | 6.15 [0.90–13.94] | 0.2744 | 23.10 [9.25–35.75] | 32.60 [18.70–52.60] | 6.10 [3.85–15.70] | 0.0186 | 0.05 | -0.091 | -0.523 to 0.352 | 0.455 | non-pCR | 0.7160 | 0.9807 | Evaluable |
| Early change (T1–T2) | Panel 2 | Naive B cells (IgD+CD27−) | Population frequency | 16 | 11 | 25.60 [11.22–35.50] | 28.75 [7.77–36.20] | 3.30 [-10.45–11.95] | 0.7820 | 33.60 [22.40–40.10] | 29.30 [15.25–50.10] | -5.40 [-14.60–5.08] | 0.6221 | 8.70 | 0.142 | -0.318 to 0.602 | 0.571 | pCR | 0.5536 | 0.9807 | Evaluable |
| Early change (T1–T2) | Panel 2 | Double-negative B cells (IgD−CD27−) | Population frequency | 16 | 11 | 20.40 [13.88–27.62] | 30.35 [24.90–50.35] | 3.20 [-3.06–18.23] | 0.2979 | 16.40 [12.95–24.35] | 20.20 [15.65–33.35] | 4.60 [-6.55–15.65] | 0.4131 | -1.40 | 0.045 | -0.420 to 0.489 | 0.523 | pCR | 0.8653 | 0.9807 | Evaluable |
| Early change (T1–T2) | Panel 2 | CD38− double-negative B cells | Population frequency | 16 | 11 | 19.80 [13.60–29.95] | 22.80 [13.60–28.18] | 0.75 [-6.38–9.30] | 0.8603 | 38.90 [25.55–47.30] | 31.50 [14.05–37.60] | -11.40 [-17.10–2.00] | 0.1055 | 12.15 | 0.250 | -0.205 to 0.670 | 0.625 | pCR | 0.2942 | | Evaluable |
| Early change (T1–T2) | Panel 2 | CD38+ double-negative B cells | Population frequency | 16 | 11 | 77.05 [66.50–83.15] | 75.65 [68.15–84.15] | -0.05 [-10.22–7.62] | 0.9800 | 57.80 [48.10–71.70] | 61.60 [58.50–82.25] | 10.30 [-3.05–16.85] | 0.1748 | -10.35 | -0.216 | -0.670 to 0.239 | 0.392 | non-pCR | 0.3676 | | Evaluable |
| Early change (T1–T2) | Panel 2 | CD38− naive B cells | Population frequency | 11 | 10 | 16.60 [11.23–23.50] | 10.00 [8.15–18.45] | -3.80 [-7.45–1.17] | 0.0830 | 20.60 [15.03–24.77] | 16.35 [9.49–19.23] | -7.40 [-12.29–1.28] | 0.0840 | 3.60 | 0.200 | -0.309 to 0.691 | 0.600 | pCR | 0.4679 | | Evaluable |
| Early change (T1–T2) | Panel 2 | CD38+ naive B cells | Population frequency | 11 | 10 | 78.30 [70.80–85.20] | 87.20 [77.30–89.85] | 5.50 [1.70–8.20] | 0.0674 | 73.40 [64.20–81.70] | 80.75 [77.50–88.70] | 8.95 [-1.12–18.10] | 0.0645 | -3.45 | -0.200 | -0.709 to 0.364 | 0.400 | non-pCR | 0.4679 | | Evaluable |
| Early change (T1–T2) | Panel 2 | Unswitched-memory B cells (IgD+CD27+) | Population frequency | 16 | 11 | 5.29 [2.04–10.15] | 2.10 [1.39–3.91] | -2.25 [-7.27–0.28] | 0.0076 | 7.03 [3.57–7.75] | 2.97 [2.54–4.32] | -2.71 [-4.80–0.72] | 0.0420 | 0.47 | -0.068 | -0.489 to 0.398 | 0.466 | non-pCR | 0.7897 | 0.9807 | Evaluable |
| Early change (T1–T2) | Panel 2 | CD38− unswitched-memory B cells | Population frequency | 10 | 9 | 62.55 [58.85–66.45] | 55.55 [49.75–62.17] | -6.35 [-13.25–2.05] | 0.0488 | 62.20 [56.10–65.00] | 54.00 [37.30–54.60] | -11.60 [-19.00–8.20] | 0.0078 | 5.25 | 0.333 | -0.200 to 0.800 | 0.667 | pCR | 0.2428 | | Evaluable |
| Early change (T1–T2) | Panel 2 | CD38+ unswitched-memory B cells | Population frequency | 10 | 9 | 28.45 [27.55–34.45] | 39.20 [32.75–45.55] | 6.40 [4.25–16.23] | 0.0273 | 31.70 [25.00–33.70] | 41.50 [39.90–58.10] | 14.90 [7.40–24.90] | 0.0039 | -8.50 | -0.444 | -0.867 to 0.067 | 0.278 | non-pCR | 0.1128 | | Evaluable |
| Early change (T1–T2) | Panel 2 | Mature dendritic cells (HLA-DR+CD83+) | Population frequency | 16 | 11 | 7.13 [1.52–12.70] | 9.85 [6.16–12.47] | 1.05 [-2.77–4.14] | 0.4332 | 4.98 [2.48–9.64] | 7.16 [5.20–10.84] | 1.88 [-0.60–4.25] | 0.2061 | -0.83 | -0.114 | -0.545 to 0.330 | 0.443 | non-pCR | 0.6448 | 0.9807 | Evaluable |
| Early change (T1–T2) | Panel 2 | Immature dendritic cells (HLA-DR+CD83−) | Population frequency | 16 | 11 | 12.30 [7.05–18.00] | 10.10 [7.09–13.45] | -3.50 [-6.08–2.14] | 0.1208 | 14.10 [9.89–19.45] | 10.00 [6.57–12.15] | -5.21 [-7.33–2.00] | 0.0537 | 1.71 | 0.159 | -0.273 to 0.591 | 0.580 | pCR | 0.5052 | 0.9807 | Evaluable |
| Early change (T1–T2) | Panel 2 | CD38 balance in switched-memory B cells | CD38 compositional balance | 14 | 10 | 45.54 [40.05–54.88] | 50.05 [44.01–57.41] | -0.89 [-13.09–10.89] | 1.0000 | 47.21 [37.66–49.55] | 46.70 [43.55–53.48] | 3.31 [-1.16–6.43] | 0.0645 | -4.20 | -0.243 | -0.686 to 0.257 | 0.379 | non-pCR | 0.3407 | 0.9807 | Evaluable |
| Early change (T1–T2) | Panel 2 | CD38 balance in double-negative B cells | CD38 compositional balance | 16 | 11 | 79.66 [68.89–85.94] | 76.84 [70.75–86.09] | -0.62 [-9.84–6.92] | 0.9399 | 59.77 [50.49–73.70] | 66.17 [60.87–85.41] | 11.71 [-2.43–17.62] | 0.1230 | -12.33 | -0.250 | -0.670 to 0.193 | 0.375 | non-pCR | 0.2942 | 0.9807 | Evaluable |
| Early change (T1–T2) | Panel 2 | CD38 balance in naive B cells | CD38 compositional balance | 11 | 10 | 82.51 [74.89–88.37] | 89.71 [80.73–91.68] | 3.77 [1.38–7.83] | 0.0830 | 78.08 [72.06–84.44] | 83.20 [80.13–90.29] | 8.01 [-1.39–13.87] | 0.0645 | -4.24 | -0.218 | -0.709 to 0.309 | 0.391 | non-pCR | 0.4262 | 0.9807 | Evaluable |
| Early change (T1–T2) | Panel 2 | CD38 balance in unswitched-memory B cells | CD38 compositional balance | 10 | 9 | 30.77 [29.28–36.94] | 41.77 [34.50–47.59] | 6.82 [2.99–16.04] | 0.0273 | 32.78 [27.67–37.18] | 43.32 [42.22–61.02] | 13.94 [8.02–23.85] | 0.0039 | -7.12 | -0.400 | -0.822 to 0.133 | 0.300 | non-pCR | 0.1564 | 0.9807 | Evaluable |
| Late change (T2–T3) | Panel 1 | Total CD3+ T cells | Population frequency | 14 | 8 | 76.00 [69.55–83.67] | 75.35 [70.90–82.58] | 0.15 [-5.28–3.95] | 0.8613 | 83.50 [75.53–95.08] | 85.85 [74.83–96.92] | -1.35 [-3.38–2.50] | 0.7422 | 1.50 | -0.018 | -0.518 to 0.500 | 0.491 | non-pCR | 0.9734 | 0.9734 | Evaluable |
| Late change (T2–T3) | Panel 1 | CD4+ T cells | Population frequency | 14 | 8 | 45.65 [38.35–57.75] | 48.15 [43.00–56.00] | 3.20 [-9.25–9.85] | 0.8077 | 52.60 [44.90–63.08] | 49.95 [27.48–56.95] | -5.45 [-11.32–2.48] | 0.2500 | 8.65 | 0.268 | -0.250 to 0.714 | 0.634 | pCR | 0.3301 | 0.9734 | Evaluable |
| Late change (T2–T3) | Panel 1 | CD4+CCR7+ T cells | Population frequency | 14 | 8 | 53.40 [41.52–73.88] | 49.20 [29.00–67.00] | -3.00 [-24.43–5.92] | 0.4263 | 65.90 [25.68–89.03] | 59.35 [48.05–71.12] | 0.50 [-45.62–42.52] | 1.0000 | -3.50 | -0.107 | -0.714 to 0.500 | 0.446 | non-pCR | 0.7135 | 0.9734 | Evaluable |
| Late change (T2–T3) | Panel 1 | CD4+CD161+ T cells | Population frequency | 14 | 8 | 11.55 [6.91–13.20] | 10.45 [8.01–15.97] | -1.15 [-2.95–6.95] | 0.7148 | 5.29 [3.56–11.97] | 8.76 [5.09–17.57] | 1.25 [-4.34–7.64] | 0.6406 | -2.40 | -0.071 | -0.607 to 0.482 | 0.464 | non-pCR | 0.8154 | 0.9734 | Evaluable |
| Late change (T2–T3) | Panel 1 | CD4+TIM-3+ T cells | Population frequency | 14 | 8 | 7.49 [1.13–15.43] | 1.68 [0.79–22.32] | 0.57 [-9.26–8.74] | 0.8077 | 8.06 [0.73–15.55] | 6.75 [2.35–14.47] | 1.57 [-3.83–6.27] | 0.8438 | -1.00 | -0.071 | -0.571 to 0.482 | 0.464 | non-pCR | 0.8154 | 0.9734 | Evaluable |
| Late change (T2–T3) | Panel 1 | CD4+TIGIT+ T cells | Population frequency | 14 | 8 | 13.30 [9.12–24.98] | 17.60 [10.70–36.62] | 6.25 [-7.26–18.74] | 0.4263 | 10.49 [5.83–53.10] | 15.20 [12.78–27.32] | 5.81 [-39.58–23.58] | 0.9453 | 0.44 | 0.089 | -0.446 to 0.607 | 0.545 | pCR | 0.7639 | 0.9734 | Evaluable |
| Late change (T2–T3) | Panel 1 | CD4+CD127+CD27+ T cells | Population frequency | 14 | 8 | 68.95 [57.02–71.35] | 63.80 [55.48–71.95] | -1.55 [-11.80–8.95] | 0.8077 | 72.25 [65.67–76.38] | 73.85 [67.97–76.83] | 1.95 [-0.53–8.07] | 0.4609 | -3.50 | -0.134 | -0.643 to 0.357 | 0.433 | non-pCR | 0.6327 | 0.9734 | Evaluable |
| Late change (T2–T3) | Panel 1 | CD4+CD127+CD27+CD62L+CCR7+ T cells | Population frequency | 14 | 8 | 0.98 [0.17–3.26] | 0.36 [0.26–0.95] | -0.07 [-1.94–0.35] | 0.3575 | 0.43 [0.12–0.96] | 0.78 [0.37–1.38] | 0.67 [-0.72–1.17] | 0.5469 | -0.73 | -0.357 | -0.821 to 0.179 | 0.321 | non-pCR | 0.1876 | 0.9734 | Evaluable |
| Late change (T2–T3) | Panel 1 | CD8+ T cells | Population frequency | 14 | 8 | 26.25 [23.15–34.42] | 32.05 [20.73–41.35] | 1.15 [-3.90–4.65] | 0.4631 | 25.40 [14.28–30.50] | 23.45 [14.60–34.55] | 3.95 [-3.73–5.25] | 0.6406 | -2.80 | -0.054 | -0.589 to 0.464 | 0.473 | non-pCR | 0.8676 | 0.9734 | Evaluable |
| Late change (T2–T3) | Panel 1 | CD8+CCR7+ T cells | Population frequency | 14 | 8 | 30.35 [19.90–65.72] | 28.25 [20.95–38.15] | -0.50 [-23.10–8.71] | 0.6257 | 37.00 [16.62–90.25] | 46.70 [30.12–59.15] | 4.20 [-45.08–35.25] | 0.8438 | -4.70 | -0.054 | -0.643 to 0.518 | 0.473 | non-pCR | 0.8676 | 0.9734 | Evaluable |
| Late change (T2–T3) | Panel 1 | CD8+CD161+ T cells | Population frequency | 14 | 8 | 5.95 [3.28–9.44] | 5.87 [4.47–9.26] | 0.68 [-2.64–2.90] | 0.7609 | 5.70 [3.50–9.22] | 8.35 [6.99–13.08] | 3.27 [-2.28–9.65] | 0.3125 | -2.60 | -0.286 | -0.786 to 0.321 | 0.357 | non-pCR | 0.2973 | 0.9734 | Evaluable |
| Late change (T2–T3) | Panel 1 | CD8+TIM-3+ T cells | Population frequency | 14 | 8 | 2.58 [0.67–9.49] | 0.77 [0.49–4.09] | -0.51 [-8.80–0.46] | 0.4631 | 6.33 [0.92–9.87] | 6.09 [2.41–19.05] | 4.02 [-1.21–12.07] | 0.4609 | -4.53 | -0.304 | -0.804 to 0.232 | 0.348 | non-pCR | 0.2667 | 0.9734 | Evaluable |
| Late change (T2–T3) | Panel 1 | CD8+TIGIT+ T cells | Population frequency | 14 | 8 | 29.75 [22.27–48.33] | 36.00 [27.12–50.27] | 4.45 [-10.13–14.18] | 0.5016 | 28.10 [8.88–65.45] | 34.40 [21.25–42.02] | 11.99 [-28.40–12.72] | 0.8438 | -7.55 | 0.089 | -0.464 to 0.625 | 0.545 | pCR | 0.7639 | 0.9734 | Evaluable |
| Late change (T2–T3) | Panel 1 | CD8+CD127+CD27+ T cells | Population frequency | 14 | 8 | 36.85 [24.00–48.67] | 36.60 [29.23–50.95] | -3.75 [-9.88–18.70] | 0.7609 | 52.60 [43.25–67.95] | 51.15 [45.02–62.58] | -0.05 [-7.33–9.43] | 0.6406 | -3.70 | -0.036 | -0.536 to 0.429 | 0.482 | non-pCR | 0.9203 | 0.9734 | Evaluable |
| Late change (T2–T3) | Panel 1 | CD8+CD127+CD27+CD62L+CCR7+ T cells | Population frequency | 14 | 8 | 1.00 [0.25–3.96] | 0.32 [0.17–0.57] | -0.43 [-0.98–0.01] | 0.0085 | 0.22 [0.12–1.25] | 0.82 [0.37–1.03] | 0.29 [-0.74–0.90] | 0.7422 | -0.73 | -0.429 | -0.929 to 0.179 | 0.286 | non-pCR | 0.1100 | 0.9734 | Evaluable |
| Late change (T2–T3) | Panel 1 | CD8+CD62L+CCR7+ T cells | Population frequency | 14 | 8 | 0.85 [0.13–2.45] | 0.27 [0.14–0.65] | -0.40 [-1.58–0.05] | 0.0785 | 0.60 [0.15–1.68] | 0.48 [0.33–1.21] | -0.10 [-1.34–0.78] | 0.8438 | -0.30 | -0.250 | -0.732 to 0.321 | 0.375 | non-pCR | 0.3650 | 0.9734 | Evaluable |
| Late change (T2–T3) | Panel 1 | CD8+CD62L+CCR7+CD127+ T cells | Population frequency | 3 | 0 | 0.51 [0.41–2.77] | 0.58 [0.34–0.67] | 0.07 [-2.43–0.26] | 1.0000 | | | | | | | | | | | | *Not evaluable: <5 paired patients in one group* |
| Late change (T2–T3) | Panel 2 | Total CD3+ T cells | Population frequency | 14 | 8 | 59.80 [43.75–66.70] | 52.50 [32.95–67.67] | -7.45 [-31.90–17.77] | 0.4631 | 62.85 [60.27–73.97] | 64.00 [28.18–73.80] | -10.90 [-34.70–6.33] | 0.5469 | 3.45 | 0.089 | -0.464 to 0.589 | 0.545 | pCR | 0.7639 | 0.9155 | Evaluable |
| Late change (T2–T3) | Panel 2 | NK cells | Population frequency | 14 | 8 | 2.21 [1.19–4.11] | 2.84 [2.04–3.87] | 0.30 [-1.72–2.15] | 0.5416 | 1.68 [1.28–2.98] | 3.20 [0.86–5.32] | 0.39 [-1.04–3.97] | 0.4609 | -0.09 | -0.143 | -0.643 to 0.358 | 0.429 | non-pCR | 0.6163 | 0.9155 | Evaluable |
| Late change (T2–T3) | Panel 2 | Total B cells | Population frequency | 14 | 8 | 4.85 [1.72–5.69] | 0.83 [0.47–2.04] | -2.02 [-5.22–0.52] | 0.0085 | 2.60 [0.74–4.62] | 0.51 [0.36–0.70] | -0.90 [-3.03–0.01] | 0.1484 | -1.12 | -0.161 | -0.661 to 0.357 | 0.420 | non-pCR | 0.5699 | 0.9155 | Evaluable |
| Late change (T2–T3) | Panel 2 | CD20+CD38+ mature B cells | Population frequency | 14 | 8 | 36.80 [22.43–42.80] | 6.26 [0.61–15.35] | -26.60 [-38.55–10.59] | 0.0002 | 22.50 [12.28–35.73] | 11.12 [4.53–23.82] | -4.12 [-19.47–3.85] | 0.3125 | -22.48 | -0.482 | -0.929 to 0.071 | 0.259 | non-pCR | 0.0698 | 0.5237 | Evaluable |
| Late change (T2–T3) | Panel 2 | CD20+CD38− memory B cells | Population frequency | 14 | 8 | 25.65 [15.72–31.75] | 8.46 [0.94–17.70] | -9.75 [-20.86–6.29] | 0.0004 | 30.35 [14.72–42.75] | 29.85 [8.51–39.62] | 4.10 [-12.60–9.78] | 0.9453 | -13.85 | -0.393 | -0.875 to 0.179 | 0.304 | non-pCR | 0.1450 | 0.5956 | Evaluable |
| Late change (T2–T3) | Panel 2 | CD20−CD38+ plasmablasts | Population frequency | 14 | 8 | 25.35 [17.70–33.58] | 62.50 [51.23–88.90] | 34.35 [21.07–45.40] | 0.0001 | 21.50 [18.93–33.58] | 46.60 [36.80–63.02] | 14.35 [-7.89–40.02] | 0.3125 | 20.00 | 0.357 | -0.196 to 0.857 | 0.679 | pCR | 0.1876 | 0.5956 | Evaluable |
| Late change (T2–T3) | Panel 2 | Switched-memory B cells (IgD−CD27+) | Population frequency | 14 | 8 | 22.15 [14.50–28.50] | 22.00 [8.13–42.55] | -0.75 [-11.56–16.58] | 1.0000 | 22.60 [17.75–32.23] | 30.35 [13.66–44.35] | 3.38 [-7.71–21.57] | 0.6406 | -4.13 | -0.071 | -0.607 to 0.482 | 0.464 | non-pCR | 0.8154 | 0.9155 | Evaluable |
| Late change (T2–T3) | Panel 2 | CD38+ switched-memory B cells | Population frequency | 10 | 6 | 52.10 [48.42–57.17] | 59.90 [51.80–69.83] | 12.00 [-0.90–14.32] | 0.0645 | 43.05 [34.27–70.50] | 45.90 [31.45–54.05] | -9.95 [-21.95–10.75] | 0.5625 | 21.95 | 0.367 | -0.333 to 1.000 | 0.683 | pCR | 0.2635 | | Evaluable |
| Late change (T2–T3) | Panel 2 | CD38− switched-memory B cells | Population frequency | 10 | 6 | 42.15 [37.75–46.80] | 34.60 [26.90–41.88] | -10.55 [-12.31–0.50] | 0.0371 | 52.60 [26.55–62.52] | 49.00 [42.55–64.75] | 11.15 [-10.40–22.02] | 0.5625 | -21.70 | -0.400 | -1.000 to 0.333 | 0.300 | non-pCR | 0.2198 | | Evaluable |
| Late change (T2–T3) | Panel 2 | CD83+HLA-DRhi APC-like B cells | Population frequency | 14 | 8 | 26.75 [20.82–46.08] | 20.65 [15.75–33.48] | -4.25 [-31.52–10.10] | 0.6257 | 28.10 [17.20–48.67] | 30.25 [15.90–39.20] | 0.10 [-40.33–18.80] | 0.6406 | -4.36 | 0.054 | -0.536 to 0.571 | 0.527 | pCR | 0.8676 | 0.9155 | Evaluable |
| Late change (T2–T3) | Panel 2 | Naive B cells (IgD+CD27−) | Population frequency | 14 | 8 | 33.30 [17.20–46.38] | 0.57 [0.30–5.24] | -30.14 [-35.44–16.13] | 0.0001 | 17.60 [10.47–33.38] | 3.40 [0.82–6.36] | -14.47 [-31.76–4.07] | 0.0547 | -15.68 | -0.321 | -0.857 to 0.250 | 0.339 | non-pCR | 0.2382 | 0.5956 | Evaluable |
| Late change (T2–T3) | Panel 2 | Double-negative B cells (IgD−CD27−) | Population frequency | 14 | 8 | 28.25 [20.77–40.98] | 64.35 [45.92–88.38] | 30.35 [17.10–49.83] | 0.0001 | 38.00 [22.32–46.45] | 57.30 [27.75–68.78] | 14.25 [-6.80–49.58] | 0.3125 | 16.10 | 0.250 | -0.375 to 0.786 | 0.625 | pCR | 0.3650 | 0.7822 | Evaluable |
| Late change (T2–T3) | Panel 2 | CD38− double-negative B cells | Population frequency | 13 | 8 | 26.90 [17.90–27.70] | 9.19 [1.93–10.30] | -16.40 [-20.56–5.45] | 0.0024 | 33.40 [15.80–38.98] | 16.85 [12.82–19.40] | -9.56 [-26.45–6.10] | 0.2500 | -6.84 | -0.058 | -0.654 to 0.520 | 0.471 | non-pCR | 0.8596 | | Evaluable |
| Late change (T2–T3) | Panel 2 | CD38+ double-negative B cells | Population frequency | 13 | 8 | 70.20 [68.50–78.30] | 89.30 [88.10–97.30] | 16.50 [6.70–21.70] | 0.0061 | 61.35 [57.27–81.30] | 80.40 [75.80–86.10] | 11.25 [-5.45–29.77] | 0.1953 | 5.25 | 0.038 | -0.558 to 0.635 | 0.519 | pCR | 0.9135 | | Evaluable |
| Late change (T2–T3) | Panel 2 | CD38− naive B cells | Population frequency | 3 | 1 | 8.80 [8.41–12.25] | 7.81 [6.55–10.76] | -0.99 [-5.70–2.34] | 0.7500 | 16.70 [16.70–16.70] | 27.50 [27.50–27.50] | 10.80 [10.80–10.80] | | -11.79 | | | | | | | *Not evaluable: <5 paired patients in one group* |
| Late change (T2–T3) | Panel 2 | CD38+ naive B cells | Population frequency | 3 | 1 | 89.00 [84.45–89.45] | 86.70 [84.90–90.05] | -2.30 [-4.55–5.60] | 1.0000 | 81.20 [81.20–81.20] | 70.40 [70.40–70.40] | -10.80 [-10.80–10.80] | | 8.50 | | | | | | | *Not evaluable: <5 paired patients in one group* |
| Late change (T2–T3) | Panel 2 | Unswitched-memory B cells (IgD+CD27+) | Population frequency | 14 | 8 | 2.10 [1.30–3.95] | 0.11 [0.00–0.70] | -2.06 [-3.95–0.49] | 0.0006 | 1.20 [0.61–2.89] | 0.91 [0.57–1.32] | 0.00 [-1.35–0.22] | 0.6406 | -2.06 | -0.589 | -0.929 to -0.143 | 0.205 | non-pCR | 0.0240 | 0.3603 | Evaluable |
| Late change (T2–T3) | Panel 2 | CD38− unswitched-memory B cells | Population frequency | 0 | 1 | | | | | 54.30 [54.30–54.30] | 60.20 [60.20–60.20] | 5.90 [5.90–5.90] | | | | | | | | | *Not evaluable: <5 paired patients in one group* |
| Late change (T2–T3) | Panel 2 | CD38+ unswitched-memory B cells | Population frequency | 0 | 1 | | | | | 41.50 [41.50–41.50] | 35.20 [35.20–35.20] | -6.30 [-6.30–6.30] | | | | | | | | | *Not evaluable: <5 paired patients in one group* |
| Late change (T2–T3) | Panel 2 | Mature dendritic cells (HLA-DR+CD83+) | Population frequency | 14 | 8 | 8.94 [5.70–10.30] | 8.44 [5.40–13.45] | 1.40 [-0.95–5.44] | 0.4263 | 8.52 [6.10–14.72] | 10.38 [4.62–12.88] | -1.76 [-6.38–3.60] | 0.7422 | 3.17 | 0.196 | -0.375 to 0.714 | 0.598 | pCR | 0.4822 | 0.9041 | Evaluable |
| Late change (T2–T3) | Panel 2 | Immature dendritic cells (HLA-DR+CD83−) | Population frequency | 14 | 8 | 9.95 [6.08–12.93] | 12.45 [6.36–15.50] | 0.55 [-2.01–6.55] | 0.4631 | 10.29 [3.83–13.40] | 8.30 [2.92–15.32] | 2.19 [-11.01–5.25] | 0.7422 | -1.64 | 0.071 | -0.536 to 0.661 | 0.536 | pCR | 0.8154 | 0.9155 | Evaluable |
| Late change (T2–T3) | Panel 2 | CD38 balance in switched-memory B cells | CD38 compositional balance | 10 | 6 | 55.28 [50.86–60.24] | 62.18 [56.27–72.20] | 11.40 [-1.22–14.07] | 0.0645 | 45.08 [35.41–72.49] | 48.68 [32.72–55.65] | -10.73 [-22.04–11.42] | 0.5625 | 22.13 | 0.400 | -0.333 to 1.000 | 0.700 | pCR | 0.2198 | 0.5956 | Evaluable |
| Late change (T2–T3) | Panel 2 | CD38 balance in double-negative B cells | CD38 compositional balance | 13 | 8 | 72.15 [71.41–81.39] | 90.67 [89.63–98.06] | 17.00 [5.59–21.40] | 0.0024 | 64.77 [59.51–83.86] | 82.50 [79.89–87.05] | 10.43 [-6.01–28.38] | 0.1953 | 6.57 | 0.038 | -0.558 to 0.596 | 0.519 | pCR | 0.9155 | 0.9155 | Evaluable |
| Late change (T2–T3) | Panel 2 | CD38 balance in naive B cells | CD38 compositional balance | 3 | 1 | 91.00 [87.29–91.40] | 91.74 [88.79–93.18] | 0.73 [-2.61–5.89] | 0.7500 | 82.94 [82.94–82.94] | 71.91 [71.91–71.91] | -11.03 [-11.03–11.03] | | 11.77 | | | | | | | *Not evaluable: <5 paired patients in one group* |
| Late change (T2–T3) | Panel 2 | CD38 balance in unswitched-memory B cells | CD38 compositional balance | 0 | 1 | | | | | 43.32 [43.32–43.32] | 36.90 [36.90–36.90] | -6.42 [-6.42–6.42] | | | | | | | | | *Not evaluable: <5 paired patients in one group* |
| Overall change (T1–T3) | Panel 1 | Total CD3+ T cells | Population frequency | 14 | 5 | 75.45 [68.60–82.55] | 75.35 [67.58–78.22] | -1.20 [-12.50–7.03] | 0.5935 | 73.30 [69.30–79.50] | 73.10 [66.70–75.40] | 0.60 [-16.30–2.10] | 1.0000 | -1.80 | 0.171 | -0.515 to 0.801 | 0.586 | pCR | 0.6105 | 0.6977 | Evaluable |
| Overall change (T1–T3) | Panel 1 | CD4+ T cells | Population frequency | 14 | 4 | 55.65 [44.90–60.10] | 51.85 [44.42–56.77] | -4.35 [-9.72–0.72] | 0.3258 | 62.90 [58.02–64.78] | 54.60 [46.65–57.33] | -5.05 [-13.93–1.65] | 0.6250 | 0.70 | | | | | | | *Not evaluable: <5 paired patients in one group* |
| Overall change (T1–T3) | Panel 1 | CD4+CCR7+ T cells | Population frequency | 14 | 4 | 63.10 [53.05–73.58] | 46.85 [29.00–67.00] | -18.65 [-38.33–3.62] | 0.0906 | 68.60 [48.27–85.60] | 56.05 [44.83–65.20] | -7.85 [-21.23–2.08] | 0.3750 | -10.80 | | | | | | | *Not evaluable: <5 paired patients in one group* |
| Overall change (T1–T3) | Panel 1 | CD4+CD161+ T cells | Population frequency | 14 | 4 | 10.95 [3.31–14.95] | 8.70 [5.42–13.60] | 0.52 [-3.88–3.16] | 0.9032 | 7.25 [5.75–9.35] | 8.76 [7.41–11.17] | 0.17 [-1.51–3.66] | 0.8750 | 0.35 | | | | | | | *Not evaluable: <5 paired patients in one group* |
| Overall change (T1–T3) | Panel 1 | CD4+TIM-3+ T cells | Population frequency | 14 | 4 | 1.83 [0.22–25.68] | 1.68 [0.88–10.39] | -0.40 [-15.55–1.90] | 0.4263 | 1.18 [0.24–2.65] | 3.17 [2.35–7.32] | 2.95 [1.08–6.67] | 0.3750 | -3.35 | | | | | | | *Not evaluable: <5 paired patients in one group* |
| Overall change (T1–T3) | Panel 1 | CD4+TIGIT+ T cells | Population frequency | 14 | 4 | 26.15 [15.38–32.45] | 20.50 [10.70–30.77] | -6.55 [-16.88–9.01] | 0.4263 | 10.53 [5.96–16.25] | 15.20 [13.32–20.00] | 4.53 [-0.15–11.13] | 0.3750 | -11.08 | | | | | | | *Not evaluable: <5 paired patients in one group* |
| Overall change (T1–T3) | Panel 1 | CD4+CD127+CD27+ T cells | Population frequency | 14 | 4 | 67.95 [56.92–73.40] | 65.15 [55.62–70.72] | -2.60 [-6.05–0.68] | 0.5830 | 84.75 [76.33–85.83] | 77.45 [70.40–80.50] | -4.80 [-14.88–6.12] | 0.7500 | 2.20 | | | | | | | *Not evaluable: <5 paired patients in one group* |
| Overall change (T1–T3) | Panel 1 | CD4+CD127+CD27+CD62L+CCR7+ T cells | Population frequency | 14 | 7 | 0.47 [0.24–5.96] | 0.45 [0.28–0.82] | -0.07 [-2.43–0.46] | 0.2958 | 1.38 [0.54–6.61] | 1.64 [1.02–7.58] | 0.69 [0.24–2.86] | 0.2969 | -0.77 | -0.551 | -0.939 to -0.041 | 0.224 | non-pCR | 0.0461 | 0.3688 | Evaluable |
| Overall change (T1–T3) | Panel 1 | CD8+ T cells | Population frequency | 14 | 4 | 26.80 [22.98–32.55] | 32.05 [21.23–37.55] | 2.70 [-2.38–8.25] | 0.2412 | 28.20 [24.32–28.80] | 35.70 [33.67–38.30] | 10.50 [6.85–15.00] | 0.1250 | -7.80 | | | | | | | *Not evaluable: <5 paired patients in one group* |
| Overall change (T1–T3) | Panel 1 | CD8+CCR7+ T cells | Population frequency | 14 | 5 | 39.60 [26.30–59.20] | 26.15 [20.95–36.70] | -16.00 [-31.85–4.50] | 0.0494 | 51.10 [30.60–55.30] | 31.00 [27.50–51.00] | -3.30 [-7.30–3.10] | 0.4375 | -12.70 | -0.343 | -0.829 to 0.200 | 0.329 | non-pCR | 0.2976 | 0.6977 | Evaluable |
| Overall change (T1–T3) | Panel 1 | CD8+CD161+ T cells | Population frequency | 14 | 5 | 5.47 [3.99–10.66] | 6.46 [3.74–11.20] | -0.03 [-4.37–2.63] | 0.8552 | 4.99 [4.39–5.22] | 7.14 [5.50–9.97] | 2.15 [1.88–3.40] | 0.1250 | -2.18 | -0.371 | -0.857 to 0.114 | 0.314 | non-pCR | 0.2566 | 0.6977 | Evaluable |
| Overall change (T1–T3) | Panel 1 | CD8+TIM-3+ T cells | Population frequency | 14 | 5 | 0.95 [0.34–16.48] | 0.83 [0.52–1.85] | 0.06 [-15.38–0.53] | 0.6698 | 2.43 [2.13–3.20] | 4.55 [2.70–7.64] | 0.74 [-1.66–2.42] | 1.0000 | -0.69 | -0.200 | -0.857 to 0.486 | 0.400 | non-pCR | 0.5593 | 0.6977 | Evaluable |
| Overall change (T1–T3) | Panel 1 | CD8+TIGIT+ T cells | Population frequency | 14 | 5 | 33.80 [27.65–60.20] | 38.05 [27.12–45.77] | -5.30 [-18.93–10.25] | 0.5016 | 17.60 [13.60–29.80] | 32.40 [21.60–38.40] | 14.80 [-8.20–24.80] | 0.6250 | -20.10 | -0.286 | -0.857 to 0.344 | 0.357 | non-pCR | 0.3913 | 0.6977 | Evaluable |
| Overall change (T1–T3) | Panel 1 | CD8+CD127+CD27+ T cells | Population frequency | 14 | 5 | 44.75 [33.42–51.60] | 36.80 [32.02–46.83] | -5.40 [-16.20–4.50] | 0.2676 | 50.80 [47.40–70.70] | 47.30 [47.20–69.40] | -0.20 [-3.50–8.70] | 1.0000 | -5.20 | -0.229 | -0.857 to 0.429 | 0.386 | non-pCR | 0.4998 | 0.6977 | Evaluable |
| Overall change (T1–T3) | Panel 1 | CD8+CD127+CD27+CD62L+CCR7+ T cells | Population frequency | 14 | 4 | 0.39 [0.17–2.33] | 0.44 [0.21–0.69] | -0.05 [-0.76–0.32] | 0.4631 | 1.16 [0.69–3.00] | 0.82 [0.72–3.42] | 0.04 [-0.17–1.00] | 0.6250 | -0.09 | | | | | | | *Not evaluable: <5 paired patients in one group* |
| Overall change (T1–T3) | Panel 1 | CD8+CD62L+CCR7+ T cells | Population frequency | 14 | 5 | 0.29 [0.16–4.25] | 0.35 [0.18–0.67] | -0.01 [-3.47–0.21] | 0.2489 | 1.49 [1.03–2.46] | 0.46 [0.37–1.91] | -1.03 [-2.25–0.58] | 0.6250 | 1.02 | 0.086 | -0.543 to 0.714 | 0.543 | pCR | 0.8230 | 0.8230 | Evaluable |
| Overall change (T1–T3) | Panel 1 | CD8+CD62L+CCR7+CD127+ T cells | Population frequency | 1 | 1 | 1.38 [1.38–1.38] | 0.07 [0.07–0.07] | -1.31 [-1.31–1.31] | | 0.83 [0.83–0.83] | 1.19 [1.19–1.19] | 0.36 [0.36–0.36] | | -1.67 | | | | | | | *Not evaluable: <5 paired patients in one group* |
| Overall change (T1–T3) | Panel 2 | Total CD3+ T cells | Population frequency | 13 | 6 | 67.10 [59.20–77.20] | 60.90 [32.10–74.90] | -1.50 [-35.90–8.50] | 0.2439 | 69.70 [66.95–71.33] | 54.10 [33.40–64.45] | -13.95 [-37.23–6.28] | 0.3125 | 12.45 | 0.205 | -0.385 to 0.769 | 0.603 | pCR | 0.5214 | 0.7821 | Evaluable |
| Overall change (T1–T3) | Panel 2 | NK cells | Population frequency | 13 | 6 | 3.56 [1.25–3.99] | 3.04 [2.30–3.87] | 0.47 [-0.52–1.07] | 0.5417 | 1.87 [1.41–2.12] | 3.20 [2.23–3.52] | 1.57 [0.66–1.81] | 0.0625 | -1.11 | -0.487 | -0.924 to 0.026 | 0.256 | non-pCR | 0.1061 | 0.7791 | Evaluable |
| Overall change (T1–T3) | Panel 2 | Total B cells | Population frequency | 13 | 6 | 5.25 [3.61–7.08] | 0.98 [0.39–1.19] | -4.75 [-6.11–3.44] | 0.0061 | 5.70 [5.55–6.24] | 1.20 [0.47–4.54] | -4.43 [-5.19–0.46] | 0.1563 | -0.32 | 0.000 | -0.590 to 0.565 | 0.500 | Tie | 1.0000 | 1.0000 | Evaluable |
| Overall change (T1–T3) | Panel 2 | CD20+CD38+ mature B cells | Population frequency | 13 | 6 | 23.40 [13.80–33.10] | 10.20 [4.94–15.50] | -10.99 [-21.49–1.27] | 0.0024 | 33.35 [28.70–41.90] | 25.95 [9.45–34.10] | -10.40 [-15.53–5.98] | 0.1563 | -0.59 | -0.103 | -0.641 to 0.462 | 0.449 | non-pCR | 0.7654 | 0.9567 | Evaluable |
| Overall change (T1–T3) | Panel 2 | CD20+CD38− memory B cells | Population frequency | 13 | 6 | 25.00 [10.50–35.20] | 7.91 [3.77–13.50] | -15.15 [-26.59–3.20] | 0.0024 | 36.10 [27.55–45.33] | 30.10 [15.80–37.42] | -0.85 [-12.78–2.00] | 0.5625 | -14.30 | -0.436 | -0.872 to 0.128 | 0.282 | non-pCR | 0.1518 | 0.7791 | Evaluable |
| Overall change (T1–T3) | Panel 2 | CD20−CD38+ plasmablasts | Population frequency | 13 | 6 | 30.30 [15.30–63.40] | 60.40 [54.60–74.30] | 32.20 [9.60–51.71] | 0.0134 | 19.40 [11.50–22.15] | 44.30 [25.82–57.08] | 15.38 [13.90–30.92] | 0.2188 | 16.82 | 0.256 | -0.282 to 0.744 | 0.628 | pCR | 0.4155 | 0.7791 | Evaluable |
| Overall change (T1–T3) | Panel 2 | Switched-memory B cells (IgD−CD27+) | Population frequency | 13 | 6 | 23.10 [17.10–36.00] | 22.60 [12.00–37.60] | -3.34 [-8.20–1.20] | 0.4548 | 36.15 [34.42–40.88] | 32.35 [16.48–40.95] | -4.92 [-12.53–5.26] | 0.8438 | 1.58 | 0.026 | -0.538 to 0.590 | 0.513 | pCR | 0.9661 | 1.0000 | Evaluable |
| Overall change (T1–T3) | Panel 2 | CD38+ switched-memory B cells | Population frequency | 7 | 5 | 56.10 [44.65–60.00] | 59.90 [49.70–66.70] | 9.90 [0.35–16.05] | 0.2188 | 46.70 [35.40–49.60] | 47.40 [40.60–51.20] | -6.10 [-6.60–6.80] | 1.0000 | 16.00 | 0.371 | -0.314 to 1.000 | 0.686 | pCR | 0.3434 | | Evaluable |
| Overall change (T1–T3) | Panel 2 | CD38− switched-memory B cells | Population frequency | 7 | 5 | 41.20 [35.55–50.65] | 40.10 [29.80–46.40] | -8.50 [-15.45–0.80] | 0.3750 | 48.60 [44.00–60.00] | 47.40 [42.30–54.70] | 3.40 [-4.00–6.10] | 1.0000 | -11.90 | -0.371 | -1.000 to 0.314 | 0.314 | non-pCR | 0.3434 | | Evaluable |
| Overall change (T1–T3) | Panel 2 | CD83+HLA-DRhi APC-like B cells | Population frequency | 13 | 6 | 18.50 [9.92–24.90] | 21.70 [17.10–34.80] | 0.70 [-5.80–23.30] | 0.4143 | 9.25 [7.37–23.48] | 29.35 [21.73–40.35] | 19.92 [12.80–24.34] | 0.0625 | -19.22 | -0.333 | -0.846 to 0.205 | 0.333 | non-pCR | 0.2727 | 0.7791 | Evaluable |
| Overall change (T1–T3) | Panel 2 | Naive B cells (IgD+CD27−) | Population frequency | 13 | 6 | 16.80 [1.32–31.30] | 2.00 [0.28–4.89] | -12.82 [-28.31–0.00] | 0.0068 | 20.55 [10.26–28.85] | 5.16 [3.23–9.10] | -13.72 [-22.98–1.02] | 0.1563 | 0.90 | -0.128 | -0.692 to 0.437 | 0.436 | non-pCR | 0.7012 | 0.9562 | Evaluable |
| Overall change (T1–T3) | Panel 2 | Double-negative B cells (IgD−CD27−) | Population frequency | 13 | 6 | 26.90 [17.00–72.40] | 62.60 [47.80–81.10] | 22.80 [8.10–50.30] | 0.0215 | 28.15 [19.62–37.65] | 49.50 [29.20–69.80] | 10.90 [5.92–28.48] | 0.0938 | 11.90 | 0.256 | -0.256 to 0.744 | 0.628 | pCR | 0.4155 | 0.7791 | Evaluable |
| Overall change (T1–T3) | Panel 2 | CD38− double-negative B cells | Population frequency | 12 | 6 | 17.05 [12.33–26.10] | 8.31 [2.98–10.32] | -10.93 [-15.21–4.03] | 0.0122 | 30.40 [21.10–41.73] | 11.84 [8.61–16.73] | -18.80 [-29.63–7.10] | 0.1563 | 7.88 | 0.194 | -0.444 to 0.806 | 0.597 | pCR | 0.5532 | | Evaluable |
| Overall change (T1–T3) | Panel 2 | CD38+ double-negative B cells | Population frequency | 12 | 6 | 80.00 [71.03–86.30] | 89.75 [87.38–96.03] | 12.45 [4.33–17.88] | 0.0269 | 65.90 [52.58–75.47] | 86.80 [78.95–89.47] | 19.95 [10.20–32.70] | 0.1563 | -7.50 | -0.250 | -0.861 to 0.389 | 0.375 | non-pCR | 0.4371 | | Evaluable |
| Overall change (T1–T3) | Panel 2 | CD38− naive B cells | Population frequency | 3 | 2 | 21.50 [15.12–21.55] | 13.70 [10.76–18.20] | -0.94 [-4.37–0.08] | 0.7500 | 24.00 [17.25–30.75] | 19.80 [15.95–23.65] | -4.20 [-7.10–1.30] | 1.0000 | 3.26 | | | | | | | *Not evaluable: <5 paired patients in one group* |
| Overall change (T1–T3) | Panel 2 | CD38+ naive B cells | Population frequency | 3 | 2 | 72.30 [71.55–80.45] | 83.10 [77.90–84.90] | 1.90 [0.00–6.35] | 0.5000 | 71.45 [63.83–79.08] | 78.75 [74.58–82.92] | 7.30 [3.85–10.75] | 0.5000 | -5.40 | | | | | | | *Not evaluable: <5 paired patients in one group* |
| Overall change (T1–T3) | Panel 2 | Unswitched-memory B cells (IgD+CD27+) | Population frequency | 13 | 6 | 2.29 [0.51–10.00] | 0.14 [0.00–0.60] | -1.36 [-9.87–0.14] | 0.0093 | 4.55 [3.29–6.70] | 1.16 [0.49–1.43] | -2.38 [-4.71–1.87] | 0.0625 | 1.03 | 0.013 | -0.487 to 0.538 | 0.506 | pCR | 1.0000 | 1.0000 | Evaluable |
| Overall change (T1–T3) | Panel 2 | CD38− unswitched-memory B cells | Population frequency | 0 | 2 | | | | | 54.40 [47.15–61.65] | 52.60 [48.80–56.40] | -1.80 [-5.25–1.65] | 1.0000 | | | | | | | | *Not evaluable: <5 paired patients in one group* |
| Overall change (T1–T3) | Panel 2 | CD38+ unswitched-memory B cells | Population frequency | 0 | 2 | | | | | 40.50 [33.40–47.60] | 42.10 [38.65–45.55] | 1.60 [-2.05–5.25] | 1.0000 | | | | | | | | *Not evaluable: <5 paired patients in one group* |
| Overall change (T1–T3) | Panel 2 | Mature dendritic cells (HLA-DR+CD83+) | Population frequency | 13 | 6 | 4.18 [1.86–15.70] | 10.00 [5.18–14.00] | -0.41 [-1.70–11.65] | 0.5879 | 3.42 [0.82–5.95] | 10.19 [5.83–11.57] | 5.49 [1.18–9.87] | 0.0625 | -5.90 | -0.359 | -0.795 to 0.179 | 0.321 | non-pCR | 0.2441 | 0.7791 | Evaluable |
| Overall change (T1–T3) | Panel 2 | Immature dendritic cells (HLA-DR+CD83−) | Population frequency | 13 | 6 | 7.35 [6.59–17.60] | 14.00 [6.29–15.40] | 2.86 [-1.70–5.40] | 0.2163 | 13.25 [10.09–17.15] | 13.00 [3.67–15.22] | -1.26 [-5.58–0.37] | 0.4375 | 4.12 | 0.410 | -0.103 to 0.872 | 0.705 | pCR | 0.1791 | 0.7791 | Evaluable |

| | | | | | | | | | | | | | | | | | | | | |
|---|---|---|---|---|---|---|---|---|---|---|---|---|---|---|---|---|---|---|---|---|
| Overall change (T1–T3) | Panel 2 | CD38 balance in switched-memory B cells | CD38 compositional balance | 7 | 5 | 57.66 [46.80–62.78] | 59.90 [51.77–69.13] | 9.76 [-0.39–16.23] | 0.3750 | 49.00 [37.11–53.97] | 50.00 [42.60–54.76] | -5.10 [-6.40–5.58] | 1.0000 | 14.86 | 0.371 | -0.257 to 1.000 | 0.686 | pCR | 0.3434 | 0.7791 | Evaluable |
| Overall change (T1–T3) | Panel 2 | CD38 balance in double-negative B cells | CD38 compositional balance | 12 | 6 | 82.32 [73.13–87.29] | 91.52 [89.35–96.98] | 11.18 [3.97–15.75] | 0.0161 | 68.28 [55.75–78.16] | 88.01 [82.47–91.22] | 19.95 [7.77–31.59] | 0.1563 | -8.77 | -0.222 | -0.806 to 0.444 | 0.389 | non-pCR | 0.4936 | 0.7821 | Evaluable |
| Overall change (T1–T3) | Panel 2 | CD38 balance in naive B cells | CD38 compositional balance | 3 | 2 | 77.08 [76.85–84.05] | 85.85 [81.03–88.79] | 0.72 [0.15–4.75] | 0.5000 | 74.59 [67.28–81.89] | 79.86 [75.88–83.83] | 5.27 [1.94–8.60] | 1.0000 | -4.54 | | | | | | | *Not evaluable: <5 paired patients in one group* |
| Overall change (T1–T3) | Panel 2 | CD38 balance in unswitched-memory B cells | CD38 compositional balance | 0 | 2 | | | | | 42.72 [35.18–50.27] | 44.51 [40.70–48.32] | 1.79 [-1.95–5.53] | 1.0000 | | | | | | | | *Not evaluable: <5 paired patients in one group* |

Between-group analyses required at least five paired patients in each response group. Within-group Wilcoxon P values are reported separately and may be available when the between-group comparison is not evaluable. APC, antigen-presenting cell; AUC, area under the receiver operating characteristic curve; CI, confidence interval; FDR, false discovery rate; IQR, interquartile range, presented as Q1–Q3; n, number of paired patients; NK, natural killer; pCR, pathological complete response; non-pCR, absence of pCR; q, Benjamini–Hochberg-adjusted P value; T1, baseline before treatment; T2, after completion of the first treatment block; T3, after completion of neoadjuvant treatment and before surgery.
