## Supplementary Figures for "CIRCULATING IMMUNE DYNAMICS DURING NEOADJUVANT CHEMOIMMUNOTHERAPY IN EARLY TRIPLE-NEGATIVE BREAST CANCER"

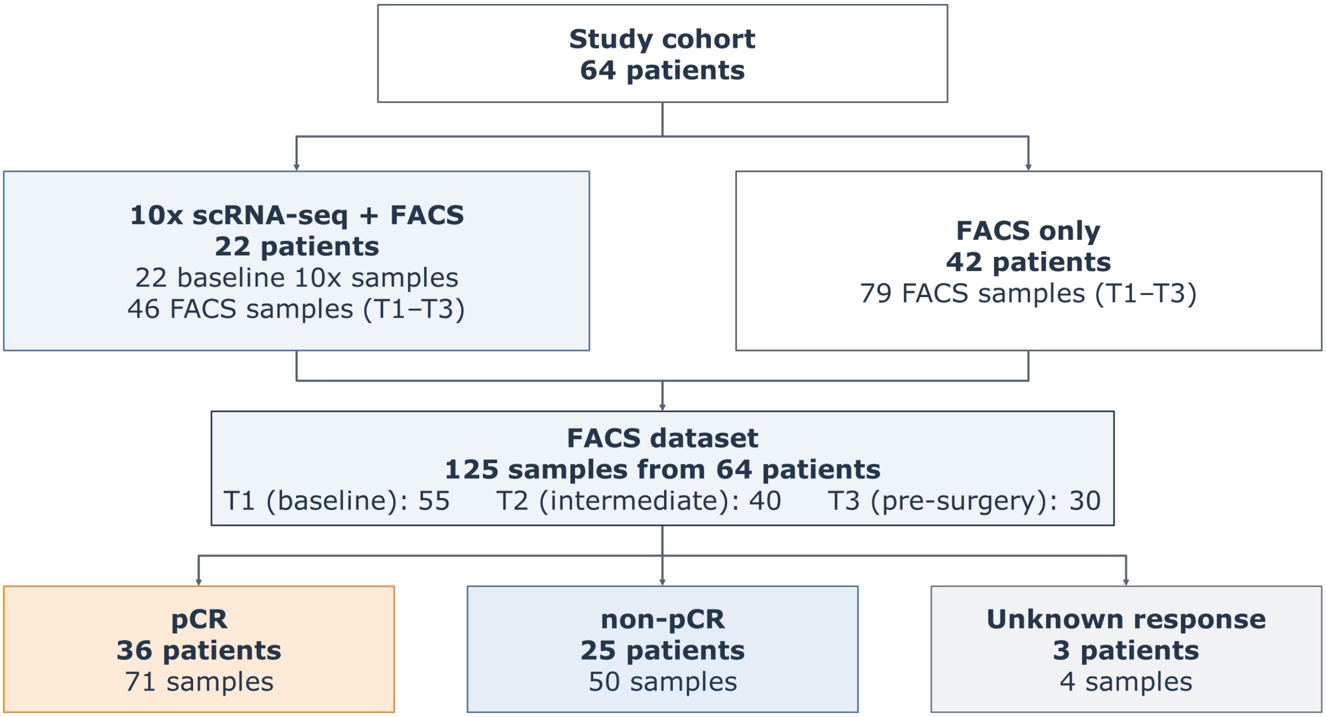


**Supplementary Figure 1. Study cohort and sample allocation for single-cell RNA sequencing and flow cytometry.** Samples with unknown pathological response were excluded from response-stratified comparisons.

pCR, pathological complete response.

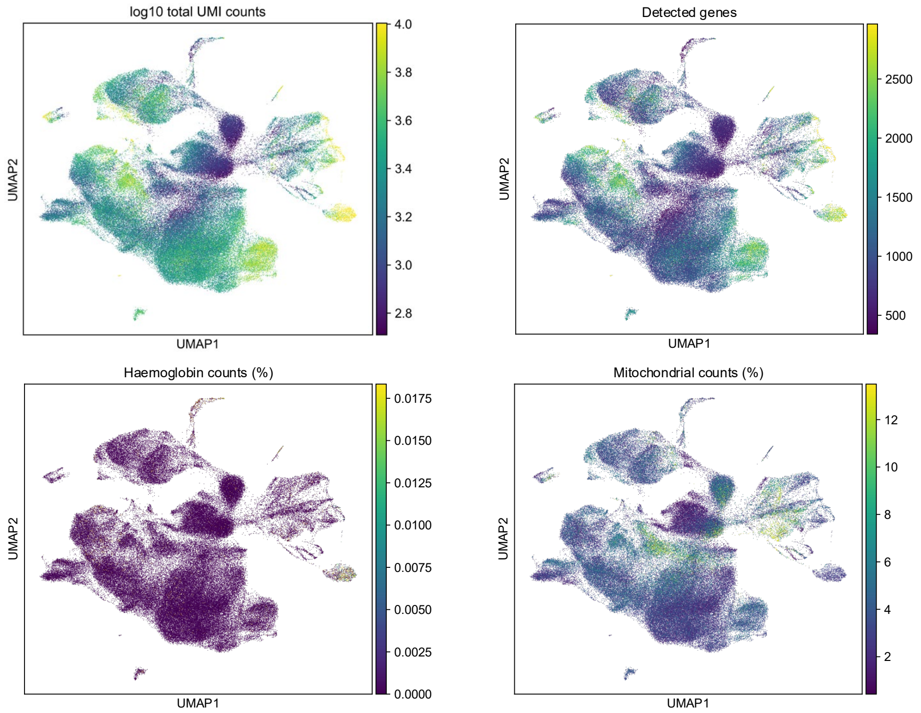


**Supplementary Figure 2. Quality-control metrics across the global peripheral blood mononuclear cell atlas.** UMAP representations of the 135,271 quality-controlled baseline peripheral blood mononuclear cells, colored according to log10-transformed total UMI counts, number of detected genes, percentage of counts assigned to hemoglobin genes and percentage of counts assigned to mitochondrial genes. Each point represents one cell, and the color scale indicates the value of the corresponding quality-control metric.

| Pathway (n = pCR / non-pCR) | T cells 14 / 8 | Cytotoxic T/NK 11 / 7 | T/NK intermediate 14 / 7 | NK cells 10 / 6 | B cells 14 / 8 | Myeloid/APC 10 / 5 |
|---|---|---|---|---|---|---|
| **GO biological processes** | | | | | | |
| T-cell receptor signaling | | | | | | 0.013 |
| T-cell activation | | | | | | |
| T-cell differentiation | | | | | | |
| B-cell activation | | | | | | |
| NK cell-mediated immunity | | | | 0.031 | | |
| Peptide antigen processing and presentation | | | | | 0.026 | |
| Phagocytosis | | | | | | |
| Lymphocyte migration | | | | | | |
| Leukocyte chemotaxis | | | | | | |
| **Hallmark gene sets** | | | | | | |
| Adipogenesis | | | | | | |
| Allograft rejection | | | | | | |
| Androgen response | | | | | | |
| Apical junction | | | | | | |
| Apical surface | | | - | | 0.009 | |
| Apoptosis | | 0.002 | | | 0.004 | |
| Bile acid metabolism | | | | | | |
| Cholesterol homeostasis | 0.002 | 0.002 | - | | 0.005 | |
| Coagulation | | | | | | |
| Complement | | | | | | 0.010 |
| DNA repair | 0.00031 | 0.010 | | 0.004 | 0.00061 | |
| E2F targets | 0.017 | 0.008 | | 0.007 | 0.00038 | |
| Epithelial-mesenchymal transition | | 0.042 | 0.032 | | | |
| Estrogen response (early) | | | 0.003 | 0.045 | | |
| Estrogen response (late) | | | | | | |
| Fatty acid metabolism | | | | 0.032 | | |
| G2/M checkpoint | | | | | 0.029 | |
| Glycolysis | | | | | | |
| Hedgehog signaling | | - | - | - | - | - |
| Heme metabolism | | | | | | |
| Hypoxia | | 0.020 | | | | |
| IL-2/STAT5 signaling | | | | | | |
| IL-6/JAK-STAT3 signaling | | | | | 0.044 | 0.00041 |
| Inflammatory response | | | | | 0.002 | 0.00063 |
| Interferon alpha response | | | | | | |
| Interferon gamma response | | | | | | |
| KRAS signaling (DN) | | | | 0.006 | | |
| KRAS signaling (UP) | | | | | | |
| Mitotic spindle | 0.025 | 0.037 | | 0.00064 | | |
| mTORC1 signaling | 0.00015 | 8.8e-07 | | 0.029 | 0.0002 | |
| MYC targets (v1) | 4e-05 | 6.5e-07 | 0.035 | 9.9e-05 | 1.1e-06 | |
| MYC targets (v2) | 0.048 | | - | | 0.012 | |
| Myogenesis | | | | | | |
| Notch signaling | | | - | | | |
| Oxidative phosphorylation | 0.001 | 0.010 | | 1.5e-05 | 0.004 | 0.008 |
| p53 pathway | 0.025 | 0.00028 | | | 0.013 | |
| Peroxisome | | | | | | |
| PI3K/AKT/mTOR signaling | | | | | | |
| Protein secretion | | | | | | |
| Reactive oxygen species pathway | | | - | 0.020 | 0.006 | |
| Spermatogenesis | | | | | | |
| TGF-beta signaling | | | | | | |
| TNF-alpha signaling via NF-kappaB | 6e-05 | 5.1e-10 | | | 2.4e-09 | 0.008 |
| Unfolded protein response | 0.037 | 0.0002 | | | 0.00013 | |
| UV response (DN) | 0.00057 | 0.006 | 0.001 | 0.00028 | 0.022 | |
| UV response (UP) | | 0.00032 | | | 2.3e-05 | |
| WNT/beta-catenin signaling | | | - | | | |
| Xenobiotic metabolism | | | | | | |

**Signed $-\log_{10}$ (nominal CAMERA *P*)**

**Towards non-pCR** — **Towards pCR**

−2 −1 0 1 2

Combination not tested

Numbers show nominal P < 0.05

**Supplementary Figure 3. Baseline pathway enrichment by pathological response across PBMC populations.** CAMERA was applied to patient-level pseudobulk profiles from pretreatment samples, yielding 331 evaluable comparisons across 57 gene sets and six cell populations. Colors represent signed $-\log_{10}$ nominal P values, with red indicating enrichment toward pCR and blue toward non-pCR. Outlined cells show nominal P < 0.05, and dashes indicate untested combinations. Benjamini–Hochberg correction was applied across all 331 comparisons.

Supplementary Figure 4

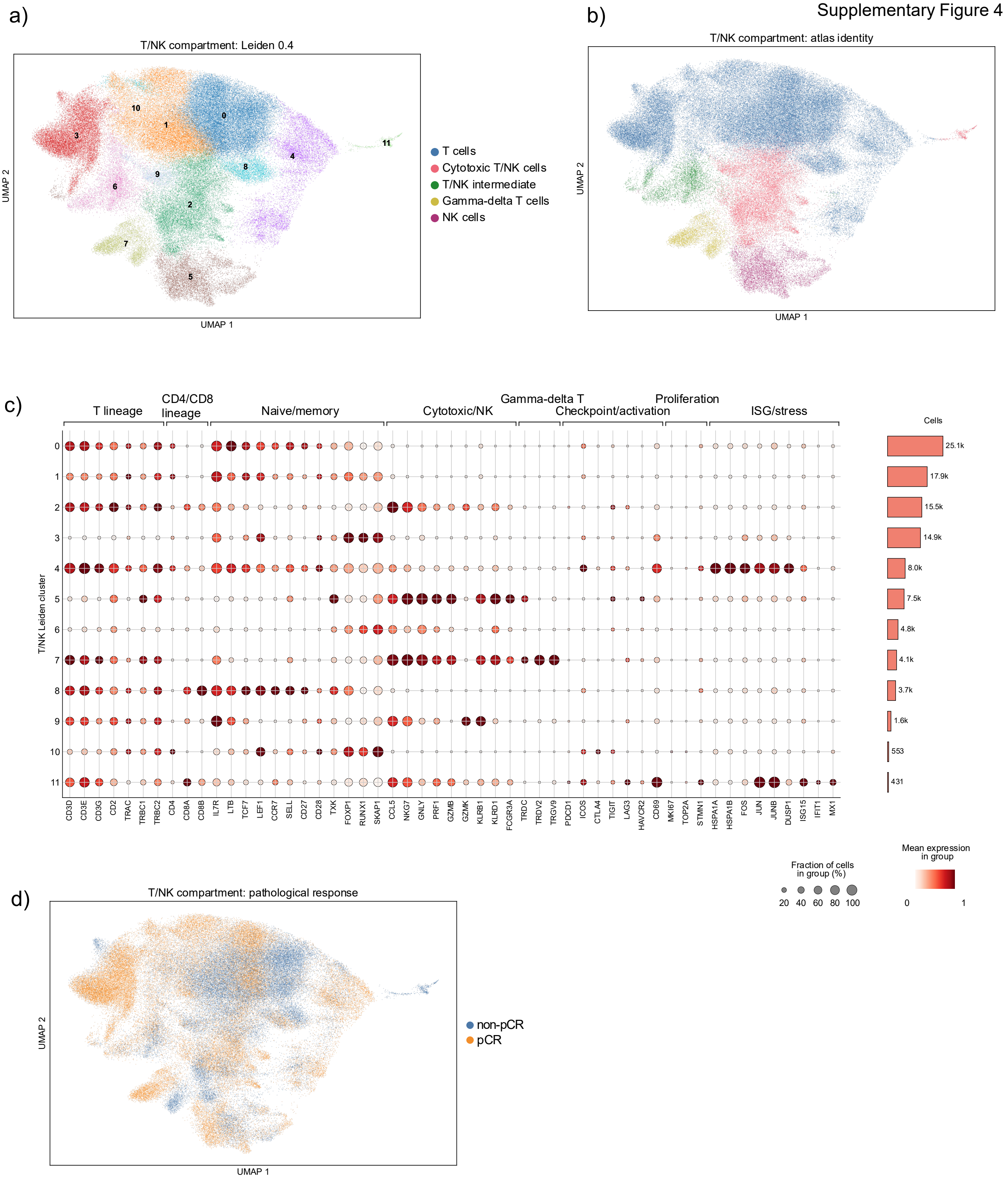


**Supplementary Figure 4. Baseline single-cell characterization of the circulating T/NK compartment. (a)** UMAP of the combined T/NK compartment after reclustering at Leiden resolution 0.4, showing 12 subclusters, C0–C11. **(b)** T/NK UMAP colored by the original atlas annotation. **(c)** Expression of genes associated with T-cell lineage, naïve/memory, cytotoxic/NK, γδ T-cell, checkpoint/activation, proliferation and ISG/stress programs across T/NK subclusters. Dot size indicates the fraction of cells expressing each gene, color intensity indicates mean expression and adjacent bars show cell counts. **(d)** T/NK UMAP colored by pathological response.

pCR, pathological complete response

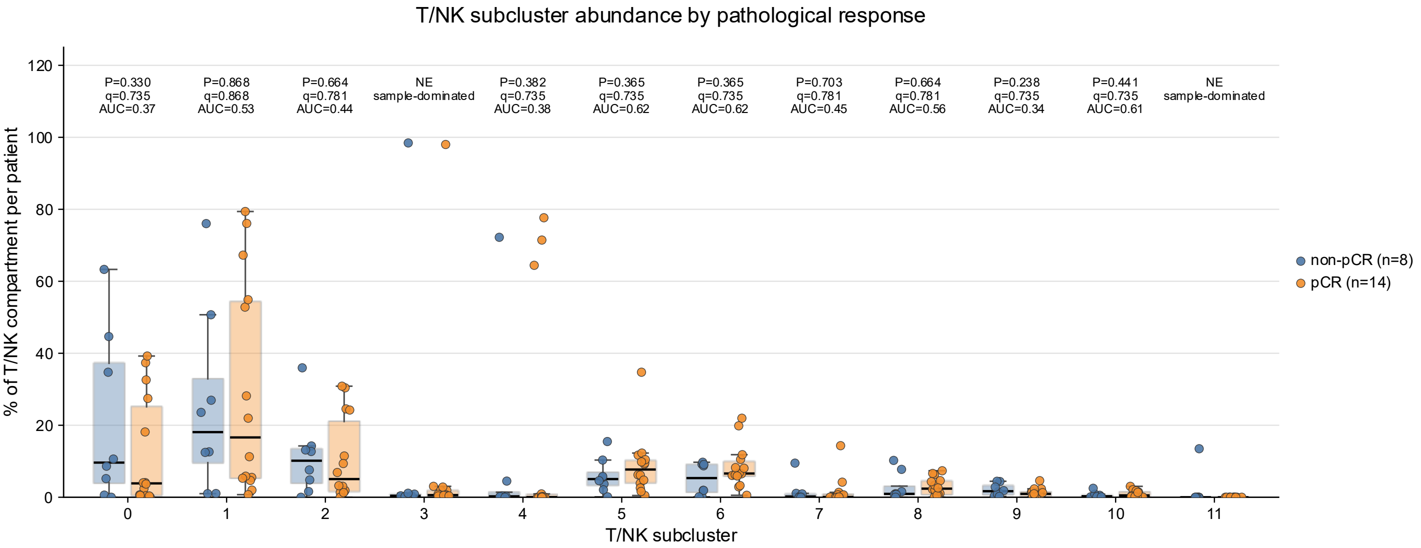


**Supplementary Figure 5. Patient-level abundance of T/NK subclusters according to pathological response.** Relative abundance of Leiden subclusters C0–C11 within the baseline T/NK compartment, shown separately for non-pCR (n=8) and pCR (n=14) patients. For each patient, subcluster abundance was calculated as the percentage of the total T/NK compartment assigned to the corresponding subcluster. C3 and C11 were retained descriptively but excluded from statistical inference because they were sample-dominated. The remaining subclusters were compared using two-sided Mann–Whitney U tests, with Benjamini–Hochberg correction across the ten evaluable subclusters. AUC values above 0.5 indicate higher abundance in pCR and values below 0.5 indicate higher abundance in non-pCR. Individual points represent patients; boxes show the median and interquartile range.

AUC, area under the receiver operating characteristic curve; NE, non-evaluable; pCR, pathological complete response.

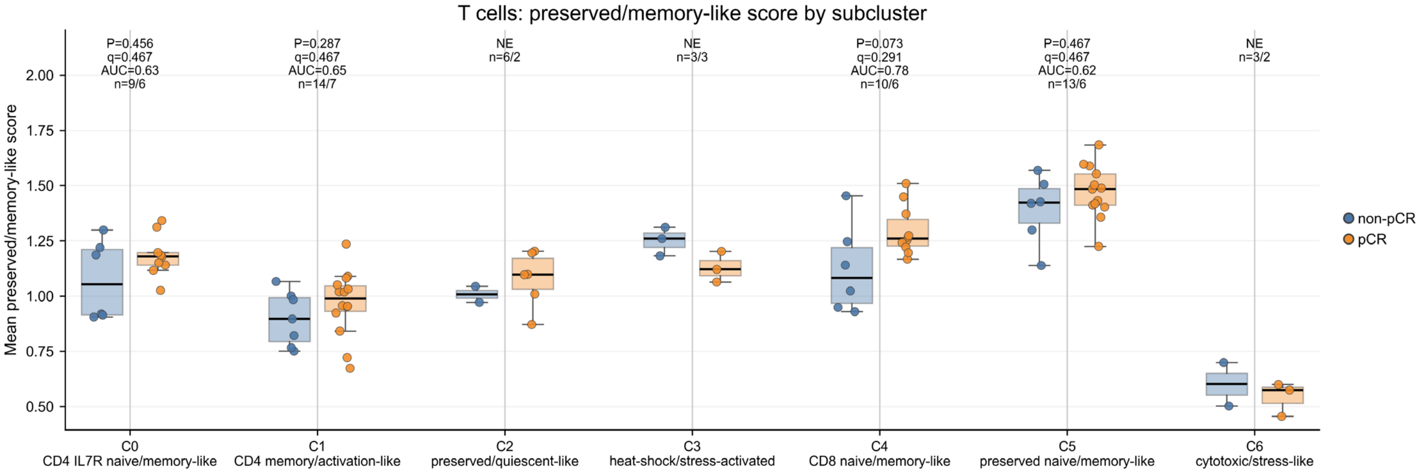


**Supplementary Figure 6. Preserved/memory-like program scores across T-cell subclusters.** Patient-level preserved/memory-like scores across T-cell subclusters C0–C6. The score was defined as the mean log-normalized expression of *IL7R, CCR7, SELL, TCF7, LEF1, LTB, CD27, CD28, TXK, FOXP1, RUNX1* and *SKAP1* across all cells contributed by each patient to the corresponding subcluster. Sample sizes are reported as pCR/non-pCR. Analyses required at least 30 cells per patient and at least five evaluable patients in each response group. C2 was excluded because it was sample-dominated, whereas C3 and C6 did not meet the minimum group size. P values were obtained using two-sided Mann–Whitney U tests and q values by Benjamini–Hochberg correction across the four evaluable subclusters, C0, C1, C4 and C5. AUC values above 0.5 indicate higher scores in pCR. Individual points represent patients; boxes show the median and interquartile range.

AUC, area under the receiver operating characteristic curve; NE, non-evaluable; pCR, pathological complete response

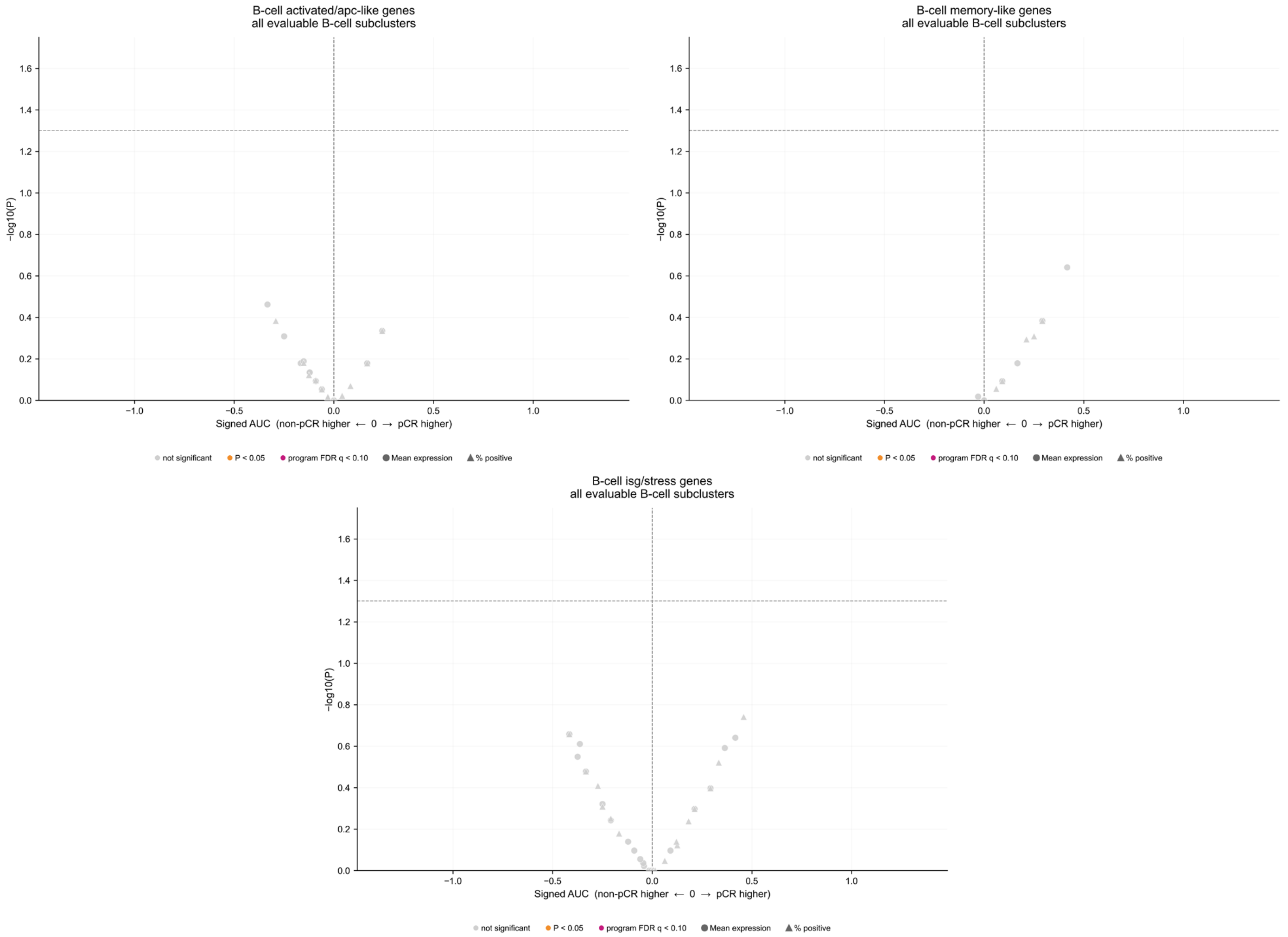


**Supplementary Figure 7. Gene-level analyses of memory-like, activated/APC-like and ISG/stress B-cell programs.** Volcano plots summarize patient-level gene–subcluster comparisons for the activated/APC-like, memory-like and ISG/stress programs across the two evaluable B-cell subclusters, C1 and C2. For each program gene, mean log-normalized expression and the percentage of cells with detectable raw-count expression were evaluated. Circles represent mean-expression comparisons and triangles represent percentage-positive comparisons. Groups were compared using two-sided Mann–Whitney U tests. The x-axis shows signed AUC, calculated as 2 × (AUC − 0.5), with positive values indicating higher values in pCR and negative values indicating higher values in non-pCR. The y-axis shows −log10(P). Gray points denote P≥0.05, orange points denote nominal P<0.05 and red points denote associations meeting q<0.10 after Benjamini–Hochberg correction across all genes, both evaluable subclusters and both expression metrics within the corresponding functional program. No nominal gene-level associations were detected in these three programs.

AUC, area under the receiver operating characteristic curve; pCR, pathological complete response.

### Supplementary Figure 8

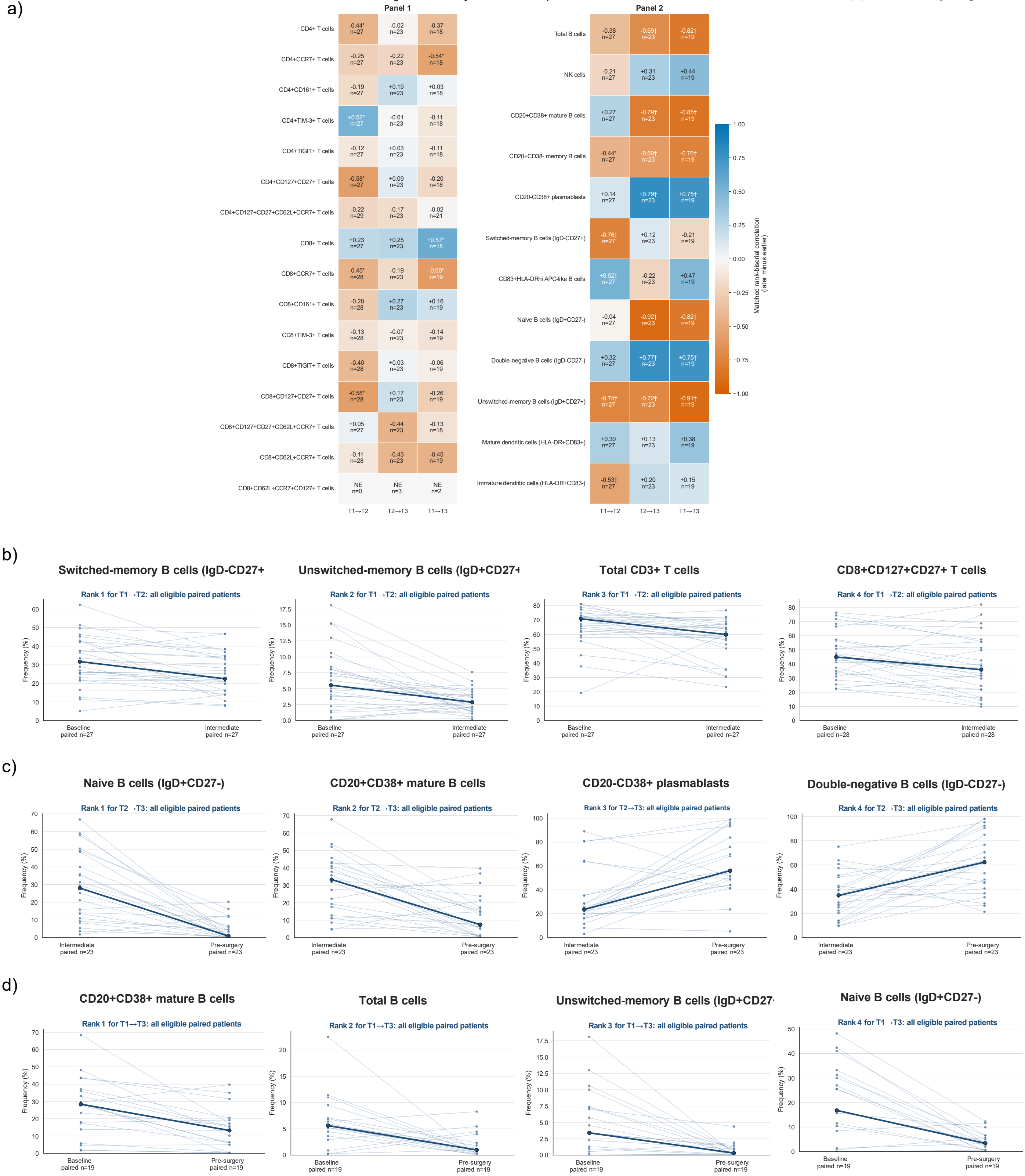


**Supplementary Figure 8. Treatment-associated longitudinal remodeling of circulating immune populations in the complete flow-cytometry cohort. (a)** Heatmaps summarizing longitudinal changes in the complete cohort for the T-cell populations included in Panel 1 and the B-cell, natural killer-cell and dendritic-cell populations included in Panel 2. Changes were evaluated between baseline and the intermediate treatment time point (T1–T2), between the intermediate and pre-surgery time points (T2–T3), and across the complete neoadjuvant period (T1–T3). Each heatmap cell reports the matched rank-biserial correlation and the number of evaluable paired patients. Positive values, shown in blue, indicate an increase at the later time point, whereas negative values, shown in orange, indicate a decrease. Top four immune populations (lowest nominal P values) for **(b)** T1–T2, **(c)** T2–T3 and **(d)** T1–T3. Thin lines connect paired measurements; thick lines indicate medians. Comparisons used two-sided Wilcoxon signed-rank tests with Benjamini–Hochberg correction within each panel and interval. *P<0.05; †q<0.05. NE, fewer than five evaluable pairs.

APC, antigen-presenting cell; NE, not evaluable; NK, natural killer.

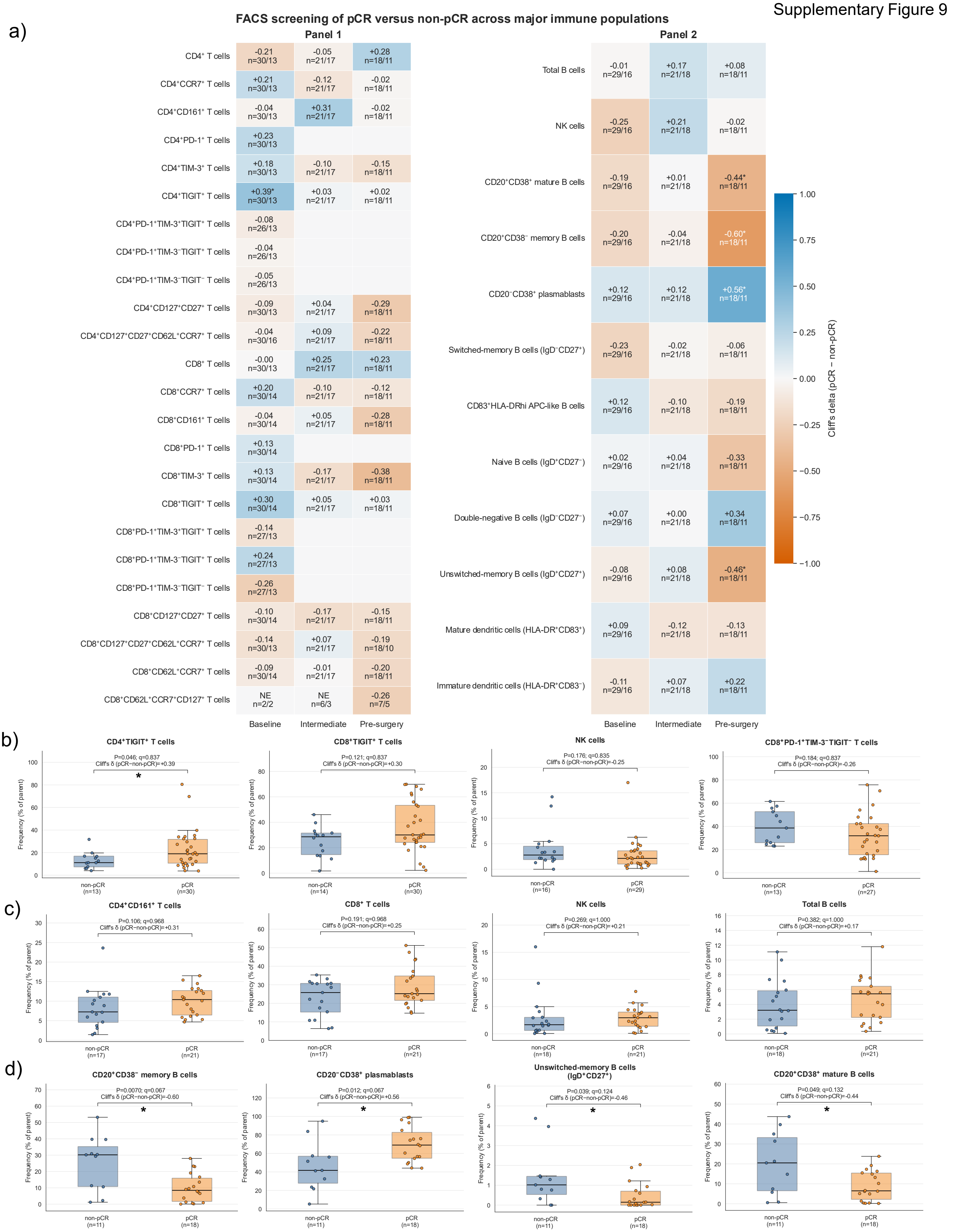


**Supplementary Figure 9. Cross-sectional flow-cytometric screening identifies time-dependent immune patterns associated with pathological response. (a)** Heatmaps summarizing pCR versus non-pCR comparisons for the immune populations evaluated in Panels 1 and 2 at baseline, the intermediate treatment timepoint and pre-surgery. Cells report Cliff's delta, calculated as pCR minus non-pCR, and the numbers of evaluable patients in each response group. Positive values indicate higher frequencies in pCR and negative values indicate higher frequencies in non-pCR. NE indicates that the prespecified minimum sample size was not reached. **(b–d)** Four selected immune populations from the broad flow-cytometry screen at **(b)** baseline, **(c)** the intermediate treatment timepoint and **(d)** pre-surgery. Sample sizes in the heatmaps are reported as pCR/non-pCR. Blank cells indicate PD-1-containing phenotypes not analyzed after baseline. Benjamini–Hochberg correction was applied within each flow-cytometry panel and timepoint.

Supplementary Figure 10

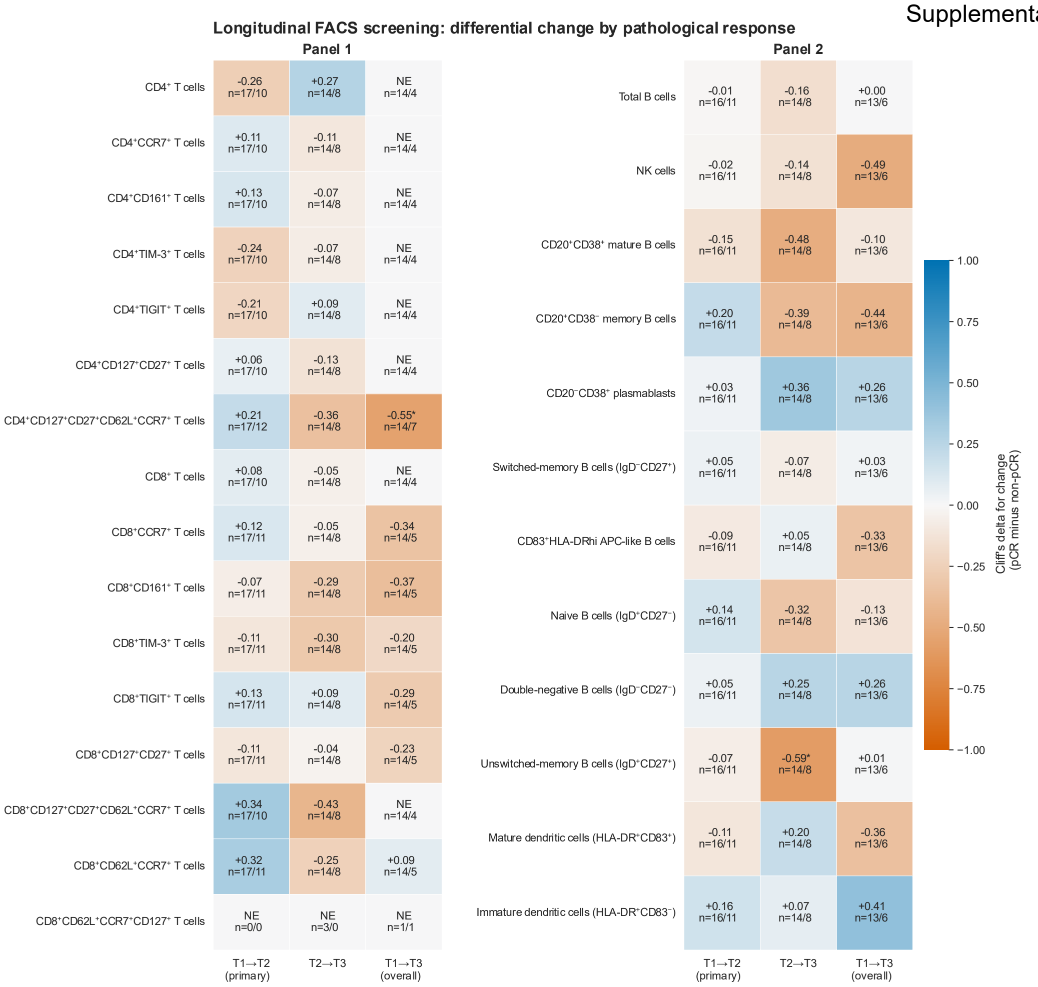


**Supplementary Figure 10. Response-associated differences in longitudinal changes in circulating immune populations.** Heatmaps summarize differences between pCR and non-pCR patients in the longitudinal changes observed from T1 to T2, T2 to T3 and T1 to T3. Analyses are shown separately for the T-cell populations included in Panel 1 and the B-cell, natural killer-cell and dendritic-cell populations included in Panel 2. For each patient, change was calculated as the later minus the earlier measurement and compared between response groups using a two-sided Mann–Whitney U test. Each cell reports Cliff's delta and the number of paired patients in the pCR/non-pCR groups. Cliff's delta was oriented as pCR minus non-pCR. Positive values, shown in blue, indicate a greater increase or smaller decrease in pCR, whereas negative values, shown in orange, indicate a greater increase or smaller decrease in non-pCR. * denotes nominal P<0.05 and † denotes q<0.05 after Benjamini–Hochberg correction within each flow-cytometry panel and time interval. NE indicates fewer than five paired patients in either response group.

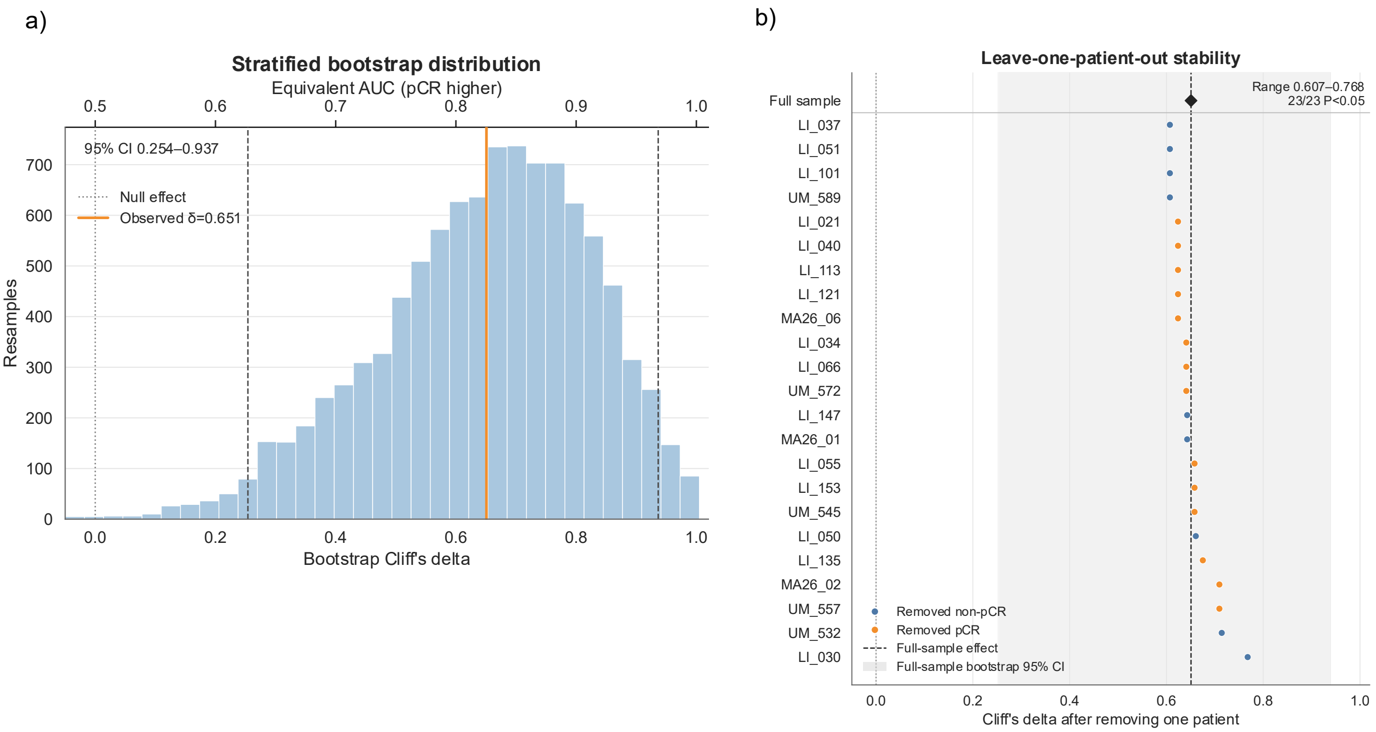


**Supplementary Figure 11. Robustness of the pre-surgery switched-memory B-cell CD38 balance. (a)** Stratified bootstrap distribution of Cliff's delta for the T3 switched-memory CD38 balance, defined as 100 × $CD38^{+}/(CD38^{+} + CD38^{-})$. A total of 10,000 patient-level bootstrap samples were generated with stratification by pathological-response group. The orange line indicates the observed effect (Cliff's δ=0.651), dashed lines indicate the bootstrap 95% confidence interval (0.254–0.937), and the null value is δ=0, equivalent to an AUC of 0.5. The corresponding AUC was 0.825, with a bootstrap 95% confidence interval of 0.627–0.968. **(b)** Leave-one-patient-out analysis in which the comparison was repeated after sequential exclusion of each of the 23 patients with an evaluable T3 measurement. Cliff's δ ranged from 0.607 to 0.768 and the AUC from 0.804 to 0.884; all 23 comparisons remained nominally significant. Points show the resulting Cliff's delta after excluding the indicated patient and are colored according to the response group of the excluded patient. The vertical dashed line indicates the full-cohort estimate, and the shaded region represents its bootstrap 95% confidence interval.

AUC, area under the receiver operating characteristic curve; CI, confidence interval; pCR, pathological complete response; T3, post-neoadjuvant, pre-surgery time point.
